# Molecular characterization of liver steatosis through circulating proteins, metabolites and lipids

**DOI:** 10.64898/2025.12.01.25341334

**Authors:** Marilyn De Graeve, Essi Hantikainen, Philippine Louail, Vinicius Verri Hernandes, Cristian Pattaro, Francisco S. Domingues, Peter P. Pramstaller, Herbert Tilg, Markus Ralser, Christoph Grander, Johannes Rainer

## Abstract

**Background:** Metabolic dysfunction-associated steatotic liver disease (MASLD) is among the most prevalent liver disorders in Western countries. While hepatic steatosis is prevalent, molecular mechanisms associated with steatosis grades and how nutrition modulates related metabolic disturbances remain poorly understood.

**Methods:** We quantified 175 metabolites and lipids and 148 proteins in serum samples of 327 participants of the CHRIS NAFLD study to identify circulating biomarkers of hepatic steatosis severity. Participants, all without any strong clinical manifestation of liver disease, were stratified by steatosis grade based on ultrasound-derived measurement. Associations between dietary intake and steatosis were also investigated, and a potential influence of diet on the identified biomarkers was evaluated.

**Results:** Hepatic steatosis was positively correlated with BMI and a higher proportion of T2D and MASLD cases was present in the high grade steatosis group. A considerable number of plasma proteins, metabolites, and in particular lipids were found significantly associated with steatosis, mainly reflecting a broader metabolically unhealthy phenotype. Adherence to a plant-based diet was associated with lower steatosis grades and was unrelated to the identified circulating steatosis biomarkers.

**Conclusions:** Our results suggest that circulating metabolic signatures of steatosis reflect broader metabolic dysregulation and are independent of dietary intake. These findings support the use of blood-based multi-omics profiling to discriminate between metabolically healthy and unhealthy individuals, and may thus improve MASLD risk stratification.

**Highlights:**

- Biomarkers for hepatic steatosis severity identified through multi-omics profiling
- Steatosis-associated profiles reflect broad metabolic dysfunction
- Adherence to plant-based diet is associated with lower steatosis grade
- Identified steatosis blood biomarkers are independent of dietary habits

**Graphical abstract:** 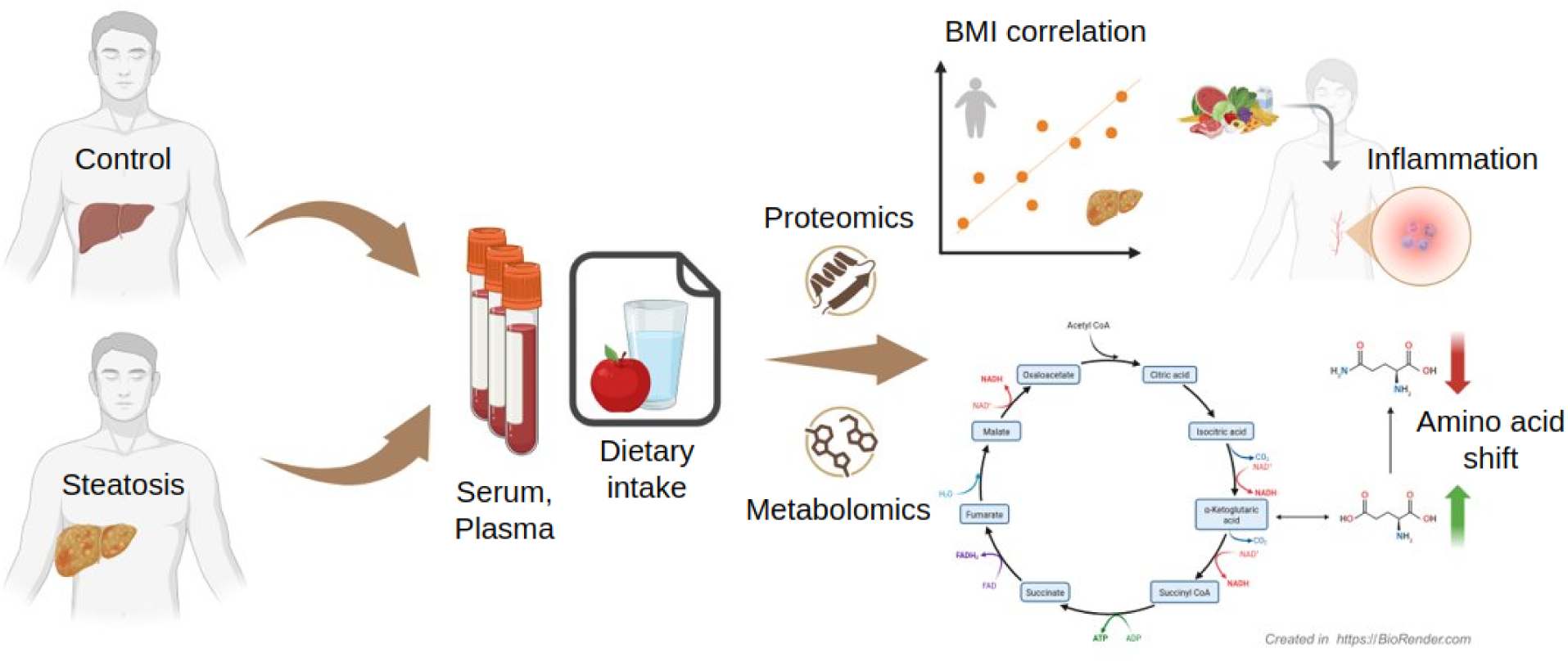

## Introduction

Hepatic steatosis, characterized by the accumulation of triglycerides in hepatocytes, is a key feature in diagnosing metabolic dysfunction-associated steatotic liver disease (MASLD). MASLD has become a significant public health concern due to its rising prevalence and its potential to progress into more severe liver conditions, including metabolic dysfunction-associated steatohepatitis (MASH), cirrhosis and hepatocellular carcinoma (HCC). Steatosis is considered a hepatic manifestation of metabolic syndrome (MetS), and is strongly associated with obesity, type 2 diabetes (T2D) as well as chronic cardiovascular diseases (CVD) (1–4).

Dietary factors play an essential role in the development as well as management of MASLD. Previous literature has shown variations in macronutrient intake by MASLD status (5). Steatosis is closely and bidirectionally linked to hepatic and peripheral insulin resistance, particularly in individuals with obesity (6). Insulin resistance leads to increased lipolysis in adipose tissue, as well as altered *de novo* lipogenesis in hepatocytes, resulting in triglyceride accumulation (7). Therefore, plant-based diets are gaining attention as a sustainable and health-conscious alternative for managing liver disease (8).

Hepatic steatosis is commonly assessed using indirect or semi-quantitative methods, such as liver enzyme levels, imaging surrogates like computer tomography attenuation, or scoring systems based on clinical and metabolic parameters. However, these approaches lack the sensitivity and specificity to accurately quantify liver fat content, particularly in early or moderate stages of steatosis (9–12). More recent studies have adopted non-invasive methods like controlled attenuation parameter (CAP), obtained during a liver stiffness measurement (FibroScan®), to provide a feasible, more precise, quantitative assessment of hepatic fat (13–15).

Advances in multi-omics technologies, such as proteomics, metabolomics, and lipidomics, offer new avenues to study the molecular mechanisms underlying steatosis and its progression. The application of high-throughput omics technologies to study steatosis not only enhances our understanding of its molecular drivers but also paves the way for targeted prevention and treatment strategies to halt or reverse the progression of liver disease.

To further explore the link between steatosis and its proteomic and metabolic influences, we use cross-sectional data from the CHRIS NAFLD study cohort (n=327 participants) (16), with participants recruited from the Cooperative Health Research in South Tyrol (CHRIS) study (17,18), that includes individuals with varying liver steatosis grades. By combining plasma proteomics (148 proteins) with serum metabolite and lipid profiling (175 metabolites), we investigate how such circulating biomarkers could help classify liver steatosis severity and assess the role of diet.

## Methods

### Study cohort

The Cooperative Health Research in South Tyrol (CHRIS) study is a population-based cohort of 13,393 adults aged 18 and over, recruited in the alpine Val Venosta/Vinschgau district in South Tyrol (Italy) (17,18). For the CHRIS NAFLD study 356 participants of the CHRIS study with and without a diagnosis of T2D were recalled and additional phenotyping and sample collection performed (16).

### Classification of steatosis

To evaluate the presence of hepatic steatosis and hepatic fibrosis, different non-invasive measurement tools were used. For liver steatosis assessment, participants underwent abdominal transient elastography (Fibroscan^®^, Echosens, France) after overnight fasting (15). The presence and severity of steatosis was evaluated as documented by Ballestri et al. (19). Steatosis was quantified using CAP (20), and individuals were classified into four groups, corresponding to varying steatosis grades. Control participants, having no abnormally increased liver fatty change, were defined as having a CAP score from ≥0 to <238 dB/m. Grade 1, 2, and 3 steatosis was defined as a CAP of ≥238 to ≤260 dB/m, >260 to ≤290 dB/m, and >290 to ≤400 dB/m, respectively (21).

### Proteomics and metabolomics, including lipids, data

Plasma proteomics data was generated using the same mass spectrometry based-approach and methodology used in (22). Absolute concentrations for 175 metabolites and lipids were measured in serum samples of participants using a targeted metabolomics approach employing the Biocrates AbsoluteIDQ® p180 kit following the same procedures and methods described in (23). Full results and lists of quantified proteins, metabolites, and lipids are provided in Supplementary Files 2 and 3, respectively.

The methodology for the additional untargeted metabolomics data is provided in the Supplement in section “Untargeted metabolomics data analysis”.

### Dietary intake

Dietary intake was assessed using the self-administered, semi-quantitative Global Allergy and Asthma European Network (GA2LEN) food frequency questionnaire (FFQ) (24), which has been described in detail elsewhere (25). The FFQ records the consumption of 229 foods and beverages that were categorized into 32 sections. Participants were asked to report their average consumption frequencies for the past 12 months for each item, considering standard portion sizes. Consumption frequency of specific food items was transformed into grams per day and converted into specific macronutrient estimates using the latest edition of the McCance & Widdowson’s Food Composition Tables (26). Total energy intake (TEI, in kcal) was estimated as energy coming from proteins (4 kcal/g), fat (9 kcal/g), carbohydrates expressed as monosaccharides (3.75 kcal/g) and alcohol (7 kcal/g) (27).

Daily protein, carbohydrate and fat intake was estimated expressed as percentage of TEI per day (protein %energy = (protein in g × 4 kcal/g)/TEI in kcal) × 100; carbohydrates %energy = (carbohydrates in g × 3.75 kcal/g)/TEI in kcal) × 100; fat %energy = (fat in g × 9 kcal/g)/TEI in kcal) × 100). We additionally distinguished between saturated, polyunsaturated, monounsaturated and trans fatty acids.

For the Plant-based Diet Index (PDI) score, the FFQ items were categorized into eighteen food groups as outlined by Satja et al. (28). The consumption of the index was expressed in servings/day. After categorizing intake into quintiles, a positive score was assigned to the “healthy” and “less-healthy”/“unhealthy” plant foods, and a reverse score to the animal-based foods. For the positively scored items participants with an intake above the highest quintile received a score of 5 and those below the lowest quintile intake received a score of 1. Reverse values were assigned for the reversely scored items. For each participant, the individual scores of the 18 food groups were then summed up to create the final score.

### Statistical data analysis

The data analysis workflows are available in the GitHub repository https://github.com/EuracBiomedicalResearch/nafld_proteo_metabolomics. Data analysis was conducted in R v4.5.2. Steatosis measurements with a Fibroscan liver stiffness interquartile range score above 30 were removed. The final data sets used in all analyses were restricted to samples without missing values in any of the main covariates (steatosis, age, sex) and available metabolomics, proteomics and FFQ data and consisted of data from n=327 participants. Protein and metabolite abundances were log2 transformed and had log-normal distributions. Only the metabolomics data had missing values (1.24%), no imputation was performed.

Multiple linear regression analyses were performed for the proteomics data (148 proteins), targeted metabolomics data (175 metabolites and lipids), FFQ derived macronutrients (7 items), and the PDI score. A separate model was fitted for each of these analytes explaining the response variable (e.g. abundance of a protein) by the covariates steatosis grade (categorical), age (numeric) and sex (binary). For dietary intake, the estimated daily total energy intake (kcal) was additionally included as an explanatory variable in the models. Significant response variables were defined based on the coefficient for steatosis grade 3, which represents the difference between individuals with the highest steatosis grade (n = 81) and the base level (n = 140). All p-values were adjusted for multiple hypothesis testing using the Benjamini-Hochberg method for a strong control of the false discovery rate (FDR). Associations with an adjusted p-value<0.05 were considered statistically significant. For increased confidence, we required for protein, metabolite and lipid associations, in addition to the statistical significance, that also the observed difference in concentrations was more than 50% of the coefficient of variation (CV) for a particular analyte. This CV represents the technical variance of a measurement estimated on the available study-internal quality control samples.

### Sensitivity analyses

To evaluate potential confounding of the results by T2D the coefficients for steatosis from the models were compared to those from fitted models that included in addition T2D as an explanatory (categorical) variable.

The influence and potential confounding of the PDI score on the steatosis results was evaluated by comparing coefficients for steatosis between the linear models with and without PDI score as additional explanatory variables.

### Pathway analysis

For pathway analysis, UniProt and HMDB IDs for significant proteins and metabolites were submitted to the RAMP-DB (https://rampdb.nih.gov/pathway-enrichment, v3.0.7) online tool (29). This resource integrates multiple pathway databases, including KEGG, Reactome, and WikiPathways, to perform robust enrichment analyses. Results were exported and served as the basis for downstream clustering and curation.

The enriched pathways for proteins and metabolites were manually reviewed and grouped into biologically meaningful pathway clusters. Particular attention was given to recurring themes across pathways, allowing for aggregation into higher-level biological processes (e.g., amino acid metabolism, lipid metabolism, coagulation cascade).

## Results

### Study sample characteristics and general data overview

For the present analysis data from 327 participants of the CHRIS NAFLD study cohort were used (see Table 1). From these, 140 (42.8%) showed no signs of steatosis, while 187 (57.2%) were categorized in steatosis grades 1-3. In the present cohort, a higher percentage of men were classified with steatosis grade 3, while the percentage of women is higher in the control group. For anthropometric traits, body mass index (BMI), body fat, visceral fat and waist circumference were all strongly, and positively, related with steatosis grade (Table 1 and Supplementary Figure S1).

**Table 1.** Demographics and relevant clinical parameters of the CHRIS NAFLD study cohort. Values are presented as mean (with standard deviation in brackets), absolute numbers or percentages per steatosis grade. * Values with significant differences (p-value < 0.05) compared to controls.

|  | **Controls** | **Grade 1** | **Grade 2** | **Grade 3** |
| --- | --- | --- | --- | --- |
| **n** | 140 | 52 | 54 | 81 |
| **% Women** | 55.0 | 55.8 | 51.9 | 42.0 |
| **Age** | 67.1 (12.1) | 70.6 (9.13) | 69.4 (9.01) | 65.8 (9.37) |
| **BMI** | 26.0 (4.07) | 26.9 (3.18) | 29.6 (5.24) * | 30.8 (4.94) * |
| **Visceral fat** | 9.76 (3.94) | 11.1 (3.45) * | 12.6 (3.88) * | 14 (4.39) * |
| **Body fat** | 30.7 (10.1) | 32.5 (8.92) | 34.8 (10.1) * | 35.8 (10.2) * |
| **Waist circumference** | 88.2 (12.1) | 90.7 (9.77) | 98.0 (11.7) * | 102.0 (11.2) * |

We next evaluated the relationship between steatosis and other liver health related traits in the present data set. While no relationship between steatosis grade and the liver fibrosis score FIB4 was present (Figure 1A), FibroScan quantified liver stiffness was significantly higher in steatosis grade 2 and 3 compared to controls (Figure 1B). Also, the percentage of participants diagnosed with T2D or MASLD was higher in steatosis grade 3, although a considerable proportion of controls were also positive for T2D and MASLD (Figure 1C and 1D). Related to these diagnoses, also the percentage of individuals taking T2D medication, and to a lesser extent, lipid modifying medications, was higher in grade 3 steatosis than in controls (34% *vs* 18% for T2D and 34% *vs* 26% for lipid modifying medication).

**Figure 1:**
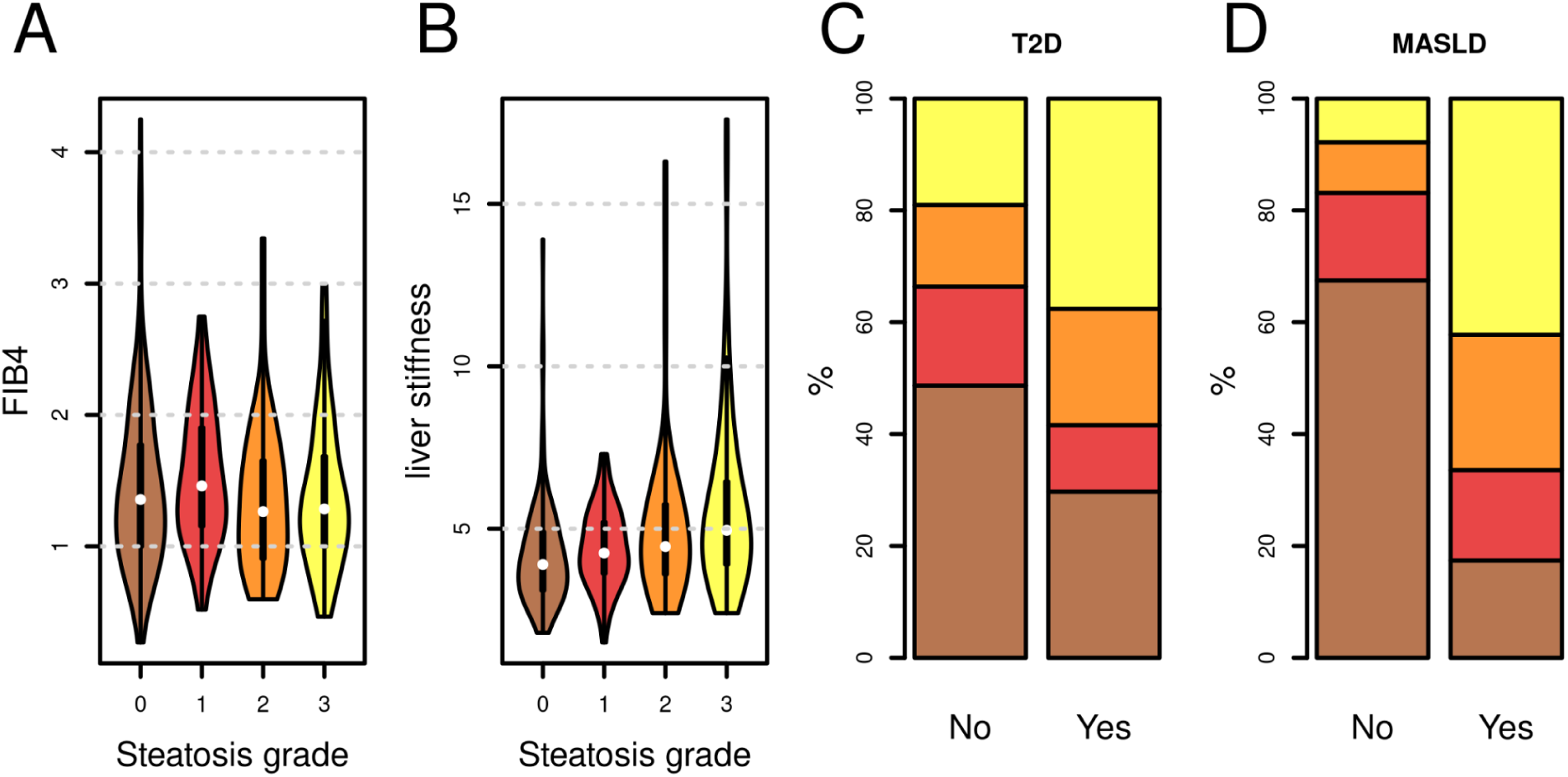
Relationship between liver steatosis and liver health-related traits. A) Estimated liver fibrosis score (FIB4) across steatosis grades. B) FibroScan quantified liver stiffness across steatosis grades. Differences for participants with steatosis grade 2 and 3 compared to controls are significant (p-value 0.006 and < 0.001, respectively). C) Percentages of participants diagnosed with T2D per steatosis grade (represented in the colors from panel A). D) Percentages of participants diagnosed with MASLD per steatosis grade (represented in the colors from panel A).

Using a targeted metabolomics approach, we quantified 175 metabolites and lipids in the participants’ blood serum samples. An additional untargeted metabolomics setup was employed for detection of small polar molecules. To complement these, also 146 proteins from the high abundance plasma proteome were measured in blood plasma samples of the same participants. These proteins are mainly involved in nutrient transport, coagulation and immune system activity. Principal component analysis (PCA) of the z-score transformed metabolite, respectively protein abundances suggest a separation of participants by steatosis grade on principal components 3 and 4 (see Figure 2B and 2D). Otherwise, no clear pattern or clustering was observed in the PCA.

**Figure 2:**
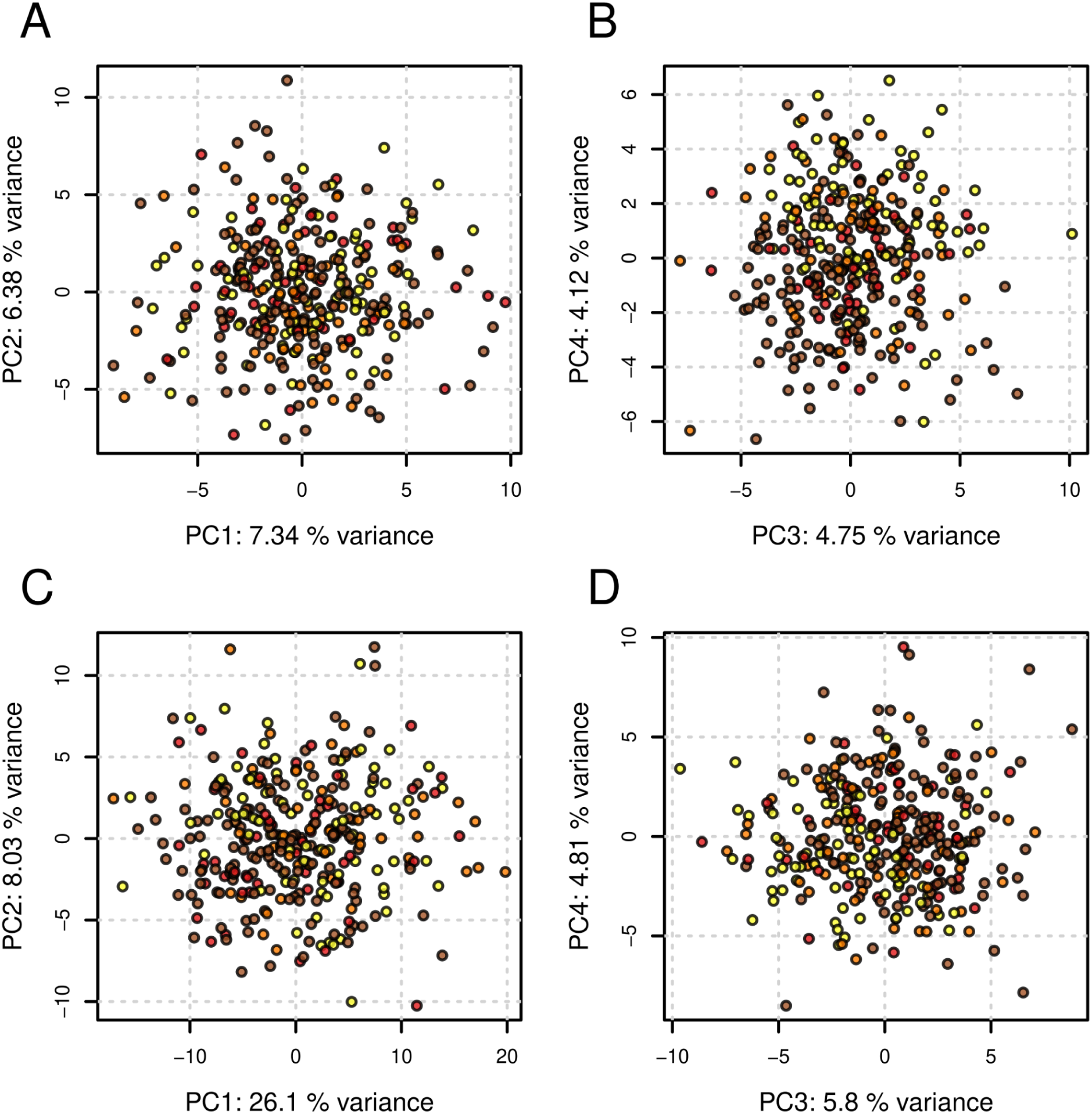
Principal component analysis of the proteomics and metabolomics data sets. Panels A) and B) show the results for the proteomics, C) and D) for the metabolomics data. The color depicts the steatosis grade for each participant (brown, red, orange and yellow for controls and steatosis grades 1 to 3). A slight separation by steatosis grade is visible for both data sets in PC3 and PC4 (panels B and D).

### Circulating proteins associated with liver steatosis

Using multiple linear regression analyses, we next evaluated associations between circulating proteins and steatosis grade, adjusting in addition for the participants’ age and sex. From the tested 148 proteins, 2, 5, and 24 were found to have significantly different abundances between steatosis grade 1, 2, and 3 compared to the control group (see Figure 3A and Supplementary Table S1; full results available in Supplementary File 2 and Supplementary Figure S2). The 24 proteins associated with grade 3 steatosis include 5 complement components and -factors, 4 serine proteinase inhibitors, 2 apolipoproteins and 2 Inter-alpha-trypsin inhibitor heavy chain proteins all showing the same trend of differential abundance, albeit to a lesser extent, already with lower steatosis grades. A pathway enrichment analysis of steatosis grade 3 associated proteins resulted in 12 significant (but partially redundant Wikipaths and reactome) pathways (Supplementary Figure S3). The most enriched and relevant one was the complement and coagulation cascades pathway comprising 7 of the 24 significant proteins (Supplementary Figure S4).

**Figure 3:**
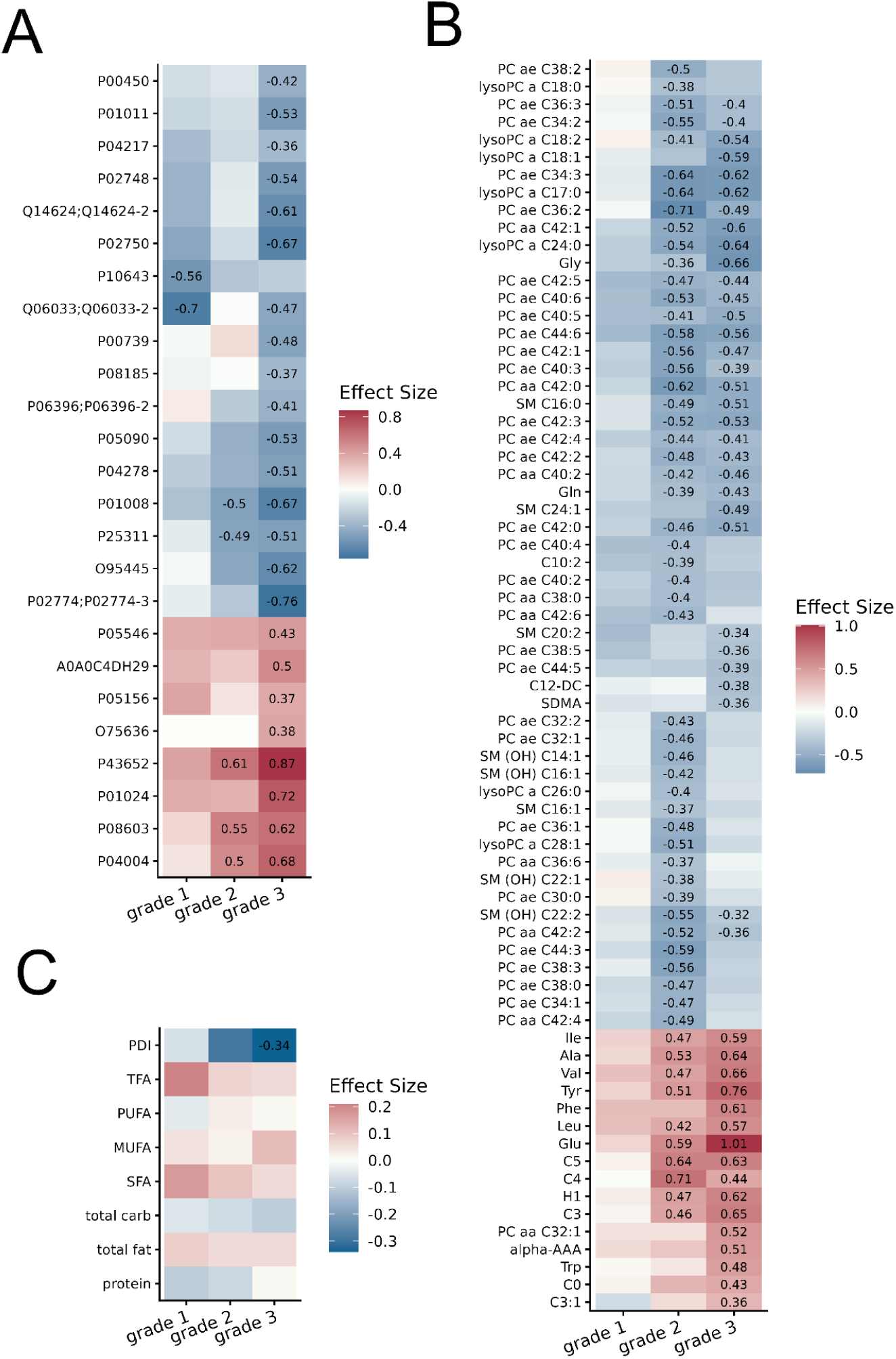
Association of circulating proteins, metabolites and lipids and dietary characteristics with steatosis grades. Shown are effect sizes (color coded) from the different linear regression analyses, with the effect size value displayed for significant associations between the respective variable and steatosis grade. A) Circulating proteins with a significant association with at least one steatosis grade. B) Metabolites and lipids with a significant association with at least one steatosis grade. C) Effect sizes for all tested macronutrient groups and the plant-based diet index (PDI). TFA: trans-fatty acids, PUFA: poly-unsaturated fatty acids, MUFA: mono-unsaturated fatty acids, SFA: saturated fatty acids, total carb: total carbohydrates.

Note that due to the unbalanced distribution of T2D cases across steatosis grades (Figure 1C), the relationship between BMI (Supplementary Figure S1C) and the investigated phenotype in our cohort, we did not adjust for these variables in our regression models. However, as shown by a sensitivity analysis, the influence of T2B on the proteomics association results was only marginal (Supplementary Figure S5).

### Metabolites and lipids associated with liver steatosis

From the 175 investigated metabolites and lipids, none, 58, and 48 were found significantly associated with liver steatosis grade 1, 2, and 3, respectively (Supplementary Table S2). Similar to the proteomics result, the effect size and significance levels increased for most metabolites with steatosis grade (See Figure 3B and Supplementary Figure S6; full results available in Supplementary File 3). However, some lipids showed, in line with previous observations (30), a stronger association with steatosis grade 2 than 3. The strongest associations with the largest effect sizes were for amino acids, 8 of them, including glutamate, having higher concentrations in steatosis grade 3, while 2 were negatively associated with steatosis. For the lipid data, most of the significant molecules were phosphocholines (PC) with all, except PC aa C32:1, showing lower abundances in steatosis grade 3 compared to control individuals. The effect sizes for these associations were similar to those from linear models accounting also for T2D (See Supplementary Figure S7) suggesting independence of the results from this potential confounder. The exception was hexose sugar (H1) that showed some difference in its effect size (See Supplementary Figure S7).

We next conducted a pathway enrichment analysis for the steatosis grade 3 associated metabolites and lipids which revealed significant pathways grouped into 15 clusters (Supplementary Figure S8). A large set of significantly associated metabolites were related to the amino acid metabolism, summarized in the biosynthesis of amino acids pathway (Supplementary Figure S9). For completeness, we also analyzed the untargeted metabolomics data generated for the present study cohort. A total of 10,882 LC-MS features were analyzed, with 8 being significantly associated with steatosis grade 3 (Supplementary Table S3). Due to the lack of available MS2 data, however, the identified LC-MS features, i.e., ions of molecules characterized by their measured mass-to-charge ratio and retention time, could not be annotated to metabolites and we hence applied the Mummichog algorithm for a tentative annotation through common biochemical pathways. This analysis revealed 14 enriched pathways (Supplementary Table S4) with putative annotations of 3 significantly discriminating metabolites: N-Acetylmethionine, 1-Pyrroline-4-hydroxy-2-carboxylate and L-4-Hydroxyglutamate semialdehyde, all related to amino acid metabolism (full results and related methodology are presented in Supplementary Section “Untargeted metabolomics data analysis”).

### Associations between dietary characteristics and liver steatosis

Finally, we investigated a possible relationship between dietary habits and steatosis. We summarized macronutrient intake and the plant-based diet index (PDI) from the participants’ food frequency questionnaires (FFQ) and conducted linear regression analyses adjusting in addition for the estimated daily total energy intake. While none of the macronutrient groups was significantly associated with any of the steatosis grades (Supplementary Table S5), the PDI was significantly, and strongly negative, associated with liver steatosis grade 3 (p-value 0.0099, effect size −0.35). Associations for steatosis grade 1 and 2 also had negative coefficients that did however not reach statistical significance (Figure 3C and Supplementary Table S6).

To evaluate a potential influence of dietary habits on the identified statosis-associated proteins, metabolites or lipids, we repeated the regression analyses including the PDI as a covariate and compared the resulting effect sizes with those from the original analysis. The results from the models with and without adjustment for PDI were highly similar, arguing against an influence of dietary intake on the present results (Supplementary Figures S10 and S11). Therefore, in our cohort, dietary habits did not explain the observed differences in concentrations of circulating proteins, metabolites, and lipids.

## Discussion

In this study, we investigated the relationships between circulating protein, metabolite, and lipid profiles with hepatic steatosis severity, using samples from a well-characterized, fasting, single-site population cohort. Our analyses identified a set of proteins and metabolites with significantly different abundances in participants with grade 3 steatosis. These are discussed in detail in Table 2.

**Table 2.** Circulating proteins, metabolites, and lipids associated with grade 3 steatosis. Variable: significant protein, metabolite or lipid. Direction: abundance in high grade steatosis relative to control individuals. Discussion: summary of the protein, metabolite or lipid function in context of the relevant literature.

| Variable | Direction | Discussion |
| --- | --- | --- |
| **Proteins** |  |  |
| ITIHs | Lower | Interestingly, ITIH4 was previously described to be an acute-phase protein and novel biomarker for cholestatic liver disease (32). Furthermore, ITIH4 was proposed as a biomarker for fibrosis in hepatitis C (33,34). ITIH3 was recently described as an important regulator of hepatic steatosis, as knock-down of ITIH3 in a murine model led to increased hepatic triglyceride accumulation in different models of MASLD (35). |
| APOs | Lower | The APOD and APOM proteins were thus far not discussed in steatosis research, but expression of the gene Apo M has been reported to be significantly lower in steatotic liver disease, independently if MASLD is present or not (36). As the liver is a major organ in fatty acid metabolism, these results are not unexpected. |
| SERPINS | Depending molecular species | The significant SERPINS have not yet been described in relation to steatosis, except for SERPINE1 (UniProt P05121), which was not measured in this study (31). |
| Complement compounds, CFs and immunoglobulins | Depending molecular species | As complement compounds, CFs and immunoglobulins are involved in the inflammatory processes, these differences suggest changes in the immune response pathways during steatosis development. Increased levels of C3 in grade 3 steatosis stimulates triglyceride synthesis and glucose transport in adipocytes, regulating fat storage and playing a role in postprandial triglyceride clearance (37,38). |
| AFM and VTN | Higher | These findings were consistent with results from murine studies on their protein equivalents (AFM60 and VTN)(39), and were retrieved as functional biomarkers of hepatic steatosis in humans (31). Therefore, a shared biology between mice and humans in hepatic steatosis development was suggested. |
| AZGP1 | Lower | AZGP1 is normally responsible for lipid degradation stimulation in adipocytes but is less abundant in steatosis participants, suggesting lipid metabolism dysfunction (i.e. more triglycerides). |
| HPR | Lower | HPR is the main plasma protein associated with APOL-I-containing high-density lipoprotein (HDL), so a downregulation of this protein suggests lipid metabolism dysfunction as well. |
| GSN | Lower | GSN is overexpressed in MASH patients and reported to have a protective role in MASH through F-actin regulation and P53 degradation (40). Though in this study, the protein abundance is lower in grade 3 steatosis. |
| GC, LRG1, SHBG, A1BG and CP | Lower | No direct links between these plasma proteins and liver steatosis have been reported so far. As most significant proteins were reported to be, among other functions, involved in the immune response pathways, a (low-grade) pro-inflammatory state is expected in the steatosis phenotype. |
| **Metabolites** |  |  |
| AAs and Biogenic amines | Depending on molecular species | Glutamate has also been reported to be positively associated with higher BMI (23). Branched AAs (Ile, Leu, Val) on the other hand have been reported to be decreased in patients with hepatic encephalopathy in advanced chronic liver disease (41). Furthermore, they are also associated with metabolic health (42) and with insulin resistance, having an increased concentration in blood of diabetic mice, rats, and humans (43). Alteration of amino acid concentrations, as well as changes in biogenic amine abundances, have been attributed and found associated with numerous different metabolically unhealthy phenotypes. These are therefore disregarded as potential molecular drivers specific for hepatic steatosis in humans. |
| H1 | Higher | The carbohydrate metabolism is affected by liver steatosis. A relationship of these hexose sugar with the higher proportion of T2D cases in the steatosis grade 3 category of the present cohort can however not be ruled out. |
| (Acyl)carnitines | Higher, except for C12-DC | Acylcarnitines are primarily involved in the beta oxidation of fatty acids, facilitating the transport of acetyl-CoA across mitochondrial membranes. Generally, dysregulation of acylcarnitine production indicates a dysregulated immune system or damage to the liver (44). |
| **Lipids** |  |  |
| PCs | Lower, except for PC aa C32:1 | PCs are key components of the lipid bilayer of cells, as well as being involved in metabolism and signaling, such as in the early stages of the inflammatory cascade (45). |
| LysoPCs | Lower | As lysoPCs have a number of anti-inflammatory effects, such as binding G protein-coupled lysophospholipid receptors. Decreased levels of lysoPCs have been observed in liver cirrhosis with increased mortality risk (46). Decreased lysoPCs have also been associated with pre-diabetes (47), obesity (48), and type 2 diabetes (49,50), although the underlying mechanisms linking these lipids with the disease progression are poorly understood. Similar mechanisms are expected to occur in grade 3 steatosis. |
| SMs | Lower | SMs are major components of low-density lipoprotein (LDL) particles, and were reported to be associated with increased risk of type 2 diabetes (49,51), although the association appears to be BMI dependent (52). The hydroxylated sphingomyelin has been cited as a pre-diagnostic circulating marker, reduced with higher colorectal cancer risk and higher in breast cancer patients (53,54). |

Although defined by different clinical criteria, the group of steatosis grade 3 participants was in our cohort largely overlapping the group of participants diagnosed with MASLD. This confirms and expands on an earlier report of circulating multi-protein signatures of hepatic steatosis being strongly associated with liver disease and multi-system metabolic outcomes (31). Similarly, also the proportion of T2D cases was higher in steatosis grade 3, but a sensitivity analysis argued against a strong influence of T2D on the present association results. The circulating proteome and metabolome association results might therefore not be exclusively related to a progression of steatosis, but reflect a more general transition from a metabolically healthy to unhealthy phenotype. Ultimately, the inherent relationship between the liver-health related phenotypes that include steatosis, fibrosis, MASLD and T2D makes the identification of clear, phenotype-specific, markers challenging.

As expected, a higher adherence to a plant-based diet was negatively associated with steatosis, but apart from this, no other relationship between dietary habits and steatosis was found in the present data set. The absent association of any fatty acid-related macronutrient intake with steatosis, and the independence of the circulating proteome and lipidome association results from the plant-based dietary index suggest that the observed changes in circulating molecules, such as higher triglyceride levels, most likely originate from secretion from the liver and not from the diet.

Given the individual variability in disease progression, with some patients progressing slowly from steatosis to MASH or fibrosis, and others doing so rapidly, there is a clinical need for predictive biomarkers that can identify individuals at risk. Our findings offer potential candidates, but longitudinal data and subsequent mechanistic validations are required. Moreover, the non-annotated metabolites identified in our untargeted analysis represent a largely unexplored space. Targeted follow-up studies, incorporating MS/MS validation and pathway analysis, could uncover novel pathways involved in hepatic dysfunction and systemic metabolic health.

Summarizing, our findings support the concept that the circulating biomolecular landscape of steatosis is not only influenced by hepatic pathology but is also shaped by systemic metabolic health. As such, distinguishing between different metabolically healthy and unhealthy states may provide a more mechanistically relevant framework for biomarker discovery than strictly dichotomous steatosis phenotyping alone.

### Limitations of the study

Despite the strengths of our cohort design, such as fasting-state sample collection, comprehensive multi-omic molecular phenotyping, liver health phenotyping as performed in clinical practice, and recruitment from a single demographically consistent population, several limitations must be acknowledged.

First, we relied exclusively on plasma and serum samples, limiting the resolution of tissue-specific insights. While most of the measured plasma proteins are known to originate from the liver, small molecules like lipids and metabolites have more diverse systemic sources. As such, their circulating levels may not accurately reflect liver-specific alterations. The inclusion of liver biopsies would allow deeper molecular profiling, but is, for ethical reasons, not possible in a (generally healthy) population cohort.

Another limitation is the inherent relationship between metabolic health-related phenotypes and diseases such as T2D or MASLD. Due to the complete or partial collinearity of these phenotypes, their influence and confounding could not be completely accounted for. Through sensitivity analyses, we tried to evaluate their potential influences, but this does not guarantee complete independence of the result.

Further, while robust and reliable state-of-the-art methodology was employed, only a limited set of plasma proteins was investigated. These comprise however important and relevant proteins for the present phenotype that are involved in nutrient transport, coagulation, and immune system activity. Also the metabolome coverage, like in most metabolomics studies, was limited in the present study lacking some potentially interesting metabolite classes, such as bile acids, which might also help to assess a potential contribution by the microbiome.

Biochemical pathway analysis in general suffers from a lack of completeness of biochemical pathways, both regarding specificity in the organism, tissue or cellular location, as well as incompleteness regarding regulatory/signaling/transport events. Interpretation of pathway analysis results presently requires extensive manual evaluation and expert know-how. This applies in particular to metabolites and lipids, which can originate either from food uptake or metabolism and biosynthesis, or even from microbial intermediate processes. Current pathway enrichment methods, designed originally for transcriptomics, are less useful for metabolomics and lipidomics data.

The dietary assessment in the present study is, like in most other cross-sectional or cohort studies, based entirely on FFQs. Such data can however be affected by recall errors and its temporal alignment with the fasting blood samples is unclear.

Finally, the untargeted metabolomics approach, while broad in scope, suffers from incomplete metabolite annotation. The absence of MS/MS data limited our ability to assign identities to a substantial portion of detected features. These non-annotated molecules may hold important biological relevance, particularly in steatosis progression, but further research and data would be required to uncover them.

## Conclusion

In conclusion, through a multi-omics approach we identified circulating proteins, metabolites and lipids associated with liver steatosis that present valuable insights into the biomolecular signatures of hepatic steatosis. Our study however also highlights the complexity of interpreting such data in the context of metabolic comorbidities. A shift toward understanding metabolic health as a continuum, supported by robust molecular phenotyping, may improve our capacity to identify individuals at risk and develop targeted interventions for hepatic steatosis, MASH or fibrosis and more generally MASLD.

## Data Availability

Data and samples can be requested for clearly defined research activities via the CHRIS Portal (https://chrisportal.eurac.edu/). Contact the corresponding author for details. Full results of the present analyses are provided in the supplementary documents. The data analysis workflows are available at https://github.com/EuracBiomedicalResearch/nafld_proteo_metabolomics.

## Acknowledgments

The CHRIS study thank all study participants, the Healthcare System of the Autonomous Province of Bolzano Bozen - South Tyrol, and all Eurac Research staff involved in the study (https://www.eurac.edu/chrisack). Bioresource Impact Factor Code: BRIF6107. The CHRIS study was funded by the Autonomous Province of Bolzano/Bozen - South Tyrol - Department of Innovation, Research, University and Museums and supported by the European Regional Development Fund (FESR1157).

## Author Contributions

Conceptualization, MDG, JR, CG; Data curation, MDG, JR; Formal analysis, MDG, ND, EH, PL, CP, HT, MR, JR, CG; Funding acquisition, CP, FSD, PPP, HT, JR, GC; Investigation, MDG, ND, EH, PL, VVH, JR, CG; Methodology, MDG, ND, EH, PL, VVH, CP, HT, MR, JR, CG; Project administration, MDG, JR, CG; Resources, MDG, ND, EH, PL, CP, FSD, MR, PPP, HT, MR, JR, CG; Software, MDG, EH, PL, JR; Supervision, EH, FSD, CP, JR, CG; Validation, MDG, EH, CP, JR, CG; Visualization, MDG, JR; Writing – original draft, MDG, EH, PP, JR, CG; Writing – review and editing, MDG, ND, EH, PL, VVH, CP, FSD, PPP, HT; MR, JR, CG. All authors have read and agreed to the published version of the manuscript.

## Institutional Review Board Statement

The Ethics Committee of the Healthcare System of the Autonomous Province of - Bolzano/Bozen – South Tyrol approved the CHRIS NAFLD protocol on 22 September 2016 (85–2016). The study conforms to the Declaration of Helsinki, and with national and institutional legal and ethical requirements.

## Informed Consent Statement

Informed consent was obtained from all subjects involved in the study.

