## supplementary_figures for "Molecular characterization of liver steatosis through circulating proteins, metabolites and lipids" (from another version of this preprint: version 1, 2025-12-02)

**Title**

**Short Title**

Blood-based profiling of liver steatosis

**Authors**

Marilyn De Graeve[1], Essi Hantikainen[1], Philippine Louail[1], Vinicius Verri Hernandes[1,2], Cristian Pattaro[1], Francisco S. Domingues[1], Peter P. Pramstaller[1], Herbert Tilg[3], Markus Ralser[4,5,6], Christoph Grander[3,*], Johannes Rainer[1,*]

**Affiliations**

[1]Institute for Biomedicine, Eurac Research, 39100 Bolzano, Italy

[2]Faculty of Chemistry, Department of Food Chemistry and Toxicology, University of Vienna, Währinger Straße 38, 1090 Vienna, Austria

[3]Department of Internal Medicine I, Gastroenterology, Hepatology, Endocrinology & Metabolism, Medical University Innsbruck, 6020 Innsbruck, Austria.

[4]Institute of Biochemistry, Charité Universitätsmedizin Berlin, 10117 Berlin, Germany

[5]Max Planck Institute for Molecular Genetics, 14195 Berlin, Germany

[6]The Wellcome Centre for Human Genetics, Nuffield Department of Medicine, University of Oxford, Oxford OX3 7BN, UK

* Contributed equally

**Corresponding author**

#### Supplementary Figures

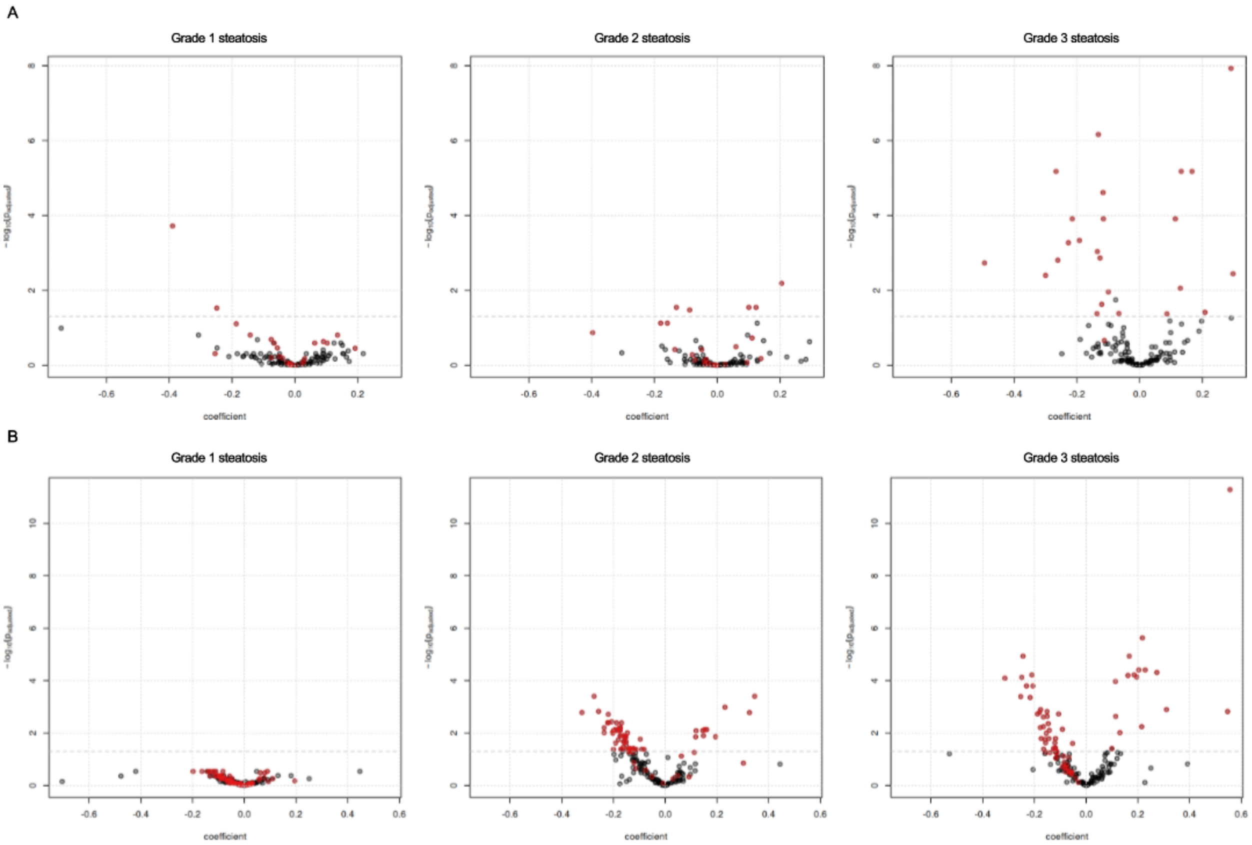


Figure S1. Results from the association analysis for proteins (A) and metabolites and lipids (B) visualized as Volcano plots. Shown are differences in abundance on the x-axis against its significance on the y-axis. The coefficient (x-axis) represents the log2 difference in average abundance between individuals with different steatosis grade and controls, positive values indicating higher abundance in steatosis, negative values higher abundance in control individuals. Red color depicts significant proteins and metabolites, respectively.

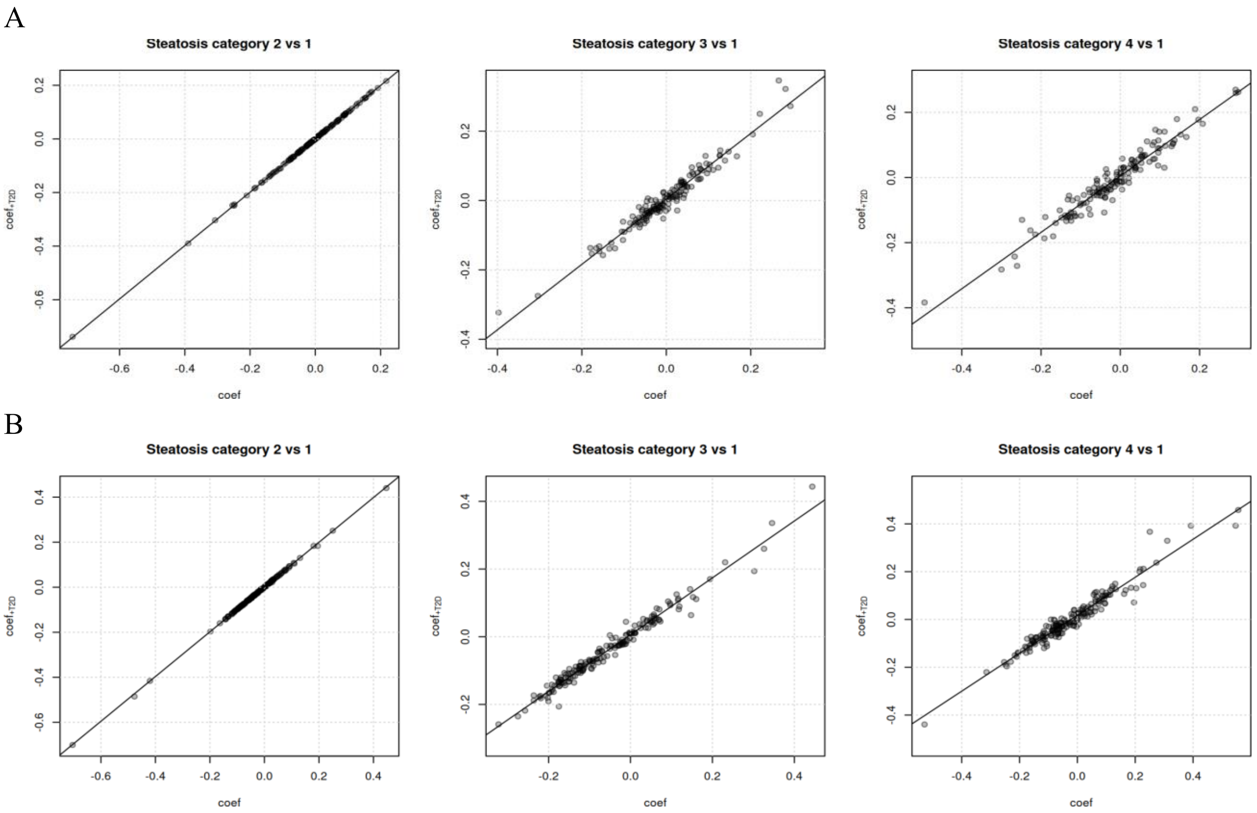


Figure S2. Sensitivity analysis comparing the coefficients from the association analysis for proteins (A) and metabolites and lipids (B) with (y-axis) and without (x-axis) T2D adjustment.

A

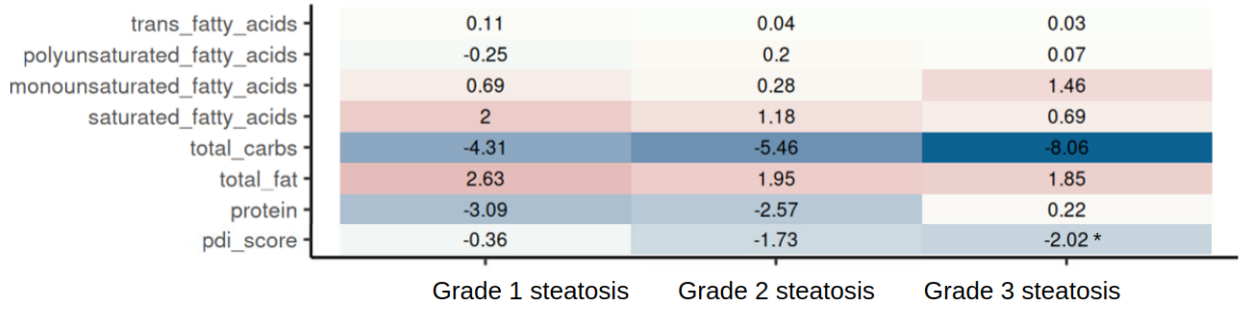


B

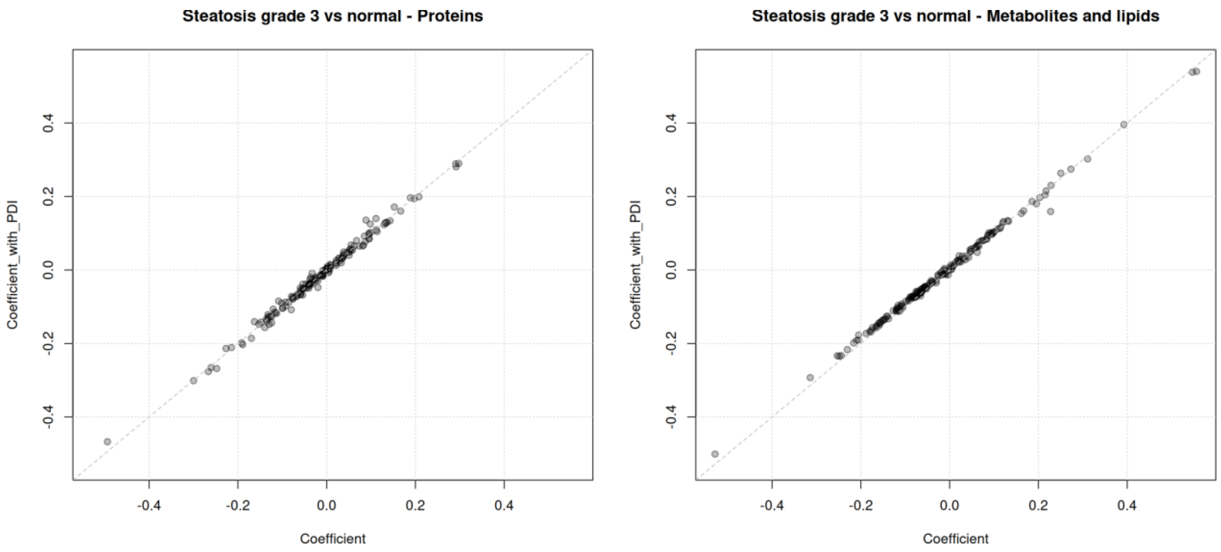


Figure S3. Effect of the dietary intake on the steatosis characterization. Hierarchical clustered heatmap depicting trends among the control versus different steatosis grade comparisons of dietary intake variables. Asterisks indicate significantly altered variables. The coefficients for each variable of the control vs steatosis grades comparison were shown, positive values indicating higher abundance in steatosis, negative values higher abundance in control individuals (A). Comparison of coefficients with and without PDI adjustment in the grade 3 steatosis association analysis for proteins, and metabolites and lipids, respectively (B).

A

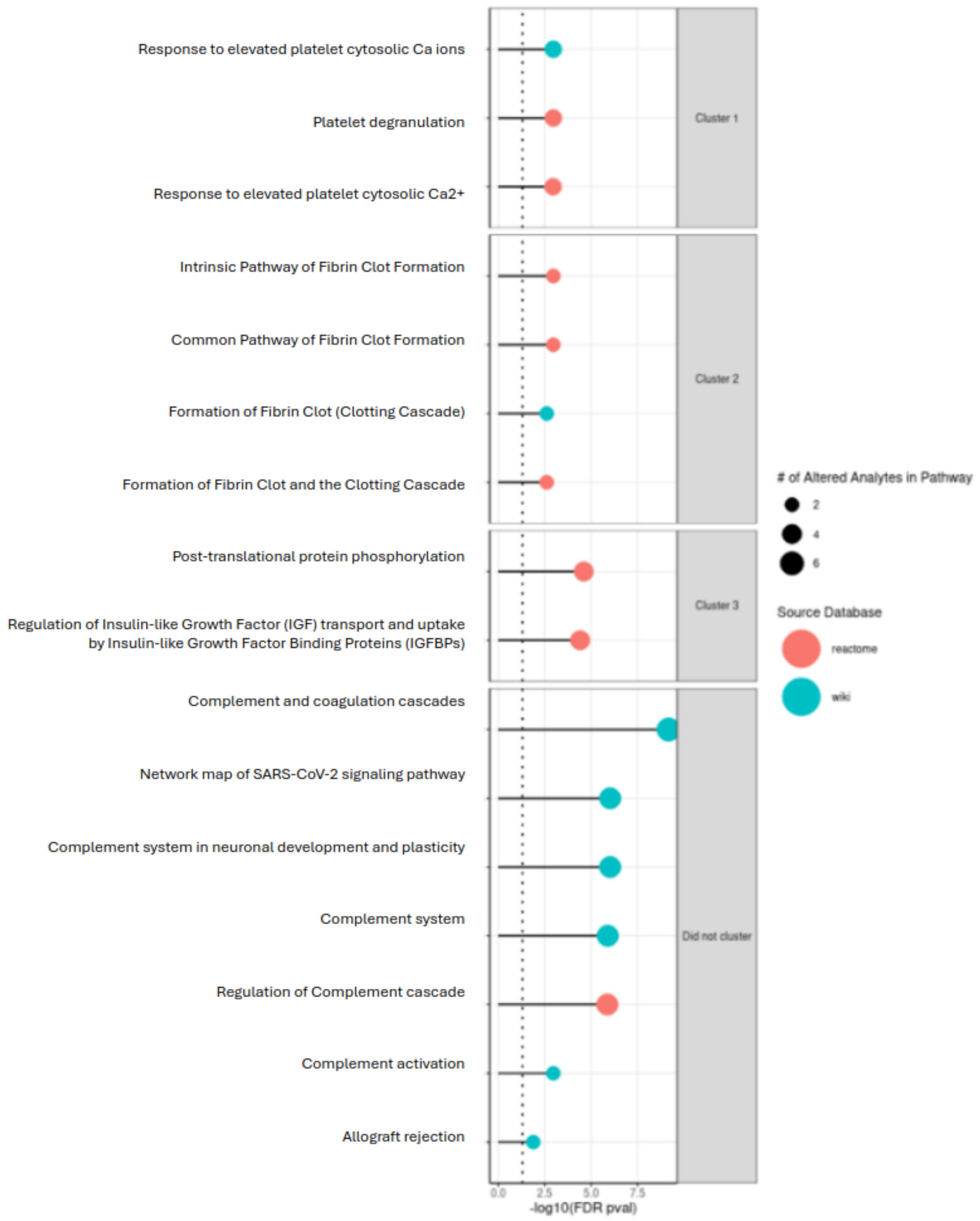
Response to elevated platelet cytosolic Ca ions
Platelet degranulation
Response to elevated platelet cytosolic Ca2+
Intrinsic Pathway of Fibrin Clot Formation
Common Pathway of Fibrin Clot Formation
Formation of Fibrin Clot (Clotting Cascade)
Formation of Fibrin Clot and the Clotting Cascade
Post-translational protein phosphorylation
Regulation of Insulin-like Growth Factor (IGF) transport and uptake by Insulin-like Growth Factor Binding Proteins (IGFBPs)
Complement and coagulation cascades
Network map of SARS-CoV-2 signaling pathway
Complement system in neuronal development and plasticity
Complement system
Regulation of Complement cascade
Complement activation
Allograft rejection
Cluster 1
Cluster 2
Cluster 3
Did not cluster
0.0
2.5
5.0
7.5
-log10(FDR pval)
### of Altered Analytes in Pathway
2
4
6
Source Database
reactome
wiki

B

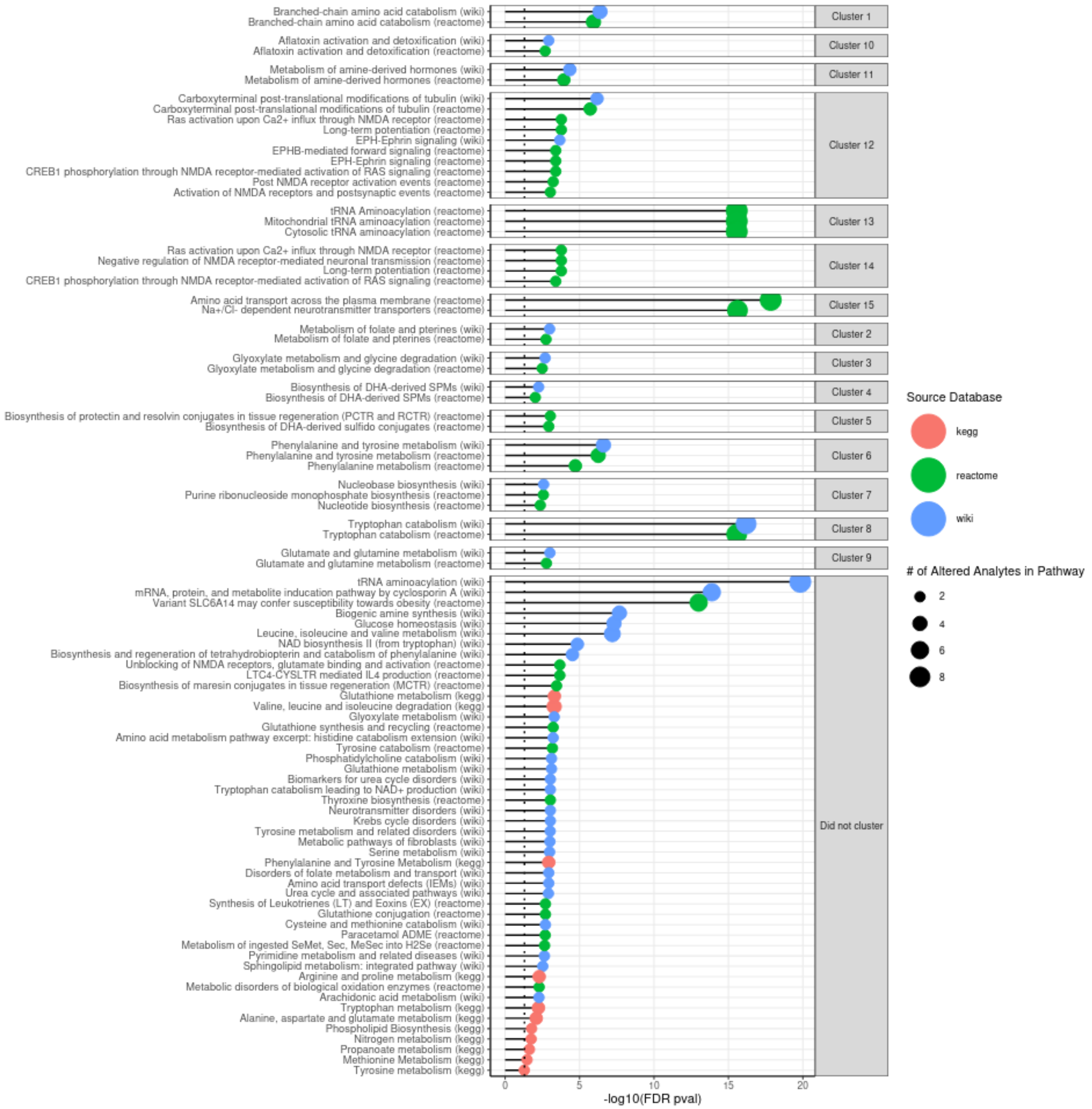


Figure S4. Pathway enrichment analysis using Ramp-DB, original not manually curated output of proteins (A) and metabolites and lipids (B). There is no overlap in affected pathways between proteins and metabolites. The Phosphatidylcholine catabolism (wiki) and phospholipid biosynthesis (KEGG) representing the significantly altered PCs, belong to the "Did not cluster" cluster, but are not depicted on the figure due to a maximum of 100 input identifiers on Ramp-DB.
