## supplementary_table_captions for "Molecular characterization of liver steatosis through circulating proteins, metabolites and lipids" (from another version of this preprint: version 1, 2025-12-02)

Table S1. Relatively quantified Scanning SWATH proteomics data. Alternative names of the proteins and associated genes can be consulted at https://www.uniprot.org.

Table S2. Absolutely quantified metabolites and lipids. Alternative names of the analytes can be consulted with the provider (Biocrates AbsoluteIDQ® p180 kit).

Table S3. Significant proteins when comparing control (n=140, CAP<238 dB/m) versus different grades of steatosis: grade 1 (n=52, CAP>238 dB/m and CAP>260 dB/m), grade 2 (n=54, CAP>260 dB/m and CAP<290 dB/m), grade 3 steatosis (n=81, CAP>290 dB/m) using linear regression analysis with age and sex as covariates. $CV_{QC}$, coefficient of variance of pooled quality control samples from the CHRIS study. Alternative nomenclature can be consulted on UniProt (https://www.uniprot.org/). org/). Formula: protein ~ sex + age/10 + steatosis grade.

Table S4. Significant metabolites and lipids when comparing control (n=140, CAP<238 dB/m) versus different grades of steatosis: grade 1 (n=52, CAP>238 dB/m and CAP>260 dB/m), grade 2 (n=54, CAP>260 dB/m and CAP<290 dB/m), grade 3 steatosis (n=81, CAP>290 dB/m) using linear regression analysis with age and sex as covariates. $CV_{QC}$, coefficient of variance of pooled quality control samples from the CHRIS study. Alternative nomenclature can be consulted with the provider (Biocrates p180 kit). Formula: metabolite/lipid ~ sex + age/10 + steatosis grade.

Table S5. Dietary intake variables when comparing control (n=140, CAP<238 dB/m) versus grade 3 steatosis (n=81, CAP>290 dB/m) using linear regression analysis with age, sex and estimated daily total energy intake as covariates. Asterisks, variables with a confidence interval outside of zero. PDI, Plant-based diet index. Formula: macronutrient/PDI ~ sex + age/10 + steatosis grade + estimated daily total energy intake.

Table S6. Pathway names for significantly altered proteins, metabolites and lipids between control and grade 3 steatosis.
