## supplementary_table1 for "Molecular characterization of liver steatosis through circulating proteins, metabolites and lipids" (from another version of this preprint: version 1, 2025-12-02)

| Uniprot ID | Description | PeptideN | eptideNam | Protein.Id | rotein.Nam | Gene | dified.Sequ | pped.Sequ | Precursor.Id |
|---|---|---|---|---|---|---|---|---|---|
| P02768 | Serum albumin | 187 | AAC(UniM | P02768:H | ALBU HU | ALB | AAC(UniM | AACLLPK | [illegible] |
| P02766 | Transthyretin | 4 | AADDTW | P02766:A | TTHY HU | TTR | AADDTW | AADDTW | [illegible] |
| P19827 | Inter-alpha-trypsin inhibitor heavy chain H1 | 36 | AAISGEN | P19827:F | ITIH1 HU | ITIH1 | AAISGEN | AAISGEN | [illegible] |
| P01023 | Alpha-2-macroglobulin | 119 | AAQVTIQ | P01023+P | A2MG HU | A2M | AAQVTIQ | AAQVTIQ | [illegible] |
| P01042;P01042-2 | Kininogen-1 | 25 | AATGEC( | P01042:P | KNG1 HU | KNG1 | AATGEC( | AATGECT | [illegible] |
| P02649 | Apolipoprotein E | 27 | AATVGSL | P02649:H | APOE HU | APOE | AATVGSL | AATVGSL | [illegible] |
| P01024 | Complement C3 | 178 | AAVYHH | P01024+P | CO3 HUN | C3 | AAVYHH | AAVYHH | [illegible] |
| P04196 | Histidine-rich glycoprotein | 21 | ADLFYD | P04196 | HRG HU | HRG | ADLFYD | ADLFYD | [illegible] |
| P01011 | Alpha-1-antichymotrypsin | 25 | ADLSGIT | P01011;G | AACT HU | SERPINA3 | ADLSGIT | ADLSGIT | [illegible] |
| P02787 | Serotransferrin | 130 | ADRDQY | P02787;C | TRFE HU | TF | ADRDQY | ADRDQY | [illegible] |
| P01834 | Immunoglobulin kappa constant | 19 | ADYEKH | P01834 | IGKC HU | IGKC | ADYEKH | ADYEKH | [illegible] |
| Q14624;Q14624-2 | Inter-alpha-trypsin inhibitor heavy chain H4 | 52 | AEAQAQ | Q14624;Q | ITIH4 HU | ITIH4 | AEAQAQ | AEAQAQ | [illegible] |
| P07360 | Complement component C8 gamma chain | 6 | AEATTLH | P07360:O | CO8G HU | C8G | AEATTLH | AEATTLH | [illegible] |
| P00450 | Ceruloplasmin | 49 | AEEEHLC | P00450:H | CERU HU | CP | AEEEHLC | AEEEHLC | [illegible] |
| P02647 | Apolipoprotein A-I | 69 | AELQEG | P02647:F | APOA1 H | APOA1 | AELQEG | AELQEG | [illegible] |
| P02760 | Protein AMBP | 5 | AFIQLWA | P02760:S | AMBP HU | AMBP | AFIQLWA | AFIQLWA | [illegible] |
| P01008 | Antithrombin-III | 39 | AFLEVNE | P01008:O | ANT3 HU | SERPINC1 | AFLEVNE | AFLEVNE | [illegible] |
| O95445 | Apolipoprotein M | 2 | AFLLTPR | O95445:O | APOM HU | APOM | AFLLTPR | AFLLTPR | AFLLTPR2+SLTSC(UniMod:4)LDSK2 |
| P01031 | Complement C5 | 20 | AFTEC(Ui | P01031 | CO5 HUN | C5 | AFTEC(Ui | AFTECCV | [illegible] |
| P01042-2 | Isoform LMW of Kininogen-1 | 3 | AGAEPAS | P01042-2; | KNG1 HU | KNG1 | AGAEPAS | AGAEPAS | [illegible] |
| P06396;P06396-2 | Gelsolin | 22 | AGALNSN | P06396;P | GELS HU | GSN | AGALNSN | AGALNSN | [illegible] |
| P25311 | Zinc-alpha-2-glycoprotein | 15 | AGEVQEI | P25311+P | ZA2G HU | AZGP1 | AGEVQEI | AGEVQEI | [illegible] |
| P22792 | Carboxypeptidase N subunit 2 | 2 | AGGSWD | P22792 | CPN2 HU | CPN2 | AGGSWD | AGGSWD | [illegible] |
| Q96PD5 | N-acetylmuramoyl-L-alanine amidase | 9 | AGLLRPD | Q96PD5;Q | PGRP2 H | PGLYRP2 | AGLLRPD | AGLLRPD | [illegible] |
| P19823 | Inter-alpha-trypsin inhibitor heavy chain H2 | 46 | AHVSFKF | P19823;Q | ITIH2 HU | ITIH2 | AHVSFKF | AHVSFKF | [illegible] |
| P02675 | Fibrinogen beta chain | 82 | AHYGGF | P02675:D | FIBB HU | FGB | AHYGGF | AHYGGF | [illegible] |
| P02748 | Complement component C9 | 13 | AIEDYINI | P02748 | CO9 HUN | C9 | AIEDYINI | AIEDYINI | [illegible] |
| P05155;P05155-3 | Plasma protease C1 inhibitor | 14 | AIMEKLE | P05155:P | IC1 HUM | SERPING1 | AIMEKLE | AIMEKLE | [illegible] |
| P43652 | Afamin | 16 | AIPVTQY | P43652 | AFAM HU | AFM | AIPVTQY | AIPVTQY | [illegible] |
| P02679;P02679-2 | Fibrinogen gamma chain | 44 | AIQLTYN | P02679:P | FIBG HU | FGG | AIQLTYN | AIQLTYN | [illegible] |
| P01861 | Immunoglobulin heavy constant gamma 4 | 11 | AKGQPRI | P01861:A | IGHG4 H | IGHG4 | AKGQPRI | AKGQPRI | [illegible] |
| P06727 | Apolipoprotein A-IV | 52 | AKIDQN | P06727 | APOA4 H | APOA4 | AKIDQN | AKIDQN | [illegible] |
| O14791;O14791-2;O14791-3 | Apolipoprotein L1 | 6 | ALADGV | O14791;O | APOL1 H | APOL1 | ALADGV | ALADGV | [illegible] |
| P00751 | Complement factor B | 35 | ALFVSEE | P00751;E | CFAB HU | CFB | ALFVSEE | ALFVSEE | [illegible] |
| P02750 | Leucine-rich alpha-2-glycoprotein | 7 | ALGHLDI | P02750 | A2GL HU | LRG1 | ALGHLDI | ALGHLDI | [illegible] |
| P20851;P20851-2 | C4b-binding protein beta chain | 1 | ALLAFQE | P20851;P | C4BPB H | C4BPB | ALLAFQE | ALLAFQE | ALLAFQESK2 |
| P13671 | Complement component C6 | 4 | ALNHLPL | P13671 | CO6 HUN | C6 | ALNHLPL | ALNHLPL | [illegible] |
| P01019 | Angiotensinogen | 8 | ALQDQL | P01019 | ANGT HU | AGT | ALQDQL | ALQDQL | [illegible] |
| P35858;P35858-2 | Insulin-like growth factor-binding protein complex acid labile subunit | 4 | ALRDFAI | P35858:P | ALS HUN | IGFALS | ALRDFAI | ALRDFAI | [illegible] |
| P02671 | Fibrinogen alpha chain | 85 | ALTDMP | P02671:P | FIBA HU | FGA | ALTDMP | ALTDMP | [illegible] |
| P04114 | Apolipoprotein B-100 | 106 | ALVDTLK | P04114+P | APOB HU | APOB | ALVDTLK | ALVDTLK | [illegible] |
| P36955 | Pigment epithelium-derived factor | 11 | ALYYDLI | P36955:I | PEDF HU | SERPINF1 | ALYYDLI | ALYYDLI | [illegible] |
| P07357 | Complement component C8 alpha chain | 5 | AM(UniM | P07357 | CO8A HU | C8A | AM(UniM | AMAVED | [illegible] |
| P02751 | Fibronectin | 27 | AQITGYR | P02751:P | FINC HU | FN1 | AQITGYR | AQITGYR | [illegible] |
| P02765 | Alpha-2-HS-glycoprotein | 15 | AQLVPLP | P02765:C | FETUA H | AHSG | AQLVPLP | AQLVPLP | [illegible] |
| P00488 | Coagulation factor XIII A chain | 2 | AQMDLS | P00488 | F13A HU | F13A1 | AQMDLS | AQMDLS | [illegible] |
| A0A0C4DH29 | Immunoglobulin heavy variable 1-3 | 1 | ASGYTFT | A0A0C4D | HV103 H | IGHV1-3 | ASGYTFT | ASGYTFT | [illegible] |
| P01602 | Immunoglobulin kappa variable 1-5 | 3 | ASQSISSV | P01602 | KV105 H | IGKV1-5 | ASQSISSV | ASQSISSV | [illegible] |
| P01619 | Immunoglobulin kappa variable 3-20 | 3 | ASQSVSS | P01619 | KV320 H | IGKV3-20 | ASQSVSS | ASQSVSS | [illegible] |
| P10909;P10909-2;P10909-4;P10909-5 | Clusterin | 23 | ASSIIDEL | P10909;P | CLUS HU | CLU | ASSIIDEL | ASSIIDEL | [illegible] |
| P00746 | Complement factor D | 2 | ATLGPAV | P00746;K | CFAD HU | CFD | ATLGPAV | ATLGPAV | [illegible] |
| P15814 | Immunoglobulin lambda-like polypeptide 1 | 2 | ATPSVTL | P15814;C | IGLL1 HU | IGLL1 | ATPSVTL | ATPSVTL | [illegible] |
| P00747 | Plasminogen | 17 | ATTVTGT | P00747+P | PLMN HU | PLG | ATTVTGT | ATTVTGT | [illegible] |
| P08571 | Monocyte differentiation antigen CD14 | 1 | ATVNPSA | P08571 | CD14 HU | CD14 | ATVNPSA | ATVNPSA | ATVNPSAPR2 |
| P02749 | Beta-2-glycoprotein 1 | 7 | ATVVYQ | P02749+P | APOH HU | APOH | ATVVYQ | ATVVYQ | [illegible] |
| P04217;P04217-2 | Alpha-1B-glycoprotein | 13 | ATWSGA | P04217:P | A1BG HU | A1BG | ATWSGA | ATWSGA | [illegible] |
| P00738 | Haptoglobin | 27 | AVGDKLI | P00738:J | HPT HUN | HP | AVGDKLI | AVGDKLI | [illegible] |
| P06681 | Complement C2 | 2 | AVISPGFI | P06681:P | CO2 HUN | C2 | AVISPGFI | AVISPGFI | [illegible] |
| P05543 | Thyroxine-binding globulin | 5 | AVLHIGE | P05543 | THBG HU | SERPINA7 | AVLHIGE | AVLHIGE | [illegible] |
| P01009 | Alpha-1-antitrypsin | 61 | AVLTIDE | P01009;A | A1AT HU | SERPINA1 | AVLTIDE | AVLTIDE | [illegible] |
| P04004 | Vitronectin | 23 | AVRPGYI | P04004+P | VTNC HU | VTN | AVRPGYI | AVRPGYI | [illegible] |
| P05154 | Plasma serine protease inhibitor | 1 | AVVEVDI | P05154;G | IPSP HUN | SERPINA5 | AVVEVDI | AVVEVD | [illegible] |
| P01859 | Immunoglobulin heavy constant gamma 2 | 15 | C(UniMod | P01859;A | IGHG2 H | IGHG2 | C(UniMod | CCVECPI | [illegible] |
| P08603 | Complement factor H | 39 | C(UniMod | P08603+P | CFAH HU | CFH | C(UniMod | CFEGFGI | [illegible] |
| P04003 | C4b-binding protein alpha chain | 26 | C(UniMod | P04003+P | C4BPA H | C4BPA | C(UniMod | CHPGYKI | [illegible] |
| P01860 | Immunoglobulin heavy constant gamma 3 | 18 | C(UniMod | P01860:A | IGHG3 H | IGHG3 | C(UniMod | CPAPELL | [illegible] |
| P02790 | Hemopexin | 34 | C(UniMod | P02790+P | HEMO HU | HPX | C(UniMod | CSPHLVL | [illegible] |
| P01591 | Immunoglobulin J chain | 6 | C(UniMod | P01591:D | IGJ HUM | JCHAIN | C(UniMod | CYTAVVI | [illegible] |
| P02656 | Apolipoprotein C-III | 7 | DALSSVC | P02656:B | APOC3 H | APOC3 | DALSSVC | DALSSVC | [illegible] |
| P01877 | Immunoglobulin heavy constant alpha 2 | 5 | DASGATF | P01877:A | IGHA2 H | IGHA2 | DASGATF | DASGATF | [illegible] |
| P01876 | Immunoglobulin heavy constant alpha 1 | 7 | DASGVTF | P01876:A | IGHA1 H | IGHA1 | DASGVTF | DASGVTF | [illegible] |
| P01857 | Immunoglobulin heavy constant gamma 1 | 13 | DELTKN( | P01857;A | IGHG1 H | IGHG1 | DELTKN( | DELTKN( | [illegible] |
| P01871 | Immunoglobulin heavy constant mu | 39 | DGFFGNI | P01871;P | IGHM HU | IGHM | DGFFGNI | DGFFGNI | [illegible] |
| P00734 | Prothrombin | 25 | DKLAAC( | P00734;C | THRB HU | F2 | DKLAAC( | DKLAACI | [illegible] |
| A0A0J9YX35 | Immunoglobulin heavy variable 3-64D | 3 | DNSKNTI | A0A0J9Y | HV64D H | IGHV3-64D | DNSKNTI | DNSKNTI | [illegible] |
| P02753 | Retinol-binding protein 4 | 9 | DPNGLPF | P02753;Q | RET4 HU | RBP4 | DPNGLPF | DPNGLPF | [illegible] |
| P02746 | Complement C1q subcomponent subunit B | 7 | DQTIRFD | P02746;A | C1QB HU | C1QB | DQTIRFD | DQTIRFD | [illegible] |
| P08697 | Alpha-2-antiplasmin | 11 | DSFHLDE | P08697:P | A2AP HU | SERPINF2 | DSFHLDE | DSFHLDE | [illegible] |
| P06310 | Immunoglobulin kappa variable 2-30 | 3 | DSGVPDI | P06310 | KV230 H | IGKV2-30 | DSGVPDI | DSGVPDI | [illegible] |
| P0C0L4 | Complement C4-A | 2 | DSSTWLT | P0C0L4:P | CO4A HU | C4A | DSSTWLT | DSSTWLT | [illegible] |
| P03952 | Plasma kallikrein | 9 | DSVTGTL | P03952:C | KLKB1 H | KLKB1 | DSVTGTL | DSVTGTL | [illegible] |
| P07358 | Complement component C8 beta chain | 5 | DTMVED | P07358:F | CO8B HU | C8B | DTMVED | DTMVED | [illegible] |
| P02654 | Apolipoprotein C-I | 5 | EFGNTLE | P02654:K | APOC1 H | APOC1 | EFGNTLE | EFGNTLE | [illegible] |
| P02774;P02774-3 | Vitamin D-binding protein | 26 | EFSHLGK | P02774:P | VTDB HU | GC | EFSHLGK | EFSHLGK | [illegible] |
| P68871 | Hemoglobin subunit beta | 7 | EFTPPVQ | P68871+P | HBB HUN | HBB | EFTPPVQ | EFTPPVQ | [illegible] |
| P23142 | Fibulin-1 | 1 | EFTRPEE | P23142 | FBLN1 H | FBLN1 | EFTRPEE | EFTRPEE | EFTRPEEIIFLR3 |
| Q9HCU4 | Cadherin EGF LAG seven-pass G-type receptor 2 | 1 | EGHVM(U | Q9HCU4 | CELR2 H | CELSR2 | EGHVM(U | EGHVML | [illegible] |
| Q16610;Q16610-4 | Extracellular matrix protein 1 | 2 | ELLALIQI | Q16610;Q | ECM1 HU | ECM1 | ELLALIQI | ELLALIQI | [illegible] |
| P02652 | Apolipoprotein A-II | 8 | EQLTPLIK | P02652;V | APOA2 H | APOA2 | EQLTPLIK | EQLTPLIK | [illegible] |
| P00748 | Coagulation factor XII | 5 | EQPPSLT | P00748 | FA12 HU | F12 | EQPPSLT | EQPPSLT | [illegible] |
| P27169 | Serum paraoxonase/arylesterase 1 | 14 | EVQPVEI | P27169;F | PON1 HU | PON1 | EVQPVEI | EVQPVEI | [illegible] |
| P80108 | Phosphatidylinositol-glycan-specific phospholipase D | 6 | FGSSLITV | P80108 | PHLD HU | GPLD1 | FGSSLITV | FGSSLITV | [illegible] |
| P51884 | Lumican | 9 | FNALQYI | P51884 | LUM HU | LUM | FNALQYI | FNALQYI | [illegible] |
| P02747 | Complement C1q subcomponent subunit C | 4 | FQSVFTV | P02747 | C1QC HU | C1QC | FQSVFTV | FQSVFTV | [illegible] |
| A0A075B6I0 | Immunoglobulin lambda variable 8-61 | 4 | FSGSILGN | A0A075B | LV861 HU | IGLV8-61 | FSGSILGN | FSGSILGN | [illegible] |
| A0A075B6H9 | Immunoglobulin lambda variable 4-69 | 1 | FSGSSSG | A0A075B | LV469 HU | IGLV4-69 | FSGSSSG | FSGSSSG | FSGSSSGAER2 |
| P05546 | Heparin cofactor 2 | 12 | FTVDRPF | P05546 | HEP2 HU | SERPIND1 | FTVDRPF | FTVDRPF | [illegible] |
| P22352 | Glutathione peroxidase 3 | 2 | FYTFLK2 | P22352;A | GPX3 HU | GPX3 | FYTFLK+ | FYTFLK+ | [illegible] |
| P06396 | Gelsolin | 1 | GASQAG | P06396 | GELS HU | GSN | GASQAG | GASQAG | [illegible] |
| P09871 | Complement C1s subcomponent | 9 | GFQVVV | P09871;H | C1S HUN | C1S | GFQVVV | GFQVVV | [illegible] |
| P05156 | Complement factor I | 6 | GLETSLA | P05156;E | CFAI HU | CFI | GLETSLA | GLETSLA | [illegible] |
| A0A0B4J1U7 | Immunoglobulin heavy variable 6-1 | 1 | GLEWLGI | A0A0B4J1 | HV601 H | IGHV6-1 | GLEWLGI | GLEWLGI | GLEWLGR2 |
| A0A0J9YXX1 | Immunoglobulin heavy variable 5-10-1 | 1 | GLEWMG | A0A0J9Y | HV5X1 H | IGHV5-10-1 | GLEWMG | GLEWMG | GLEWMGR2 |
| P01780 | Immunoglobulin heavy variable 3-7 | 2 | GLEWVA | P01780 | HV307 H | IGHV3-7 | GLEWVA | GLEWVA | [illegible] |
| A0A0A0MS15 | Immunoglobulin heavy variable 3-49 | 2 | GLEWVG | A0A0A0M | HV349 H | IGHV3-49 | GLEWVG | GLEWVG | [illegible] |
| P00736 | Complement C1r subcomponent | 7 | GLTLHLK | P00736:B | C1R HUN | C1R | GLTLHLK | GLTLHLK | [illegible] |
| Q06033;Q06033-2 | Inter-alpha-trypsin inhibitor heavy chain H3 | 3 | GMTNINI | Q06033:O | ITIH3 HU | ITIH3 | GMTNINI | GMTNINI | [illegible] |
| P08519 | Apolipoprotein(a) | 1 | GTYSTTV | P08519:A | APOA HU | LPA | GTYSTTV | GTYSTTV | GTYSTTVTGR2 |
| P08185 | Corticosteroid-binding globulin | 3 | HLVALSP | P08185:G | CBG HUN | SERPINA6 | HLVALSP | HLVALSP | [illegible] |
| P04217 | Alpha-1B-glycoprotein | 9 | HQFLLTG | P04217;M | A1BG HU | A1BG | HQFLLTG | HQFLLTG | [illegible] |
| P04278 | Sex hormone-binding globulin | 1 | IALGGLLI | P04278;P | SHBG HU | SHBG | IALGGLLI | IALGGLLI | [illegible] |
| P15169 | Carboxypeptidase N catalytic chain | 4 | IHILPSMN | P15169 | CBPN HU | CPN1 | IHILPSMN | IHILPSMN | [illegible] |
| P05090 | Apolipoprotein D | 10 | IKVLNQE | P05090;C | APOD HU | APOD | IKVLNQE | IKVLNQE | [illegible] |
| P10643 | Complement component C7 | 5 | ILPLTVC( | P10643 | CO7 HUN | C7 | ILPLTVC( | ILPLTVCI | [illegible] |
| Q14624 | Inter-alpha-trypsin inhibitor heavy chain H4 | 4 | IPKPEASI | Q14624+C | ITIH4 HU | ITIH4 | IPKPEASI | IPKPEASI | [illegible] |
| P02763 | Alpha-1-acid glycoprotein 1 | 11 | IPKSDVV | P02763 | A1AG1 H | ORM1 | IPKSDVV | IPKSDVV | [illegible] |
| P20742 | Pregnancy zone protein | 1 | IQHPFTV | P20742;P | PZP HUN | PZP | IQHPFTV | IQHPFTV | [illegible] |
| P18428 | Lipopolysaccharide-binding protein | 1 | ITDKGLQ | P18428 | LBP HUN | LBP | ITDKGLQ | ITDKGLQ | [illegible] |
| O43866 | CD5 antigen-like | 5 | IWLDNVI | O43866 | CD5L HU | CD5L | IWLDNVI | IWLDNVI | [illegible] |
| P02745 | Complement C1q subcomponent subunit A | 1 | KGHIYQC | P02745 | C1QA HU | C1QA | KGHIYQC | KGHIYQC | [illegible] |
| P07225 | Vitamin K-dependent protein S | 9 | KVESELII | P07225;G | PROS HU | PROS1 | KVESELII | KVESELII | [illegible] |
| P29622 | Kallistatin | 4 | LGFTDLF | P29622 | KAIN HU | SERPINA4 | LGFTDLF | LGFTDLF | [illegible] |
| O75636 | Ficolin-3 | 3 | LLGEVDI | O75636:O | FCN3 HU | FCN3 | LLGEVDI | LLGEVDI | [illegible] |
| P01701 | Immunoglobulin lambda variable 1-51 | 1 | LLIYDNN | P01701 | LV151 HU | IGLV1-51 | LLIYDNN | LLIYDNN | [illegible] |
| P01703 | Immunoglobulin lambda variable 1-40 | 1 | LLIYGNSI | P01703 | LV140 HU | IGLV1-40 | LLIYGNSI | LLIYGNSI | [illegible] |
| P06312 | Immunoglobulin kappa variable 4-1 | 3 | LLIYWAS | P06312 | KV401 H | IGKV4-1 | LLIYWAS | LLIYWAS | [illegible] |
| P01705 | Immunoglobulin lambda variable 2-23 | 1 | LMIYEGS | P01705 | LV223 HU | IGLV2-23 | LMIYEGS | LMIYEGS | LMIYEGSK2 |
| P49908 | Selenoprotein P | 1 | LPTDSEL | P49908;A | SEPP1 HU | SELENOP | LPTDSEL | LPTDSEL | LPTDSELAPR2 |
| O75882-2 | Isoform 2 of Attractin | 2 | LTLTPWV | O75882;O | ATRN HU | ATRN | LTLTPWV | LTLTPWV | [illegible] |
| Q9UGM5 | Fetuin-B | 1 | LVVLPFP | Q9UGM5: | FETUB H | FETUB | LVVLPFP | LVVLPFP | LVVLPFPK2 |
| P05452 | Tetranectin | 1 | MFEELK2 | P05452:E | TETN HU | CLEC3B | MFEELK | MFEELK | MFEELK2 |
| A0A0B4J1V2 | Immunoglobulin heavy variable 2-26 | 1 | MGVSWII | A0A0B4J1 | HV226 H | IGHV2-26 | MGVSWII | MGVSWII | MGVSWIR2 |
| P00742 | Coagulation factor X | 2 | MLEVPY | P00742 | FA10 HU | F10 | MLEVPY | MLEVPY | [illegible] |
| P00739 | Haptoglobin-related protein | 6 | NYAEVGI | P00739:P | HPTR HU | HPR | NYAEVGI | NYAEVG | [illegible] |
| P01599 | Immunoglobulin kappa variable 1-17 | 1 | RLIYAAS | P01599 | KV117 H | IGKV1-17 | RLIYAAS | RLIYAAS | [illegible] |
| P00740 | Coagulation factor IX | 1 | SALVLQY | P00740:P | FA9 HUN | F9 | SALVLQY | SALVLQY | SALVLQYLR2 |
| P19652 | Alpha-1-acid glycoprotein 2 | 4 | SDVMYTI | P19652 | A1AG2 H | ORM2 | SDVMYTI | SDVMYTI | [illegible] |
| P05160 | Coagulation factor XIII B chain | 2 | SFYFPMS | P05160 | F13B HU | F13B | SFYFPMS | SFYFPMS | [illegible] |
| A0A075B6K4 | Immunoglobulin lambda variable 3-10 | 1 | SGQAPVI | A0A075B | LV310 HU | IGLV3-10 | SGQAPVI | SGQAPVI | [illegible] |
| A0A0C4DH31 | Immunoglobulin heavy variable 1-18 | 1 | SLRSDDT | A0A0C4D | HV118 H | IGHV1-18 | SLRSDDT | SLRSDDT | [illegible] |
| P80748 | Immunoglobulin lambda variable 3-21 | 1 | SVHWYQ | P80748 | LV321 HU | IGLV3-21 | SVHWYQ | SVHWYQ | [illegible] |
| A0A0B4J1Y9 | Immunoglobulin heavy variable 3-72 | 1 | TEDTAVY | A0A0B4J1 | HV372 H | IGHV3-72 | TEDTAVY | TEDTAVY | [illegible] |
| P23142;P23142-4 | Fibulin-1 | 1 | TGYYFD( | P23142:P | FBLN1 H | FBLN1 | TGYYFD( | TGYYFD( | TGYYFDGISR2 |
| P69905 | Hemoglobin subunit alpha | 4 | VGAHAG | P69905 | HBA HUN | HBA1 | VGAHAG | VGAHAG | [illegible] |
| A0A075B6J9 | Immunoglobulin lambda variable 2-18 | 1 | VSWYQQ | A0A075B | LV218 HU | IGLV2-18 | VSWYQQ | VSWYQQ | [illegible] |
| A0A0C4DH34 | Immunoglobulin heavy variable 4-28 | 2 | VTMSVD | A0A0C4D | HV428 H | IGHV4-28 | VTMSVD | VTMSVD | [illegible] |
| B9A064 | Immunoglobulin lambda-like polypeptide 5 | 2 | VTVLGQI | B9A064:A | IGLL5 HU | IGLL5 | VTVLGQI | VTVLGQI | [illegible] |
| P06276 | Cholinesterase | 1 | YLTLNTE | P06276:H | CHLE HU | BCHE | YLTLNTE | YLTLNTE | YLTLNTESTR2 |

4)FAEEGKK3+ADDKETC(UniMod:4)FAEEGKK4+AEFAEVSK1+AEFAEVSK2+AEFAEVSKLVTDLTK2+AEFAEVSKLVTDLTK3+AEFAEVSKLVTDLTK4+AFKAWAVAR2+ALVLIAFAQYLQQC(UniMod:4)PFEDHVK3+ALVLIAFAQYLQQC(UniMod:4)PFEDHVK4+ALVLIAFAQYLQQC(UniMod:4)PFEDHVKLVNEVTEFAK5+ATKEQLK1+ATKEQLKAVMDDFAAFVEK3+ATKEQLKAVMDDFAAFVEK4+AVM(UniMod:35)DDFAAFVEK2+AVM(UniMod:35)DDFAAFVEK3+AVMDDFAAFVEK2+AVMDDFAAFVEK3+C(UniMod:4)C(UniMod:4)AAADPHEC(UniMod:4)YAK2+C(UniMod:4)C(UniMod:4)AAADPHEC(UniMod:4)YAK3+C(UniMod:4)C(

EGLEK3+FEVQVTVPK2+FQVDNNNR2+FRVVSMDENFHPLNELIPLVYIQDPK4+FRVVSMDENFHPLNELIPLVYIQDPKGNR5+FSGQLNSHGC(UniMod:4)FYQQVK3+GEAFTLK1+GHFSISIPVK2+GHFSISIPVKSDIAPVAR4+GPTQEFK1+GPTQEFK2+GVPIPNKVIFIR2+GVPIPNKVIFIR3+HYDGSYSTFGER2+HYDGSYSTFGER3+IAQWQSFQLEGGLK2+IAQWQSFQLEGGLK3+IAQWQSFQLEGGLKQFSFPLSSEPFQGSYK4+KDTVIKPLLVEPEGLEK4+KLSFYYLIMAK3+KYSDASDC(UniMod:4)HGEDSQAFC(UniMod:4)EK3+KYSDASDC(UniMod:4)HGEDSQAFC(UniMod:4)EK4+LHTEAQIQEEGTVVELTGR3+LHTEAQIQEEGTVVELTGR4+LLIY

SR3+DFDFVPPVVR2+DKNRWEDPGK2+DSC(UniMod:4)VGSLVVK2+EALKLEEK2+EGVQKEDIPPADLSDQVPDTESETR4+EVVADSVWVDVK2+EVVADSVWVDVKDSC(UniMod:4)VGSLVVK3+FISLGEAC(UniMod:4)KK2+FYHPEKEDGKLNK2+FYHPEKEDGKLNK3+FYHPEKEDGKLNK4+FYYIYNEK2+GLEVTITAR2+GQGTLSVVTMYHAK2+GQGTLSVVTMYHAK3+GVFVLNK1+GVFVLNKK2+GYTQQLAFR2+GYTQQLAFRQPSSAFAAFVK3+GYTQQLAFRQPSSAFAAFVKR3+GYTQQLAFRQPSSAFAAFVKR4+HQQTVTIPPK2+HQQTVTIPPKSSLSVPYVIVPLKTGLQEVEVK4+HQQTVTIPPKSSLSVPYVIVPLKTGLQEVEVK5+IFTVNH

(UniMod:4)LAK2+DGAGDVAFVK2+DKSKEFQLFSSPHGK4+DLLFKDSAHGFLK2+DLLFKDSAHGFLK3+DLLFKDSAHGFLKVPPR3+DLLFKDSAHGFLKVPPR4+DLLFRDDTVC(UniMod:4)LAK2+DLLFRDDTVC(UniMod:4)LAK3+DSAHGFLK1+DSAHGFLK2+DSAHGFLKVPPR2+DSAHGFLKVPPR3+DSGFQMNQLR2+DSGFQMNQLR3+DYELLC(UniMod:4)LDGTRKPVEEYANC(UniMod:4)HLAR5+EDPQTFYYAVAVVK2+EDPQTFYYAVAVVKK3+EFQLFSSPHGK2+EFQLFSSPHGK3+EFQLFSSPHGKDLLFK3+EFQLFSSPHGKDLLFK4+EGTC(UniMod:4)PEAPTDEC(UniMod:4)KPVK3+EGTC(UniMod:4)PEAPTDEC(UniMod:4)KPVKWC(Un

TGQYER4+HRQGPVNLLSDPEQGVEVTGQYEREK4+HRQGPVNLLSDPEQGVEVTGQYEREK5+ILDDLSPR2+ITFELVYEELLK2+ITFELVYEELLK3+ITFELVYEELLKR3+LALDNGGLAR2+LGVYELLLK2+LLFKGSEMVVAGK3+LQDRGPDVLTATVSGK2+LQDRGPDVLTATVSGK3+M(UniMod:35)NFRPGVLSSR3+MNFRPGVLSSR2+MNFRPGVLSSR3+NM(UniMod:35)EQFQVSVSVAPNAK2+NMEQFQVSVSVAPNAK2+NMEQFQVSVSVAPNAK3+NPLVWVHASPEHVVVTR3+NPLVWVHASPEHVVVTR4+NVVFVIDK2+NVVFVIDKSGSMSGR2+NVVFVIDKSGSMSGR3+QGPVNLLSDPEQGVEVTGQYER3+QGPVNLLSDPEQGVEVTGQYER

PR4+HYYIGIIETTWDYASDHGEK3+HYYIGIIETTWDYASDHGEK4+HYYIGIIETTWDYASDHGEKK4+HYYIGIIETTWDYASDHGEKK5+IYHSHIDAPK2+KAEEEHLGILGPQLHADVGDKVK3+KAEEEHLGILGPQLHADVGDKVK4+KAEEEHLGILGPQLHADVGDKVK5+KAEEEHLGILGPQLHADVGDKVK6+KALYLQYTDETFR2+KALYLQYTDETFR3+KERGPEEEHLGILGPVIWAEVGDTIR5+KLISVDTEHSNIYLQNGPDR3+KLISVDTEHSNIYLQNGPDR4+LISVDTEHSNIYLQNGPDR3+LISVDTEHSNIYLQNGPDR4+LVYREYTDASFTNR3+LVYREYTDASFTNRK4+M(UniMod:35)YYSAVDPTKDIFTGLIGPMK3+MYYSAVDPTKDIFTGLIGPMK3+N
GRDYVSQFEGSALGK4+DYVSQFEGSALGK2+DYVSQFEGSALGK3+EQLGPVTQEFWDNLEKETEGLRQEMSK4+ETEGLRQEMSK3+ETEGLRQEMSKDLEEVK3+KWQEEMELYR2+KWQEEMELYR3+LAEYHAK1+LAEYHAK2+LEALKENGGAR2+LHELQEK1+LHELQEK2+LLDNWDSVTSTFSK2+LLDNWDSVTSTFSK3+LREQLGPVTQEFWDNLEK3+LSPLGEEM(UniMod:35)RDR3+LSPLGEEMR2+LSPLGEEMRDR2+LSPLGEEMRDR3+QEMSKDLEEVKAK3+QGLLPVLESFK2+QGLLPVLESFK3+QKLHELQEK2+QKVEPLR2+QKVEPLRAELQEGAR2+QKVEPLRAELQEGAR3+QKVEPLRAELQEGAR4+THLAPYSDELR2+THLAPYSDELR3+

DSSVKPYR3+GGETSEM(UniMod:35)YLIQPDSSVKPYR4+GGETSEMYLIQPDSSVKPYR3+GGETSEMYLIQPDSSVKPYR4+GHRPLDK2+GSWYSMR1+GSWYSMR2+GSWYSMRK2+HGTDDGVVWMNWK2+HGTDDGVVWMNWK3+HQLYIDETVNSNIPTNLR3+HQLYIDETVNSNIPTNLR4+IQKLESDVSAQMEYC(UniMod:4)R3+IRPFFPQQ2+KAPDAGGC(UniMod:4)LHADPDLGVLC(UniMod:4)PTGC(UniMod:4)QLQEALLQQERPIR5+KGGETSEM(UniMod:35)YLIQPDSSVKPYR5+KGGETSEMYLIQPDSSVKPYR3+KGGETSEMYLIQPDSSVKPYR4+KGGETSEMYLIQPDSSVKPYR5+KQC(UniMod:4)SKEDGGGWWYNR3+KQC(UniMod:4)SKED

QHHLGGAK5+IVNLKEK1+IVNLKEK2+KMLEEIMK2+KNWIQYK2+LDGSVDFK2+LDGSVDFKK2+LDGSVDFKKNWIQYK4+LTIGEGQQHHLGGAK2+LTIGEGQQHHLGGAK3+M(UniMod:35)LEEIMK2+M(UniMod:35)LEEIMKYEASILTHDSSIR3+M(UniMod:35)LEEIMKYEASILTHDSSIR4+MLEEIMK1+MLEEIMK2+MLEEIMKYEASILTHDSSIR3+MLEEIMKYEASILTHDSSIR4+NWIQYKEGFGHLSPTGTTEFWLGNEK4+QSGLYFIKPLK2+QSGLYFIKPLK3+RLDGSVDFK2+RLDGSVDFKK2+TRWYSMK2+TSTADYAMFK2+VAQLEAQC(UniMod:4)QEPC(UniMod:4)KDTVQIHDITGK4+VAQLEAQC(UniMod:4)QEPC(UniMod:4)KDTVQIHDITGK

VEGHLSFLEK3+LGPHAGDVEGHLSFLEK4+LKEEIGKELEELR3+LKEEIGKELEELR4+LLPHANEVSQK2+LLPHANEVSQK3+LNHQLEGLTFQMK3+LTPYADEFK2+LTPYADEFKVK2+LTPYADEFKVK3+LVPFATELHER2+LVPFATELHER3+NAEELKAR2+QLTPYAQR2+RVEPYGENFNK2+RVEPYGENFNK3+SELTQQLNALFQDK2+SELTQQLNALFQDKLGEVNTYAGDLQK4+SLAELGGHLDQQVEEFR3+SLAELGGHLDQQVEEFRR3+SLAELGGHLDQQVEEFRR4+SLAPYAQDTQEK2+SLAPYAQDTQEKLNHQLEGLTFQMK4+TQVNTQAEQLR2+TQVNTQAEQLRR3+VEPYGENFNK2+VKIDQTVEELRR3+VLRENADSLQASLRPHADELK4+

FR4+EYHTEKLVTSK2+EYHTEKLVTSK3+GDFSSANNR2+GDFSSANNRDNTYNR2+GDFSSANNRDNTYNR3+GDSTFESK2+GGSTSYGTGSETESPR2+GGSTSYGTGSETESPR3+GGSTSYGTGSETESPRNPSSAGSWNSGSSGPGSTGNR4+GLIDEVNQDFTNR2+GLIDEVNQDFTNR3+GLIDEVNQDFTNRINK3+GSESGIFTNTK2+GSESGIFTNTKESSSHHPGIAEFPSR4+GSESGIFTNTKESSSHHPGIAEFPSR5+HPDEAAFFDTASTGK2+HPDEAAFFDTASTGK3+HQSAC(UniMod:4)KDSDWPFC(UniMod:4)SDEDWNYKC(UniMod:4)PSGC(UniMod:4)R4+HQSAC(UniMod:4)KDSDWPFC(UniMod:4)SDEDWNYKC(UniMod:4)PSGC(UniMod:4)R5+LEVDIDIK2+
IFNMAR2+EIQIYKK2+EQHLFLPFSYK2+EVYGFNPEGK2+FFGEGTKK2+FIIPSPK1+FIIPSPK2+FPEVDVLTK2+FSVPAGIVIPSFQALTAR3+FVTQAEGAK2+GFEPTLEALFGK2+GIISALLVPPETEEAK2+GIISALLVPPETEEAK3+GMTRPLSTLISSSQSC(UniMod:4)QYTLDAK3+GNVATEISTER2+IADFELPTIIVPEQTIEIPSIK3+IADFELPTIIVPEQTIEIPSIK4+IEDGTLASK2+IEGNLIFDPNNYLPK2+IEGNLIFDPNNYLPK3+ILGEELGFASLHDLQLLGK3+INC(UniMod:4)KVELEVPQLC(UniMod:4)SFILK3+ITLPDFR2+IYSLWEHSTK2+IYSLWEHSTK3+KGNVATEISTER2+KGNVATEISTER3+KGNVATEISTERDLGQC(UniMod:4)DR4+KLTISEQNIQR2+KL

DR3+KLSSWVLLM(UniMod:35)K3+KLSSWVLLMK2+KLSSWVLLMK3+KLYHSEAFTVNFGDTEEAKK4+KLYHSEAFTVNFGDTEEAKK5+KQINDYVEK2+KQINDYVEKGTQGK2+KQINDYVEKGTQGK3+KQINDYVEKGTQGK4+LGMFNIQHC(UniMod:4)K2+LGMFNIQHC(UniMod:4)K3+LGMFNIQHC(UniMod:4)KK2+LGMFNIQHC(UniMod:4)KK3+LQHLENELTHDIITK4+LSITGTYDLK2+LSSWVLLMK2+LVDKFLEDVK2+LVDKFLEDVK3+LVDKFLEDVKK2+LVDKFLEDVKK3+LYHSEAFTVNFGDTEEAK3+LYHSEAFTVNFGDTEEAKK3+LYHSEAFTVNFGDTEEAKK4+LYHSEAFTVNFGDTEEAKKQINDYVEK4+LYHSEAFTVNFGDTEE

GGWWYNR4+KREEAPSLRPAPPPISGGGYR3+KREEAPSLRPAPPPISGGGYR4+KREEAPSLRPAPPPISGGGYR5+KWDPYK2+KWDPYKQGFGNVATNTDGK3+KWDPYKQGFGNVATNTDGK4+LESDVSAQMEYC(UniMod:4)R2+M(UniMod:35)GPTELLIEMEDWKGDKVK3+M(UniMod:35)GPTELLIEMEDWKGDKVK4+MGPTELLIEMEDWK2+MGPTELLIEMEDWK3+MGPTELLIEMEDWKGDK3+MGPTEL
