## supplementary_table2 for "Molecular characterization of liver steatosis through circulating proteins, metabolites and lipids" (from another version of this preprint: version 1, 2025-12-02)

| Analyte name | Analyte class | Biochemical name |
| --- | --- | --- |
| ADMA | biogenic amines | Asymmetric dimethylarginine |
| Ala | aminoacids | Alanine |
| alpha-AAA | biogenic amines | alpha-Aminoadipic acid |
| Arg | aminoacids | Arginine |
| Asn | aminoacids | Asparagine |
| Asp | aminoacids | Aspartate |
| Cit | aminoacids | Citrulline |
| Creatinine | biogenic amines | Creatinine |
| DOPA | biogenic amines | Dihydroxyphenylalanine |
| Gln | aminoacids | Glutamine |
| Glu | aminoacids | Glutamate |
| Gly | aminoacids | Glycine |
| His | aminoacids | Histidine |
| Histamine | biogenic amines | Histamine |
| Ile | aminoacids | Isoleucine |
| Kynurenine | biogenic amines | Kynurenine |
| Leu | aminoacids | Leucine |
| Lys | aminoacids | Lysine |
| Met | aminoacids | Methionine |
| Met-SO | biogenic amines | Methioninesulfoxide |
| Orn | aminoacids | Ornithine |
| Phe | aminoacids | Phenylalanine |
| Pro | aminoacids | Proline |
| Putrescine | biogenic amines | Putrescine |
| Sarcosine | biogenic amines | Sarcosine |
| SDMA | biogenic amines | Symmetric dimethylarginine |
| Ser | aminoacids | Serine |
| Serotonin | biogenic amines | Serotonin |
| Spermidine | biogenic amines | Spermidine |
| Spermine | biogenic amines | Spermine |
| t4-OH-Pro | biogenic amines | trans-4-Hydroxyproline |
| Taurine | biogenic amines | Taurine |
| Thr | aminoacids | Threonine |
| Trp | aminoacids | Tryptophan |
| Tyr | aminoacids | Tyrosine |
| Val | aminoacids | Valine |
| C0 | acylcarnitines | Carnitine |
| C10 | acylcarnitines | Decanoylcarnitine |
| C10:1 | acylcarnitines | Decenoylcarnitine |
| C10:2 | acylcarnitines | Decadienylcarnitine |
| C12 | acylcarnitines | Dodecanoylcarnitine |
| C12-DC | acylcarnitines | Dodecanedioylcarnitine |
| C12:1 | acylcarnitines | Dodecenoylcarnitine |
| C14 | acylcarnitines | Tetradecanoylcarnitine |
| C14:1 | acylcarnitines | Tetradecenoylcarnitine |
| C14:1-OH | acylcarnitines | Hydroxytetradecenoylcarnitine |

| | | |
|---|---|---|
| C14:2 | acylcarnitines | Tetradecadienylcarnitine |
| C14:2-OH | acylcarnitines | Hydroxytetradecadienylcarnitine |
| C16 | acylcarnitines | Hexadecanoylcarnitine |
| C16-OH | acylcarnitines | Hydroxyhexadecanoylcarnitine |
| C16:1 | acylcarnitines | Hexadecenoylcarnitine |
| C16:1-OH | acylcarnitines | Hydroxyhexadecenoylcarnitine |
| C18 | acylcarnitines | Octadecanoylcarnitine |
| C18:1 | acylcarnitines | Octadecenoylcarnitine |
| C2 | acylcarnitines | Acetylcarnitine |
| C3 | acylcarnitines | Propionylcarnitine |
| C3-DC (C4-OH) | acylcarnitines | Hydroxybutyrylcarnitine |
| C3:1 | acylcarnitines | Propenoylcarnitine |
| C4 | acylcarnitines | Butyrylcarnitine |
| C4:1 | acylcarnitines | Butenylcarnitine |
| C5 | acylcarnitines | Valerylcarnitine |
| C5-DC (C6-OH) | acylcarnitines | Glutarylcarnitine (Hydroxyhexanoylcarnitine) |
| C5-M-DC | acylcarnitines | Methylglutarylcarnitine |
| C5-OH (C3-DC-M) | acylcarnitines | Hydroxyvalerylcarnitine (Methylmalonylcarnitine) |
| C5:1 | acylcarnitines | Tiglylcarnitine |
| C5:1-DC | acylcarnitines | Glutaconylcarnitine |
| C6 (C4:1-DC) | acylcarnitines | Hexanoylcarnitine (Fumarylcarnitine) |
| C6:1 | acylcarnitines | Hexenoylcarnitine |
| C8 | acylcarnitines | Octanoylcarnitine |
| C9 | acylcarnitines | Nonaylcarnitine |
| H1 | sugars | Hexose |
| lysoPC a C14:0 | glycerophospholipids | lysoPhosphatidylcholine acyl C14:0 |
| lysoPC a C16:0 | glycerophospholipids | lysoPhosphatidylcholine acyl C16:0 |
| lysoPC a C16:1 | glycerophospholipids | lysoPhosphatidylcholine acyl C16:1 |
| lysoPC a C17:0 | glycerophospholipids | lysoPhosphatidylcholine acyl C17:0 |
| lysoPC a C18:0 | glycerophospholipids | lysoPhosphatidylcholine acyl C18:0 |
| lysoPC a C18:1 | glycerophospholipids | lysoPhosphatidylcholine acyl C18:1 |
| lysoPC a C18:2 | glycerophospholipids | lysoPhosphatidylcholine acyl C18:2 |
| lysoPC a C20:3 | glycerophospholipids | lysoPhosphatidylcholine acyl C20:3 |
| lysoPC a C20:4 | glycerophospholipids | lysoPhosphatidylcholine acyl C20:4 |
| lysoPC a C24:0 | glycerophospholipids | lysoPhosphatidylcholine acyl C24:0 |
| lysoPC a C26:0 | glycerophospholipids | lysoPhosphatidylcholine acyl C26:0 |
| lysoPC a C26:1 | glycerophospholipids | lysoPhosphatidylcholine acyl C26:1 |
| lysoPC a C28:0 | glycerophospholipids | lysoPhosphatidylcholine acyl C28:0 |
| lysoPC a C28:1 | glycerophospholipids | lysoPhosphatidylcholine acyl C28:1 |
| PC aa C24:0 | glycerophospholipids | Phosphatidylcholine diacyl C24:0 |
| PC aa C26:0 | glycerophospholipids | Phosphatidylcholine diacyl C26:0 |
| PC aa C28:1 | glycerophospholipids | Phosphatidylcholine diacyl C28:1 |
| PC aa C30:0 | glycerophospholipids | Phosphatidylcholine diacyl C30:0 |
| PC aa C30:2 | glycerophospholipids | Phosphatidylcholine diacyl C30:2 |
| PC aa C32:0 | glycerophospholipids | Phosphatidylcholine diacyl C32:0 |
| PC aa C32:1 | glycerophospholipids | Phosphatidylcholine diacyl C32:1 |
| PC aa C32:2 | glycerophospholipids | Phosphatidylcholine diacyl C32:2 |

| | | |
|---|---|---|
| PC aa C32:3 | glycerophospholipids | Phosphatidylcholine diacyl C32:3 |
| PC aa C34:1 | glycerophospholipids | Phosphatidylcholine diacyl C34:1 |
| PC aa C34:2 | glycerophospholipids | Phosphatidylcholine diacyl C34:2 |
| PC aa C34:3 | glycerophospholipids | Phosphatidylcholine diacyl C34:3 |
| PC aa C34:4 | glycerophospholipids | Phosphatidylcholine diacyl C34:4 |
| PC aa C36:0 | glycerophospholipids | Phosphatidylcholine diacyl C36:0 |
| PC aa C36:1 | glycerophospholipids | Phosphatidylcholine diacyl C36:1 |
| PC aa C36:2 | glycerophospholipids | Phosphatidylcholine diacyl C36:2 |
| PC aa C36:3 | glycerophospholipids | Phosphatidylcholine diacyl C36:3 |
| PC aa C36:4 | glycerophospholipids | Phosphatidylcholine diacyl C36:4 |
| PC aa C36:5 | glycerophospholipids | Phosphatidylcholine diacyl C36:5 |
| PC aa C36:6 | glycerophospholipids | Phosphatidylcholine diacyl C36:6 |
| PC aa C38:0 | glycerophospholipids | Phosphatidylcholine diacyl C38:0 |
| PC aa C38:1 | glycerophospholipids | Phosphatidylcholine diacyl C38:1 |
| PC aa C38:3 | glycerophospholipids | Phosphatidylcholine diacyl C38:3 |
| PC aa C38:4 | glycerophospholipids | Phosphatidylcholine diacyl C38:4 |
| PC aa C38:5 | glycerophospholipids | Phosphatidylcholine diacyl C38:5 |
| PC aa C38:6 | glycerophospholipids | Phosphatidylcholine diacyl C38:6 |
| PC aa C40:1 | glycerophospholipids | Phosphatidylcholine diacyl C40:1 |
| PC aa C40:2 | glycerophospholipids | Phosphatidylcholine diacyl C40:2 |
| PC aa C40:3 | glycerophospholipids | Phosphatidylcholine diacyl C40:3 |
| PC aa C40:4 | glycerophospholipids | Phosphatidylcholine diacyl C40:4 |
| PC aa C40:5 | glycerophospholipids | Phosphatidylcholine diacyl C40:5 |
| PC aa C40:6 | glycerophospholipids | Phosphatidylcholine diacyl C40:6 |
| PC aa C42:0 | glycerophospholipids | Phosphatidylcholine diacyl C42:0 |
| PC aa C42:1 | glycerophospholipids | Phosphatidylcholine diacyl C42:1 |
| PC aa C42:2 | glycerophospholipids | Phosphatidylcholine diacyl C42:2 |
| PC aa C42:4 | glycerophospholipids | Phosphatidylcholine diacyl C42:4 |
| PC aa C42:5 | glycerophospholipids | Phosphatidylcholine diacyl C42:5 |
| PC aa C42:6 | glycerophospholipids | Phosphatidylcholine diacyl C42:6 |
| PC ae C30:0 | glycerophospholipids | Phosphatidylcholine acyl-alkyl C30:0 |
| PC ae C30:1 | glycerophospholipids | Phosphatidylcholine acyl-alkyl C30:1 |
| PC ae C30:2 | glycerophospholipids | Phosphatidylcholine acyl-alkyl C30:2 |
| PC ae C32:1 | glycerophospholipids | Phosphatidylcholine acyl-alkyl C32:1 |
| PC ae C32:2 | glycerophospholipids | Phosphatidylcholine acyl-alkyl C32:2 |
| PC ae C34:0 | glycerophospholipids | Phosphatidylcholine acyl-alkyl C34:0 |
| PC ae C34:1 | glycerophospholipids | Phosphatidylcholine acyl-alkyl C34:1 |
| PC ae C34:2 | glycerophospholipids | Phosphatidylcholine acyl-alkyl C34:2 |
| PC ae C34:3 | glycerophospholipids | Phosphatidylcholine acyl-alkyl C34:3 |
| PC ae C36:0 | glycerophospholipids | Phosphatidylcholine acyl-alkyl C36:0 |
| PC ae C36:1 | glycerophospholipids | Phosphatidylcholine acyl-alkyl C36:1 |
| PC ae C36:2 | glycerophospholipids | Phosphatidylcholine acyl-alkyl C36:2 |
| PC ae C36:3 | glycerophospholipids | Phosphatidylcholine acyl-alkyl C36:3 |
| PC ae C36:4 | glycerophospholipids | Phosphatidylcholine acyl-alkyl C36:4 |
| PC ae C36:5 | glycerophospholipids | Phosphatidylcholine acyl-alkyl C36:5 |
| PC ae C38:0 | glycerophospholipids | Phosphatidylcholine acyl-alkyl C38:0 |
| PC ae C38:1 | glycerophospholipids | Phosphatidylcholine acyl-alkyl C38:1 |

| | | |
|---|---|---|
| PC ae C38:2 | glycerophospholipids | Phosphatidylcholine acyl-alkyl C38:2 |
| PC ae C38:3 | glycerophospholipids | Phosphatidylcholine acyl-alkyl C38:3 |
| PC ae C38:4 | glycerophospholipids | Phosphatidylcholine acyl-alkyl C38:4 |
| PC ae C38:5 | glycerophospholipids | Phosphatidylcholine acyl-alkyl C38:5 |
| PC ae C38:6 | glycerophospholipids | Phosphatidylcholine acyl-alkyl C38:6 |
| PC ae C40:1 | glycerophospholipids | Phosphatidylcholine acyl-alkyl C40:1 |
| PC ae C40:2 | glycerophospholipids | Phosphatidylcholine acyl-alkyl C40:2 |
| PC ae C40:3 | glycerophospholipids | Phosphatidylcholine acyl-alkyl C40:3 |
| PC ae C40:4 | glycerophospholipids | Phosphatidylcholine acyl-alkyl C40:4 |
| PC ae C40:5 | glycerophospholipids | Phosphatidylcholine acyl-alkyl C40:5 |
| PC ae C40:6 | glycerophospholipids | Phosphatidylcholine acyl-alkyl C40:6 |
| PC ae C42:0 | glycerophospholipids | Phosphatidylcholine acyl-alkyl C42:0 |
| PC ae C42:1 | glycerophospholipids | Phosphatidylcholine acyl-alkyl C42:1 |
| PC ae C42:2 | glycerophospholipids | Phosphatidylcholine acyl-alkyl C42:2 |
| PC ae C42:3 | glycerophospholipids | Phosphatidylcholine acyl-alkyl C42:3 |
| PC ae C42:4 | glycerophospholipids | Phosphatidylcholine acyl-alkyl C42:4 |
| PC ae C42:5 | glycerophospholipids | Phosphatidylcholine acyl-alkyl C42:5 |
| PC ae C44:3 | glycerophospholipids | Phosphatidylcholine acyl-alkyl C44:3 |
| PC ae C44:4 | glycerophospholipids | Phosphatidylcholine acyl-alkyl C44:4 |
| PC ae C44:5 | glycerophospholipids | Phosphatidylcholine acyl-alkyl C44:5 |
| PC ae C44:6 | glycerophospholipids | Phosphatidylcholine acyl-alkyl C44:6 |
| SM (OH) C14:1 | sphingolipids | Hydroxysphingomyeline C14:1 |
| SM (OH) C16:1 | sphingolipids | Hydroxysphingomyeline C16:1 |
| SM (OH) C22:1 | sphingolipids | Hydroxysphingomyeline C22:1 |
| SM (OH) C22:2 | sphingolipids | Hydroxysphingomyeline C22:2 |
| SM (OH) C24:1 | sphingolipids | Hydroxysphingomyeline C24:1 |
| SM C16:0 | sphingolipids | Sphingomyeline C16:0 |
| SM C16:1 | sphingolipids | Sphingomyeline C16:1 |
| SM C18:0 | sphingolipids | Sphingomyeline C18:0 |
| SM C18:1 | sphingolipids | Sphingomyeline C18:1 |
| SM C20:2 | sphingolipids | Sphingomyeline C20:2 |
| SM C24:0 | sphingolipids | Sphingomyeline C24:0 |
| SM C24:1 | sphingolipids | Sphingomyeline C24:1 |
| SM C26:0 | sphingolipids | Sphingomyeline C26:0 |
| SM C26:1 | sphingolipids | Sphingomyeline C26:1 |
