## supplementary_table3 for "Molecular characterization of liver steatosis through circulating proteins, metabolites and lipids" (from another version of this preprint: version 1, 2025-12-02)

| Uniprot ID | Gene | cv_qc_chri | oef_SexMa | oef_Age_1 | liver_fat_( | liver_fat_( | liver_fat_( | alue_SexM | value_Age_ | e_liver_fat | e_liver_fat | e_liver_fat | adj_SexM | .adj_Age_1 | liver_fat_ | liver_fat_ | liver_fat_ | ctSize_SexM | ctSize_Age | ze_liver_fa | ze_liver_fa | ze_liver_fa | ficant_Sex | ificant_Age | nt_liver_fa | nt_liver_fa | nt_liver_fa | perc_SexM | perc_Age | rc_liver_fa | rc_liver_fa | rc_liver_fat_cat14 |
|---|---|---|---|---|---|---|---|---|---|---|---|---|---|---|---|---|---|---|---|---|---|---|---|---|---|---|---|---|---|---|---|---|
| P02768 | ALB | 4.737056 | 0.071658 | -0.01472 | 0.036652 | -0.02179 | 0.000692 | 1.23E-05 | 0.054683 | 0.122728 | 0.349962 | 0.97289 | 0.000121 | 0.126454 | 0.490913 | 0.849036 | 0.987963 | 0.478585 | -0.09828 | 0.244786 | -0.14554 | 0.004625 | TRUE | FALSE | FALSE | FALSE | FALSE | 5.092388 | 1.025232 | 2.573043 | 1.521907 | 0.048009 |
| P02766 | TTR | 10.01682 | 0.329469 | -0.1405 | 0.146201 | -0.08527 | -0.07666 | 4.11E-13 | 4.51E-11 | 0.02286 | 0.175788 | 0.164003 | 3.04E-11 | 6.07E-10 | 0.255459 | 0.700478 | 0.40454 | 0.733455 | -0.31277 | 0.325468 | -0.18982 | -0.17067 | TRUE | TRUE | FALSE | FALSE | FALSE | 25.65511 | 10.22863 | 10.66514 | 6.08839 | 5.457669 |
| P19827 | ITIH1 | 3.376043 | 0.032991 | -0.03273 | -0.01494 | -0.04526 | -0.03941 | 0.078419 | 0.00025 | 0.586324 | 0.094248 | 0.095643 | 0.168203 | 0.001232 | 0.885469 | 0.536489 | 0.277552 | 0.191946 | -0.19045 | -0.08694 | -0.26331 | -0.22931 | FALSE | TRUE | FALSE | FALSE | FALSE | 2.313133 | 2.294924 | 1.041184 | 3.186742 | 2.769546 |
| P01023 | A2M | 3.529184 | -0.03587 | 0.145625 | -0.0263 | -0.04771 | 0.038451 | 0.414887 | 1.44E-11 | 0.683795 | 0.45228 | 0.488538 | 0.596148 | 2.13E-10 | 0.926973 | 0.934694 | 0.731172 | -0.08522 | 0.345965 | -0.06247 | -0.11336 | 0.09135 | FALSE | TRUE | FALSE | FALSE | FALSE | 2.517441 | 10.62101 | 1.839413 | 3.362584 | 2.701077 |
| P01042;P0 | KNG1 | 4.208245 | -0.01699 | -0.04029 | -0.01517 | -0.00515 | -0.007 | 0.349454 | 3.92E-06 | 0.569131 | 0.844094 | 0.75995 | 0.544413 | 2.76E-05 | 0.885469 | 0.96269 | 0.920514 | -0.10135 | -0.2403 | -0.0905 | -0.03072 | -0.04174 | FALSE | TRUE | FALSE | FALSE | FALSE | 1.184845 | 2.832142 | 1.057342 | 0.357671 | 0.48633 |
| P02649 | APOE | 6.889968 | -0.02112 | -0.01101 | -0.05121 | -0.10236 | -0.03602 | 0.624903 | 0.589836 | 0.419479 | 0.101202 | 0.508856 | 0.751916 | 0.715539 | 0.787894 | 0.553717 | 0.731172 | -0.05461 | -0.02848 | -0.13242 | -0.26468 | -0.09313 | FALSE | FALSE | FALSE | FALSE | FALSE | 1.474534 | 0.76637 | 3.613449 | 7.35275 | 2.527938 |
| P01024 | C3 | 2.435395 | -0.00755 | -0.02079 | 0.063204 | 0.059557 | 0.132412 | 0.698121 | 0.024536 | 0.027617 | 0.034667 | 1.35E-07 | 0.826576 | 0.069834 | 0.255459 | 0.32067 | 6.65E-06 | -0.04133 | -0.11377 | 0.345859 | 0.325898 | 0.724568 | FALSE | FALSE | FALSE | FALSE | TRUE | 0.524898 | 1.451623 | 4.478384 | 4.214548 | 9.612496 |
| P04196 | HRG | 4.591216 | -0.02736 | 0.015093 | 0.067769 | 0.010352 | -0.00852 | 0.425904 | 0.353162 | 0.179542 | 0.834552 | 0.844141 | 0.598361 | 0.500643 | 0.583261 | 0.96269 | 0.947857 | -0.08886 | 0.049023 | 0.220112 | 0.033623 | -0.02768 | FALSE | FALSE | FALSE | FALSE | FALSE | 1.914484 | 1.05169 | 4.809474 | 0.720129 | 0.592423 |
| P01011 | SERPINA: | 5.494796 | -0.00573 | 0.026668 | -0.04997 | -0.04021 | -0.12647 | 0.823791 | 0.029179 | 0.186875 | 0.279661 | 0.00012 | 0.889934 | 0.079684 | 0.583261 | 0.78094 | 0.001365 | -0.02421 | 0.112594 | -0.21098 | -0.16979 | -0.53398 | FALSE | FALSE | FALSE | FALSE | TRUE | 0.3983 | 1.865687 | 3.524472 | 2.826661 | 9.162228 |
| P02787 | TF | 3.795468 | 0.005596 | -0.03604 | 0.028948 | 0.005889 | -0.02412 | 0.798229 | 0.000563 | 0.367973 | 0.852111 | 0.38294 | 0.878542 | 0.002451 | 0.745583 | 0.96269 | 0.674705 | 0.028135 | -0.18121 | 0.145548 | 0.029607 | -0.12125 | FALSE | TRUE | FALSE | FALSE | FALSE | 0.388615 | 2.529578 | 2.026766 | 0.408996 | 1.685604 |
| P01834 | IGKC | 3.809914 | 0.107304 | 0.008737 | 0.009501 | 0.036663 | 0.008672 | 0.005298 | 0.629242 | 0.865644 | 0.506698 | 0.857434 | 0.020105 | 0.738443 | 0.959934 | 0.942137 | 0.947857 | 0.310794 | 0.025305 | 0.02752 | 0.106191 | 0.025118 | TRUE | FALSE | FALSE | FALSE | FALSE | 7.721298 | 0.607416 | 0.660761 | 2.573855 | 0.602915 |
| Q14624;Q | ITIH4 | 3.771327 | -0.06328 | 0.006235 | -0.07547 | -0.0196 | -0.11565 | 0.001808 | 0.512684 | 0.011058 | 0.50002 | 7.41E-06 | 0.008921 | 0.659802 | 0.204566 | 0.942137 | 0.000123 | -0.33432 | 0.032941 | -0.39869 | -0.10354 | -0.61099 | TRUE | FALSE | FALSE | FALSE | TRUE | 4.483963 | 0.433126 | 5.370079 | 1.367692 | 8.346315 |
| P07360 | C8G | 9.407207 | 0.029549 | -0.03466 | 0.124636 | 0.081432 | 0.083025 | 0.421575 | 0.046856 | 0.021409 | 0.125328 | 0.074138 | 0.598361 | 0.115577 | 0.255459 | 0.59834 | 0.24859 | 0.088764 | -0.10411 | 0.374407 | 0.244622 | 0.249407 | FALSE | FALSE | FALSE | FALSE | FALSE | 2.069278 | 2.431221 | 9.023284 | 5.806795 | 5.923663 |
| P00450 | CP | 4.164603 | -0.14425 | 0.00847 | -0.0393 | -0.0301 | -0.09965 | 1.02E-08 | 0.465864 | 0.275849 | 0.395742 | 0.001415 | 1.89E-07 | 0.615605 | 0.648026 | 0.874176 | 0.011022 | -0.61368 | 0.036034 | -0.16721 | -0.12804 | -0.42397 | TRUE | FALSE | FALSE | FALSE | TRUE | 10.51523 | 0.588804 | 2.761695 | 2.108073 | 7.151606 |
| P02647 | APOA1 | 20.20619 | -0.06056 | -0.01334 | -0.01235 | -0.068 | -0.0763 | 0.002396 | 0.15497 | 0.670937 | 0.017836 | 0.002436 | 0.010744 | 0.264661 | 0.926973 | 0.219978 | 0.018027 | -0.3304 | -0.07279 | -0.0674 | -0.37097 | -0.41628 | FALSE | FALSE | FALSE | FALSE | FALSE | 4.28706 | 0.929133 | 0.859966 | 4.82594 | 5.431111 |
| P02760 | AMBP | 4.399126 | 0.029219 | 0.020331 | -0.01495 | -0.02919 | -0.05434 | 0.147377 | 0.033398 | 0.613117 | 0.315424 | 0.033077 | 0.269281 | 0.086717 | 0.907413 | 0.843224 | 0.135984 | 0.160138 | 0.111426 | -0.08193 | -0.15998 | -0.29784 | FALSE | FALSE | FALSE | FALSE | FALSE | 2.045931 | 1.419197 | 1.041553 | 2.04396 | 3.838681 |
| P01008 | SERPINC1 | 3.687444 | 0.00469 | -0.03029 | -0.05556 | -0.08791 | -0.11677 | 0.800475 | 0.000626 | 0.042053 | 0.001129 | 9.9E-07 | 0.878542 | 0.002646 | 0.345768 | 0.033414 | 2.44E-05 | 0.026758 | -0.17285 | -0.31701 | -0.50163 | -0.66627 | FALSE | TRUE | FALSE | TRUE | TRUE | 0.325585 | 2.122009 | 3.926145 | 6.28329 | 8.43036 |
| O95445 | APOM | 7.380478 | 0.027681 | -0.05441 | -0.01176 | -0.15878 | -0.21453 | 0.454177 | 0.002013 | 0.828512 | 0.003116 | 6.04E-06 | 0.628208 | 0.007641 | 0.959934 | 0.075088 | 0.000123 | 0.079876 | -0.15699 | -0.03392 | -0.45816 | -0.61904 | FALSE | TRUE | FALSE | FALSE | TRUE | 1.937246 | 3.843132 | 0.818232 | 11.63406 | 16.03281 |
| P01031 | C5 | 4.00853 | 0.031782 | 0.017025 | -0.06746 | 0.011138 | 0.005722 | 0.123312 | 0.081111 | 0.026176 | 0.707742 | 0.825691 | 0.228127 | 0.156754 | 0.255459 | 0.94573 | 0.947857 | 0.170313 | 0.091234 | -0.36152 | 0.059686 | 0.030662 | FALSE | FALSE | FALSE | FALSE | FALSE | 2.227406 | 1.187079 | 4.787197 | 0.775017 | 0.397399 |
| P01042-2 | KNG1 | 8.713785 | -0.02315 | -0.03065 | -0.04521 | 0.024683 | -0.07572 | 0.528581 | 0.078411 | 0.402131 | 0.641524 | 0.103098 | 0.680261 | 0.156754 | 0.783097 | 0.94573 | 0.293433 | -0.06997 | -0.09263 | -0.13661 | 0.074587 | -0.22882 | FALSE | FALSE | FALSE | FALSE | FALSE | 1.617915 | 2.147464 | 3.183058 | 1.725592 | 5.388727 |
| P06396;P0 | GSN | 5.238846 | 0.037322 | -0.00713 | 0.0232 | -0.07968 | -0.1206 | 0.249294 | 0.641416 | 0.625404 | 0.088709 | 0.003357 | 0.411317 | 0.741637 | 0.913703 | 0.525157 | 0.023658 | 0.126992 | -0.02426 | 0.078939 | -0.27111 | -0.41035 | FALSE | FALSE | FALSE | FALSE | TRUE | 2.620687 | 0.495456 | 1.621066 | 5.678062 | 8.718608 |
| P25311 | AZGP1 | 4.831434 | 0.194995 | 0.030633 | -0.02604 | -0.13024 | -0.13466 | 1.84E-12 | 0.015403 | 0.505133 | 0.000774 | 7.46E-05 | 9.05E-11 | 0.047713 | 0.869298 | 0.02862 | 0.00092 | 0.738637 | 0.116039 | -0.09865 | -0.49336 | -0.5101 | TRUE | TRUE | FALSE | TRUE | TRUE | 14.47201 | 2.146047 | 1.821548 | 9.447871 | 9.783674 |
| P22792 | CPN2 | 10.9367 | -0.12809 | -0.05134 | 0.088564 | -0.07746 | 0.032623 | 0.009364 | 0.027438 | 0.219164 | 0.27415 | 0.598175 | 0.030128 | 0.076619 | 0.606561 | 0.78094 | 0.804817 | -0.28631 | -0.11476 | 0.197956 | -0.17313 | 0.072918 | FALSE | FALSE | FALSE | FALSE | FALSE | 9.284828 | 3.623029 | 6.331118 | 5.515548 | 2.286996 |
| Q96PD5 | PGLYRP2 | 5.85233 | 0.02579 | 0.031117 | 0.009274 | 0.126868 | -0.0038 | 0.392306 | 0.029612 | 0.83395 | 0.003748 | 0.920324 | 0.577305 | 0.079684 | 0.959934 | 0.075088 | 0.986433 | 0.093718 | 0.113075 | 0.0337 | 0.46102 | -0.01382 | FALSE | FALSE | FALSE | FALSE | FALSE | 1.803711 | 2.180303 | 0.644889 | 9.192057 | 0.263998 |
| P19823 | ITIH2 | 4.289194 | -0.00185 | -0.01054 | -0.02315 | -0.03445 | -0.04113 | 0.92009 | 0.227197 | 0.392557 | 0.19589 | 0.077775 | 0.958446 | 0.357714 | 0.774645 | 0.700478 | 0.250231 | -0.01117 | -0.06369 | -0.13987 | -0.20813 | -0.2485 | FALSE | FALSE | FALSE | FALSE | FALSE | 0.128262 | 0.733347 | 1.617658 | 2.416539 | 2.892 |
| P02675 | FGB | 14.60239 | -0.09337 | 0.041309 | -0.00478 | -0.00244 | 0.062095 | 0.057652 | 0.075695 | 0.947098 | 0.972532 | 0.316159 | 0.135437 | 0.153463 | 0.978434 | 0.989577 | 0.598651 | -0.2114 | 0.093531 | -0.01082 | -0.00552 | 0.140592 | FALSE | FALSE | FALSE | FALSE | FALSE | 6.685807 | 2.904736 | 0.331711 | 0.169064 | 4.398046 |
| P02748 | C9 | 4.66574 | 0.005731 | 0.123002 | -0.14233 | -0.03929 | -0.19155 | 0.873378 | 3.41E-12 | 0.007344 | 0.449233 | 3.13E-05 | 0.929928 | 6.31E-11 | 0.155921 | 0.934694 | 0.000464 | 0.016099 | 0.345508 | -0.3998 | -0.11035 | -0.53805 | FALSE | TRUE | FALSE | FALSE | TRUE | 0.398047 | 8.899847 | 10.36876 | 2.760498 | 14.1989 |
| P05155;P0 | SERPING: | 8.237328 | 0.139246 | 0.035706 | 0.107816 | 0.078272 | 0.073343 | 0.005726 | 0.132534 | 0.143328 | 0.279372 | 0.246519 | 0.020427 | 0.233513 | 0.558227 | 0.78094 | 0.493039 | 0.305507 | 0.07834 | 0.236551 | 0.17173 | 0.160915 | TRUE | FALSE | FALSE | FALSE | FALSE | 10.1329 | 2.505838 | 7.759581 | 5.575257 | 5.215164 |
| P43652 | AFM | 4.787243 | 0.010841 | -0.06352 | 0.135651 | 0.204767 | 0.29079 | 0.751808 | 0.000108 | 0.007349 | 4.39E-05 | 7.93E-11 | 0.862539 | 0.000615 | 0.155921 | 0.006493 | 1.17E-08 | 0.03229 | -0.18919 | 0.404048 | 0.609918 | 0.866146 | FALSE | TRUE | FALSE | TRUE | TRUE | 0.754256 | 4.501004 | 9.858805 | 15.25003 | 22.33103 |
| P02679;P0 | FGG | 13.76115 | -0.09971 | 0.040261 | -0.0036 | -0.02144 | 0.045523 | 0.034725 | 0.071205 | 0.958453 | 0.752192 | 0.443572 | 0.096969 | 0.14663 | 0.978434 | 0.951491 | 0.73008 | -0.23503 | 0.094898 | -0.00848 | -0.05054 | 0.107301 | FALSE | FALSE | FALSE | FALSE | FALSE | 7.155833 | 2.82997 | 0.249779 | 1.497444 | 3.205719 |
| P01861 | IGHG4 | 6.241279 | 0.613661 | -0.02325 | 0.032155 | 0.060595 | -0.15274 | 1.18E-06 | 0.691819 | 0.859774 | 0.734858 | 0.329313 | 1.34E-05 | 0.78761 | 0.959934 | 0.94573 | 0.601709 | 0.535234 | -0.02028 | 0.028045 | 0.052851 | -0.13322 | TRUE | FALSE | FALSE | FALSE | FALSE | 53.01367 | 1.624446 | 2.253816 | 4.289554 | 11.16789 |
| P06727 | APOA4 | 3.520802 | 0.043957 | 0.027432 | 0.068023 | -0.007 | -0.04045 | 0.363937 | 0.231201 | 0.338615 | 0.92017 | 0.507861 | 0.555285 | 0.360187 | 0.736986 | 0.989577 | 0.731172 | 0.101299 | 0.063219 | 0.15676 | -0.01613 | -0.09322 | FALSE | FALSE | FALSE | FALSE | FALSE | 3.093738 | 1.919668 | 4.827898 | 0.486226 | 2.843386 |
| O14791;O | APOL1 | 7.431628 | -0.01624 | -0.15882 | 0.071671 | 0.147417 | -0.00223 | 0.709121 | 1.51E-13 | 0.262646 | 0.019475 | 0.967623 | 0.829548 | 3.72E-12 | 0.63724 | 0.221721 | 0.987963 | -0.0383 | -0.3745 | 0.168997 | 0.347601 | -0.00526 | FALSE | TRUE | FALSE | FALSE | FALSE | 1.132273 | 11.63773 | 5.093348 | 10.75848 | 0.154715 |
| P00751 | CFB | 3.749243 | -0.04042 | -0.01177 | -0.00072 | 0.096754 | 0.050544 | 0.12192 | 0.340254 | 0.985091 | 0.010539 | 0.125335 | 0.228127 | 0.493702 | 0.985091 | 0.155984 | 0.326552 | -0.17111 | -0.04984 | -0.00303 | 0.409614 | 0.213983 | FALSE | FALSE | FALSE | FALSE | FALSE | 2.841094 | 0.819414 | 0.049608 | 6.936481 | 3.565564 |
| P02750 | LRG1 | 5.418132 | -0.14678 | 0.110354 | -0.18672 | -0.0716 | -0.26654 | 0.000276 | 1.25E-08 | 0.001583 | 0.214749 | 2.26E-07 | 0.001858 | 1.23E-07 | 0.078082 | 0.722338 | 6.68E-06 | -0.36703 | 0.275937 | -0.46688 | -0.17904 | -0.66648 | TRUE | TRUE | FALSE | FALSE | TRUE | 10.7099 | 7.949299 | 13.81713 | 5.088256 | 20.29204 |
| P20851;P2 | C4BPB | 11.32209 | -0.14641 | 0.006415 | -0.1093 | -0.01559 | 0.021397 | 0.001923 | 0.772219 | 0.112709 | 0.817633 | 0.717446 | 0.009179 | 0.846827 | 0.490913 | 0.96269 | 0.88485 | -0.3443 | 0.015085 | -0.25704 | -0.03666 | 0.050318 | FALSE | FALSE | FALSE | FALSE | FALSE | 10.68108 | 0.445626 | 7.87049 | 1.086395 | 1.494149 |
| P13671 | C6 | 7.011245 | -0.18342 | 0.011972 | -0.06418 | -0.06141 | -0.04516 | 5.97E-08 | 0.444293 | 0.186834 | 0.198784 | 0.279687 | 7.37E-07 | 0.592391 | 0.583261 | 0.700478 | 0.53758 | -0.59124 | 0.038591 | -0.20687 | -0.19794 | -0.14557 | TRUE | FALSE | FALSE | FALSE | FALSE | 13.55728 | 0.833288 | 4.548854 | 4.34824 | 3.179807 |
| P01019 | AGT | 5.297532 | -0.07509 | -0.0904 | 0.023755 | -0.02513 | 0.034717 | 0.071843 | 6.18E-06 | 0.697414 | 0.675668 | 0.508698 | 0.156364 | 4.16E-05 | 0.926973 | 0.94573 | 0.731172 | -0.19481 | -0.23453 | 0.061626 | -0.06518 | 0.090063 | FALSE | TRUE | FALSE | FALSE | FALSE | 5.342866 | 6.466856 | 1.660201 | 1.756842 | 2.435557 |
| P35858;P3 | IGFALS | 10.06249 | -0.11905 | -0.1702 | -0.00965 | 0.014969 | -0.0318 | 0.00793 | 1.37E-14 | 0.882875 | 0.816063 | 0.572142 | 0.02608 | 4.05E-13 | 0.959934 | 0.96269 | 0.804817 | -0.27015 | -0.38624 | -0.02189 | 0.033969 | -0.07216 | FALSE | TRUE | FALSE | FALSE | FALSE | 8.601863 | 12.52173 | 0.670794 | 1.04299 | 2.228623 |
| P02671 | FGA | 15.53572 | -0.08809 | 0.045745 | -0.02739 | 0.003854 | 0.052119 | 0.099418 | 0.070575 | 0.726546 | 0.96007 | 0.439082 | 0.198835 | 0.14663 | 0.926973 | 0.989577 | 0.73008 | -0.18363 | 0.095363 | -0.05711 | 0.008035 | 0.108649 | FALSE | FALSE | FALSE | FALSE | FALSE | 6.295968 | 3.221633 | 1.916903 | 0.26752 | 3.678636 |
| P04114 | APOB | 4.949376 | 0.054908 | -0.04936 | 0.093196 | -0.00701 | 0.067496 | 0.179336 | 0.010998 | 0.120736 | 0.905268 | 0.190871 | 0.319781 | 0.037853 | 0.490913 | 0.989577 | 0.443111 | 0.148169 | -0.1332 | 0.251488 | -0.01892 | 0.182137 | FALSE | TRUE | FALSE | FALSE | FALSE | 3.879283 | 3.480674 | 6.673055 | 0.487271 | 4.789631 |
| P36955 | SERPINF1 | 6.541181 | 0.173219 | 0.028939 | -0.01794 | 0.074534 | 0.096 | 2.01E-08 | 0.042883 | 0.685047 | 0.087085 | 0.011964 | 2.98E-07 | 0.109346 | 0.926973 | 0.525157 | 0.06558 | 0.600518 | 0.100326 | -0.06218 | 0.258395 | 0.332816 | TRUE | FALSE | FALSE | FALSE | FALSE | 12.75715 | 2.026147 | 1.250993 | 5.302061 | 6.880633 |
| P07357 | C8A | 6.092441 | 0.000913 | -0.00913 | -0.05285 | 0.016781 | -0.02637 | 0.975264 | 0.512347 | 0.222249 | 0.693077 | 0.478267 | 0.987734 | 0.659802 | 0.606561 | 0.94573 | 0.731172 | 0.003466 | -0.03466 | -0.20055 | 0.063678 | -0.10008 | FALSE | FALSE | FALSE | FALSE | FALSE | 0.063338 | 0.635075 | 3.731404 | 1.169989 | 1.844964 |
| P02751 | FN1 | 30.54984 | -0.1695 | 0.004107 | 0.153002 | 0.024916 | 0.197123 | 0.006806 | 0.88912 | 0.094965 | 0.781609 | 0.012566 | 0.022893 | 0.920208 | 0.488242 | 0.956017 | 0.066421 | -0.29914 | 0.007248 | 0.270016 | 0.043971 | 0.347881 | FALSE | FALSE | FALSE | FALSE | FALSE | 12.4671 | 0.285087 | 11.18806 | 1.742043 | 14.64099 |
| P02765 | AHSG | 4.653773 | 0.03881 | -0.04896 | 0.025074 | -0.0655 | 0.001794 | 0.165152 | 0.000244 | 0.540849 | 0.104757 | 0.959402 | 0.298079 | 0.001232 | 0.885469 | 0.553717 | 0.987963 | 0.151371 | -0.19097 | 0.097796 | -0.25545 | 0.006996 | FALSE | TRUE | FALSE | FALSE | FALSE | 2.726633 | 3.452019 | 1.753198 | 4.644395 | 0.124406 |
| P00488 | F13A1 | 15.11746 | 0.076622 | 0.008338 | 0.007992 | -0.06479 | -0.13925 | 0.274484 | 0.801336 | 0.938058 | 0.521729 | 0.115929 | 0.441561 | 0.853221 | 0.977694 | 0.942137 | 0.323725 | 0.122091 | 0.013286 | 0.012735 | -0.10323 | -0.22189 | FALSE | FALSE | FALSE | FALSE | FALSE | 5.454596 | 0.579602 | 0.555504 | 4.592971 | 10.1334 |
| A0A0C4D | IGHV1-3 | 21.37678 | -0.03331 | 0.014844 | 0.191972 | 0.139084 | 0.297579 | 0.612932 | 0.633664 | 0.047696 | 0.143878 | 0.000391 | 0.743556 | 0.738443 | 0.352951 | 0.665437 | 0.003612 | -0.05562 | 0.024785 | 0.320533 | 0.232227 | 0.496866 | FALSE | FALSE | FALSE | FALSE | TRUE | 2.335937 | 1.034215 | 14.23238 | 10.12058 | 22.90804 |
| P01602 | IGKV1-5 | 7.286084 | 0.116451 | -0.01078 | -0.0533 | 0.038783 | -0.02267 | 0.060089 | 0.712139 | 0.556805 | 0.663531 | 0.771241 | 0.135529 | 0.804553 | 0.885469 | 0.94573 | 0.920514 | 0.210242 | -0.01947 | -0.09622 | 0.070019 | -0.04092 | FALSE | FALSE | FALSE | FALSE | FALSE | 8.406475 | 0.750123 | 3.763415 | 2.72468 | 1.583514 |
| P01619 | IGKV3-20 | 8.423756 | 0.001305 | -0.00657 | -0.13163 | 0.037464 | 0.085146 | 0.981075 | 0.800594 | 0.103886 | 0.637061 | 0.220617 | 0.987734 | 0.853221 | 0.490913 | 0.94573 | 0.475228 | 0.002637 | -0.01328 | -0.26595 | 0.075697 | 0.172038 | FALSE | FALSE | FALSE | FALSE | FALSE | 0.090496 | 0.456657 | 9.552777 | 2.630826 | 6.079491 |
| P10909;P1 | CLU | 3.13556 | 0.036475 | -0.03124 | -0.01413 | -0.05209 | -0.05061 | 0.037207 | 0.000181 | 0.581178 | 0.039188 | 0.022065 | 0.098192 | 0.000956 | 0.885469 | 0.341166 | 0.09896 | 0.225999 | -0.19358 | -0.08758 | -0.32278 | -0.31361 | FALSE | TRUE | FALSE | FALSE | FALSE | 2.560472 | 2.189203 | 0.984526 | 3.676922 | 3.570605 |
| P00746 | CFD | 13.28874 | 0.108421 | 0.259758 | -0.13798 | 0.03415 | 0.111639 | 0.102372 | 2.98E-15 | 0.156582 | 0.720912 | 0.182319 | 0.202013 | 1.44E-13 | 0.579354 | 0.94573 | 0.442348 | 0.165523 | 0.396565 | -0.21065 | 0.052136 | 0.170437 | FALSE | TRUE | FALSE | FALSE | FALSE | 7.804739 | 19.72776 | 10.03609 | 2.395337 | 8.04553 |
| P15814 | IGLL1 | 11.92332 | 0.098083 | 0.029917 | 0.086233 | 0.103151 | -0.01173 | 0.035499 | 0.174279 | 0.206968 | 0.12483 | 0.841492 | 0.097292 | 0.293106 | 0.606561 | 0.59834 | 0.947857 | 0.232951 | 0.071054 | 0.204808 | 0.244989 | -0.02787 | FALSE | FALSE | FALSE | FALSE | FALSE | 7.03503 | 2.095339 | 6.159501 | 7.411725 | 0.816606 |
| P00747 | PLG | 3.260475 | -0.0115 | -0.05415 | -0.04889 | -0.01891 | -0.03449 | 0.582346 | 8.5E-08 | 0.111762 | 0.530678 | 0.191615 | 0.718226 | 7.4E-07 | 0.490913 | 0.942137 | 0.443111 | -0.05859 | -0.27602 | -0.24919 | -0.0964 | -0.17577 | FALSE | TRUE | FALSE | FALSE | FALSE | 0.800017 | 3.82498 | 3.446808 | 1.31963 | 2.419183 |
| P08571 | CD14 | 18.39787 | 0.069159 | 0.009326 | -0.13819 | -0.05753 | -0.13197 | 0.310754 | 0.772444 | 0.167994 | 0.558779 | 0.125767 | 0.494533 | 0.846827 | 0.583261 | 0.942137 | 0.326552 | 0.113188 | 0.015263 | -0.22617 | -0.09416 | -0.21598 | FALSE | FALSE | FALSE | FALSE | FALSE | 4.910484 | 0.648508 | 10.05255 | 4.068497 | 9.578631 |
| P02749 | APOH | 9.273493 | 0.104971 | 0.051521 | 0.026179 | -0.02229 | 0.053071 | 0.014991 | 0.011629 | 0.678082 | 0.71919 | 0.327934 | 0.047083 | 0.039115 | 0.926973 | 0.94573 | 0.601709 | 0.268491 | 0.131777 | 0.066959 | -0.05701 | 0.135743 | FALSE | TRUE | FALSE | FALSE | FALSE | 7.547287 | 3.635664 | 1.831132 | 1.556894 | 3.747105 |
| P04217;P0 | A1BG | 5.30256 | -0.13583 | 0.01699 | -0.02468 | -0.03983 | -0.03959 | 3.64E-10 | 0.088018 | 0.423779 | 0.189509 | 0.136072 | 8.97E-09 | 0.164895 | 0.787894 | 0.700478 | 0.347219 | -0.67539 | 0.084479 | -0.12272 | -0.19803 | -0.19684 | TRUE | FALSE | FALSE | FALSE | FALSE | 9.872843 | 1.18465 | 1.72556 | 2.799172 | 2.782015 |
| P00738 | HP | 5.034891 | -0.04355 | 0.028159 | 0.008539 | 0.167269 | 0.09544 | 0.500392 | 0.357152 | 0.928267 | 0.073522 | 0.242307 | 0.656917 | 0.500643 | 0.974352 | 0.473098 | 0.493039 | -0.0751 | 0.048552 | 0.014723 | 0.288407 | 0.16456 | FALSE | FALSE | FALSE | FALSE | FALSE | 3.064928 | 1.971015 | 0.593648 | 12.29306 | 6.839155 |
| P06681 | C2 | 10.68615 | 0.057574 | 0.025721 | -0.0036 | 0.042463 | 0.0216 | 0.222879 | 0.249588 | 0.9586 | 0.532954 | 0.716811 | 0.388072 | 0.380814 | 0.978434 | 0.942137 | 0.88485 | 0.136295 | 0.060889 | -0.00851 | 0.100523 | 0.051133 | FALSE | FALSE | FALSE | FALSE | FALSE | 4.071395 | 1.798799 | 0.249534 | 2.987037 | 1.508431 |
| P05543 | SERPINA' | 9.138962 | -0.08499 | -0.01245 | -0.00997 | -0.01363 | 0.027169 | 0.080366 | 0.587244 | 0.888588 | 0.845618 | 0.657127 | 0.169917 | 0.715539 | 0.959934 | 0.96269 | 0.845694 | -0.1957 | -0.02868 | -0.02296 | -0.03137 | 0.062559 | FALSE | FALSE | FALSE | FALSE | FALSE | 6.068219 | 0.866969 | 0.693698 | 0.948932 | 1.901053 |
| P01009 | SERPINA: | 3.97514 | -0.02493 | 0.018299 | -0.02441 | 0.041712 | -0.03532 | 0.428556 | 0.219603 | 0.597377 | 0.358787 | 0.374242 | 0.598361 | 0.349802 | 0.893048 | 0.849036 | 0.667323 | -0.08822 | 0.064754 | -0.08639 | 0.147604 | -0.12499 | FALSE | FALSE | FALSE | FALSE | FALSE | 1.743124 | 1.276467 | 1.706601 | 2.933425 | 2.478399 |
| P04004 | VTN | 4.276639 | 0.041273 | -0.06996 | 0.027332 | 0.123325 | 0.16674 | 0.097658 | 6.89E-09 | 0.454219 | 0.00066 | 1.97E-07 | 0.197991 | 7.85E-08 | 0.81981 | 0.02862 | 6.68E-06 | 0.16724 | -0.2835 | 0.110752 | 0.499719 | 0.675637 | FALSE | TRUE | FALSE | TRUE | TRUE | 2.902136 | 4.969012 | 1.912596 | 8.924235 | 12.25188 |
| P05154 | SERPINA: | 19.87533 | 0.17966 | -0.01992 | 0.017595 | 0.005009 | 0.136662 | 0.033392 | 0.616879 | 0.886756 | 0.967103 | 0.19876 | 0.095038 | 0.736275 | 0.959934 | 0.989577 | 0.450197 | 0.236985 | -0.02627 | 0.023209 | 0.006607 | 0.180268 | FALSE | FALSE | FALSE | FALSE | FALSE | 13.26167 | 1.390197 | 1.227062 | 0.347767 | 9.935879 |
| P01859 | IGHG2 | 4.035425 | 0.188962 | -0.0619 | 0.037029 | 0.042525 | 0.0546 | 0.000912 | 0.021053 | 0.655293 | 0.601983 | 0.443968 | 0.00519 | 0.061094 | 0.926973 | 0.942137 | 0.73008 | 0.365591 | -0.11976 | 0.071642 | 0.082275 | 0.105637 | TRUE | FALSE | FALSE | FALSE | FALSE | 13.99431 | 4.384093 | 2.599896 | 2.991503 | 3.857136 |
| P08603 | CFH | 3.964787 | 0.023917 | -0.00597 | 0.030707 | 0.100466 | 0.113553 | 0.227014 | 0.523168 | 0.290608 | 0.000488 | 7.5E-06 | 0.390676 | 0.666133 | 0.672032 | 0.02862 | 0.000123 | 0.129942 | -0.03246 | 0.166835 | 0.545841 | 0.616943 | FALSE | FALSE | FALSE | TRUE | TRUE | 1.671611 | 0.414944 | 2.151277 | 7.211986 | 8.188955 |
| P04003 | C4BPA | 4.31948 | -0.17485 | -0.01185 | -0.00189 | -0.04039 | 0.020278 | 1.93E-08 | 0.409339 | 0.966129 | 0.356857 | 0.596734 | 2.98E-07 | 0.550747 | 0.979363 | 0.849036 | 0.804817 | -0.61373 | -0.04161 | -0.00664 | -0.14178 | 0.071174 | TRUE | FALSE | FALSE | FALSE | FALSE | 12.88496 | 0.825102 | 0.131281 | 2.839474 | 1.415469 |
| P01860 | IGHG3 | 8.561555 | 0.158826 | 0.096965 | 0.128136 | -0.03582 | -0.0594 | 0.060438 | 0.015565 | 0.301249 | 0.768608 | 0.576997 | 0.135529 | 0.047713 | 0.675529 | 0.95433 | 0.804817 | 0.207492 | 0.126676 | 0.167398 | -0.04679 | -0.0776 | FALSE | TRUE | FALSE | FALSE | FALSE | 11.63786 | 6.952143 | 9.288085 | 2.51369 | 4.203063 |
| P02790 | HPX | 3.94387 | -0.06873 | 0.005178 | 0.009672 | 0.012477 | 0.038508 | 0.000591 | 0.580838 | 0.739628 | 0.662731 | 0.124393 | 0.003801 | 0.715539 | 0.927669 | 0.94573 | 0.326552 | -0.38139 | 0.028732 | 0.053668 | 0.069232 | 0.213678 | TRUE | FALSE | FALSE | FALSE | FALSE | 4.879533 | 0.359556 | 0.672652 | 0.868575 | 2.705126 |
| P01591 | JCHAIN | 8.013247 | 0.046332 | 0.021293 | -0.1284 | -0.0489 | -0.05845 | 0.493495 | 0.505836 | 0.196584 | 0.616403 | 0.493632 | 0.656917 | 0.659802 | 0.593763 | 0.94573 | 0.731172 | 0.076623 | 0.035215 | -0.21234 | -0.08088 | -0.09667 | FALSE | FALSE | FALSE | FALSE | FALSE | 3.263594 | 1.486894 | 9.307746 | 3.44794 | 4.134938 |
| P02656 | APOC3 | 4.536047 | -0.05076 | -0.06066 | 0.051887 | 0.046313 | 0.133547 | 0.24399 | 0.003422 | 0.416954 | 0.461031 | 0.0155 | 0.410347 | 0.01266 | 0.787894 | 0.934694 | 0.079105 | -0.12774 | -0.15266 | 0.130589 | 0.116559 | 0.336109 | FALSE | TRUE | FALSE | FALSE | FALSE | 3.580715 | 4.294103 | 3.662003 | 3.262219 | 9.698718 |
| P01877 | IGHA2 | 5.89438 | 0.422429 | 0.029359 | -0.03718 | 0.126837 | -0.05867 | 4.18E-09 | 0.375204 | 0.717359 | 0.20949 | 0.50645 | 8.83E-08 | 0.509452 | 0.926973 | 0.721034 | 0.731172 | 0.639172 | 0.044423 | -0.05625 | 0.191915 | -0.08878 | TRUE | FALSE | FALSE | FALSE | FALSE | 34.01821 | 2.055849 | 2.610516 | 9.189694 | 4.15063 |
| P01876 | IGHA1 | 6.57185 | 0.316006 | 0.061715 | 0.091363 | 0.030297 | -0.00911 | 8.25E-06 | 0.062226 | 0.372792 | 0.763531 | 0.917598 | 8.72E-05 | 0.135653 | 0.745583 | 0.95433 | 0.986433 | 0.489383 | 0.095575 | 0.141489 | 0.046919 | -0.01411 | TRUE | FALSE | FALSE | FALSE | FALSE | 24.48796 | 4.37058 | 6.537594 | 2.122237 | 0.633551 |
| P01857 | IGHG1 | 6.131651 | -0.02312 | 0.036128 | -0.02258 | 0.061699 | 0.001106 | 0.598564 | 0.082613 | 0.726139 | 0.330453 | 0.984071 | 0.732128 | 0.156754 | 0.926973 | 0.843224 | 0.988572 | -0.05869 | 0.091723 | -0.05732 | 0.156645 | 0.002808 | FALSE | FALSE | FALSE | FALSE | FALSE | 1.615242 | 2.535785 | 1.57722 | 4.369391 | 0.0767 |
| P01871 | IGHM | 5.475662 | -0.27863 | -0.10415 | -0.24791 | -0.04357 | -0.12935 | 0.000971 | 0.008917 | 0.044433 | 0.718512 | 0.221559 | 0.005321 | 0.032189 | 0.346108 | 0.94573 | 0.475228 | -0.36022 | -0.13465 | -0.3205 | -0.05632 | -0.16722 | TRUE | TRUE | FALSE | FALSE | FALSE | 21.30445 | 7.486303 | 18.74859 | 3.065808 | 9.379992 |
| P00734 | F2 | 3.510323 | -0.0759 | -0.1093 | 0.009804 | -0.04497 | -0.04618 | 0.006612 | 2.51E-15 | 0.810086 | 0.262549 | 0.188441 | 0.022759 | 1.44E-13 | 0.959934 | 0.78094 | 0.443111 | -0.27399 | -0.39455 | 0.035392 | -0.16234 | -0.16672 | TRUE | TRUE | FALSE | FALSE | FALSE | 5.401642 | 7.870103 | 0.681862 | 3.166112 | 3.252988 |
| A0A0J9Y: | IGHV3-64 | 9.656367 | 0.011382 | -0.05345 | -0.01661 | -0.04352 | -0.05413 | 0.843866 | 0.051214 | 0.844809 | 0.601822 | 0.45816 | 0.905016 | 0.120312 | 0.959934 | 0.942137 | 0.731172 | 0.021963 | -0.10315 | -0.03205 | -0.08398 | -0.10445 | FALSE | FALSE | FALSE | FALSE | FALSE | 0.792058 | 3.774574 | 1.157819 | 3.062731 | 3.823312 |
| P02753 | RBP4 | 4.74165 | 0.200785 | -0.03374 | 0.053872 | -0.01146 | -0.02392 | 2.66E-08 | 0.04359 | 0.298017 | 0.821603 | 0.590603 | 3.58E-07 | 0.109346 | 0.675529 | 0.96269 | 0.804817 | 0.605951 | -0.10181 | 0.162581 | -0.0346 | -0.0722 | TRUE | FALSE | FALSE | FALSE | FALSE | 14.93239 | 2.365932 | 3.804727 | 0.797784 | 1.672124 |
| P02746 | C1QB | 6.563073 | 0.119608 | 0.027463 | -0.08001 | -0.0036 | -0.06942 | 0.000161 | 0.06471 | 0.082835 | 0.936652 | 0.080013 | 0.001324 | 0.138799 | 0.488242 | 0.989577 | 0.251955 | 0.414201 | 0.095102 | -0.27706 | -0.01245 | -0.2404 | TRUE | FALSE | FALSE | FALSE | FALSE | 8.643981 | 1.921793 | 5.702368 | 0.24951 | 4.929511 |
| P08697 | SERPINF2 | 6.155718 | -0.07825 | -0.08665 | 0.066392 | 0.02407 | 0.03682 | 0.002946 | 1.37E-11 | 0.084307 | 0.523492 | 0.264779 | 0.012184 | 2.13E-10 | 0.488242 | 0.942137 | 0.515623 | -0.30825 | -0.34135 | 0.26153 | 0.094816 | 0.145041 | FALSE | TRUE | FALSE | FALSE | FALSE | 5.573819 | 6.190424 | 4.709474 | 1.682396 | 2.585008 |
| P06310 | IGKV2-30 | 8.121169 | 0.038594 | -0.05046 | -0.15469 | -0.09947 | -0.09276 | 0.541846 | 0.092384 | 0.096459 | 0.276228 | 0.245679 | 0.69132 | 0.170911 | 0.488242 | 0.78094 | 0.493039 | 0.067757 | -0.08859 | -0.27157 | -0.17463 | -0.16286 | FALSE | FALSE | FALSE | FALSE | FALSE | 2.711255 | 3.559574 | 11.31789 | 7.138129 | 6.641131 |
| P0C0L4 | C4A | 9.393978 | 0.067879 | 0.065846 | 0.069887 | 0.043315 | -0.0198 | 0.394209 | 0.081138 | 0.550056 | 0.706232 | 0.843789 | 0.577305 | 0.156754 | 0.885469 | 0.94573 | 0.947857 | 0.095007 | 0.092161 | 0.097817 | 0.060625 | -0.02772 | FALSE | FALSE | FALSE | FALSE | FALSE | 4.817477 | 4.669841 | 4.963444 | 3.04786 | 1.382034 |
| P03952 | KLKB1 | 6.576317 | -0.13681 | -0.07857 | 0.089975 | 0.05034 | -0.07905 | 2.26E-05 | 3.19E-07 | 0.054896 | 0.273603 | 0.049788 | 0.000209 | 2.62E-06 | 0.386886 | 0.78094 | 0.193911 | -0.44477 | -0.25542 | 0.2925 | 0.163649 | -0.25699 | TRUE | TRUE | FALSE | FALSE | FALSE | 9.947392 | 5.597066 | 6.435195 | 3.550863 | 5.632327 |
| P07358 | C8B | 8.845528 | 0.025166 | 0.001728 | 0.072544 | 0.030439 | 0.099068 | 0.4902 | 0.920172 | 0.175945 | 0.563 | 0.031928 | 0.656917 | 0.945732 | 0.583261 | 0.942137 | 0.135009 | 0.076791 | 0.005274 | 0.22136 | 0.092882 | 0.302297 | FALSE | FALSE | FALSE | FALSE | FALSE | 1.759671 | 0.119871 | 5.156912 | 2.132289 | 7.108147 |
| P02654 | APOC1 | 15.69465 | 0.202722 | -0.10272 | 0.169718 | -0.04609 | -0.13249 | 0.001118 | 0.000489 | 0.061648 | 0.60475 | 0.089532 | 0.00591 | 0.002192 | 0.414722 | 0.942137 | 0.268307 | 0.353605 | -0.17917 | 0.296036 | -0.08039 | -0.23109 | FALSE | TRUE | FALSE | FALSE | FALSE | 15.08679 | 7.379522 | 12.48383 | 3.24603 | 9.618134 |
| P02774;P0 | GC | 3.065501 | 0.021117 | -0.05095 | -0.01622 | -0.04983 | -0.13139 | 0.232369 | 2.98E-09 | 0.531784 | 0.051282 | 9.28E-09 | 0.395294 | 3.68E-08 | 0.885469 | 0.379489 | 6.87E-07 | 0.121696 | -0.29365 | -0.09347 | -0.28715 | -0.75718 | FALSE | TRUE | FALSE | FALSE | TRUE | 1.474508 | 3.595029 | 1.130556 | 3.514066 | 9.534871 |
| P68871 | HBB | 8.566037 | 0.192152 | -0.00849 | 0.053397 | 0.080897 | 0.055121 | 0.003696 | 0.784823 | 0.580276 | 0.394194 | 0.506711 | 0.014397 | 0.853221 | 0.885469 | 0.874176 | 0.731172 | 0.323168 | -0.01428 | 0.089805 | 0.136056 | 0.092704 | TRUE | FALSE | FALSE | FALSE | FALSE | 14.24664 | 0.590433 | 3.770555 | 5.767573 | 3.894591 |
| P23142 | FBLN1 | 17.24096 | 0.022653 | 0.131108 | 0.020139 | -0.00627 | -0.03271 | 0.748721 | 0.000107 | 0.846188 | 0.951026 | 0.714005 | 0.862539 | 0.000615 | 0.959934 | 0.989577 | 0.88485 | 0.035078 | 0.20302 | 0.031185 | -0.0097 | -0.05065 | FALSE | TRUE | FALSE | FALSE | FALSE | 1.58257 | 9.513444 | 1.40572 | 0.435365 | 2.29313 |
| Q9HCU4 | CELSR2 | 22.87162 | -0.02209 | 0.074621 | -0.1833 | -0.04631 | -0.14805 | 0.781179 | 0.047976 | 0.117146 | 0.686639 | 0.140869 | 0.875868 | 0.116402 | 0.490913 | 0.94573 | 0.353367 | -0.03083 | 0.104142 | -0.25581 | -0.06462 | -0.20662 | FALSE | FALSE | FALSE | FALSE | FALSE | 1.543169 | 5.308434 | 13.54751 | 3.261695 | 10.80715 |
| Q16610;Q | ECM1 | 11.9559 | 0.012066 | 0.000302 | 0.111779 | 0.009123 | 0.006965 | 0.801498 | 0.989389 | 0.113348 | 0.895206 | 0.908454 | 0.878542 | 0.996119 | 0.490913 | 0.989577 | 0.986433 | 0.028111 | 0.000703 | 0.260405 | 0.021253 | 0.016226 | FALSE | FALSE | FALSE | FALSE | FALSE | 0.839892 | 0.02093 | 8.05597 | 0.634341 | 0.483939 |

| | | | | | | | | | | | | | | | | | | | | | | | | | | | | | | | | |
|---|---|---|---|---|---|---|---|---|---|---|---|---|---|---|---|---|---|---|---|---|---|---|---|---|---|---|---|---|---|---|---|---|
| P02652 | APOA2 | 4.086108 | 0.057751 | -0.04767 | 0.022317 | -0.03588 | -0.05325 | 0.005695 | 1.87E-06 | 0.464294 | 0.231586 | 0.042817 | 0.020427 | 1.38E-05 | 0.827897 | 0.761659 | 0.171268 | 0.296515 | -0.24473 | 0.114582 | -0.18424 | -0.27339 | FALSE | TRUE | FALSE | FALSE | FALSE | 4.084221 | 3.359101 | 1.558906 | 2.518513 | 3.759755 |
| P00748 | F12 | 5.827374 | 0.004494 | -0.07593 | 0.151826 | 0.074408 | 0.058528 | 0.926114 | 0.001021 | 0.033468 | 0.287768 | 0.33894 | 0.958446 | 0.004085 | 0.291367 | 0.788699 | 0.611745 | 0.010186 | -0.1721 | 0.34411 | 0.168643 | 0.132652 | FALSE | TRUE | FALSE | FALSE | FALSE | 0.311998 | 5.404099 | 11.09748 | 5.292868 | 4.14026 |
| P27169 | PON1 | 5.658883 | -0.01155 | -0.12573 | -0.02511 | 0.004308 | -0.10131 | 0.807308 | 4.16E-08 | 0.717877 | 0.949721 | 0.090644 | 0.878542 | 3.85E-07 | 0.926973 | 0.989577 | 0.268307 | -0.02604 | -0.28352 | -0.05663 | 0.009714 | -0.22846 | FALSE | TRUE | FALSE | FALSE | FALSE | 0.803541 | 9.105775 | 1.755874 | 0.299024 | 7.274843 |
| P80108 | GPLD1 | 9.234715 | 0.358575 | -0.03489 | -0.07527 | -0.1215 | -0.03808 | 1.32E-14 | 0.097457 | 0.248777 | 0.058667 | 0.497058 | 1.96E-12 | 0.176205 | 0.63481 | 0.413466 | 0.731172 | 0.818244 | -0.07961 | -0.17176 | -0.27726 | -0.08689 | TRUE | FALSE | FALSE | FALSE | FALSE | 28.21589 | 2.447731 | 5.355949 | 8.786728 | 2.674351 |
| P51884 | LUM | 7.285832 | 0.023423 | 0.136714 | -0.0464 | -0.02705 | -0.00894 | 0.501565 | 2.89E-15 | 0.364688 | 0.590656 | 0.838843 | 0.656917 | 1.44E-13 | 0.745583 | 0.942137 | 0.947857 | 0.068403 | 0.399253 | -0.1355 | -0.079 | -0.02612 | FALSE | TRUE | FALSE | FALSE | FALSE | 1.636813 | 9.939838 | 3.268307 | 1.892703 | 0.621809 |
| P02747 | C1QC | 5.284922 | 0.066304 | 0.023408 | -0.04159 | 0.025494 | 0.006798 | 0.028443 | 0.10141 | 0.34769 | 0.55797 | 0.858194 | 0.084191 | 0.180828 | 0.739805 | 0.942137 | 0.947857 | 0.243433 | 0.08594 | -0.1527 | 0.093601 | 0.024959 | FALSE | FALSE | FALSE | FALSE | FALSE | 4.703089 | 1.635723 | 2.924884 | 1.782833 | 0.472323 |
| A0A075B | IGLV8-61 | 7.802192 | 0.136794 | -0.05229 | -0.11292 | -0.01514 | -0.12398 | 0.10459 | 0.189387 | 0.360825 | 0.900733 | 0.243345 | 0.203676 | 0.314936 | 0.745583 | 0.989577 | 0.493039 | 0.180959 | -0.06917 | -0.14937 | -0.02003 | -0.164 | FALSE | FALSE | FALSE | FALSE | FALSE | 9.945951 | 3.69062 | 8.14111 | 1.054987 | 8.973485 |
| A0A075B | IGLV4-69 | 17.07488 | 0.082661 | -0.03906 | 0.049242 | -0.07329 | 0.035944 | 0.373753 | 0.374284 | 0.718025 | 0.584575 | 0.759122 | 0.558742 | 0.509452 | 0.926973 | 0.942137 | 0.920514 | 0.099567 | -0.04704 | 0.059313 | -0.08828 | 0.043296 | FALSE | FALSE | FALSE | FALSE | FALSE | 5.896932 | 2.744148 | 3.472115 | 5.211425 | 2.52277 |
| P05546 | SERPIND | 3.997182 | -0.06499 | -0.08693 | 0.103202 | 0.11089 | 0.129421 | 0.037232 | 8.48E-09 | 0.024325 | 0.013901 | 0.001073 | 0.098192 | 8.97E-08 | 0.255459 | 0.187028 | 0.008819 | -0.21765 | -0.29112 | 0.345611 | 0.371357 | 0.433416 | FALSE | TRUE | FALSE | FALSE | TRUE | 4.607823 | 6.210691 | 7.415468 | 7.989394 | 9.385447 |
| P22352 | GPX3 | 11.71894 | 0.067291 | -0.08645 | 0.008138 | -0.10507 | -0.16284 | 0.217973 | 0.000897 | 0.919076 | 0.182644 | 0.018535 | 0.384048 | 0.003686 | 0.971595 | 0.700478 | 0.087701 | 0.134729 | -0.17309 | 0.016293 | -0.21036 | -0.32603 | FALSE | TRUE | FALSE | FALSE | FALSE | 4.774766 | 6.175362 | 0.565662 | 7.554428 | 11.9488 |
| P06396 | GSN | 14.14976 | 0.096011 | 0.05092 | -0.01894 | -0.05658 | -0.10841 | 0.055517 | 0.031926 | 0.796403 | 0.433137 | 0.086512 | 0.132525 | 0.084375 | 0.959934 | 0.929048 | 0.266744 | 0.211894 | 0.112378 | -0.0418 | -0.12487 | -0.23925 | FALSE | FALSE | FALSE | FALSE | FALSE | 6.88143 | 3.592501 | 1.321438 | 3.99986 | 7.803769 |
| P09871 | C1S | 5.821158 | 0.04717 | 0.012536 | -0.04649 | 0.019254 | -0.05847 | 0.061928 | 0.293333 | 0.209436 | 0.59655 | 0.066673 | 0.136796 | 0.434133 | 0.606561 | 0.942137 | 0.234944 | 0.206996 | 0.055013 | -0.20401 | 0.084493 | -0.2566 | FALSE | FALSE | FALSE | FALSE | FALSE | 3.32361 | 0.872733 | 3.274958 | 1.343545 | 4.136419 |
| P05156 | CFI | 5.653273 | -0.08285 | 0.014829 | 0.090693 | 0.0253 | 0.086929 | 0.001253 | 0.218916 | 0.015762 | 0.491393 | 0.007165 | 0.006397 | 0.349802 | 0.233271 | 0.942137 | 0.042414 | -0.35365 | 0.0633 | 0.387139 | 0.107998 | 0.37107 | TRUE | FALSE | FALSE | FALSE | TRUE | 5.910752 | 1.033173 | 6.488189 | 1.769153 | 6.210694 |
| A0A0B4J1 | IGHV6-1 | 7.593105 | 0.106646 | 0.024187 | 0.173246 | 0.282393 | 0.088405 | 0.471731 | 0.729965 | 0.425889 | 0.187076 | 0.636348 | 0.646447 | 0.818446 | 0.787894 | 0.700478 | 0.826136 | 0.080504 | 0.018258 | 0.130778 | 0.213169 | 0.066734 | FALSE | FALSE | FALSE | FALSE | FALSE | 7.672212 | 1.690672 | 12.75926 | 21.62104 | 6.319412 |
| A0A0J9Y | IGHV5-10 | 9.62165 | 0.142272 | -0.07248 | -0.74416 | 0.266421 | -0.2478 | 0.397873 | 0.362552 | 0.002765 | 0.272792 | 0.243553 | 0.577305 | 0.501474 | 0.102321 | 0.78094 | 0.493039 | 0.092747 | -0.04725 | -0.48511 | 0.173679 | -0.16154 | FALSE | FALSE | FALSE | FALSE | FALSE | 10.3642 | 5.152573 | 67.49943 | 20.28199 | 18.73911 |
| P01780 | IGHV3-7 | 11.72861 | -0.06863 | -0.11526 | 0.085884 | 0.077483 | 0.142847 | 0.272925 | 0.000117 | 0.349908 | 0.390857 | 0.071032 | 0.441561 | 0.000644 | 0.739805 | 0.874176 | 0.244484 | -0.11953 | -0.20075 | 0.149586 | 0.134954 | 0.2488 | FALSE | TRUE | FALSE | FALSE | FALSE | 4.871956 | 8.317131 | 6.133786 | 5.517573 | 10.40821 |
| A0A0A0M | IGHV3-49 | 11.65333 | 0.113385 | 0.007304 | -0.05232 | 0.085287 | -0.00955 | 0.083298 | 0.813113 | 0.585395 | 0.365855 | 0.907799 | 0.173635 | 0.859577 | 0.885469 | 0.849036 | 0.986433 | 0.193522 | 0.012467 | -0.0893 | 0.145566 | -0.01629 | FALSE | FALSE | FALSE | FALSE | FALSE | 8.176309 | 0.507587 | 3.693372 | 6.089906 | 0.663944 |
| P00736 | C1R | 7.556028 | 0.050405 | 0.012679 | -0.00638 | 0.055444 | -0.00855 | 0.084793 | 0.358568 | 0.881582 | 0.188498 | 0.816449 | 0.174296 | 0.500643 | 0.959934 | 0.700478 | 0.947857 | 0.192093 | 0.048321 | -0.02432 | 0.211299 | -0.03257 | FALSE | FALSE | FALSE | FALSE | FALSE | 3.555551 | 0.882734 | 0.443286 | 3.917921 | 0.59423 |
| Q06033;Q | ITIH3 | 12.0474 | -0.14937 | 0.210083 | -0.38961 | -0.00708 | -0.26054 | 0.005797 | 3.89E-15 | 1.29E-06 | 0.927351 | 0.000149 | 0.020427 | 1.44E-13 | 0.000191 | 0.989577 | 0.001573 | -0.26901 | 0.378342 | -0.70166 | -0.01275 | -0.46921 | FALSE | TRUE | TRUE | FALSE | TRUE | 10.90871 | 15.67549 | 31.00425 | 0.492041 | 19.79288 |
| P08519 | LPA | 28.25457 | -0.324 | 0.113161 | -0.10606 | 0.026009 | 0.111143 | 0.109636 | 0.237063 | 0.720789 | 0.928945 | 0.663088 | 0.210728 | 0.365473 | 0.926973 | 0.989577 | 0.846009 | -0.17888 | 0.062478 | -0.05856 | 0.01436 | 0.061364 | FALSE | FALSE | FALSE | FALSE | FALSE | 25.17948 | 8.159555 | 7.628739 | 1.81914 | 8.008351 |
| P08185 | SERPINA | 8.529931 | -0.14989 | -0.06755 | -0.01771 | -0.00412 | -0.13622 | 0.000185 | 0.000362 | 0.760904 | 0.942587 | 0.006797 | 0.001438 | 0.001675 | 0.946335 | 0.989577 | 0.041913 | -0.40204 | -0.18119 | -0.04751 | -0.01105 | -0.36536 | TRUE | TRUE | FALSE | FALSE | TRUE | 10.94879 | 4.793745 | 1.235444 | 0.286037 | 9.902053 |
| P04217 | A1BG | 4.678584 | -0.12586 | 0.021321 | -0.06614 | -0.03622 | -0.06641 | 2.16E-10 | 0.019416 | 0.019482 | 0.191802 | 0.006442 | 6.38E-09 | 0.057472 | 0.255459 | 0.700478 | 0.041455 | -0.67419 | 0.114208 | -0.35428 | -0.194 | -0.3557 | TRUE | FALSE | FALSE | FALSE | TRUE | 9.116128 | 1.488865 | 4.691248 | 2.542159 | 4.710523 |
| P04278 | SHBG | 17.35827 | -0.30944 | 0.085542 | -0.25388 | -0.39739 | -0.49393 | 0.003041 | 0.081895 | 0.096125 | 0.008264 | 0.000189 | 0.012184 | 0.156754 | 0.488242 | 0.135896 | 0.001866 | -0.31992 | 0.088439 | -0.26247 | -0.41085 | -0.51066 | TRUE | FALSE | FALSE | FALSE | TRUE | 23.92278 | 6.108655 | 19.24059 | 31.71268 | 40.82792 |
| P15169 | CPN1 | 10.10741 | -0.17779 | -0.04198 | -0.09147 | -0.02215 | 0.028831 | 0.000206 | 0.061787 | 0.189166 | 0.745972 | 0.629802 | 0.001523 | 0.135653 | 0.583261 | 0.951491 | 0.824873 | -0.40832 | -0.09641 | -0.21007 | -0.05088 | 0.066214 | TRUE | FALSE | FALSE | FALSE | FALSE | 13.1149 | 2.952577 | 6.545502 | 1.547469 | 2.018495 |
| P05090 | APOD | 6.557527 | 0.312164 | 0.033894 | -0.07452 | -0.18015 | -0.22682 | 3.39E-12 | 0.097627 | 0.240122 | 0.004059 | 3.98E-05 | 1.26E-10 | 0.176205 | 0.630184 | 0.075088 | 0.000535 | 0.735229 | 0.079829 | -0.17552 | -0.42431 | -0.53422 | TRUE | FALSE | FALSE | FALSE | TRUE | 24.15685 | 2.377164 | 5.30121 | 13.30053 | 17.02523 |
| P10643 | C7 | 8.942739 | 0.000729 | 0.11265 | -0.24886 | -0.13497 | -0.1126 | 0.987734 | 8.43E-07 | 0.000403 | 0.049355 | 0.060735 | 0.987734 | 6.57E-06 | 0.029827 | 0.379489 | 0.219238 | 0.001637 | 0.252771 | -0.55841 | -0.30286 | -0.25266 | FALSE | TRUE | TRUE | FALSE | FALSE | 0.05057 | 8.12128 | 18.82688 | 9.807298 | 8.117557 |
| Q14624 | ITIH4 | 7.645648 | 0.01823 | 0.038822 | -0.11949 | -0.04631 | -0.09896 | 0.574945 | 0.011972 | 0.012698 | 0.323749 | 0.016316 | 0.715058 | 0.039375 | 0.208807 | 0.843224 | 0.080492 | 0.06153 | 0.131036 | -0.4033 | -0.15633 | -0.334 | FALSE | TRUE | FALSE | FALSE | FALSE | 1.2716 | 2.727471 | 8.634777 | 3.262361 | 7.099771 |
| P02763 | ORM1 | 5.802807 | -0.00198 | 0.037532 | -0.07535 | 0.010325 | -0.08514 | 0.963963 | 0.071334 | 0.242706 | 0.870504 | 0.12495 | 0.983907 | 0.14663 | 0.630184 | 0.976019 | 0.326552 | -0.00502 | 0.094981 | -0.1907 | 0.026129 | -0.21546 | FALSE | FALSE | FALSE | FALSE | FALSE | 0.13754 | 2.635663 | 5.361968 | 0.718228 | 6.079121 |
| P20742 | PZP | 15.95912 | -0.24542 | 0.046718 | -0.0667 | -0.06343 | 0.008794 | 0.002384 | 0.218726 | 0.571177 | 0.583798 | 0.930772 | 0.010744 | 0.349802 | 0.885469 | 0.942137 | 0.986433 | -0.33779 | 0.064303 | -0.09181 | -0.0873 | 0.012104 | TRUE | FALSE | FALSE | FALSE | FALSE | 18.5435 | 3.291275 | 4.732162 | 4.49447 | 0.611417 |
| P18428 | LBP | 18.94902 | 0.046561 | 0.099586 | 0.045414 | 0.040806 | -0.01977 | 0.569806 | 0.010546 | 0.705699 | 0.729934 | 0.848314 | 0.714672 | 0.037161 | 0.926973 | 0.94573 | 0.947857 | 0.06306 | 0.134875 | 0.061507 | 0.055266 | -0.02678 | FALSE | TRUE | FALSE | FALSE | FALSE | 3.279986 | 7.146574 | 3.197943 | 2.868817 | 1.379885 |
| O43866 | CD5L | 8.893295 | -0.16866 | -0.06879 | -0.30707 | -0.06464 | -0.18929 | 0.0304 | 0.061685 | 0.007375 | 0.563982 | 0.054021 | 0.088219 | 0.135653 | 0.155921 | 0.942137 | 0.205001 | -0.23724 | -0.09677 | -0.43193 | -0.09093 | -0.26626 | FALSE | FALSE | FALSE | FALSE | FALSE | 12.40131 | 4.884026 | 23.71912 | 4.582637 | 14.02051 |
| P02745 | C1QA | 12.0242 | 0.096531 | 0.02811 | -0.04281 | -0.02535 | -0.05925 | 0.021401 | 0.155578 | 0.485372 | 0.674169 | 0.26162 | 0.06464 | 0.264661 | 0.85518 | 0.94573 | 0.515623 | 0.256079 | 0.074571 | -0.11357 | -0.06726 | -0.15718 | FALSE | FALSE | FALSE | FALSE | FALSE | 6.919909 | 1.967543 | 3.012016 | 1.772869 | 4.192512 |
| P07225 | PROS1 | 7.188543 | 0.003521 | -0.02948 | -0.00778 | -0.02175 | 0.003367 | 0.911799 | 0.050611 | 0.867654 | 0.635486 | 0.933112 | 0.957066 | 0.120312 | 0.959934 | 0.94573 | 0.986433 | 0.012352 | -0.10342 | -0.02728 | -0.07629 | 0.011812 | FALSE | FALSE | FALSE | FALSE | FALSE | 0.244376 | 2.064489 | 0.540534 | 1.518944 | 0.233672 |
| P29622 | SERPINA | 8.402829 | 0.144667 | -0.02143 | 0.048045 | -0.01431 | 0.094943 | 0.000687 | 0.28377 | 0.438625 | 0.814397 | 0.075585 | 0.004067 | 0.42855 | 0.801438 | 0.96269 | 0.24859 | 0.373959 | -0.0554 | 0.124195 | -0.03699 | 0.245424 | TRUE | FALSE | FALSE | FALSE | FALSE | 10.54754 | 1.496655 | 3.38631 | 0.996694 | 6.802303 |
| O75636 | FCN3 | 8.837166 | 0.053388 | -0.06877 | 0.002441 | -0.00103 | 0.207845 | 0.367891 | 0.014597 | 0.977612 | 0.990427 | 0.00573 | 0.555591 | 0.046964 | 0.984262 | 0.994466 | 0.038548 | 0.098399 | -0.12675 | 0.004499 | -0.00189 | 0.383077 | FALSE | TRUE | FALSE | FALSE | TRUE | 3.769897 | 4.882245 | 0.169358 | 0.071135 | 15.49619 |
| P01701 | IGLV1-51 | 15.48794 | -0.01185 | -0.07074 | -0.08083 | 0.127873 | 0.029406 | 0.882256 | 0.062327 | 0.491555 | 0.268576 | 0.770924 | 0.932671 | 0.135653 | 0.855884 | 0.78094 | 0.920514 | -0.01648 | -0.09835 | -0.11238 | 0.177775 | 0.040881 | FALSE | FALSE | FALSE | FALSE | FALSE | 0.824871 | 5.025623 | 5.762906 | 9.268158 | 2.05915 |
| P01703 | IGLV1-40 | 17.9083 | 0.117592 | 0.000122 | -0.20996 | -0.10267 | 0.004102 | 0.250125 | 0.997991 | 0.162122 | 0.48625 | 0.974613 | 0.411317 | 0.997991 | 0.583261 | 0.942137 | 0.987963 | 0.128489 | 0.000133 | -0.22942 | -0.11219 | 0.004483 | FALSE | FALSE | FALSE | FALSE | FALSE | 8.492246 | 0.008432 | 15.66564 | 7.376076 | 0.284764 |
| P06312 | IGKV4-1 | 11.05918 | 0.053487 | -0.06724 | -0.09705 | -0.16577 | -0.03767 | 0.361943 | 0.015797 | 0.260025 | 0.050771 | 0.610783 | 0.555285 | 0.047713 | 0.63724 | 0.379489 | 0.809842 | 0.10053 | -0.12638 | -0.1824 | -0.31157 | -0.0708 | FALSE | TRUE | FALSE | FALSE | FALSE | 3.777024 | 4.771184 | 6.958288 | 12.17646 | 2.645336 |
| P01705 | IGLV2-23 | 14.81073 | -0.04552 | -0.05518 | 0.080396 | 0.004087 | 0.004524 | 0.631744 | 0.219808 | 0.564269 | 0.976205 | 0.96989 | 0.754017 | 0.349802 | 0.885469 | 0.989577 | 0.987963 | -0.05366 | -0.06505 | 0.094775 | 0.004819 | 0.005333 | FALSE | FALSE | FALSE | FALSE | FALSE | 3.20559 | 3.898979 | 5.730824 | 0.283725 | 0.314058 |
| P49908 | SELENOP | 14.68113 | 0.119625 | -0.09052 | -0.02979 | 0.031104 | -0.03889 | 0.050153 | 0.001816 | 0.738999 | 0.723391 | 0.612853 | 0.123711 | 0.007073 | 0.927669 | 0.94573 | 0.809842 | 0.215892 | -0.16336 | -0.05376 | 0.056134 | -0.07019 | FALSE | TRUE | FALSE | FALSE | FALSE | 8.645228 | 6.475066 | 2.086201 | 2.179351 | 2.732576 |
| O75882-2 | ATRN | 8.503868 | 0.010684 | -0.01784 | 0.032854 | 0.047456 | 0.037553 | 0.769414 | 0.301105 | 0.53929 | 0.367151 | 0.414447 | 0.873815 | 0.441223 | 0.885469 | 0.849036 | 0.713234 | 0.032803 | -0.05478 | 0.100875 | 0.145711 | 0.115305 | FALSE | FALSE | FALSE | FALSE | FALSE | 0.743275 | 1.244246 | 2.303381 | 3.344117 | 2.637175 |
| Q9UGM5 | FETUB | 15.33002 | -0.31801 | -0.01527 | -0.16294 | -0.15874 | -0.12455 | 4.61E-05 | 0.675216 | 0.150341 | 0.153964 | 0.200763 | 0.000401 | 0.774667 | 0.570526 | 0.690507 | 0.450197 | -0.44845 | -0.02153 | -0.22977 | -0.22386 | -0.17563 | TRUE | FALSE | FALSE | FALSE | FALSE | 24.66078 | 1.064083 | 11.95649 | 11.6315 | 9.016506 |
| P05452 | CLEC3B | 6.95538 | -0.0485 | -0.06004 | 0.010055 | -0.0576 | -0.09196 | 0.117506 | 5.07E-05 | 0.824735 | 0.197358 | 0.018962 | 0.22296 | 0.000313 | 0.959934 | 0.700478 | 0.087701 | -0.16928 | -0.20957 | 0.035094 | -0.20105 | -0.32098 | FALSE | TRUE | FALSE | FALSE | FALSE | 3.418874 | 4.249697 | 0.699382 | 4.07352 | 6.582075 |
| A0A0B4J1 | IGHV2-26 | 7.953549 | 0.160141 | 0.005193 | -0.02586 | 0.063537 | 0.081096 | 0.041842 | 0.888682 | 0.822346 | 0.574665 | 0.412898 | 0.104959 | 0.920208 | 0.959934 | 0.942137 | 0.713234 | 0.227235 | 0.007369 | -0.03669 | 0.090156 | 0.115072 | FALSE | FALSE | FALSE | FALSE | FALSE | 11.73967 | 0.360616 | 1.80838 | 4.502442 | 5.782124 |
| P00742 | F10 | 8.426214 | 0.002871 | -0.12131 | 0.089063 | -0.00972 | -0.03139 | 0.932542 | 4.1E-13 | 0.07444 | 0.842565 | 0.463611 | 0.958446 | 8.68E-12 | 0.479008 | 0.96269 | 0.731172 | 0.008731 | -0.36886 | 0.270819 | -0.02957 | -0.09545 | FALSE | TRUE | FALSE | FALSE | FALSE | 0.199233 | 8.771901 | 6.367935 | 0.67622 | 2.199671 |
| P00739 | HPR | 5.951066 | 0.203356 | -0.11731 | -0.01241 | 0.094946 | -0.29985 | 0.002645 | 0.000258 | 0.899857 | 0.327669 | 0.000459 | 0.011512 | 0.001232 | 0.965064 | 0.843224 | 0.003997 | 0.322698 | -0.18616 | -0.01969 | 0.150666 | -0.47583 | TRUE | TRUE | FALSE | FALSE | TRUE | 15.13734 | 8.471369 | 0.863872 | 6.802524 | 23.102 |
| P01599 | IGKV1-17 | 11.47207 | 0.074199 | 0.019654 | -0.12428 | -0.0294 | 0.096061 | 0.331868 | 0.586675 | 0.268358 | 0.789768 | 0.31955 | 0.522516 | 0.715539 | 0.640596 | 0.958079 | 0.598651 | 0.10826 | 0.028676 | -0.18134 | -0.0429 | 0.140158 | FALSE | FALSE | FALSE | FALSE | FALSE | 5.277624 | 1.371623 | 8.996621 | 2.058718 | 6.88512 |
| P00740 | F9 | 15.05167 | 0.216283 | -0.04474 | 0.21766 | 0.294123 | 0.291473 | 0.01527 | 0.286927 | 0.095523 | 0.022181 | 0.009631 | 0.047083 | 0.428941 | 0.488242 | 0.234483 | 0.054824 | 0.266419 | -0.05511 | 0.268116 | 0.362303 | 0.359039 | TRUE | FALSE | FALSE | FALSE | FALSE | 16.17363 | 3.14985 | 16.2846 | 22.6139 | 22.38889 |
| P19652 | ORM2 | 5.01087 | 0.033044 | -0.03555 | -0.01805 | 0.034192 | -0.01442 | 0.523927 | 0.147788 | 0.812503 | 0.647649 | 0.825494 | 0.680186 | 0.257325 | 0.959934 | 0.94573 | 0.947857 | 0.071268 | -0.07666 | -0.03893 | 0.073742 | -0.03111 | FALSE | FALSE | FALSE | FALSE | FALSE | 2.3169 | 2.494426 | 1.259081 | 2.39829 | 1.004808 |
| P05160 | F13B | 15.38294 | 0.033123 | -0.00192 | 0.093668 | -0.1766 | 0.050264 | 0.558114 | 0.942786 | 0.259635 | 0.031044 | 0.481364 | 0.705991 | 0.962292 | 0.63724 | 0.306305 | 0.731172 | 0.064828 | -0.00376 | 0.183327 | -0.34564 | 0.098378 | FALSE | FALSE | FALSE | FALSE | FALSE | 2.322448 | 0.133138 | 6.707947 | 13.02171 | 3.54546 |
| A0A075B | IGLV3-10 | 19.42063 | -0.04005 | 0.024823 | -0.07342 | -0.15014 | -0.16953 | 0.711842 | 0.628376 | 0.644606 | 0.337527 | 0.215817 | 0.829548 | 0.738443 | 0.92623 | 0.846678 | 0.475228 | -0.04133 | 0.025619 | -0.07578 | -0.15495 | -0.17497 | FALSE | FALSE | FALSE | FALSE | FALSE | 2.814668 | 1.73546 | 5.220936 | 10.96739 | 12.46951 |
| A0A0C4D | IGHV1-18 | 18.42498 | 0.293546 | 0.025541 | -0.1656 | 0.035362 | -0.06615 | 0.000644 | 0.526605 | 0.186371 | 0.773781 | 0.538788 | 0.003974 | 0.666133 | 0.583261 | 0.95433 | 0.766737 | 0.378157 | 0.032903 | -0.21334 | 0.045555 | -0.08521 | TRUE | FALSE | FALSE | FALSE | FALSE | 22.5649 | 1.786127 | 12.16361 | 2.481419 | 4.691765 |
| P80748 | IGLV3-21 | 17.22441 | 0.1924 | 0.00708 | -0.06995 | 0.221516 | 0.098271 | 0.052188 | 0.879586 | 0.629714 | 0.12097 | 0.430861 | 0.126619 | 0.920208 | 0.913703 | 0.59834 | 0.73008 | 0.216019 | 0.007949 | -0.07853 | 0.248711 | 0.110336 | FALSE | FALSE | FALSE | FALSE | FALSE | 14.26627 | 0.491969 | 4.96787 | 16.59585 | 7.049011 |
| A0A0B4J1 | IGHV3-72 | 25.3251 | 0.189553 | -0.04067 | 0.164537 | 0.092962 | 0.005001 | 0.037817 | 0.344879 | 0.218466 | 0.478999 | 0.965255 | 0.098192 | 0.495554 | 0.606561 | 0.942137 | 0.987963 | 0.231632 | -0.0497 | 0.201064 | 0.113599 | 0.006111 | FALSE | FALSE | FALSE | FALSE | FALSE | 14.041 | 2.859154 | 12.08066 | 6.655763 | 0.347228 |
| P23142;P2 | FBLN1 | 10.99977 | 0.108796 | 0.122829 | 0.015946 | 0.000576 | -0.05362 | 0.059614 | 9.03E-06 | 0.850431 | 0.994466 | 0.460892 | 0.135529 | 5.81E-05 | 0.959934 | 0.994466 | 0.731172 | 0.204 | 0.230312 | 0.0299 | 0.001081 | -0.10055 | FALSE | TRUE | FALSE | FALSE | FALSE | 7.832821 | 8.88676 | 1.111437 | 0.039968 | 3.786925 |
| P69905 | HBA1 | 10.03185 | 0.225461 | 0.009087 | 0.012507 | 0.080647 | 0.001366 | 0.003046 | 0.799343 | 0.910245 | 0.459782 | 0.988572 | 0.012184 | 0.853221 | 0.969182 | 0.934694 | 0.988572 | 0.330074 | 0.013303 | 0.01831 | 0.118068 | 0.002 | TRUE | FALSE | FALSE | FALSE | FALSE | 16.91505 | 0.631847 | 0.870688 | 5.749243 | 0.09473 |
| A0A075B | IGLV2-18 | 17.29441 | 0.236377 | -0.02958 | -0.04326 | -0.30421 | -0.07978 | 0.041594 | 0.588725 | 0.798863 | 0.069007 | 0.584697 | 0.104959 | 0.715539 | 0.959934 | 0.464229 | 0.804817 | 0.22679 | -0.02838 | -0.04151 | -0.29188 | -0.07654 | FALSE | FALSE | FALSE | FALSE | FALSE | 17.8031 | 2.07152 | 3.044086 | 23.47461 | 5.685545 |
| A0A0C4D | IGHV4-28 | 11.19706 | 0.047284 | 0.016656 | 0.101751 | 0.040635 | 0.188706 | 0.485867 | 0.603713 | 0.307236 | 0.67805 | 0.02807 | 0.656917 | 0.726419 | 0.678671 | 0.94573 | 0.122185 | 0.077544 | 0.027315 | 0.166868 | 0.066641 | 0.309471 | FALSE | FALSE | FALSE | FALSE | FALSE | 3.331768 | 1.161193 | 7.307478 | 2.856679 | 13.97407 |
| B9A064 | IGLL5 | 6.79604 | 0.17192 | -0.00108 | -0.08238 | -0.05408 | 0.0212 | 0.000234 | 0.960715 | 0.225411 | 0.417866 | 0.716397 | 0.001649 | 0.973876 | 0.606561 | 0.909472 | 0.88485 | 0.406668 | -0.00255 | -0.19487 | -0.12792 | 0.050147 | TRUE | FALSE | FALSE | FALSE | FALSE | 12.65567 | 0.074696 | 5.876449 | 3.819706 | 1.480313 |
| P06276 | BCHE | 17.37964 | -0.01846 | -0.00944 | 0.155612 | -0.05186 | 0.152386 | 0.773444 | 0.755486 | 0.098968 | 0.575177 | 0.060338 | 0.873815 | 0.840691 | 0.488242 | 0.942137 | 0.219238 | -0.03197 | -0.01636 | 0.269588 | -0.08984 | 0.264 | FALSE | FALSE | FALSE | FALSE | FALSE | 1.287479 | 0.656747 | 11.38938 | 3.660052 | 11.14063 |
