## supplementary_table4 for "Molecular characterization of liver steatosis through circulating proteins, metabolites and lipids" (from another version of this preprint: version 1, 2025-12-02)

| nalyte_nan | nalyte_cla | hemical_n | v_qc_chri | ef_SexMa | oef_Age_1 | liver_fat_( | liver_fat_( | liver_fat_( | alue_SexM | alue_Age_ | e_liver_fat | e_liver_fat | e_liver_fat | adj_SexM | adj_Age_1 | liver_fat_ | liver_fat_ | liver_fat_ | tSize_Sex | ctSize_Age | ze_liver_fa | ze_liver_fa | ze_liver_fa | ficant_Sex | ificant_Ag | nt_liver_fa | nt_liver_fa | nt_liver_fa | perc_Sex | perc_Age | c_liver_fat | c_liver_fat | c_liver_fat_cat14 |
|---|---|---|---|---|---|---|---|---|---|---|---|---|---|---|---|---|---|---|---|---|---|---|---|---|---|---|---|---|---|---|---|---|---|
| ADMA | biogenic ar | Asymmetri | 3.313832 | -0.08149 | 0.041809 | 0.071201 | 0.07026 | 0.030914 | 0.305152 | 0.267466 | 0.541354 | 0.53994 | 0.758257 | 0.434626 | 0.483123 | 0.814023 | 0.675927 | 0.829344 | -0.1157 | 0.059364 | 0.101096 | 0.09976 | 0.043894 | FALSE | FALSE | FALSE | FALSE | FALSE | 5.810706 | 2.940396 | 5.059056 | 4.990568 | 2.165897 |
| Ala | aminoacids | Alanine | 2.394819 | 0.021139 | 0.010477 | 0.055576 | 0.152717 | 0.18568 | 0.495808 | 0.475443 | 0.222941 | 0.000723 | 3.13E-06 | 0.633331 | 0.665621 | 0.609605 | 0.00738 | 6.09E-05 | 0.073292 | 0.036326 | 0.192694 | 0.529499 | 0.643791 | FALSE | FALSE | FALSE | TRUE | TRUE | 1.476017 | 0.728865 | 3.927418 | 11.16608 | 13.73532 |
| alpha-AAA | biogenic ar | alpha-Ami | 10.74347 | 0.50624 | -0.12266 | 0.195415 | 0.302089 | 0.547128 | 1.43E-05 | 0.02464 | 0.255273 | 0.065947 | 0.000182 | 6.25E-05 | 0.079852 | 0.664344 | 0.13917 | 0.001513 | 0.47471 | -0.11502 | 0.183244 | 0.283274 | 0.513051 | TRUE | FALSE | FALSE | FALSE | TRUE | 42.03433 | 8.87407 | 14.50531 | 23.29283 | 46.11735 |
| Arg | aminoacids | Arginine | 1.912311 | -0.06945 | 0.002267 | -0.00769 | -0.00263 | -0.03962 | 0.012885 | 0.863068 | 0.850425 | 0.947718 | 0.259115 | 0.032679 | 0.932326 | 0.936423 | 0.976053 | 0.365687 | -0.27701 | 0.009043 | -0.03069 | -0.01049 | -0.15803 | TRUE | FALSE | FALSE | FALSE | FALSE | 4.93198 | 0.157291 | 0.53479 | 0.182461 | 2.784477 |
| Asn | aminoacids | Asparagin | 1.569123 | 0.057346 | -0.0062 | -0.01257 | -0.06517 | -0.04636 | 0.015387 | 0.578189 | 0.716299 | 0.055923 | 0.119635 | 0.037925 | 0.744033 | 0.864499 | 0.12388 | 0.217768 | 0.269016 | -0.02907 | -0.05897 | -0.30573 | -0.21747 | TRUE | FALSE | FALSE | FALSE | FALSE | 4.054965 | 0.430494 | 0.875088 | 4.620988 | 3.265455 |
| Asp | aminoacids | Aspartate | 2.732266 | -0.14369 | -0.02543 | -0.02423 | 0.051628 | 0.062345 | 0.010937 | 0.339029 | 0.768954 | 0.524382 | 0.379524 | 0.028999 | 0.544313 | 0.894638 | 0.664977 | 0.491976 | -0.28344 | -0.05015 | -0.04779 | 0.10184 | 0.12298 | TRUE | FALSE | FALSE | FALSE | FALSE | 10.47255 | 1.777966 | 1.693642 | 3.643351 | 4.41613 |
| Cit | aminoacids | Citrulline | 2.019113 | 0.021082 | 0.134157 | -0.01354 | -0.05033 | -0.10786 | 0.63894 | 8.83E-10 | 0.83734 | 0.437803 | 0.057826 | 0.750433 | 1.7E-08 | 0.936423 | 0.584851 | 0.134926 | 0.049244 | 0.313375 | -0.03163 | -0.11755 | -0.25194 | FALSE | TRUE | FALSE | FALSE | FALSE | 1.471987 | 9.745168 | 0.942998 | 3.549869 | 7.762729 |
| Creatinine | biogenic ar | Creatinine | 2.357409 | 0.318777 | 0.038208 | -0.02308 | -0.03812 | -0.0693 | 7.64E-21 | 0.011338 | 0.620483 | 0.405506 | 0.084357 | 1.34E-18 | 0.043133 | 0.82889 | 0.558769 | 0.171657 | 0.974229 | 0.116771 | -0.07053 | -0.11651 | -0.21179 | TRUE | TRUE | FALSE | FALSE | FALSE | 24.72728 | 2.683792 | 1.612604 | 2.67769 | 4.920828 |
| DOPA | biogenic ar | Dihydroxy | 1.463391 | -0.03891 | 0.015497 | -0.04432 | -0.03163 | -0.01879 | 0.196513 | 0.276653 | 0.316111 | 0.466496 | 0.620858 | 0.301665 | 0.483123 | 0.703263 | 0.604717 | 0.734122 | -0.14435 | 0.05749 | -0.16441 | -0.11734 | -0.06969 | FALSE | FALSE | FALSE | FALSE | FALSE | 2.733761 | 1.079944 | 3.119473 | 2.216655 | 1.310656 |
| Gln | aminoacids | Glutamine | 2.426557 | 0.029506 | 0.047791 | -0.04876 | -0.08158 | -0.09156 | 0.18969 | 9.5E-06 | 0.139983 | 0.01228 | 0.001359 | 0.296391 | 8.31E-05 | 0.544767 | 0.041788 | 0.007209 | 0.140092 | 0.226909 | -0.23153 | -0.38736 | -0.43471 | FALSE | TRUE | FALSE | TRUE | TRUE | 2.066246 | 3.36809 | 3.437849 | 5.817962 | 6.55195 |
| Glu | aminoacids | Glutamate | 1.877868 | 0.098461 | -0.05604 | 0.109888 | 0.326031 | 0.556288 | 0.076222 | 0.033088 | 0.177259 | 5.65E-05 | 2.93E-14 | 0.14821 | 0.098142 | 0.551116 | 0.001648 | 5.13E-12 | 0.178118 | -0.10137 | 0.19879 | 0.589798 | 1.006339 | FALSE | FALSE | FALSE | TRUE | TRUE | 7.063069 | 3.960657 | 7.914418 | 25.35598 | 47.04806 |
| Gly | aminoacids | Glycine | 1.871623 | -0.25466 | 0.018795 | -0.10764 | -0.13204 | -0.24395 | 2.01E-11 | 0.278811 | 0.046165 | 0.012985 | 2.42E-07 | 3.52E-10 | 0.483123 | 0.421046 | 0.041788 | 1.17E-05 | -0.69216 | 0.051085 | -0.29256 | -0.35889 | -0.66307 | TRUE | FALSE | FALSE | TRUE | TRUE | 19.30529 | 1.311308 | 7.746176 | 9.58449 | 18.42337 |
| His | aminoacids | Histidine | 1.770452 | 0.112055 | -0.01931 | 0.13064 | -0.06679 | -0.06423 | 0.015996 | 0.378203 | 0.055337 | 0.317887 | 0.2722 | 0.038879 | 0.590943 | 0.421046 | 0.465692 | 0.38108 | 0.266179 | -0.04588 | 0.310326 | -0.15866 | -0.15257 | TRUE | FALSE | FALSE | FALSE | FALSE | 8.076658 | 1.347648 | 9.477898 | 4.738599 | 4.552417 |
| Histamine | biogenic ar | Histamine | 0.496982 | 0.001468 | -0.00665 | 0.004513 | 0.00974 | -0.00105 | 0.645109 | 9.44E-05 | 0.314729 | 0.035471 | 0.804907 | 0.751259 | 0.000661 | 0.703263 | 0.086215 | 0.860943 | 0.086344 | -0.39109 | 0.265429 | 0.572792 | -0.06158 | FALSE | TRUE | FALSE | FALSE | FALSE | 0.10182 | 0.462022 | 0.313334 | 0.677398 | 0.072604 |
| Ile | aminoacids | Isoleucine | 1.845524 | 0.296192 | 0.00846 | 0.075717 | 0.160408 | 0.203386 | 1.31E-17 | 0.584711 | 0.115672 | 0.000759 | 1.33E-06 | 7.66E-16 | 0.744033 | 0.510292 | 0.00738 | 3.91E-05 | 0.866288 | 0.024744 | 0.221452 | 0.469153 | 0.594854 | TRUE | FALSE | FALSE | TRUE | TRUE | 22.78989 | 0.588145 | 5.388434 | 11.76029 | 15.13977 |
| Kynurenin | biogenic ar | Kynurenin | 3.337567 | 0.10544 | 0.09134 | -0.00595 | 0.096389 | 0.095606 | 0.011611 | 4.89E-06 | 0.922346 | 0.108823 | 0.069129 | 0.030072 | 4.75E-05 | 0.949618 | 0.205841 | 0.153134 | 0.270653 | 0.234462 | -0.01527 | 0.247421 | 0.245411 | TRUE | TRUE | FALSE | FALSE | FALSE | 7.582223 | 6.535956 | 0.413234 | 6.909442 | 6.85142 |
| Leu | aminoacids | Leucine | 2.025615 | 0.248168 | -0.01432 | 0.089498 | 0.117836 | 0.161674 | 7.8E-18 | 0.266209 | 0.025584 | 0.002871 | 3.64E-06 | 6.82E-16 | 0.483123 | 0.290158 | 0.013955 | 6.36E-05 | 0.87495 | -0.05048 | 0.315536 | 0.415447 | 0.570003 | TRUE | FALSE | FALSE | TRUE | TRUE | 18.76983 | 0.997448 | 6.399964 | 8.510614 | 11.85844 |
| Lys | aminoacids | Lysine | 3.534779 | 0.07599 | -0.0101 | -0.00581 | 0.03712 | 0.047001 | 0.005258 | 0.430165 | 0.88369 | 0.342208 | 0.169421 | 0.015335 | 0.630876 | 0.936423 | 0.490872 | 0.27709 | 0.309366 | -0.04114 | -0.02366 | 0.151123 | 0.191347 | TRUE | FALSE | FALSE | FALSE | FALSE | 5.408377 | 0.702814 | 0.403714 | 2.606377 | 3.311488 |
| Met | aminoacids | Methionin | 2.424235 | 0.117453 | 0.006634 | 0.021139 | 0.048871 | 0.072919 | 7.39E-06 | 0.586723 | 0.576793 | 0.189814 | 0.025665 | 3.69E-05 | 0.744033 | 0.814023 | 0.319399 | 0.071291 | 0.48967 | 0.027658 | 0.088131 | 0.203745 | 0.304006 | TRUE | FALSE | FALSE | FALSE | FALSE | 8.481787 | 0.460894 | 1.476052 | 3.445475 | 5.184286 |
| Met-SO | biogenic ar | Methionin | 25.75556 | -0.31094 | 0.195358 | -0.01355 | 0.44379 | 0.392611 | 0.069667 | 0.014738 | 0.957923 | 0.078316 | 0.067896 | 0.140138 | 0.051583 | 0.963437 | 0.153991 | 0.152331 | -0.23508 | 0.1477 | -0.01024 | 0.335527 | 0.296833 | FALSE | FALSE | FALSE | FALSE | FALSE | 24.05138 | 14.50086 | 0.943485 | 36.01729 | 31.27672 |
| Orn | aminoacids | Ornithine | 3.31438 | 0.125857 | 0.071081 | 0.077849 | 0.03124 | 0.08575 | 0.000612 | 4.6E-05 | 0.145842 | 0.552126 | 0.062659 | 0.002102 | 0.000335 | 0.544767 | 0.685263 | 0.144281 | 0.368404 | 0.208066 | 0.227877 | 0.091445 | 0.251005 | TRUE | TRUE | FALSE | FALSE | FALSE | 9.115566 | 5.050372 | 5.544337 | 2.189028 | 6.123954 |
| Phe | aminoacids | Phenylalan | 1.914453 | 0.05813 | 0.020723 | 0.062358 | 0.062618 | 0.113663 | 0.003767 | 0.028554 | 0.033732 | 0.030099 | 8.65E-06 | 0.011566 | 0.089232 | 0.327955 | 0.075247 | 0.000108 | 0.309807 | 0.110446 | 0.33234 | 0.333727 | 0.605773 | TRUE | FALSE | FALSE | FALSE | TRUE | 4.111525 | 1.446798 | 4.417084 | 4.435916 | 8.197154 |
| Pro | aminoacids | Proline | 2.639889 | 0.246681 | 0.031155 | -0.02838 | 0.069638 | 0.095735 | 4.87E-09 | 0.109202 | 0.637659 | 0.240127 | 0.065242 | 5.02E-08 | 0.238878 | 0.839025 | 0.368665 | 0.148277 | 0.630349 | 0.079611 | -0.07252 | 0.177948 | 0.244633 | TRUE | FALSE | FALSE | FALSE | FALSE | 18.64746 | 2.182979 | 1.986623 | 4.945358 | 6.860954 |
| Putrescine | biogenic ar | Putrescine | 4.355134 | 0.003216 | 0.026189 | -0.10074 | -0.09327 | -0.08894 | 0.949146 | 0.272638 | 0.174153 | 0.200474 | 0.162847 | 0.971348 | 0.483123 | 0.551116 | 0.324238 | 0.271412 | 0.007113 | 0.057928 | -0.22284 | -0.20631 | -0.19673 | FALSE | FALSE | FALSE | FALSE | FALSE | 0.223156 | 1.831833 | 7.232674 | 6.678561 | 6.358957 |
| Sarcosine | biogenic ar | Sarcosine | 2.423926 | 0.1035 | -0.01572 | 0.107762 | 0.040338 | 0.086625 | 0.061866 | 0.547892 | 0.184832 | 0.613154 | 0.2149 | 0.130441 | 0.726371 | 0.551116 | 0.740013 | 0.324202 | 0.208087 | -0.0316 | 0.216656 | 0.0811 | 0.174161 | FALSE | FALSE | FALSE | FALSE | FALSE | 7.437684 | 1.095378 | 7.755522 | 2.835499 | 6.188349 |
| SDMA | biogenic ar | Symmetric | 4.07297 | 0.070775 | 0.117352 | -0.0557 | -0.05066 | -0.12071 | 0.039993 | 3.58E-12 | 0.269774 | 0.307134 | 0.00564 | 0.088592 | 1.57E-10 | 0.670574 | 0.455495 | 0.023699 | 0.210667 | 0.349309 | -0.1658 | -0.15079 | -0.35929 | FALSE | TRUE | FALSE | FALSE | TRUE | 5.028051 | 8.47423 | 3.936411 | 3.573827 | 8.726696 |
| Ser | aminoacids | Serine | 2.24331 | -0.04997 | -0.02714 | -0.04771 | -0.07303 | -0.0649 | 0.026246 | 0.010811 | 0.147434 | 0.024384 | 0.022202 | 0.061241 | 0.042042 | 0.544767 | 0.064655 | 0.063695 | -0.24256 | -0.13174 | -0.2316 | -0.35456 | -0.31506 | FALSE | TRUE | FALSE | FALSE | FALSE | 3.524023 | 1.898745 | 3.362082 | 5.192728 | 4.601179 |
| Serotonin | biogenic ar | Serotonin | 1.963043 | -0.09423 | -0.18145 | 0.250795 | -0.13782 | -0.07495 | 0.456591 | 0.002624 | 0.176717 | 0.449645 | 0.639583 | 0.614642 | 0.013918 | 0.551116 | 0.59612 | 0.74618 | -0.08218 | -0.15823 | 0.218704 | -0.12018 | -0.06536 | FALSE | TRUE | FALSE | FALSE | FALSE | 6.749918 | 13.40206 | 18.98622 | 10.02401 | 5.332508 |
| Spermidin | biogenic ar | Spermidin | 2.606853 | 0.049768 | -0.00595 | 0.01037 | -0.00956 | 0.0181 | 0.07476 | 0.65173 | 0.799862 | 0.81204 | 0.606773 | 0.147 | 0.798657 | 0.909598 | 0.871822 | 0.722349 | 0.199318 | -0.02383 | 0.041532 | -0.03829 | 0.072491 | FALSE | FALSE | FALSE | FALSE | FALSE | 3.509858 | 0.413211 | 0.721399 | 0.664905 | 1.262534 |
| Spermine | biogenic ar | Spermine | 7.921353 | -0.028 | 0.082765 | -0.07436 | -0.12194 | -0.05124 | 0.672661 | 0.007449 | 0.444995 | 0.201954 | 0.54378 | 0.769383 | 0.032591 | 0.764183 | 0.324238 | 0.660844 | -0.04929 | 0.145723 | -0.13092 | -0.21469 | -0.09021 | FALSE | TRUE | FALSE | FALSE | FALSE | 1.959464 | 5.904555 | 5.28934 | 8.81936 | 3.615325 |
| t4-OH-Pro | biogenic ar | trans-4-Hy | 3.533942 | 0.298795 | 0.035781 | 0.01182 | 0.097232 | 0.092348 | 3.24E-07 | 0.187393 | 0.888265 | 0.240159 | 0.202181 | 2.18E-06 | 0.372656 | 0.936423 | 0.368665 | 0.307713 | 0.557317 | 0.06674 | 0.022047 | 0.181359 | 0.172249 | TRUE | FALSE | FALSE | FALSE | FALSE | 23.01165 | 2.511173 | 0.822659 | 6.971914 | 6.61039 |
| Taurine | biogenic ar | Taurine | 1.517923 | -0.10386 | -0.03651 | 0.00887 | -0.01106 | -0.06128 | 0.001035 | 0.014379 | 0.847384 | 0.807154 | 0.122571 | 0.003483 | 0.051355 | 0.936423 | 0.871822 | 0.218877 | -0.36007 | -0.12659 | 0.030751 | -0.03834 | -0.21247 | TRUE | FALSE | FALSE | FALSE | FALSE | 7.464386 | 2.563235 | 0.616698 | 0.769425 | 4.339444 |
| Thr | aminoacids | Threonine | 1.720682 | 0.03153 | -0.04214 | -0.03828 | -0.00219 | 0.005436 | 0.327416 | 0.005909 | 0.417946 | 0.962449 | 0.893494 | 0.46208 | 0.028727 | 0.764183 | 0.979236 | 0.925215 | 0.108309 | -0.14475 | -0.13148 | -0.00751 | 0.018673 | FALSE | TRUE | FALSE | FALSE | FALSE | 2.209526 | 2.963891 | 2.688612 | 0.151617 | 0.377503 |
| Trp | aminoacids | Tryptopha | 1.774949 | 0.11383 | -0.03178 | 0.006731 | 0.02977 | 0.114545 | 8.34E-06 | 0.007886 | 0.855344 | 0.412284 | 0.000353 | 4.03E-05 | 0.03286 | 0.936423 | 0.559299 | 0.002301 | 0.474238 | -0.1324 | 0.028043 | 0.12403 | 0.477216 | TRUE | TRUE | FALSE | FALSE | TRUE | 8.209699 | 2.227306 | 0.467656 | 2.084969 | 8.263331 |
| Tyr | aminoacids | Tyrosine | 1.904165 | 0.003664 | 0.01431 | 0.059417 | 0.145511 | 0.2174 | 0.903537 | 0.317375 | 0.181297 | 0.000944 | 2.68E-08 | 0.952524 | 0.523968 | 0.551116 | 0.007942 | 2.35E-06 | 0.012889 | 0.050337 | 0.209003 | 0.511844 | 0.764717 | FALSE | FALSE | FALSE | TRUE | TRUE | 0.254304 | 0.996835 | 4.204471 | 10.61226 | 16.26365 |
| Val | aminoacids | Valine | 2.020057 | 0.181214 | -0.01164 | 0.079196 | 0.119332 | 0.166795 | 4.06E-12 | 0.328329 | 0.032625 | 0.001115 | 2.67E-07 | 8.89E-11 | 0.532015 | 0.327955 | 0.008127 | 1.17E-05 | 0.713054 | -0.0458 | 0.311626 | 0.469555 | 0.656319 | TRUE | FALSE | FALSE | TRUE | TRUE | 13.38372 | 0.810108 | 5.642904 | 8.623149 | 12.25619 |
| C0 | acylcarniti | Carnitine | 2.82499 | 0.045327 | 0.033635 | 0.017937 | 0.112584 | 0.130781 | 0.172759 | 0.032814 | 0.712885 | 0.019281 | 0.001947 | 0.279934 | 0.098142 | 0.864499 | 0.054422 | 0.009736 | 0.148787 | 0.110408 | 0.058878 | 0.369562 | 0.429295 | FALSE | FALSE | FALSE | FALSE | TRUE | 3.191688 | 2.358769 | 1.251034 | 8.11628 | 9.488615 |
| C10 | acylcarniti | Decanoylc | 2.069999 | 0.040223 | 0.1453 | -0.04716 | 0.069504 | -0.00336 | 0.483177 | 1.58E-07 | 0.575442 | 0.40119 | 0.962937 | 0.631014 | 1.85E-06 | 0.814023 | 0.557208 | 0.968471 | 0.075177 | 0.271565 | -0.08814 | 0.129903 | -0.00629 | FALSE | TRUE | FALSE | FALSE | FALSE | 2.827278 | 10.59608 | 3.322819 | 4.935607 | 0.233363 |
| C10:1 | acylcarniti | Decenoylc | 2.6472 | 0.021066 | 0.143163 | -0.03533 | -0.01164 | 0.013399 | 0.606204 | 1.1E-12 | 0.555994 | 0.843557 | 0.794992 | 0.721671 | 9.64E-11 | 0.814023 | 0.900137 | 0.858787 | 0.053456 | 0.36328 | -0.08965 | -0.02953 | 0.034 | FALSE | TRUE | FALSE | FALSE | FALSE | 1.470923 | 10.43235 | 2.478987 | 0.809803 | 0.93306 |
| C10:2 | acylcarniti | Decadieny | 4.580989 | -0.03401 | 0.050655 | -0.08254 | -0.09028 | -0.06685 | 0.169445 | 1.95E-05 | 0.023484 | 0.01179 | 0.032732 | 0.278704 | 0.000148 | 0.290158 | 0.041264 | 0.085494 | -0.14783 | 0.220166 | -0.35877 | -0.39239 | -0.29057 | FALSE | TRUE | TRUE | FALSE | FALSE | 2.385623 | 3.573494 | 5.888338 | 6.457567 | 4.742877 |
| C12 | acylcarniti | Dodecanoy | 3.721085 | 0.084503 | 0.120073 | 0.021247 | 0.02463 | 0.023559 | 0.028123 | 1.46E-10 | 0.70588 | 0.656271 | 0.62643 | 0.064758 | 3.2E-09 | 0.863839 | 0.775996 | 0.73574 | 0.229862 | 0.326619 | 0.057795 | 0.066999 | 0.064085 | FALSE | TRUE | FALSE | FALSE | FALSE | 6.032218 | 8.678965 | 1.483626 | 1.721907 | 1.646399 |
| C12-DC | acylcarniti | Dodecanec | 2.287337 | 0.00661 | 0.016791 | -0.01285 | -0.0074 | -0.05209 | 0.659116 | 0.018316 | 0.559249 | 0.732336 | 0.006165 | 0.758851 | 0.061641 | 0.814023 | 0.826831 | 0.025089 | 0.048442 | 0.123057 | -0.09416 | -0.0542 | -0.38175 | FALSE | FALSE | FALSE | FALSE | TRUE | 0.459215 | 1.170669 | 0.894496 | 0.513899 | 3.676576 |
| C12:1 | acylcarniti | Dodecenoy | 2.684317 | 0.04931 | 0.137297 | -0.05872 | -0.01666 | 0.004979 | 0.167618 | 8.75E-15 | 0.262771 | 0.746337 | 0.911962 | 0.278704 | 1.53E-12 | 0.666448 | 0.831904 | 0.938784 | 0.14082 | 0.392093 | -0.1677 | -0.04757 | 0.014219 | FALSE | TRUE | FALSE | FALSE | FALSE | 3.477012 | 9.984251 | 4.154245 | 1.161297 | 0.345713 |
| C14 | acylcarniti | Tetradecar | 8.121303 | 0.020718 | 0.082599 | 0.03251 | 0.057519 | 0.073137 | 0.473628 | 4.1E-09 | 0.443765 | 0.168469 | 0.045615 | 0.627916 | 6.53E-08 | 0.764183 | 0.28904 | 0.110869 | 0.075617 | 0.301471 | 0.118654 | 0.209936 | 0.266937 | FALSE | TRUE | FALSE | FALSE | FALSE | 1.446429 | 5.89237 | 2.278972 | 4.067491 | 5.200153 |
| C14:1 | acylcarniti | Tetradecen | 2.912874 | 0.058932 | 0.148314 | -0.07048 | 0.004168 | -0.02626 | 0.23712 | 9.7E-10 | 0.335311 | 0.953743 | 0.676068 | 0.348707 | 1.7E-08 | 0.703263 | 0.976053 | 0.763302 | 0.124903 | 0.314346 | -0.14939 | 0.008834 | -0.05566 | FALSE | TRUE | FALSE | FALSE | FALSE | 4.169415 | 10.82739 | 5.006939 | 0.289337 | 1.836973 |
| C14:1-OH | acylcarniti | Hydroxyte | 4.559527 | 0.062371 | 0.068508 | 0.030625 | -0.01889 | 0.021169 | 0.014349 | 2.48E-08 | 0.410907 | 0.605679 | 0.508406 | 0.035872 | 3.62E-07 | 0.764183 | 0.740013 | 0.622175 | 0.259859 | 0.285432 | 0.127597 | -0.07872 | 0.088198 | FALSE | TRUE | FALSE | FALSE | FALSE | 4.418011 | 4.86319 | 2.145489 | 1.318179 | 1.478136 |
| C14:2 | acylcarniti | Tetradecad | 3.731478 | 0.077088 | 0.152931 | -0.03678 | -0.07804 | -0.02585 | 0.084258 | 2.88E-12 | 0.573956 | 0.225291 | 0.645718 | 0.160274 | 1.57E-10 | 0.814023 | 0.352017 | 0.748349 | 0.17904 | 0.355187 | -0.08542 | -0.18125 | -0.06004 | FALSE | TRUE | FALSE | FALSE | FALSE | 5.488667 | 11.18256 | 2.582089 | 5.558175 | 1.80789 |
| C14:2-OH | acylcarniti | Hydroxyte | 4.236342 | 0.00865 | 0.019831 | -0.00321 | 0.054048 | 0.025496 | 0.699985 | 0.06248 | 0.922486 | 0.095898 | 0.368377 | 0.78524 | 0.160795 | 0.949618 | 0.184418 | 0.482029 | 0.042792 | 0.098107 | -0.01586 | 0.267379 | 0.126132 | FALSE | FALSE | FALSE | FALSE | FALSE | 0.60137 | 1.384096 | 0.222493 | 3.8174 | 1.782982 |
| C16 | acylcarniti | Hexadecan | 9.386632 | 0.080386 | 0.095903 | 0.002367 | 0.057014 | 0.099164 | 0.008902 | 1.36E-10 | 0.957931 | 0.196752 | 0.010544 | 0.024341 | 3.2E-09 | 0.963437 | 0.324238 | 0.039261 | 0.271291 | 0.323659 | 0.007989 | 0.192416 | 0.334664 | FALSE | TRUE | FALSE | FALSE | FALSE | 5.730065 | 6.873402 | 0.164216 | 4.031068 | 7.115248 |
| C16-OH | acylcarniti | Hydroxyhe | 4.406227 | 0.014309 | 0.050057 | -0.02454 | -0.0114 | -0.0108 | 0.502072 | 1.09E-06 | 0.432994 | 0.710949 | 0.688145 | 0.636686 | 1.19E-05 | 0.764183 | 0.818526 | 0.767041 | 0.072559 | 0.253832 | -0.12445 | -0.05779 | -0.05474 | FALSE | TRUE | FALSE | FALSE | FALSE | 0.996751 | 3.53055 | 1.715672 | 0.793125 | 0.751121 |
| C16:1 | acylcarniti | Hexadecen | 3.740384 | -0.00608 | 0.098669 | -0.05205 | 0.052506 | 0.063364 | 0.838459 | 1.45E-11 | 0.234713 | 0.222644 | 0.092749 | 0.905742 | 5.09E-10 | 0.622345 | 0.351015 | 0.186563 | -0.02118 | 0.343929 | -0.18142 | 0.18302 | 0.220869 | FALSE | TRUE | FALSE | FALSE | FALSE | 0.422011 | 7.078502 | 3.67347 | 3.706484 | 4.489964 |
| C16:1-OH | acylcarniti | Hydroxyhe | 6.083342 | 0.01465 | 0.047026 | 0.009566 | 0.010884 | 0.076441 | 0.569901 | 0.000137 | 0.800446 | 0.769847 | 0.019298 | 0.697431 | 0.000923 | 0.909598 | 0.847316 | 0.059248 | 0.06189 | 0.198667 | 0.040413 | 0.04598 | 0.322932 | FALSE | TRUE | FALSE | FALSE | FALSE | 1.020639 | 3.31333 | 0.665282 | 0.757269 | 5.441382 |
| C18 | acylcarniti | Octadecan | 8.137137 | 0.156907 | 0.075146 | 0.047166 | 0.064023 | 0.066875 | 1.29E-05 | 1.01E-05 | 0.365053 | 0.211263 | 0.135615 | 5.81E-05 | 8.46E-05 | 0.737284 | 0.336101 | 0.239723 | 0.465257 | 0.222822 | 0.139857 | 0.189838 | 0.198296 | TRUE | TRUE | FALSE | FALSE | FALSE | 11.48944 | 5.346781 | 3.323357 | 4.537654 | 4.74454 |
| C18:1 | acylcarniti | Octadecen | 2.673785 | 0.080689 | 0.119959 | -0.05338 | 0.010411 | 0.021321 | 0.033957 | 9.72E-11 | 0.337909 | 0.849095 | 0.655977 | 0.076186 | 2.84E-09 | 0.703263 | 0.900556 | 0.755236 | 0.222087 | 0.330173 | -0.14693 | 0.028656 | 0.058683 | FALSE | TRUE | FALSE | FALSE | FALSE | 5.752301 | 8.670414 | 3.769627 | 0.72428 | 1.488813 |
| C2 | acylcarniti | Acetylcarn | 1.166284 | -0.02409 | 0.120261 | -0.05394 | -0.02711 | 0.045804 | 0.594583 | 4.11E-08 | 0.417174 | 0.67816 | 0.42287 | 0.714204 | 5.54E-07 | 0.764183 | 0.794096 | 0.528588 | -0.05699 | 0.284466 | -0.12759 | -0.06412 | 0.108344 | FALSE | TRUE | FALSE | FALSE | FALSE | 1.684077 | 8.693137 | 3.809629 | 1.896753 | 3.225797 |
| C3 | acylcarniti | Propionylc | 2.13717 | 0.069944 | 0.068342 | 0.019187 | 0.194377 | 0.273362 | 0.118467 | 0.001349 | 0.770092 | 0.002778 | 1.95E-06 | 0.21373 | 0.00787 | 0.894638 | 0.013889 | 4.87E-05 | 0.165162 | 0.161378 | 0.045307 | 0.45899 | 0.645499 | FALSE | TRUE | FALSE | TRUE | TRUE | 4.967607 | 4.8511 | 1.338829 | 14.42301 | 20.86209 |
| C3-DC (C4 | acylcarniti | Hydroxybu | 3.993166 | 0.170823 | 0.065402 | -0.04661 | 0.055297 | 0.049175 | 0.000269 | 0.003085 | 0.49401 | 0.409212 | 0.401369 | 0.001003 | 0.015879 | 0.811248 | 0.559299 | 0.511218 | 0.397812 | 0.152306 | -0.10855 | 0.128774 | 0.114519 | TRUE | TRUE | FALSE | FALSE | FALSE | 12.57008 | 4.637617 | 3.28382 | 3.907272 | 3.467322 |
| C3:1 | acylcarniti | Propenoylc | 7.157257 | 0.034619 | 0.02486 | -0.05562 | 0.047939 | 0.09995 | 0.25983 | 0.087558 | 0.217626 | 0.279544 | 0.010257 | 0.375788 | 0.204302 | 0.604516 | 0.421726 | 0.03902 | 0.123646 | 0.08879 | -0.19866 | 0.171221 | 0.356987 | FALSE | FALSE | FALSE | FALSE | TRUE | 2.428621 | 1.738081 | 3.930575 | 3.378695 | 7.17366 |
| C4 | acylcarniti | Butyrylcar | 3.127225 | 0.063267 | 0.082664 | 0.005094 | 0.345618 | 0.215012 | 0.218763 | 0.000749 | 0.946161 | 4.54E-06 | 0.00101 | 0.32721 | 0.00452 | 0.962664 | 0.000398 | 0.005702 | 0.129795 | 0.169588 | 0.010451 | 0.709048 | 0.441105 | FALSE | TRUE | FALSE | TRUE | TRUE | 4.482931 | 5.897152 | 0.353724 | 27.0695 | 16.07135 |
| C4:1 | acylcarniti | Butenylcar | 5.954158 | 0.039632 | 0.022836 | 0.005806 | 0.072788 | 0.082376 | 0.148534 | 0.078666 | 0.885183 | 0.066232 | 0.017654 | 0.259935 | 0.188244 | 0.936423 | 0.13917 | 0.055407 | 0.159168 | 0.091711 | 0.023319 | 0.292325 | 0.330831 | FALSE | FALSE | FALSE | FALSE | FALSE | 2.785182 | 1.595447 | 0.403271 | 5.174736 | 5.876031 |
| C5 | acylcarniti | Valerylcar | 2.662156 | 0.193581 | 0.022542 | 0.023695 | 0.231075 | 0.228521 | 2.54E-07 | 0.195658 | 0.660839 | 1.78E-05 | 1.34E-06 | 1.78E-06 | 0.380445 | 0.853813 | 0.001036 | 3.91E-05 | 0.534376 | 0.062227 | 0.06541 | 0.637878 | 0.630827 | TRUE | FALSE | FALSE | TRUE | TRUE | 14.35985 | 1.574767 | 1.655983 | 17.37091 | 17.16328 |
| C5-DC (C | acylcarniti | Glutarylca | 5.781492 | 0.051754 | 0.035962 | -0.07916 | -0.02937 | -0.03765 | 0.061083 | 0.006094 | 0.051075 | 0.460305 | 0.279437 | 0.130359 | 0.028824 | 0.421046 | 0.604717 | 0.38505 | 0.206316 | 0.14336 | -0.31558 | -0.1171 | -0.15009 | FALSE | TRUE | FALSE | FALSE | FALSE | 3.65242 | 2.523993 | 5.640414 | 2.056962 | 2.644094 |
| C5-M-DC | acylcarniti | Methylglut | 5.564686 | 0.001023 | 0.033549 | -0.03607 | -0.0246 | -0.06529 | 0.968006 | 0.005685 | 0.33549 | 0.503871 | 0.043108 | 0.980219 | 0.028423 | 0.703263 | 0.643631 | 0.106253 | 0.004419 | 0.144969 | -0.15588 | -0.1063 | -0.28212 | FALSE | TRUE | FALSE | FALSE | FALSE | 0.070904 | 2.352709 | 2.53204 | 1.719756 | 4.629394 |
| C5-OH (C | acylcarniti | Hydroxyva | 4.357196 | 0.022645 | 0.022586 | -0.02475 | -0.01439 | -0.01446 | 0.30548 | 0.031187 | 0.445409 | 0.651578 | 0.603987 | 0.434626 | 0.095749 | 0.764183 | 0.775689 | 0.722349 | 0.114067 | 0.113767 | -0.12466 | -0.07249 | -0.07282 | FALSE | FALSE | FALSE | FALSE | FALSE | 1.582033 | 1.577839 | 1.730188 | 1.002487 | 1.007088 |
| C5:1 | acylcarniti | Tiglylcarni | 4.826276 | 0.020382 | 0.022594 | -0.03672 | 0.007038 | 0.0352 | 0.511485 | 0.124453 | 0.420563 | 0.875122 | 0.369097 | 0.639356 | 0.262402 | 0.764183 | 0.919614 | 0.482029 | 0.073223 | 0.081173 | -0.13191 | 0.025286 | 0.126461 | FALSE | FALSE | FALSE | FALSE | FALSE | 1.422772 | 1.578459 | 2.577632 | 0.489047 | 2.469901 |
| C5:1-DC | acylcarniti | Glutaconyl | 6.259158 | 0.018817 | 0.058504 | -0.05702 | -0.00118 | 0.028555 | 0.489495 | 7.69E-06 | 0.15422 | 0.975944 | 0.406054 | 0.632099 | 7.09E-05 | 0.544767 | 0.981553 | 0.511218 | 0.074962 | 0.23306 | -0.22716 | -0.00472 | 0.113755 | FALSE | TRUE | FALSE | FALSE | FALSE | 1.312863 | 4.138518 | 4.031658 | 0.082127 | 1.999012 |
| C6 (C4:1- | acylcarniti | Hexanoylc | 3.187619 | -0.05248 | 0.117744 | -0.01087 | 0.116785 | 0.132833 | 0.246867 | 7.65E-08 | 0.870076 | 0.074546 | 0.020598 | 0.360014 | 9.56E-07 | 0.936423 | 0.14995 | 0.061095 | -0.12306 | 0.276114 | -0.02549 | 0.273864 | 0.311498 | FALSE | TRUE | FALSE | FALSE | FALSE | 3.7044 | 8.503704 | 0.756352 | 8.431575 | 9.644462 |
| C6:1 | acylcarniti | Hexenoylc | 5.605195 | -0.01909 | 0.024296 | -0.02601 | 0.011906 | 0.000514 | 0.472926 | 0.054014 | 0.505292 | 0.7563 | 0.98778 | 0.627916 | 0.143219 | 0.811248 | 0.837674 | 0.98778 | -0.08004 | 0.101884 | -0.10906 | 0.049925 | 0.002155 | FALSE | FALSE | FALSE | FALSE | FALSE | 1.331762 | 1.698366 | 1.819056 | 0.828656 | 0.035623 |
| C8 | acylcarniti | Octanoylc | 4.416431 | 0.010816 | 0.102627 | -0.00685 | 0.116085 | 0.042768 | 0.84882 | 0.000156 | 0.934485 | 0.156906 | 0.550448 | 0.91131 | 0.001011 | 0.956345 | 0.271867 | 0.664334 | 0.020841 | 0.197744 | -0.01319 | 0.223674 | 0.082406 | FALSE | TRUE | FALSE | FALSE | FALSE | 0.752535 | 7.372707 | 0.475764 | 8.378997 | 3.008824 |
| C9 | acylcarniti | Nonaylcar | 3.710575 | -0.00472 | 0.079285 | 0.033171 | 0.000122 | -0.00762 | 0.89259 | 2.44E-06 | 0.518082 | 0.99807 | 0.862762 | 0.951489 | 2.51E-05 | 0.814023 | 0.99807 | 0.89871 | -0.0146 | 0.245295 | 0.102626 | 0.000377 | -0.02359 | FALSE | TRUE | FALSE | FALSE | FALSE | 0.327631 | 5.649409 | 2.325881 | 0.008455 | 0.529873 |
| H1 | sugars | Hexose | 1.649726 | 0.095652 | 0.004626 | 0.028168 | 0.147863 | 0.195771 | 0.004297 | 0.7689 | 0.564416 | 0.002243 | 4.56E-06 | 0.012965 | 0.857054 | 0.814023 | 0.012662 | 7.25E-05 | 0.30521 | 0.014761 | 0.089881 | 0.471808 | 0.624675 | TRUE | FALSE | FALSE | TRUE | TRUE | 6.854787 | 0.321175 | 1.971675 | 10.7927 | 14.53359 |
| lysoPC a C | glycerophc | lysoPhospl | 3.013347 | -0.00434 | 0.002623 | 0.048344 | 0.002953 | 0.021203 | 0.768303 | 0.706647 | 0.025995 | 0.889526 | 0.254674 | 0.845617 | 0.828298 | 0.290158 | 0.92659 | 0.36234 | -0.03278 | 0.019809 | 0.365046 | 0.022301 | 0.1601 | FALSE | FALSE | FALSE | FALSE | FALSE | 0.30134 | 0.182001 | 3.407758 | 0.204928 | 1.480504 |
| lysoPC a C | glycerophc | lysoPhospl | 2.280884 | 0.126291 | 0.004909 | 0.024056 | -0.07918 | -0.07298 | 3.01E-05 | 0.728165 | 0.583202 | 0.066802 | 0.053458 | 0.000128 | 0.828298 | 0.816482 | 0.13917 | 0.128153 | 0.458168 | 0.017808 | 0.087272 | -0.28725 | -0.26475 | TRUE | FALSE | FALSE | FALSE | FALSE | 9.148425 | 0.340824 | 1.681408 | 5.641577 | 5.188413 |
| lysoPC a C | glycerophc | lysoPhospl | 3.497662 | -0.04211 | 0.021491 | 0.026213 | -0.05861 | 0.056476 | 0.331355 | 0.294642 | 0.680233 | 0.348768 | 0.301966 | 0.463897 | 0.500605 | 0.862614 | 0.496214 | 0.409644 | -0.10839 | 0.05532 | 0.067475 | -0.15087 | 0.145374 | FALSE | FALSE | FALSE | FALSE | FALSE | 2.961711 | 1.500792 | 1.833573 | 4.14616 | 3.992239 |
| lysoPC a C | glycerophc | lysoPhospl | 2.690056 | -0.0028 | 0.061724 | -0.04542 | -0.25742 | -0.24849 | 0.947494 | 0.002294 | 0.466629 | 3.43E-05 | 5.12E-06 | 0.971348 | 0.012545 | 0.777715 | 0.001501 | 7.47E-05 | -0.00695 | 0.153348 | -0.11285 | -0.63955 | -0.61736 | FALSE | TRUE | FALSE | TRUE | TRUE | 0.194112 | 4.371201 | 3.198615 | 19.53403 | 18.79652 |
| lysoPC a C | glycerophc | lysoPhospl | 3.238563 | 0.061267 | 0.007075 | 0.008854 | -0.13333 | -0.11591 | 0.11097 | 0.696652 | 0.875082 | 0.016502 | 0.017151 | 0.20229 | 0.823744 | 0.936423 | 0.049791 | 0.055407 | 0.175933 | 0.020317 | 0.025426 | -0.38288 | -0.33285 | FALSE | FALSE | FALSE | TRUE | FALSE | 4.338185 | 0.491618 | 0.615633 | 9.682495 | 8.365988 |
| lysoPC a C | glycerophc | lysoPhospl | 1.577274 | 0.19621 | 0.028299 | -0.04128 | -0.14205 | -0.23037 | 3.28E-06 | 0.149687 | 0.497839 | 0.018089 | 1.44E-05 | 1.69E-05 | 0.307136 | 0.811248 | 0.052761 | 0.00016 | 0.499276 | 0.072011 | -0.10504 | -0.36146 | -0.5862 | TRUE | FALSE | FALSE | FALSE | TRUE | 14.56848 | 1.980931 | 2.90254 | 10.3471 | 17.31351 |
| lysoPC a C | glycerophc | lysoPhospl | 2.237454 | 0.335863 | -0.00068 | 0.030764 | -0.19208 | -0.25309 | 1.71E-11 | 0.976358 | 0.663535 | 0.006005 | 3.97E-05 | 3.32E-10 | 0.993388 | 0.853813 | 0.02444 | 0.000409 | 0.712331 | -0.00143 | 0.065248 | -0.40738 | -0.53677 | TRUE | FALSE | FALSE | TRUE | TRUE | 26.21324 | 0.046805 | 2.155304 | 14.2408 | 19.17534 |
| lysoPC a C | glycerophc | lysoPhospl | 2.43977 | 0.186857 | -0.01012 | 0.030179 | -0.1164 | -0.06985 | 8.52E-06 | 0.604711 | 0.618972 | 0.051662 | 0.18111 | 4.03E-05 | 0.761327 | 0.82889 | 0.115908 | 0.290773 | 0.488689 | -0.02647 | 0.078929 | -0.30442 | -0.18268 | TRUE | FALSE | FALSE | FALSE | FALSE | 13.82812 | 0.704014 | 2.113917 | 8.402443 | 4.96075 |
| lysoPC a C | glycerophc | lysoPhospl | 2.262356 | 0.26805 | 0.01235 | -0.14286 | -0.02052 | -0.11507 | 8.35E-10 | 0.538089 | 0.022255 | 0.737354 | 0.032111 | 9.13E-09 | 0.724351 | 0.290158 | 0.82716 | 0.085144 | 0.663115 | 0.030552 | -0.35341 | -0.05076 | -0.28466 | TRUE | FALSE | FALSE | FALSE | FALSE | 20.41794 | 0.859715 | 10.40896 | 1.432347 | 8.302565 |
| lysoPC a C | glycerophc | lysoPhospl | 7.775769 | 0.046439 | 0.035091 | -0.09507 | -0.17655 | -0.20998 | 0.185257 | 0.034705 | 0.065055 | 0.000536 | 2.98E-06 | 0.294728 | 0.099562 | 0.426061 | 0.006253 | 6.09E-05 | 0.141933 | 0.107249 | -0.29056 | -0.53958 | -0.64177 | FALSE | FALSE | FALSE | TRUE | TRUE | 3.271245 | 2.462115 | 6.811623 | 13.01742 | 15.66707 |
| lysoPC a C | glycerophc | lysoPhospl | 9.180088 | 0.050832 | 0.034837 | -0.0192 | -0.14705 | -0.06257 | 0.21648 | 0.073709 | 0.75026 | 0.013559 | 0.228012 | 0.326587 | 0.18152 | 0.893167 | 0.041788 | 0.338153 | 0.136653 | 0.093655 | -0.05161 | -0.39531 | -0.1682 | FALSE | FALSE | FALSE | TRUE | FALSE | 3.586184 | 2.444132 | 1.339567 | 10.72993 | 4.432294 |
| lysoPC a C | glycerophc | lysoPhospl | 9.369488 | -0.02934 | 0.053284 | -0.06338 | -0.09951 | -0.11025 | 0.477504 | 0.006672 | 0.29614 | 0.095582 | 0.034972 | 0.628294 | 0.029937 | 0.70033 | 0.184418 | 0.090001 | -0.07811 | 0.141871 | -0.16875 | -0.26496 | -0.29356 | FALSE | TRUE | FALSE | FALSE | FALSE | 2.054371 | 3.762386 | 4.491032 | 7.141176 | 7.941891 |
| lysoPC a C | glycerophc | lysoPhospl | 6.810845 | 0.05385 | -0.01472 | 0.000386 | -0.07521 | -0.07389 | 0.140859 | 0.394173 | 0.994254 | 0.154038 | 0.109472 | 0.248993 | 0.610445 | 0.994254 | 0.269567 | 0.208234 | 0.164028 | -0.04484 | 0.001176 | -0.2291 | -0.22507 | FALSE | FALSE | FALSE | FALSE | FALSE | 3.803126 | 1.025665 | 0.026758 | 5.351523 | 5.255071 |
| lysoPC a C | glycerophc | lysoPhospl | 5.177682 | -0.24589 | 0.018222 | -0.0146 | -0.19948 | -0.08283 | 6.01E-09 | 0.349667 | 0.809063 | 0.000871 | 0.111556 | 5.84E-08 | 0.556288 | 0.913458 | 0.007942 | 0.208671 | -0.62293 | 0.046164 | -0.037 | -0.50536 | -0.20984 | TRUE | FALSE | FALSE | TRUE | FALSE | 18.58216 | 1.271068 | 1.017356 | 14.82841 | 5.90918 |
| PC aa C24 | glycerophc | Phosphatic | 128.7089 | -0.00525 | 0.050909 | 0.046744 | 0.058416 | -0.10548 | 0.930593 | 0.074682 | 0.597163 | 0.501696 | 0.165899 | 0.971348 | 0.18152 | 0.822862 | 0.643631 | 0.27389 | -0.00966 | 0.093705 | 0.086039 | 0.107523 | -0.19415 | FALSE | FALSE | FALSE | FALSE | FALSE | 0.364309 | 3.59176 | 3.293123 | 4.132211 | 7.585249 |
| PC aa C26 | glycerophc | Phosphatic | 3.789764 | 0.024707 | 0.011104 | 0.014464 | -0.03774 | -0.03008 | 0.226048 | 0.250004 | 0.628969 | 0.200057 | 0.242862 | 0.33524 | 0.470438 | 0.83386 | 0.324238 | 0.351246 | 0.13479 | 0.060577 | 0.078906 | -0.20591 | -0.16409 | FALSE | FALSE | FALSE | FALSE | FALSE | 1.727323 | 0.77263 | 1.007579 | 2.650761 | 2.106764 |
| PC aa C28 | glycerophc | Phosphatic | 3.101898 | -0.20614 | 0.014839 | 0.01579 | -0.11685 | 0.00317 | 5.69E-08 | 0.398125 | 0.771945 | 0.029698 | 0.946022 | 4.33E-07 | 0.611156 | 0.894638 | 0.075247 | 0.962523 | -0.59015 | 0.042483 | 0.045205 | -0.33452 | 0.009077 | TRUE | FALSE | FALSE | FALSE | FALSE | 15.35943 | 1.033867 | 1.100486 | 8.43619 | 0.220004 |
| PC aa C30 | glycerophc | Phosphatic | 3.157753 | -0.19989 | 0.014705 | 0.090563 | -0.03595 | 0.065855 | 5.42E-05 | 0.524994 | 0.20762 | 0.610462 | 0.28627 | 0.000216 | 0.712202 | 0.604516 | 0.740013 | 0.391384 | -0.4454 | 0.032767 | 0.201796 | -0.0801 | 0.146741 | TRUE | FALSE | FALSE | FALSE | FALSE | 14.86107 | 1.024503 | 6.478564 | 2.52306 | 4.670518 |
| PC aa C30 | glycerophc | Phosphatic | 29.12546 | -0.12023 | -0.00182 | -0.70393 | -0.17488 | 0.22752 | 0.798584 | 0.994282 | 0.321253 | 0.805449 | 0.699013 | 0.873451 | 0.999213 | 0.703263 | 0.871822 | 0.774224 | -0.06568 | -0.00099 | -0.38454 | -0.09553 | 0.124288 | FALSE | FALSE | FALSE | FALSE | FALSE | 8.690523 | 0.12618 | 62.89358 | 12.88715 | 17.08208 |

| | | | | | | | | | | | | | | | | | | | | | | | | | | | | | | | | |
|---|---|---|---|---|---|---|---|---|---|---|---|---|---|---|---|---|---|---|---|---|---|---|---|---|---|---|---|---|---|---|---|---|
| PC aa C32 glycerophc | Phosphatic | 3.269742 | -0.02742 | 0.023404 | -0.03982 | -0.07962 | -0.01061 | 0.425516 | 0.150936 | 0.430545 | 0.10939 | 0.806826 | 0.58176 | 0.307136 | 0.764183 | 0.205841 | 0.860943 | -0.08884 | 0.075818 | -0.12901 | -0.25793 | -0.03439 | FALSE | FALSE | FALSE | FALSE | FALSE | 1.918977 | 1.635495 | 2.798805 | 5.674078 | 0.738476 |
| PC aa C32 glycerophc | Phosphatic | 3.835023 | -0.32663 | 0.011211 | 0.092899 | 0.091922 | 0.310842 | 5.43E-07 | 0.71081 | 0.322539 | 0.319332 | 0.000139 | 3.39E-06 | 0.828298 | 0.703263 | 0.465692 | 0.001265 | -0.54312 | 0.018641 | 0.154473 | 0.152848 | 0.516869 | TRUE | FALSE | FALSE | FALSE | TRUE | 25.40782 | 0.780094 | 6.651104 | 6.578914 | 24.04316 |
| PC aa C32 glycerophc | Phosphatic | 4.253093 | -0.32464 | -0.03054 | 0.180584 | -0.1909 | 0.060589 | 1.2E-06 | 0.32572 | 0.061646 | 0.044535 | 0.464722 | 7.02E-06 | 0.532015 | 0.426061 | 0.10532 | 0.572721 | -0.52594 | -0.04947 | 0.292563 | -0.30927 | 0.09816 | TRUE | FALSE | FALSE | FALSE | FALSE | 25.23493 | 2.139177 | 13.33425 | 14.14749 | 4.289133 |
| PC aa C32 glycerophc | Phosphatic | 4.208173 | -0.2997 | -0.02047 | 0.007901 | -0.12412 | -0.08582 | 1.17E-12 | 0.285716 | 0.89432 | 0.034371 | 0.093964 | 2.93E-11 | 0.490199 | 0.937161 | 0.084716 | 0.186859 | -0.75625 | -0.05167 | 0.019936 | -0.31319 | -0.21655 | TRUE | FALSE | FALSE | FALSE | FALSE | 23.08867 | 1.429334 | 0.549137 | 8.983971 | 6.128807 |
| PC aa C34 glycerophc | Phosphatic | 3.383832 | -0.06834 | -0.01665 | 0.017833 | -0.00183 | 0.048224 | 0.080894 | 0.367714 | 0.755826 | 0.974072 | 0.328292 | 0.155566 | 0.579729 | 0.893713 | 0.981553 | 0.438557 | -0.19502 | -0.04752 | 0.050889 | -0.00523 | 0.137612 | FALSE | FALSE | FALSE | FALSE | FALSE | 4.851003 | 1.160956 | 1.243777 | 0.127061 | 3.399134 |
| PC aa C34 glycerophc | Phosphatic | 2.991069 | -0.0517 | -0.03341 | 0.07333 | -0.12514 | -0.02068 | 0.166473 | 0.059019 | 0.181238 | 0.020633 | 0.660621 | 0.278704 | 0.154155 | 0.551116 | 0.056418 | 0.755612 | -0.15198 | -0.09822 | 0.215571 | -0.36787 | -0.06078 | FALSE | FALSE | FALSE | FALSE | FALSE | 3.648311 | 2.342906 | 5.214206 | 9.061088 | 1.443435 |
| PC aa C34 glycerophc | Phosphatic | 2.912396 | -0.308 | -0.01113 | 0.043424 | -0.16496 | -0.04486 | 3.66E-10 | 0.621311 | 0.534845 | 0.01691 | 0.45577 | 4.58E-09 | 0.771131 | 0.814023 | 0.050157 | 0.565672 | -0.67369 | -0.02435 | 0.09498 | -0.36082 | -0.09813 | TRUE | FALSE | FALSE | FALSE | FALSE | 23.79904 | 0.774788 | 3.055648 | 12.11357 | 3.158656 |
| PC aa C34 glycerophc | Phosphatic | 3.385872 | -0.29134 | -0.04563 | 0.039319 | -0.02639 | 0.084195 | 4.08E-08 | 0.063533 | 0.605591 | 0.724397 | 0.198822 | 3.25E-07 | 0.161135 | 0.827957 | 0.823178 | 0.307713 | -0.59742 | -0.09357 | 0.080626 | -0.05411 | 0.172648 | TRUE | FALSE | FALSE | FALSE | FALSE | 22.37792 | 3.213496 | 2.762831 | 1.845783 | 6.009614 |
| PC aa C36 glycerophc | Phosphatic | 5.925132 | -0.09028 | -0.03091 | -0.08414 | -0.17128 | -0.0989 | 0.094933 | 0.22631 | 0.288448 | 0.028372 | 0.146966 | 0.174876 | 0.430481 | 0.691484 | 0.073016 | 0.254645 | -0.18487 | -0.06329 | -0.17231 | -0.35076 | -0.20253 | FALSE | FALSE | FALSE | FALSE | FALSE | 6.457426 | 2.165423 | 6.005646 | 12.60582 | 7.095482 |
| PC aa C36 glycerophc | Phosphatic | 3.595559 | -0.12656 | -0.04545 | 0.06322 | 0.022693 | 0.062564 | 0.002841 | 0.023066 | 0.306897 | 0.708852 | 0.239611 | 0.008877 | 0.076161 | 0.703263 | 0.818526 | 0.349433 | -0.32947 | -0.11831 | 0.164583 | 0.059079 | 0.162877 | TRUE | FALSE | FALSE | FALSE | FALSE | 9.168502 | 3.200176 | 4.479495 | 1.585422 | 4.432039 |
| PC aa C36 glycerophc | Phosphatic | 2.861902 | -0.09614 | -0.03951 | 0.053011 | -0.10981 | -0.01127 | 0.011685 | 0.028303 | 0.341585 | 0.04556 | 0.814 | 0.030072 | 0.089232 | 0.703263 | 0.106306 | 0.863333 | -0.27674 | -0.11371 | 0.152589 | -0.31609 | -0.03243 | TRUE | FALSE | FALSE | FALSE | FALSE | 6.891156 | 2.77612 | 3.742756 | 7.908774 | 0.783929 |
| PC aa C36 glycerophc | Phosphatic | 3.075998 | -0.0957 | -0.04465 | 0.025092 | -0.1157 | -0.01774 | 0.005771 | 0.006467 | 0.620018 | 0.020519 | 0.683402 | 0.016558 | 0.029781 | 0.82889 | 0.056418 | 0.766637 | -0.30115 | -0.1405 | 0.078956 | -0.36408 | -0.05582 | TRUE | TRUE | FALSE | FALSE | FALSE | 6.858609 | 3.143228 | 1.754426 | 8.350097 | 1.237253 |
| PC aa C36 glycerophc | Phosphatic | 2.693045 | -0.05151 | -0.03541 | -0.07897 | 0.043438 | 0.060461 | 0.170408 | 0.046677 | 0.152323 | 0.422616 | 0.202212 | 0.278704 | 0.127634 | 0.544767 | 0.568907 | 0.307713 | -0.15122 | -0.10396 | -0.23183 | 0.127522 | 0.177499 | FALSE | FALSE | FALSE | FALSE | FALSE | 3.634887 | 2.484916 | 5.626301 | 3.056677 | 4.279927 |
| PC aa C36 glycerophc | Phosphatic | 3.125412 | -0.25883 | 0.001044 | -0.07145 | -0.03743 | 0.004727 | 5.02E-05 | 0.972061 | 0.440185 | 0.680654 | 0.952609 | 0.000204 | 0.993388 | 0.764183 | 0.794096 | 0.963622 | -0.44909 | 0.001812 | -0.12397 | -0.06495 | 0.008201 | TRUE | FALSE | FALSE | FALSE | FALSE | 19.65097 | 0.072395 | 5.077177 | 2.628734 | 0.328177 |
| PC aa C36 glycerophc | Phosphatic | 4.257296 | -0.36437 | -0.0518 | -0.05488 | -0.20021 | -0.03022 | 2.32E-10 | 0.049958 | 0.502252 | 0.013165 | 0.667298 | 3.12E-09 | 0.134503 | 0.811248 | 0.041788 | 0.758293 | -0.67881 | -0.09649 | -0.10224 | -0.37299 | -0.05629 | TRUE | FALSE | FALSE | TRUE | FALSE | 28.73151 | 3.65541 | 3.877071 | 14.8865 | 2.116573 |
| PC aa C38 glycerophc | Phosphatic | 3.248528 | -0.1003 | -0.03405 | -0.13187 | -0.17091 | -0.13958 | 0.03139 | 0.121804 | 0.053775 | 0.011163 | 0.01773 | 0.07134 | 0.259947 | 0.421046 | 0.040697 | 0.055407 | -0.23575 | -0.08003 | -0.30995 | -0.4017 | -0.32807 | FALSE | FALSE | FALSE | TRUE | FALSE | 7.199862 | 2.388292 | 9.571553 | 12.577 | 10.15869 |
| PC aa C38 glycerophc | Phosphatic | 22.40724 | -0.20526 | -0.03623 | -0.42006 | -0.16844 | -0.20506 | 0.065317 | 0.488663 | 0.010279 | 0.296736 | 0.140977 | 0.135547 | 0.672554 | 0.290158 | 0.443835 | 0.246709 | -0.20759 | -0.03664 | -0.42484 | -0.17035 | -0.20738 | FALSE | FALSE | FALSE | FALSE | FALSE | 15.28965 | 2.54281 | 33.79871 | 12.38421 | 15.27307 |
| PC aa C38 glycerophc | Phosphatic | 2.923735 | -0.19272 | -0.03456 | 0.024942 | -0.02062 | 0.121323 | 2.06E-06 | 0.067646 | 0.670141 | 0.72013 | 0.016407 | 1.12E-05 | 0.169114 | 0.85602 | 0.823178 | 0.054175 | -0.51685 | -0.09269 | 0.066891 | -0.0553 | 0.325367 | TRUE | FALSE | FALSE | FALSE | FALSE | 14.29189 | 2.424496 | 1.743912 | 1.439616 | 8.773203 |
| PC aa C38 glycerophc | Phosphatic | 2.576442 | -0.1123 | -0.02045 | -0.1191 | 0.058205 | 0.094822 | 0.007217 | 0.298599 | 0.051661 | 0.332209 | 0.071383 | 0.02037 | 0.502451 | 0.421046 | 0.480467 | 0.155402 | -0.29474 | -0.05368 | -0.31257 | 0.152759 | 0.248861 | TRUE | FALSE | FALSE | FALSE | FALSE | 8.095253 | 1.42783 | 8.60558 | 4.116964 | 6.793381 |
| PC aa C38 glycerophc | Phosphatic | 2.546442 | -0.1273 | -0.00159 | -0.08793 | -0.05693 | -0.05732 | 0.000375 | 0.924312 | 0.091732 | 0.266047 | 0.200549 | 0.001368 | 0.974425 | 0.442626 | 0.404854 | 0.307713 | -0.39285 | -0.00491 | -0.27135 | -0.17569 | -0.17689 | TRUE | FALSE | FALSE | FALSE | FALSE | 9.224885 | 0.110446 | 6.284373 | 4.025166 | 4.053068 |
| PC aa C38 glycerophc | Phosphatic | 2.958901 | -0.15975 | -0.05414 | -0.11226 | -0.09472 | -0.01048 | 0.000468 | 0.011798 | 0.091606 | 0.147336 | 0.85435 | 0.001637 | 0.043927 | 0.442626 | 0.260442 | 0.89871 | -0.38209 | -0.12949 | -0.26851 | -0.22656 | -0.02506 | TRUE | TRUE | FALSE | FALSE | FALSE | 11.70901 | 3.823718 | 8.091903 | 6.785975 | 0.728911 |
| PC aa C40 glycerophc | Phosphatic | 5.841275 | -0.056 | 0.006355 | -0.06364 | -0.08891 | -0.09287 | 0.087709 | 0.681377 | 0.185805 | 0.060407 | 0.025075 | 0.165044 | 0.81672 | 0.551116 | 0.132141 | 0.070777 | -0.18909 | 0.02146 | -0.2149 | -0.30022 | -0.3136 | FALSE | FALSE | FALSE | FALSE | FALSE | 3.957961 | 0.441498 | 4.510225 | 6.356605 | 6.649126 |
| PC aa C40 glycerophc | Phosphatic | 7.90969 | 0.029393 | 0.015671 | -0.08912 | -0.16199 | -0.17688 | 0.491231 | 0.437873 | 0.155648 | 0.008903 | 0.001128 | 0.632099 | 0.6311 | 0.544767 | 0.034621 | 0.006171 | 0.075651 | 0.040333 | -0.22937 | -0.41692 | -0.45525 | FALSE | FALSE | FALSE | TRUE | TRUE | 2.058285 | 1.092159 | 6.372015 | 11.88291 | 13.04395 |
| PC aa C40 glycerophc | Phosphatic | 5.808048 | -0.00091 | -0.00091 | -0.0654 | -0.06992 | -0.06551 | 0.979778 | 0.957267 | 0.215719 | 0.178213 | 0.149294 | 0.985409 | 0.989921 | 0.604516 | 0.302789 | 0.256141 | -0.00283 | -0.00283 | -0.20325 | -0.21731 | -0.20361 | FALSE | FALSE | FALSE | FALSE | FALSE | 0.063165 | 0.063162 | 4.637525 | 4.966043 | 4.645781 |
| PC aa C40 glycerophc | Phosphatic | 3.577413 | -0.01066 | -0.01168 | -0.03537 | 0.04005 | 0.11956 | 0.814248 | 0.586239 | 0.595358 | 0.540742 | 0.037384 | 0.885053 | 0.744033 | 0.822862 | 0.675927 | 0.093459 | -0.02612 | -0.02863 | -0.08669 | 0.098154 | 0.293016 | FALSE | FALSE | FALSE | FALSE | FALSE | 0.74144 | 0.812917 | 2.482205 | 2.814934 | 8.640316 |
| PC aa C40 glycerophc | Phosphatic | 3.315088 | -0.09791 | 0.020819 | -0.04613 | 0.009498 | 0.088473 | 0.022465 | 0.30333 | 0.462304 | 0.877574 | 0.10156 | 0.053127 | 0.50555 | 0.777715 | 0.919614 | 0.197477 | -0.2537 | 0.053943 | -0.11951 | 0.024609 | 0.229233 | FALSE | FALSE | FALSE | FALSE | FALSE | 7.022544 | 1.453559 | 3.248929 | 0.66052 | 6.324403 |
| PC aa C40 glycerophc | Phosphatic | 2.991252 | -0.227 | -0.01356 | -0.09706 | -0.01843 | 0.107956 | 2.54E-06 | 0.545444 | 0.163867 | 0.787699 | 0.071929 | 1.34E-05 | 0.726371 | 0.551116 | 0.861545 | 0.155402 | -0.51457 | -0.03075 | -0.22 | -0.04177 | 0.244713 | TRUE | FALSE | FALSE | FALSE | FALSE | 17.04011 | 0.944586 | 6.958832 | 1.285458 | 7.77004 |
| PC aa C42 glycerophc | Phosphatic | 4.495367 | -0.05341 | 0.012568 | -0.08794 | -0.21995 | -0.18008 | 0.161624 | 0.485758 | 0.116638 | 7.76E-05 | 0.000211 | 0.274604 | 0.672554 | 0.510292 | 0.00194 | 0.001609 | -0.15131 | 0.035604 | -0.24912 | -0.6231 | -0.51015 | FALSE | FALSE | FALSE | TRUE | TRUE | 3.771422 | 0.874929 | 6.284891 | 16.469 | 13.29435 |
| PC aa C42 glycerophc | Phosphatic | 5.942829 | -0.02896 | -0.00403 | -0.08397 | -0.17631 | -0.20536 | 0.434019 | 0.817783 | 0.122787 | 0.001057 | 1.46E-05 | 0.588786 | 0.897861 | 0.516372 | 0.008044 | 0.00016 | -0.08465 | -0.01179 | -0.24549 | -0.51542 | -0.60034 | FALSE | FALSE | FALSE | TRUE | TRUE | 2.027437 | 0.279871 | 5.993348 | 12.99904 | 15.29713 |
| PC aa C42 glycerophc | Phosphatic | 6.703979 | 0.021573 | -0.00721 | -0.04802 | -0.18558 | -0.12705 | 0.579618 | 0.695434 | 0.401167 | 0.001056 | 0.010108 | 0.704397 | 0.823744 | 0.764183 | 0.008044 | 0.03902 | 0.060895 | -0.02035 | -0.13554 | -0.52383 | -0.35861 | FALSE | FALSE | FALSE | TRUE | TRUE | 1.506595 | 0.501084 | 3.384269 | 13.72743 | 9.205503 |
| PC aa C42 glycerophc | Phosphatic | 7.193068 | 0.004459 | 0.0131 | -0.07743 | -0.15238 | -0.05651 | 0.897119 | 0.422202 | 0.12688 | 0.002368 | 0.194761 | 0.951489 | 0.630876 | 0.516372 | 0.012839 | 0.307055 | 0.014295 | 0.041992 | -0.24821 | -0.48846 | -0.18115 | FALSE | FALSE | FALSE | TRUE | FALSE | 0.30958 | 0.912134 | 5.513808 | 11.14006 | 3.994684 |
| PC aa C42 glycerophc | Phosphatic | 6.33096 | -0.05892 | 0.000946 | -0.14354 | -0.09146 | 0.017547 | 0.156929 | 0.961637 | 0.019155 | 0.127925 | 0.737983 | 0.271906 | 0.989921 | 0.290158 | 0.230792 | 0.812245 | -0.15676 | 0.002516 | -0.38191 | -0.24336 | 0.046688 | FALSE | FALSE | FALSE | FALSE | FALSE | 4.168458 | 0.065562 | 10.46119 | 6.545145 | 1.223719 |
| PC aa C42 glycerophc | Phosphatic | 6.090835 | -0.06972 | 0.010247 | -0.13309 | -0.14984 | -0.05588 | 0.072692 | 0.576237 | 0.01983 | 0.007704 | 0.253645 | 0.144558 | 0.744033 | 0.290158 | 0.030639 | 0.36234 | -0.19798 | 0.029095 | -0.37792 | -0.42548 | -0.15868 | FALSE | FALSE | FALSE | TRUE | FALSE | 4.951494 | 0.712772 | 9.664142 | 10.94491 | 3.949556 |
| PC ae C30 glycerophc | Phosphatic | 4.611273 | -0.13361 | 0.036363 | 0.024118 | -0.17001 | -0.07808 | 0.00506 | 0.105358 | 0.72879 | 0.013325 | 0.192158 | 0.015009 | 0.23638 | 0.873549 | 0.041788 | 0.305705 | -0.30742 | 0.083664 | 0.055491 | -0.39116 | -0.17965 | TRUE | FALSE | FALSE | TRUE | FALSE | 9.703875 | 2.552512 | 1.685772 | 12.5066 | 5.561231 |
| PC ae C30 glycerophc | Phosphatic | 19.79333 | 0.181477 | -0.00078 | 0.447096 | -0.01107 | 0.25051 | 0.159641 | 0.989603 | 0.019121 | 0.953338 | 0.120706 | 0.273894 | 0.999213 | 0.290158 | 0.976053 | 0.217768 | 0.162394 | -0.0007 | 0.400083 | -0.00991 | 0.224168 | FALSE | FALSE | FALSE | FALSE | FALSE | 13.40444 | 0.053909 | 36.3293 | 0.770248 | 18.96272 |
| PC ae C30 glycerophc | Phosphatic | 5.476484 | -0.29505 | 0.022331 | -0.0339 | -0.09321 | -0.04148 | 9.62E-15 | 0.194382 | 0.525241 | 0.076145 | 0.365977 | 4.21E-13 | 0.380445 | 0.814023 | 0.151425 | 0.482029 | -0.82347 | 0.062326 | -0.0946 | -0.26015 | -0.11577 | TRUE | FALSE | FALSE | FALSE | FALSE | 22.69272 | 1.559941 | 2.377369 | 6.674339 | 2.91683 |
| PC ae C32 glycerophc | Phosphatic | 3.541761 | -0.07315 | 0.014186 | -0.03754 | -0.18187 | -0.0913 | 0.094282 | 0.491925 | 0.557833 | 0.004099 | 0.097963 | 0.174876 | 0.672554 | 0.814023 | 0.018392 | 0.192625 | -0.18463 | 0.035805 | -0.09476 | -0.45902 | -0.23043 | FALSE | FALSE | FALSE | TRUE | FALSE | 5.20124 | 0.988167 | 2.636515 | 13.43517 | 6.532964 |
| PC ae C32 glycerophc | Phosphatic | 3.859534 | -0.20778 | 0.014723 | -0.04237 | -0.16135 | -0.08118 | 2.38E-07 | 0.429546 | 0.463857 | 0.004778 | 0.103116 | 1.74E-06 | 0.630876 | 0.777715 | 0.020905 | 0.1983 | -0.55948 | 0.039642 | -0.11408 | -0.43446 | -0.21858 | TRUE | FALSE | FALSE | TRUE | FALSE | 15.49126 | 1.025719 | 2.980337 | 11.83356 | 5.788096 |
| PC ae C34 glycerophc | Phosphatic | 4.51781 | -0.15287 | 0.032933 | 0.007214 | -0.12196 | 0.009322 | 0.000391 | 0.103563 | 0.908349 | 0.048374 | 0.862615 | 0.001395 | 0.23638 | 0.946197 | 0.111388 | 0.89871 | -0.39027 | 0.084078 | 0.018417 | -0.31137 | 0.023799 | TRUE | FALSE | FALSE | FALSE | FALSE | 11.17794 | 2.30903 | 0.501296 | 8.821518 | 0.648242 |
| PC ae C34 glycerophc | Phosphatic | 3.569067 | -0.16221 | 0.01888 | -0.06403 | -0.15804 | -0.07297 | 1.09E-05 | 0.272233 | 0.23032 | 0.002751 | 0.112086 | 5.01E-05 | 0.483123 | 0.620091 | 0.013889 | 0.208671 | -0.47843 | 0.055686 | -0.18886 | -0.46612 | -0.21523 | TRUE | FALSE | FALSE | TRUE | FALSE | 11.90016 | 1.317269 | 4.538444 | 11.57689 | 5.18829 |
| PC ae C34 glycerophc | Phosphatic | 3.187138 | -0.03828 | -0.03241 | -0.01133 | -0.23615 | -0.17373 | 0.412968 | 0.143221 | 0.868827 | 0.000523 | 0.003433 | 0.569051 | 0.298377 | 0.936423 | 0.006253 | 0.01611 | -0.08907 | -0.07543 | -0.02637 | -0.54951 | -0.40428 | FALSE | FALSE | FALSE | TRUE | TRUE | 2.688803 | 2.272179 | 0.788486 | 17.7842 | 12.79741 |
| PC ae C34 glycerophc | Phosphatic | 3.068844 | -0.02087 | -0.03415 | -0.06305 | -0.32212 | -0.31414 | 0.69981 | 0.182849 | 0.427839 | 4.69E-05 | 6.04E-06 | 0.78524 | 0.3678 | 0.764183 | 0.001643 | 8.13E-05 | -0.04123 | -0.06747 | -0.12454 | -0.6363 | -0.62053 | FALSE | FALSE | FALSE | TRUE | TRUE | 1.457448 | 2.39567 | 4.466835 | 25.01677 | 24.32707 |
| PC ae C36 glycerophc | Phosphatic | 6.433383 | -0.00247 | -0.0058 | -0.04791 | -0.04897 | -0.01343 | 0.947592 | 0.743862 | 0.385106 | 0.366438 | 0.776926 | 0.971348 | 0.83446 | 0.762727 | 0.517151 | 0.844485 | -0.00736 | -0.01732 | -0.14294 | -0.14611 | -0.04006 | FALSE | FALSE | FALSE | FALSE | FALSE | 0.171231 | 0.403114 | 3.376514 | 3.452712 | 0.935083 |
| PC ae C36 glycerophc | Phosphatic | 3.989232 | -0.17505 | 0.013219 | -0.0093 | -0.1596 | -0.05388 | 1.36E-06 | 0.432469 | 0.858687 | 0.002035 | 0.230713 | 7.7E-06 | 0.630876 | 0.936423 | 0.0128 | 0.339284 | -0.52345 | 0.039527 | -0.02781 | -0.47725 | -0.16113 | TRUE | FALSE | FALSE | TRUE | FALSE | 12.90053 | 0.92046 | 0.646814 | 11.69789 | 3.805569 |
| PC ae C36 glycerophc | Phosphatic | 3.236341 | -0.15539 | 0.010933 | -0.01502 | -0.2747 | -0.18801 | 0.000145 | 0.567726 | 0.800284 | 3.68E-06 | 0.000267 | 0.000552 | 0.744033 | 0.909598 | 0.000398 | 0.001884 | -0.40182 | 0.028273 | -0.03884 | -0.71037 | -0.48618 | TRUE | FALSE | FALSE | TRUE | TRUE | 11.37203 | 0.760723 | 1.046607 | 20.97453 | 13.91908 |
| PC ae C36 glycerophc | Phosphatic | 3.065706 | -0.02659 | -0.05428 | -0.0247 | -0.20388 | -0.16013 | 0.543526 | 0.009095 | 0.70065 | 0.001361 | 0.003964 | 0.669838 | 0.037016 | 0.863839 | 0.009524 | 0.017344 | -0.06589 | -0.13451 | -0.06121 | -0.50524 | -0.39684 | FALSE | TRUE | FALSE | TRUE | TRUE | 1.860155 | 3.83386 | 1.726808 | 15.17887 | 11.73913 |
| PC ae C36 glycerophc | Phosphatic | 3.032223 | 0.112373 | -0.06208 | -0.09601 | -0.04482 | -0.05464 | 0.008435 | 0.002136 | 0.123931 | 0.464383 | 0.307907 | 0.023431 | 0.01206 | 0.516372 | 0.604717 | 0.41449 | 0.288479 | -0.15937 | -0.24648 | -0.11505 | -0.14028 | TRUE | TRUE | FALSE | FALSE | FALSE | 8.100523 | 4.397158 | 6.881532 | 3.155247 | 3.860259 |
| PC ae C36 glycerophc | Phosphatic | 3.007321 | 0.085537 | -0.03451 | -0.11077 | -0.10071 | -0.06193 | 0.057606 | 0.105095 | 0.093783 | 0.121018 | 0.275119 | 0.124458 | 0.23638 | 0.442626 | 0.222928 | 0.38211 | 0.210214 | -0.08481 | -0.27222 | -0.24749 | -0.1522 | FALSE | FALSE | FALSE | FALSE | FALSE | 6.108262 | 2.420806 | 7.980166 | 7.229858 | 4.386342 |
| PC ae C38 glycerophc | Phosphatic | 3.550554 | -0.26284 | -0.05075 | -0.09268 | -0.20223 | -0.11723 | 6.4E-09 | 0.015424 | 0.152816 | 0.00161 | 0.03575 | 5.9E-08 | 0.052925 | 0.544767 | 0.010065 | 0.09067 | -0.61694 | -0.11911 | -0.21754 | -0.47467 | -0.27517 | TRUE | FALSE | FALSE | TRUE | FALSE | 19.98402 | 3.580039 | 6.635163 | 15.04761 | 8.465374 |
| PC ae C38 glycerophc | Phosphatic | 47.93896 | 0.007009 | -0.04234 | -0.47692 | -0.14933 | -0.52844 | 0.969017 | 0.614425 | 0.072685 | 0.562387 | 0.021236 | 0.980219 | 0.768032 | 0.438284 | 0.693082 | 0.061938 | 0.004711 | -0.02846 | -0.32052 | -0.10036 | -0.35515 | FALSE | FALSE | FALSE | FALSE | FALSE | 0.487019 | 2.978339 | 39.17677 | 10.90517 | 44.23722 |
| PC ae C38 glycerophc | Phosphatic | 7.87996 | -0.12289 | -0.00865 | 0.028554 | -0.23606 | -0.15577 | 0.016323 | 0.719699 | 0.702633 | 0.001443 | 0.015866 | 0.039131 | 0.828298 | 0.863839 | 0.009712 | 0.053396 | -0.26167 | -0.01841 | 0.060801 | -0.50266 | -0.33168 | TRUE | FALSE | FALSE | TRUE | FALSE | 8.891251 | 0.601206 | 1.998909 | 17.77717 | 11.4013 |
| PC ae C38 glycerophc | Phosphatic | 3.545273 | -0.17746 | -1.6E-05 | -0.03219 | -0.18769 | -0.0853 | 7.78E-07 | 0.999213 | 0.533888 | 0.000259 | 0.055793 | 4.7E-06 | 0.999213 | 0.814023 | 0.004083 | 0.131943 | -0.53158 | -4.9E-05 | -0.09643 | -0.56224 | -0.25551 | TRUE | FALSE | FALSE | TRUE | FALSE | 13.08922 | 0.00114 | 2.256358 | 13.89425 | 6.090678 |
| PC ae C38 glycerophc | Phosphatic | 2.998136 | -0.02108 | -0.00146 | -0.11703 | -0.08267 | -0.06964 | 0.506976 | 0.922528 | 0.012484 | 0.071946 | 0.083008 | 0.638279 | 0.974425 | 0.290158 | 0.146401 | 0.170899 | -0.07353 | -0.0051 | -0.40824 | -0.28837 | -0.2429 | FALSE | FALSE | FALSE | FALSE | FALSE | 1.471772 | 0.101304 | 8.450346 | 5.897526 | 4.945138 |
| PC ae C38 glycerophc | Phosphatic | 2.661204 | 0.093806 | -0.02733 | -0.11755 | -0.07573 | -0.1188 | 0.009451 | 0.108731 | 0.026529 | 0.145091 | 0.009207 | 0.025444 | 0.238878 | 0.290158 | 0.259092 | 0.036618 | 0.28508 | -0.08307 | -0.35724 | -0.23014 | -0.36104 | TRUE | FALSE | FALSE | FALSE | TRUE | 6.718195 | 1.912642 | 8.489142 | 5.389216 | 8.583235 |
| PC ae C38 glycerophc | Phosphatic | 2.858186 | -0.05485 | -0.04976 | -0.10335 | -0.13692 | -0.08312 | 0.189245 | 0.012109 | 0.092308 | 0.023522 | 0.115192 | 0.296391 | 0.044148 | 0.442626 | 0.066329 | 0.212196 | -0.14395 | -0.1306 | -0.27122 | -0.35933 | -0.21812 | FALSE | TRUE | FALSE | FALSE | FALSE | 3.875289 | 3.509502 | 7.426393 | 9.955712 | 5.930308 |
| PC ae C40 glycerophc | Phosphatic | 4.11431 | -0.06993 | -0.04871 | -0.05532 | -0.10848 | -0.11814 | 0.069669 | 0.007739 | 0.327472 | 0.05125 | 0.015367 | 0.140138 | 0.03286 | 0.703263 | 0.115908 | 0.052729 | -0.19838 | -0.13817 | -0.15692 | -0.30775 | -0.33516 | FALSE | TRUE | FALSE | FALSE | FALSE | 4.966376 | 3.433618 | 3.908628 | 7.809245 | 8.533777 |
| PC ae C40 glycerophc | Phosphatic | 3.644111 | -0.11604 | 0.02153 | -0.09513 | -0.13523 | -0.11232 | 0.001525 | 0.210704 | 0.075134 | 0.010267 | 0.014729 | 0.004941 | 0.4052 | 0.438284 | 0.038611 | 0.051553 | -0.34654 | 0.064294 | -0.28409 | -0.40382 | -0.33542 | TRUE | FALSE | FALSE | FALSE | FALSE | 8.375881 | 1.503527 | 6.816344 | 9.826452 | 8.096545 |
| PC ae C40 glycerophc | Phosphatic | 5.15853 | -0.13241 | 0.01634 | -0.08873 | -0.16882 | -0.11824 | 4.12E-05 | 0.278831 | 0.058613 | 0.00028 | 0.003498 | 0.000172 | 0.483123 | 0.426061 | 0.004083 | 0.01611 | -0.44136 | 0.054466 | -0.29575 | -0.56273 | -0.39412 | TRUE | FALSE | FALSE | TRUE | FALSE | 9.612368 | 1.13904 | 6.343167 | 12.41399 | 8.540758 |
| PC ae C40 glycerophc | Phosphatic | 3.143324 | -0.01948 | 0.00515 | -0.1098 | -0.11267 | -0.08758 | 0.535253 | 0.728902 | 0.017749 | 0.013366 | 0.02773 | 0.664321 | 0.828298 | 0.290158 | 0.041788 | 0.075825 | -0.06847 | 0.0181 | -0.38594 | -0.396 | -0.30783 | FALSE | FALSE | FALSE | FALSE | FALSE | 1.359377 | 0.357585 | 7.90814 | 8.122476 | 6.25865 |
| PC ae C40 glycerophc | Phosphatic | 3.020174 | -0.01301 | 0.012972 | -0.12037 | -0.12252 | -0.15002 | 0.692999 | 0.405522 | 0.013279 | 0.01037 | 0.000355 | 0.78524 | 0.617099 | 0.290158 | 0.038611 | 0.002301 | -0.04316 | 0.043033 | -0.3993 | -0.40642 | -0.49765 | FALSE | FALSE | FALSE | TRUE | FALSE | 0.90593 | 0.903232 | 8.701452 | 8.863338 | 10.9583 |
| PC ae C40 glycerophc | Phosphatic | 3.07727 | -0.12303 | 0.004224 | -0.14172 | -0.19341 | -0.16515 | 0.001964 | 0.820901 | 0.014855 | 0.000758 | 0.001004 | 0.00625 | 0.897861 | 0.290158 | 0.00738 | 0.005702 | -0.33498 | 0.011502 | -0.38589 | -0.52662 | -0.44967 | TRUE | FALSE | FALSE | TRUE | FALSE | 8.901752 | 0.293228 | 10.32215 | 14.36423 | 12.12811 |
| PC ae C42 glycerophc | Phosphatic | 4.630525 | -0.00823 | 0.004757 | -0.0569 | -0.09637 | -0.10628 | 0.719038 | 0.660267 | 0.090947 | 0.00373 | 0.000269 | 0.801476 | 0.802408 | 0.442626 | 0.017175 | 0.001884 | -0.03933 | 0.022731 | -0.27189 | -0.46051 | -0.50788 | FALSE | FALSE | FALSE | TRUE | TRUE | 0.572109 | 0.330262 | 4.022709 | 6.90804 | 7.645167 |
| PC ae C42 glycerophc | Phosphatic | 5.312253 | 0.002688 | 0.004108 | -0.08479 | -0.17371 | -0.14651 | 0.937048 | 0.798584 | 0.090387 | 0.000459 | 0.000722 | 0.971348 | 0.884508 | 0.442626 | 0.006177 | 0.004356 | 0.008624 | 0.01318 | -0.27206 | -0.55741 | -0.47012 | FALSE | FALSE | FALSE | TRUE | TRUE | 0.186462 | 0.285117 | 6.052984 | 12.79539 | 10.68861 |
| PC ae C42 glycerophc | Phosphatic | 5.061613 | -0.05344 | -0.02061 | -0.0781 | -0.17391 | -0.1543 | 0.17616 | 0.269904 | 0.178128 | 0.002421 | 0.002096 | 0.282825 | 0.483123 | 0.551116 | 0.012839 | 0.010187 | -0.14789 | -0.05703 | -0.21614 | -0.48131 | -0.42703 | FALSE | FALSE | FALSE | TRUE | TRUE | 3.77355 | 1.438625 | 5.56253 | 12.81146 | 11.28811 |
| PC ae C42 glycerophc | Phosphatic | 4.184516 | -0.04619 | -0.01323 | -0.05723 | -0.1741 | -0.17561 | 0.202606 | 0.439967 | 0.281998 | 0.000953 | 0.000145 | 0.308313 | 0.6311 | 0.685413 | 0.007942 | 0.001265 | -0.13829 | -0.03961 | -0.17136 | -0.52126 | -0.52578 | FALSE | FALSE | FALSE | TRUE | TRUE | 3.253245 | 0.921293 | 4.046843 | 12.82571 | 12.94391 |
| PC ae C42 glycerophc | Phosphatic | 4.541949 | -0.01343 | -0.00293 | -0.10567 | -0.15769 | -0.14416 | 0.730765 | 0.873819 | 0.065735 | 0.005365 | 0.003628 | 0.809391 | 0.938149 | 0.426061 | 0.022253 | 0.01628 | -0.03787 | -0.00827 | -0.29806 | -0.4448 | -0.40662 | FALSE | FALSE | FALSE | TRUE | TRUE | 0.934898 | 0.203333 | 7.599584 | 11.55022 | 10.50864 |
| PC ae C42 glycerophc | Phosphatic | 3.250125 | 0.018738 | 0.012287 | -0.13475 | -0.1521 | -0.14244 | 0.595851 | 0.462272 | 0.009736 | 0.003041 | 0.005213 | 0.714204 | 0.657344 | 0.290158 | 0.014382 | 0.007837 | 0.058083 | 0.038087 | -0.41771 | -0.47148 | -0.44153 | FALSE | FALSE | FALSE | TRUE | TRUE | 1.307277 | 0.855312 | 9.790537 | 11.11851 | 10.37702 |
| PC ae C44 glycerophc | Phosphatic | 9.249492 | 0.019757 | -0.00862 | -0.07427 | -0.2191 | -0.11196 | 0.625535 | 0.652617 | 0.21192 | 0.000207 | 0.02899 | 0.739653 | 0.798657 | 0.604516 | 0.004017 | 0.078649 | 0.053516 | -0.02335 | -0.20118 | -0.59348 | -0.30327 | FALSE | FALSE | FALSE | TRUE | FALSE | 1.378852 | 0.599239 | 5.282724 | 16.40051 | 8.06962 |
| PC ae C44 glycerophc | Phosphatic | 7.581114 | 0.016805 | 0.001069 | -0.0219 | -0.06939 | -0.08341 | 0.648229 | 0.951086 | 0.685516 | 0.192273 | 0.073443 | 0.751259 | 0.989921 | 0.86306 | 0.320454 | 0.156738 | 0.050922 | 0.003238 | -0.06636 | -0.21026 | -0.25275 | FALSE | FALSE | FALSE | FALSE | FALSE | 1.171617 | 0.074098 | 1.529403 | 4.927144 | 5.951836 |
| PC ae C44 glycerophc | Phosphatic | 3.984887 | 0.058037 | 0.015416 | -0.11066 | -0.12067 | -0.15681 | 0.194406 | 0.465775 | 0.092226 | 0.061906 | 0.005688 | 0.301071 | 0.657344 | 0.442626 | 0.133748 | 0.023699 | 0.14349 | 0.038115 | -0.27359 | -0.29833 | -0.3877 | FALSE | FALSE | FALSE | FALSE | TRUE | 4.104862 | 1.074306 | 7.972077 | 8.723665 | 11.48223 |
| PC ae C44 glycerophc | Phosphatic | 3.974078 | 0.00054 | 0.039267 | -0.16335 | -0.22268 | -0.21617 | 0.9896 | 0.045868 | 0.007598 | 0.00023 | 4.52E-05 | 0.9896 | 0.12741 | 0.290158 | 0.004024 | 0.00044 | 0.001403 | 0.101995 | -0.42428 | -0.5784 | -0.5615 | FALSE | FALSE | FALSE | TRUE | TRUE | 0.037458 | 2.759178 | 11.98814 | 16.69006 | 16.16496 |
| SM (OH) ( sphingolipi | Hydroxysp | 3.347952 | -0.25097 | 0.039152 | -0.03218 | -0.17352 | -0.06883 | 3.94E-10 | 0.033958 | 0.573227 | 0.002149 | 0.161483 | 4.6E-09 | 0.099046 | 0.814023 | 0.012538 | 0.271412 | -0.66706 | 0.104065 | -0.08552 | -0.46122 | -0.18296 | TRUE | FALSE | FALSE | TRUE | FALSE | 19.00045 | 2.751001 | 2.255277 | 12.78097 | 4.886922 |
| SM (OH) ( sphingolipi | Hydroxysp | 3.409106 | -0.25542 | 0.045642 | -0.04494 | -0.15031 | -0.06311 | 2.45E-11 | 0.009361 | 0.407535 | 0.005068 | 0.1764 | 3.9E-10 | 0.03723 | 0.764183 | 0.021633 | 0.285834 | -0.70887 | 0.12667 | -0.12472 | -0.41715 | -0.17515 | TRUE | TRUE | FALSE | TRUE | FALSE | 19.36827 | 3.214212 | 3.163847 | 10.98067 | 4.471613 |
| SM (OH) ( sphingolipi | Hydroxysp | 3.687209 | -0.22203 | -0.06412 | 0.032778 | -0.14002 | -0.0404 | 1.54E-08 | 0.000452 | 0.559783 | 0.011652 | 0.403225 | 1.35E-07 | 0.002824 | 0.814023 | 0.041264 | 0.511218 | -0.60016 | -0.17332 | 0.088599 | -0.37848 | -0.1092 | TRUE | TRUE | FALSE | FALSE | FALSE | 16.6376 | 4.544843 | 2.297994 | 10.19217 | 2.83979 |
| SM (OH) ( sphingolipi | Hydroxysp | 3.711382 | -0.28724 | -0.01395 | -0.06244 | -0.20619 | -0.12045 | 2.35E-13 | 0.4326 | 0.258145 | 0.000169 | 0.011482 | 6.85E-12 | 0.630876 | 0.664344 | 0.003702 | 0.041008 | -0.76761 | -0.03729 | -0.16685 | -0.55101 | -0.3219 | TRUE | FALSE | FALSE | TRUE | FALSE | 22.03014 | 0.971858 | 4.422771 | 15.36349 | 8.707755 |
| SM (OH) ( sphingolipi | Hydroxysp | 6.140091 | -0.17695 | -0.04278 | -0.02916 | -0.12795 | -0.07913 | 7E-05 | 0.040285 | 0.651422 | 0.044343 | 0.154352 | 0.000272 | 0.113706 | 0.850737 | 0.10532 | 0.262248 | -0.4341 | -0.10494 | -0.07153 | -0.31388 | -0.19413 | TRUE | FALSE | FALSE | FALSE | FALSE | 13.04926 | 3.009519 | 2.041631 | 9.273638 | 5.638366 |
| SM C16:0 | sphingolip Sphingomy | 2.918797 | -0.10212 | 0.00496 | -0.05118 | -0.14618 | -0.15095 | 0.001399 | 0.740929 | 0.272061 | 0.001528 | 0.000191 | 0.00462 | 0.83446 | 0.670574 | 0.009901 | 0.001518 | -0.34473 | 0.016742 | -0.17277 | -0.49344 | -0.50954 | TRUE | FALSE | FALSE | TRUE | TRUE | 7.335142 | 0.344369 | 3.611289 | 10.66346 | 11.02982 |
| SM C16:1 | sphingolip Sphingomy | 2.863751 | -0.22889 | 0.005579 | -0.04262 | -0.12021 | -0.07539 | 4.61E-11 | 0.725621 | 0.387901 | 0.013611 | 0.076164 | 6.72E-10 | 0.828298 | 0.762727 | 0.041788 | 0.158674 | -0.70526 | 0.017189 | -0.13132 | -0.3704 | -0.23229 | TRUE | FALSE | FALSE | TRUE | FALSE | 17.19307 | 0.387422 | 2.998112 | 8.689262 | 5.364366 |
| SM C18:0 | sphingolip Sphingomy | 3.165277 | -0.19953 | -0.00712 | -0.01958 | -0.025 | 0.038103 | 2.91E-08 | 0.668033 | 0.704097 | 0.621841 | 0.390122 | 2.42E-07 | 0.806247 | 0.863839 | 0.745358 | 0.501995 | -0.60728 | -0.02168 | -0.05959 | -0.07608 | 0.115968 | TRUE | FALSE | FALSE | FALSE | FALSE | 14.83242 | 0.49497 | 1.366398 | 1.747692 | 2.676286 |
| SM C18:1 | sphingolip Sphingomy | 2.984159 | -0.29032 | 0.003448 | -0.0497 | -0.04607 | 0.003794 | 1.18E-13 | 0.845762 | 0.366536 | 0.394467 | 0.936055 | 4.13E-12 | 0.919306 | 0.737284 | 0.552254 | 0.957951 | -0.79705 | 0.009465 | -0.13645 | -0.12647 | 0.010415 | TRUE | FALSE | FALSE | FALSE | FALSE | 22.29148 | 0.23925 | 3.504942 | 3.244552 | 0.263294 |
| SM C20:2 | sphingolip Sphingomy | 10.2393 | -0.26224 | -0.00286 | -0.19823 | -0.11376 | -0.16402 | 4.57E-07 | 0.905633 | 0.008419 | 0.122653 | 0.011178 | 2.96E-06 | 0.966377 | 0.290158 | 0.223586 | 0.040754 | -0.54525 | -0.00594 | -0.41216 | -0.23652 | -0.34103 | TRUE | FALSE | FALSE | FALSE | FALSE | 19.93413 | 0.198307 | 14.72893 | 8.204184 | 12.04055 |
| SM C24:0 | sphingolip Sphingomy | 3.436194 | -0.06957 | -0.07956 | 0.035218 | -0.0985 | -0.05808 | 0.065837 | 1.12E-05 | 0.524958 | 0.071081 | 0.223041 | 0.135547 | 8.93E-05 | 0.814023 | 0.146344 | 0.333609 | -0.19818 | -0.22664 | 0.100318 | -0.28059 | -0.16543 | FALSE | TRUE | FALSE | FALSE | FALSE | 4.940736 | 5.6699 | 2.471157 | 7.066335 | 4.107627 |
| SM C24:1 | sphingolip Sphingomy | 3.855143 | -0.05552 | -0.03041 | -0.09757 | -0.1243 | -0.16538 | 0.12987 | 0.0796 | 0.070037 | 0.019074 | 0.00039 | 0.231911 | 0.188244 | 0.437729 | 0.054422 | 0.00244 | -0.16495 | -0.09036 | -0.28989 | -0.36932 | -0.49139 | FALSE | FALSE | FALSE | FALSE | TRUE | 3.923147 | 2.130383 | 6.996723 | 8.997867 | 12.1464 |
| SM C26:0 | sphingolip Sphingomy | 13.93955 | -0.05851 | -0.04605 | -0.09269 | -0.19808 | -0.13747 | 0.337487 | 0.110972 | 0.300754 | 0.024904 | 0.074667 | 0.468732 | 0.239755 | 0.701759 | 0.065047 | 0.15743 | -0.10604 | -0.08346 | -0.16798 | -0.35899 | -0.24915 | FALSE | FALSE | FALSE | FALSE | FALSE | 4.138892 | 3.243258 | 6.635401 | 14.71669 | 9.997508 |
| SM C26:1 | sphingolip Sphingomy | 13.07676 | -0.10345 | 0.00137 | -0.13258 | -0.12471 | -0.15928 | 0.056807 | 0.957347 | 0.096113 | 0.111248 | 0.020299 | 0.124265 | 0.989921 | 0.442626 | 0.207111 | 0.061095 | -0.21077 | 0.002791 | -0.27014 | -0.25409 | -0.32454 | FALSE | FALSE | FALSE | FALSE | FALSE | 7.433723 | 0.095008 | 9.625579 | 9.028754 | 11.67334 |
