## supplementary_table5 for "Molecular characterization of liver steatosis through circulating proteins, metabolites and lipids" (from another version of this preprint: version 1, 2025-12-02)

Table S5. Dietary intake variables when comparing control (n=140, CAP<238 dB/m) versus grade 3 steatosis (n=81, CAP>290 dB/m) using linear regression analysis with age, sex and estimated daily total energy intake as covariates. Asterisks, variables with a confidence interval outside of zero. PDI, Plant-based diet index. Formula: macronutrient/PDI ~ sex + age/10 + steatosis grade + estimated daily total energy intake

| **Dietary intake variable** | **Coefficient** | **Confidence interval** | **p-Value** | **Adjusted p-value** | **Effect Size** |
|---|---|---|---|---|---|
| **Macronutrient** | | | | | |
| Total proteins | 0.2175 | [-4.3901, 4.8250] | 0.9261 | 0.9261 | 0.007 |
| Total fats | 1.8504 | [-1.6008, 5.3016] | 0.2923 | 0.6820 | 0.0571 |
| Total carbohydrates | -8.0584 | [-17.6298, 1.5131] | 0.0986 | 0.6514 | -0.0970 |
| Saturated fatty acids | 0.6911 | [-0.9930, 2.3752] | 0.4201 | 0.7351 | 0.0574 |
| Monounsaturated fatty acids | 1.4622 | [-0.7088, 3.6332] | 0.1861 | 0.6514 | 0.1054 |
| Polyunsaturated fatty acids | 0.0684 | [-1.0762, 1.2131] | 0.9064 | 0.9261 | 0.0118 |
| Trans fatty acids | 0.0316 | [-0.0695, 0.1328] | 0.5390 | 0.7546 | 0.0575 |
| **Food index** | | | | | |
| PDI score | -2.0224 | [-3.5563, -0.4886] * | 0.0099 | / | -0.3354 |
