## supplementary_table6 for "Molecular characterization of liver steatosis through circulating proteins, metabolites and lipids" (from another version of this preprint: version 1, 2025-12-02)

Table S6. Pathway names for significantly altered proteins, metabolites and lipids between control and grade 3 steatosis.

| Pathway name | Pathway source | Pathway Id | Input Id | commonName |
|---|---|---|---|---|
| Cerebral organic acidurias, including diseases | wiki | WP4519 | hmdb:HMDB0000510 | Aminoadipic acid,2-amino-hexanedioic acid |
| Lysine catabolism | reactome | R-HSA-71064 | hmdb:HMDB0000510 | Aminoadipic acid,2-amino-hexanedioic acid |
| Lysine catabolism | wiki | WP5004 | hmdb:HMDB0000510 | Aminoadipic acid,2-amino-hexanedioic acid |
| Lysine degradation | kegg | map00310 | hmdb:HMDB0000510 | Aminoadipic acid,2-amino-hexanedioic acid |
| Vitamin B6-dependent and responsive disorders | wiki | WP4228 | hmdb:HMDB0000510 | Aminoadipic acid,2-amino-hexanedioic acid |
| NR1H2 and NR1H3-mediated signaling | reactome | R-HSA-9024446 | uniprot:P05090 | APOD |
| NR1H2 and NR1H3-mediated signaling | wiki | WP5012 | uniprot:P05090 | APOD |
| NR1H3 & NR1H2 regulate gene expression linked to cholesterol transport and efflux | reactome | R-HSA-9029569 | uniprot:P05090 | APOD |
| Transport of fatty acids | reactome | R-HSA-804914 | uniprot:P05090 | APOD |
| Transport of vitamins, nucleosides, and related molecules | reactome | R-HSA-425397 | uniprot:P05090 | APOD |
| Transport of vitamins, nucleosides, and related molecules | wiki | WP1937 | uniprot:P05090 | APOD |
| Metabolism of fat-soluble vitamins | reactome | R-HSA-6806667 | uniprot:O95445 | APOM |
| Retinoid metabolism and transport | reactome | R-HSA-975634 | uniprot:O95445 | APOM |
| Retinoid metabolism and transport | wiki | WP3839 | uniprot:O95445 | APOM |
| Miscellaneous transport and binding events | reactome | R-HSA-5223345 | uniprot:P25311 | AZGP1 |
| Miscellaneous transport and binding events | wiki | WP3320 | uniprot:P25311 | AZGP1 |
| 16p11.2 distal deletion syndrome | wiki | WP4950 | uniprot:P01024 | C3 |
| Acquired partial lipodystrophy / Barraquer-Simons syndrome | wiki | WP5104 | uniprot:P01024 | C3 |
| Activation of C3 and C5 | reactome | R-HSA-174577 | uniprot:P01024 | C3 |
| Allograft rejection | wiki | WP2328 | uniprot:P01024 | C3 |
| Alternative complement activation | reactome | R-HSA-173736 | uniprot:P01024 | C3 |
| Cell recruitment (pro-inflammatory response) | reactome | R-HSA-9664424 | uniprot:P01024 | C3 |
| Cell recruitment (pro-inflammatory response) | wiki | WP4998 | uniprot:P01024 | C3 |
| Cells and molecules involved in local acute inflammatory response | wiki | WP4493 | uniprot:P01024 | C3 |
| Chronic hyperglycemia impairment of neuron function | wiki | WP5283 | uniprot:P01024 | C3 |
| Complement activation | wiki | WP545 | uniprot:P01024 | C3 |
| Complement and coagulation cascades | wiki | WP558 | uniprot:P01024 | C3 |
| Complement system | wiki | WP2806 | uniprot:P01024 | C3 |
| Complement system in neuronal development and plasticity | wiki | WP5090 | uniprot:P01024 | C3 |
| Extrafollicular and follicular B cell activation by SARS-CoV-2 | wiki | WP5218 | uniprot:P01024 | C3 |
| Initial triggering of complement | reactome | R-HSA-166663 | uniprot:P01024 | C3 |
| Post-translational protein phosphorylation | reactome | R-HSA-8957275 | uniprot:P01024 | C3 |
| Post-translational protein phosphorylation | wiki | WP4110 | uniprot:P01024 | C3 |
| Purinergic signaling in leishmaniasis infection | reactome | R-HSA-9660826 | uniprot:P01024 | C3 |
| Regulation of Complement cascade | reactome | R-HSA-977606 | uniprot:P01024 | C3 |
| Regulation of IGF transport and uptake by IGF binding proteins | wiki | WP2799 | uniprot:P01024 | C3 |
| Regulation of Insulin-like Growth Factor (IGF) transport and uptake by Insulin-like Growth Factor Binding Proteins (IGFBPs) | reactome | R-HSA-381426 | uniprot:P01024 | C3 |
| TYROBP causal network in microglia | wiki | WP3945 | uniprot:P01024 | C3 |
| Allograft rejection | wiki | WP2328 | uniprot:P02748 | C9 |
| Complement activation | wiki | WP545 | uniprot:P02748 | C9 |
| Complement and coagulation cascades | wiki | WP558 | uniprot:P02748 | C9 |
| Complement system | wiki | WP2806 | uniprot:P02748 | C9 |
| Complement system in neuronal development and plasticity | wiki | WP5090 | uniprot:P02748 | C9 |
| Regulation of Complement cascade | reactome | R-HSA-977606 | uniprot:P02748 | C9 |
| Terminal pathway of complement | reactome | R-HSA-166665 | uniprot:P02748 | C9 |
| Complement and coagulation cascades | wiki | WP558 | uniprot:P08603 | CFH |
| Complement system | wiki | WP2806 | uniprot:P08603 | CFH |
| Complement system in neuronal development and plasticity | wiki | WP5090 | uniprot:P08603 | CFH |
| Network map of SARS-CoV-2 signaling pathway | wiki | WP5115 | uniprot:P08603 | CFH |
| Regulation of Complement cascade | reactome | R-HSA-977606 | uniprot:P08603 | CFH |
| Complement and coagulation cascades | wiki | WP558 | uniprot:P05156 | CFI |
| Complement system | wiki | WP2806 | uniprot:P05156 | CFI |
| Complement system in neuronal development and plasticity | wiki | WP5090 | uniprot:P05156 | CFI |
| Network map of SARS-CoV-2 signaling pathway | wiki | WP5115 | uniprot:P05156 | CFI |
| Regulation of Complement cascade | reactome | R-HSA-977606 | uniprot:P05156 | CFI |
| Defective CP causes aceruloplasminemia (ACERULOP) | reactome | R-HSA-5619060 | uniprot:P00450 | CP |

| | | | | |
|---|---|---|---|---|
| Ferroptosis | wiki | WP4313 | uniprot:P00450 | CP |
| Iron metabolism disorders | wiki | WP5172 | uniprot:P00450 | CP |
| Iron uptake and transport | reactome | R-HSA-917937 | uniprot:P00450 | CP |
| Iron uptake and transport | wiki | WP2670 | uniprot:P00450 | CP |
| Metal ion SLC transporters | reactome | R-HSA-425410 | uniprot:P00450 | CP |
| Neurodegeneration with brain iron accumulation (NBIA) subtypes pathway | wiki | WP4577 | uniprot:P00450 | CP |
| Porphyrin Metabolism | kegg | map00860 | uniprot:P00450 | CP |
| Post-translational protein phosphorylation | reactome | R-HSA-8957275 | uniprot:P00450 | CP |
| Post-translational protein phosphorylation | wiki | WP4110 | uniprot:P00450 | CP |
| Regulation of IGF transport and uptake by IGF binding proteins | wiki | WP2799 | uniprot:P00450 | CP |
| Regulation of Insulin-like Growth Factor (IGF) transport and uptake by Insulin-like Growth Factor Binding Proteins (IGFBPs) | reactome | R-HSA-381426 | uniprot:P00450 | CP |
| Vitamin D (calciferol) metabolism | reactome | R-HSA-196791 | uniprot:P02774 | GC |
| Vitamin D (calciferol) metabolism | wiki | WP3685 | uniprot:P02774 | GC |
| Vitamin D metabolism | wiki | WP1531 | uniprot:P02774 | GC |
| Activation of NMDA receptors and postsynaptic events | reactome | R-HSA-442755 | hmdb:HMDB0000123 | Glycine |
| Aflatoxin activation and detoxification | reactome | R-HSA-5423646 | hmdb:HMDB0000123 | Glycine |
| Aflatoxin activation and detoxification | wiki | WP3306 | hmdb:HMDB0000123 | Glycine |
| Alanine, aspartate and glutamate metabolism | kegg | map00250 | hmdb:HMDB0000123 | Glycine |
| Amino Acid conjugation | reactome | R-HSA-156587 | hmdb:HMDB0000123 | Glycine |
| Amino acid conjugation | wiki | WP715 | hmdb:HMDB0000123 | Glycine |
| Amino acid conjugation of benzoic acid | wiki | WP521 | hmdb:HMDB0000123 | Glycine |
| Amino acid transport across the plasma membrane | reactome | R-HSA-352230 | hmdb:HMDB0000123 | Glycine |
| Amino acid transport defects (IEMs) | wiki | WP5029 | hmdb:HMDB0000123 | Glycine |
| Arachidonic acid metabolism | wiki | WP2650 | hmdb:HMDB0000123 | Glycine |
| Arginine and proline metabolism | kegg | map00330 | hmdb:HMDB0000123 | Glycine |
| Aspirin ADME | reactome | R-HSA-9749641 | hmdb:HMDB0000123 | Glycine |
| Bile acid and bile salt metabolism | reactome | R-HSA-194068 | hmdb:HMDB0000123 | Glycine |
| Bile acid and bile salt metabolism | wiki | WP1788 | hmdb:HMDB0000123 | Glycine |
| Biosynthesis of DHA-derived SPMs | reactome | R-HSA-9018677 | hmdb:HMDB0000123 | Glycine |
| Biosynthesis of DHA-derived SPMs | wiki | WP4439 | hmdb:HMDB0000123 | Glycine |
| Biosynthesis of DHA-derived sulfido conjugates | reactome | R-HSA-9026395 | hmdb:HMDB0000123 | Glycine |
| Biosynthesis of maresin conjugates in tissue regeneration (MCTR) | reactome | R-HSA-9026762 | hmdb:HMDB0000123 | Glycine |
| Biosynthesis of protectin and resolvin conjugates in tissue regeneration (PCTR and RCTR) | reactome | R-HSA-9026766 | hmdb:HMDB0000123 | Glycine |
| Carboxyterminal post-translational modifications of tubulin | reactome | R-HSA-8955332 | hmdb:HMDB0000123 | Glycine |
| Carboxyterminal post-translational modifications of tubulin | wiki | WP4075 | hmdb:HMDB0000123 | Glycine |
| Carnitine synthesis | reactome | R-HSA-71262 | hmdb:HMDB0000123 | Glycine |
| Carnitine synthesis | wiki | WP4996 | hmdb:HMDB0000123 | Glycine |
| Choline catabolism | reactome | R-HSA-6798163 | hmdb:HMDB0000123 | Glycine |
| Choline catabolism | wiki | WP4090 | hmdb:HMDB0000123 | Glycine |
| Conjugation of benzoate with glycine | reactome | R-HSA-177135 | hmdb:HMDB0000123 | Glycine |
| Conjugation of carboxylic acids | reactome | R-HSA-159424 | hmdb:HMDB0000123 | Glycine |
| Conjugation of salicylate with glycine | reactome | R-HSA-177128 | hmdb:HMDB0000123 | Glycine |
| Creatine metabolism | reactome | R-HSA-71288 | hmdb:HMDB0000123 | Glycine |
| Creatine metabolism | wiki | WP5002 | hmdb:HMDB0000123 | Glycine |
| Creatine pathway | wiki | WP5190 | hmdb:HMDB0000123 | Glycine |
| CREB1 phosphorylation through NMDA receptor-mediated activation of RAS signaling | reactome | R-HSA-442742 | hmdb:HMDB0000123 | Glycine |
| Cysteine and methionine catabolism | wiki | WP4504 | hmdb:HMDB0000123 | Glycine |
| Cytosolic tRNA aminoacylation | reactome | R-HSA-379716 | hmdb:HMDB0000123 | Glycine |
| Defective GSS causes GSS deficiency | reactome | R-HSA-5579006 | hmdb:HMDB0000123 | Glycine |
| Defective SLC6A19 causes Hartnup disorder (HND) | reactome | R-HSA-5619044 | hmdb:HMDB0000123 | Glycine |
| Defective SLC6A19 causes Hartnup disorder (HND) | reactome | R-HSA-5659735 | hmdb:HMDB0000123 | Glycine |
| Disorders of folate metabolism and transport | wiki | WP4259 | hmdb:HMDB0000123 | Glycine |
| EPH-Ephrin signaling | reactome | R-HSA-2682334 | hmdb:HMDB0000123 | Glycine |
| EPH-Ephrin signaling | wiki | WP3309 | hmdb:HMDB0000123 | Glycine |
| EPHB-mediated forward signaling | reactome | R-HSA-3928662 | hmdb:HMDB0000123 | Glycine |
| Ethanol effects on histone modifications | wiki | WP3996 | hmdb:HMDB0000123 | Glycine |
| Folate metabolism | wiki | WP176 | hmdb:HMDB0000123 | Glycine |
| Gamma-glutamyl cycle for the biosynthesis and degradation of glutathione, including diseases | wiki | WP4518 | hmdb:HMDB0000123 | Glycine |
| Glutathione conjugation | reactome | R-HSA-156590 | hmdb:HMDB0000123 | Glycine |

| | | | | |
|---|---|---|---|---|
| Glutathione metabolism | wiki | WP100 | hmdb:HMDB0000123 | Glycine |
| Glutathione metabolism | kegg | map00480 | hmdb:HMDB0000123 | Glycine |
| Glutathione synthesis and recycling | reactome | R-HSA-174403 | hmdb:HMDB0000123 | Glycine |
| Glycine degradation | reactome | R-HSA-6783984 | hmdb:HMDB0000123 | Glycine |
| Glycine metabolism | wiki | WP1495 | hmdb:HMDB0000123 | Glycine |
| Glyoxylate metabolism | wiki | WP5166 | hmdb:HMDB0000123 | Glycine |
| Glyoxylate metabolism and glycine degradation | reactome | R-HSA-389661 | hmdb:HMDB0000123 | Glycine |
| Glyoxylate metabolism and glycine degradation | wiki | WP3562 | hmdb:HMDB0000123 | Glycine |
| Heme biosynthesis | reactome | R-HSA-189451 | hmdb:HMDB0000123 | Glycine |
| Heme biosynthesis | wiki | WP561 | hmdb:HMDB0000123 | Glycine |
| Hemesynthesis defects and porphyrias | wiki | WP5169 | hmdb:HMDB0000123 | Glycine |
| Long-term potentiation | reactome | R-HSA-9620244 | hmdb:HMDB0000123 | Glycine |
| LTC4-CYSLTR mediated IL4 production | reactome | R-HSA-9664535 | hmdb:HMDB0000123 | Glycine |
| MECP2 and associated Rett syndrome | wiki | WP3584 | hmdb:HMDB0000123 | Glycine |
| Metabolic disorders of biological oxidation enzymes | reactome | R-HSA-5579029 | hmdb:HMDB0000123 | Glycine |
| Metabolic pathways of fibroblasts | wiki | WP5312 | hmdb:HMDB0000123 | Glycine |
| Metabolic reprogramming in colon cancer | wiki | WP4290 | hmdb:HMDB0000123 | Glycine |
| Metabolism of folate and pterines | reactome | R-HSA-196757 | hmdb:HMDB0000123 | Glycine |
| Metabolism of folate and pterines | wiki | WP3806 | hmdb:HMDB0000123 | Glycine |
| Metabolism of ingested SeMet, Sec, MeSec into H2Se | reactome | R-HSA-2408508 | hmdb:HMDB0000123 | Glycine |
| Metabolism of porphyrins | reactome | R-HSA-189445 | hmdb:HMDB0000123 | Glycine |
| Metabolism of porphyrins | wiki | WP1852 | hmdb:HMDB0000123 | Glycine |
| Methionine Metabolism | kegg | map00270 | hmdb:HMDB0000123 | Glycine |
| Methionine metabolism leading to sulfur amino acids and related disorders | wiki | WP4292 | hmdb:HMDB0000123 | Glycine |
| Mitochondrial tRNA aminoacylation | reactome | R-HSA-379726 | hmdb:HMDB0000123 | Glycine |
| Na+/Cl- dependent neurotransmitter transporters | reactome | R-HSA-442660 | hmdb:HMDB0000123 | Glycine |
| Negative regulation of NMDA receptor-mediated neuronal transmission | reactome | R-HSA-9617324 | hmdb:HMDB0000123 | Glycine |
| Neurotransmitter receptors and postsynaptic signal transmission | wiki | WP2754 | hmdb:HMDB0000123 | Glycine |
| Nitrogen metabolism | kegg | map00910 | hmdb:HMDB0000123 | Glycine |
| Nucleobase biosynthesis | wiki | WP4084 | hmdb:HMDB0000123 | Glycine |
| Nucleotide biosynthesis | reactome | R-HSA-8956320 | hmdb:HMDB0000123 | Glycine |
| One-carbon metabolism | wiki | WP241 | hmdb:HMDB0000123 | Glycine |
| Paracetamol ADME | reactome | R-HSA-9753281 | hmdb:HMDB0000123 | Glycine |
| Porphyrin Metabolism | kegg | map00860 | hmdb:HMDB0000123 | Glycine |
| Post NMDA receptor activation events | reactome | R-HSA-438064 | hmdb:HMDB0000123 | Glycine |
| Primary bile acid biosynthesis | kegg | map00120 | hmdb:HMDB0000123 | Glycine |
| Protein-protein interactions at synapses | reactome | R-HSA-6794362 | hmdb:HMDB0000123 | Glycine |
| Proton-coupled neutral amino acid transporters | reactome | R-HSA-428559 | hmdb:HMDB0000123 | Glycine |
| Proximal tubule transport | wiki | WP4917 | hmdb:HMDB0000123 | Glycine |
| Purine ribonucleoside monophosphate biosynthesis | reactome | R-HSA-73817 | hmdb:HMDB0000123 | Glycine |
| Ras activation upon Ca2+ influx through NMDA receptor | reactome | R-HSA-442982 | hmdb:HMDB0000123 | Glycine |
| Recycling of bile acids and salts | reactome | R-HSA-159418 | hmdb:HMDB0000123 | Glycine |
| SALM protein interactions at the synapse | wiki | WP3812 | hmdb:HMDB0000123 | Glycine |
| Serine metabolism | wiki | WP4688 | hmdb:HMDB0000123 | Glycine |
| Synaptic adhesion-like molecules | reactome | R-HSA-8849932 | hmdb:HMDB0000123 | Glycine |
| Synaptic adhesion-like molecules | wiki | WP4054 | hmdb:HMDB0000123 | Glycine |
| Synthesis of bile acids and bile salts | reactome | R-HSA-192105 | hmdb:HMDB0000123 | Glycine |
| Synthesis of bile acids and bile salts via 7alpha-hydroxycholesterol | reactome | R-HSA-193368 | hmdb:HMDB0000123 | Glycine |
| Synthesis of Leukotrienes (LT) and Eoxins (EX) | reactome | R-HSA-2142691 | hmdb:HMDB0000123 | Glycine |
| Threonine catabolism | reactome | R-HSA-8849175 | hmdb:HMDB0000123 | Glycine |
| Threonine catabolism | wiki | WP3803 | hmdb:HMDB0000123 | Glycine |
| Trans-sulfuration, one-carbon metabolism and related pathways | wiki | WP2525 | hmdb:HMDB0000123 | Glycine |
| tRNA Aminoacylation | reactome | R-HSA-379724 | hmdb:HMDB0000123 | Glycine |
| tRNA aminoacylation | wiki | WP1938 | hmdb:HMDB0000123 | Glycine |
| Unblocking of NMDA receptors, glutamate binding and activation | reactome | R-HSA-438066 | hmdb:HMDB0000123 | Glycine |
| Variant SLC6A14 may confer susceptibility towards obesity | reactome | R-HSA-5619094 | hmdb:HMDB0000123 | Glycine |
| Scavenging of heme from plasma | reactome | R-HSA-2168880 | uniprot:P00739 | HPR |
| Leucine, isoleucine and valine metabolism | wiki | WP4686 | hmdb:HMDB0000736 | Isobutyryl-L-carnitine |
| Leucine, isoleucine and valine metabolism | wiki | WP4686 | hmdb:HMDB0000688 | Isovalerylcarnitine,CAR 5:0 |

| | | | | |
|---|---|---|---|---|
| Network map of SARS-CoV-2 signaling pathway | wiki | WP5115 | uniprot:Q06033 | ITIH3 |
| Platelet degranulation | reactome | R-HSA-114608 | uniprot:Q06033 | ITIH3 |
| Response to elevated platelet cytosolic Ca ions | wiki | WP1903 | uniprot:Q06033 | ITIH3 |
| Response to elevated platelet cytosolic Ca2+ | reactome | R-HSA-76005 | uniprot:Q06033 | ITIH3 |
| Network map of SARS-CoV-2 signaling pathway | wiki | WP5115 | uniprot:Q14624 | ITIH4 |
| Platelet degranulation | reactome | R-HSA-114608 | uniprot:Q14624 | ITIH4 |
| Response to elevated platelet cytosolic Ca ions | wiki | WP1903 | uniprot:Q14624 | ITIH4 |
| Response to elevated platelet cytosolic Ca2+ | reactome | R-HSA-76005 | uniprot:Q14624 | ITIH4 |
| Alanine and aspartate metabolism | wiki | WP106 | hmdb:HMDB0000161 | L-Alanine |
| Alanine metabolism | reactome | R-HSA-8964540 | hmdb:HMDB0000161 | L-Alanine |
| Alanine metabolism | wiki | WP4980 | hmdb:HMDB0000161 | L-Alanine |
| Alanine, aspartate and glutamate metabolism | kegg | map00250 | hmdb:HMDB0000161 | L-Alanine |
| Amino acid metabolism pathway excerpt: histidine catabolism extension | wiki | WP4661 | hmdb:HMDB0000161 | L-Alanine |
| Amino acid transport across the plasma membrane | reactome | R-HSA-352230 | hmdb:HMDB0000161 | L-Alanine |
| Arginine and proline metabolism | kegg | map00330 | hmdb:HMDB0000161 | L-Alanine |
| Biosynthesis and turnover of 1-deoxy-sphingoid bases | wiki | WP5179 | hmdb:HMDB0000161 | L-Alanine |
| Cytosolic tRNA aminoacylation | reactome | R-HSA-379716 | hmdb:HMDB0000161 | L-Alanine |
| Defective SLC6A19 causes Hartnup disorder (HND) | reactome | R-HSA-5619044 | hmdb:HMDB0000161 | L-Alanine |
| Defective SLC6A19 causes Hartnup disorder (HND) | reactome | R-HSA-5659735 | hmdb:HMDB0000161 | L-Alanine |
| Glutamate and glutamine metabolism | reactome | R-HSA-8964539 | hmdb:HMDB0000161 | L-Alanine |
| Glutamate and glutamine metabolism | wiki | WP4992 | hmdb:HMDB0000161 | L-Alanine |
| Glutathione metabolism | kegg | map00480 | hmdb:HMDB0000161 | L-Alanine |
| Glyoxylate metabolism | wiki | WP5166 | hmdb:HMDB0000161 | L-Alanine |
| Glyoxylate metabolism and glycine degradation | reactome | R-HSA-389661 | hmdb:HMDB0000161 | L-Alanine |
| Glyoxylate metabolism and glycine degradation | wiki | WP3562 | hmdb:HMDB0000161 | L-Alanine |
| Krebs cycle disorders | wiki | WP4236 | hmdb:HMDB0000161 | L-Alanine |
| Metabolism of amine-derived hormones | reactome | R-HSA-209776 | hmdb:HMDB0000161 | L-Alanine |
| Metabolism of amine-derived hormones | wiki | WP3548 | hmdb:HMDB0000161 | L-Alanine |
| Metabolism of ingested SeMet, Sec, MeSec into H2Se | reactome | R-HSA-2408508 | hmdb:HMDB0000161 | L-Alanine |
| Mitochondrial iron-sulfur cluster biogenesis | reactome | R-HSA-1362409 | hmdb:HMDB0000161 | L-Alanine |
| Mitochondrial iron-sulfur cluster biogenesis | wiki | WP2702 | hmdb:HMDB0000161 | L-Alanine |
| Mitochondrial tRNA aminoacylation | reactome | R-HSA-379726 | hmdb:HMDB0000161 | L-Alanine |
| Molybdenum cofactor biosynthesis | reactome | R-HSA-947581 | hmdb:HMDB0000161 | L-Alanine |
| mRNA, protein, and metabolite inducation pathway by cyclosporin A | wiki | WP3953 | hmdb:HMDB0000161 | L-Alanine |
| Na+/Cl- dependent neurotransmitter transporters | reactome | R-HSA-442660 | hmdb:HMDB0000161 | L-Alanine |
| NAD biosynthesis II (from tryptophan) | wiki | WP2485 | hmdb:HMDB0000161 | L-Alanine |
| Nucleobase catabolism | wiki | WP4042 | hmdb:HMDB0000161 | L-Alanine |
| Nucleotide catabolism | reactome | R-HSA-8956319 | hmdb:HMDB0000161 | L-Alanine |
| Phenylalanine and tyrosine metabolism | reactome | R-HSA-8963691 | hmdb:HMDB0000161 | L-Alanine |
| Phenylalanine and tyrosine metabolism | wiki | WP4981 | hmdb:HMDB0000161 | L-Alanine |
| Phenylalanine metabolism | reactome | R-HSA-8964208 | hmdb:HMDB0000161 | L-Alanine |
| Proton-coupled neutral amino acid transporters | reactome | R-HSA-428559 | hmdb:HMDB0000161 | L-Alanine |
| Pyrimidine catabolism | reactome | R-HSA-73621 | hmdb:HMDB0000161 | L-Alanine |
| Selenocompound metabolism | kegg | map00450 | hmdb:HMDB0000161 | L-Alanine |
| Thyroxine biosynthesis | reactome | R-HSA-209968 | hmdb:HMDB0000161 | L-Alanine |
| tRNA Aminoacylation | reactome | R-HSA-379724 | hmdb:HMDB0000161 | L-Alanine |
| tRNA aminoacylation | wiki | WP1938 | hmdb:HMDB0000161 | L-Alanine |
| Tryptophan catabolism | reactome | R-HSA-71240 | hmdb:HMDB0000161 | L-Alanine |
| Tryptophan catabolism | wiki | WP4994 | hmdb:HMDB0000161 | L-Alanine |
| Tryptophan catabolism leading to NAD+ production | wiki | WP4210 | hmdb:HMDB0000161 | L-Alanine |
| Tryptophan metabolism | kegg | map00380 | hmdb:HMDB0000161 | L-Alanine |
| Urea cycle and associated pathways | wiki | WP4595 | hmdb:HMDB0000161 | L-Alanine |
| Variant SLC6A14 may confer susceptibility towards obesity | reactome | R-HSA-5619094 | hmdb:HMDB0000161 | L-Alanine |
| Beta-oxidation of pristanoyl-CoA | reactome | R-HSA-389887 | hmdb:HMDB0000062 | L-Carnitine,Carnitine |
| Biosynthesis of unsaturated fatty acids | kegg | map01040 | hmdb:HMDB0000062 | L-Carnitine,Carnitine |
| Carnitine metabolism | reactome | R-HSA-200425 | hmdb:HMDB0000062 | L-Carnitine,Carnitine |
| Carnitine metabolism | wiki | WP4061 | hmdb:HMDB0000062 | L-Carnitine,Carnitine |
| Carnitine synthesis | reactome | R-HSA-71262 | hmdb:HMDB0000062 | L-Carnitine,Carnitine |
| Carnitine synthesis | wiki | WP4996 | hmdb:HMDB0000062 | L-Carnitine,Carnitine |

| | | | | |
|---|---|---|---|---|
| Diet-dependent trimethylamine/trimethylamine N-oxide metabolism | wiki | WP5219 | hmdb:HMDB0000062 | L-Carnitine,Carnitine |
| Fatty acid Metabolism | kegg | map00071 | hmdb:HMDB0000062 | L-Carnitine,Carnitine |
| Mitochondrial fatty acid oxidation disorders | wiki | WP5123 | hmdb:HMDB0000062 | L-Carnitine,Carnitine |
| Organic cation transport | reactome | R-HSA-549127 | hmdb:HMDB0000062 | L-Carnitine,Carnitine |
| Organic cation/anion/zwitterion transport | reactome | R-HSA-549132 | hmdb:HMDB0000062 | L-Carnitine,Carnitine |
| Peroxisomal lipid metabolism | reactome | R-HSA-390918 | hmdb:HMDB0000062 | L-Carnitine,Carnitine |
| Peroxisomal lipid metabolism | wiki | WP1878 | hmdb:HMDB0000062 | L-Carnitine,Carnitine |
| Signaling by Retinoic Acid | reactome | R-HSA-5362517 | hmdb:HMDB0000062 | L-Carnitine,Carnitine |
| Signaling by Retinoic Acid | wiki | WP4421 | hmdb:HMDB0000062 | L-Carnitine,Carnitine |
| Thermogenesis | wiki | WP4321 | hmdb:HMDB0000062 | L-Carnitine,Carnitine |
| 10q22q23 copy number variation | wiki | WP5402 | hmdb:HMDB0000148 | L-Glutamic acid,Glutamate |
| Activation of AMPA receptors | reactome | R-HSA-399710 | hmdb:HMDB0000148 | L-Glutamic acid,Glutamate |
| Activation of Ca-permeable Kainate Receptor | reactome | R-HSA-451308 | hmdb:HMDB0000148 | L-Glutamic acid,Glutamate |
| Activation of kainate receptors upon glutamate binding | reactome | R-HSA-451326 | hmdb:HMDB0000148 | L-Glutamic acid,Glutamate |
| Activation of kainate receptors upon glutamate binding | wiki | WP4456 | hmdb:HMDB0000148 | L-Glutamic acid,Glutamate |
| Activation of Na-permeable kainate receptors | reactome | R-HSA-451307 | hmdb:HMDB0000148 | L-Glutamic acid,Glutamate |
| Activation of NMDA receptors and postsynaptic events | reactome | R-HSA-442755 | hmdb:HMDB0000148 | L-Glutamic acid,Glutamate |
| Aflatoxin activation and detoxification | reactome | R-HSA-5423646 | hmdb:HMDB0000148 | L-Glutamic acid,Glutamate |
| Aflatoxin activation and detoxification | wiki | WP3306 | hmdb:HMDB0000148 | L-Glutamic acid,Glutamate |
| Alanine metabolism | reactome | R-HSA-8964540 | hmdb:HMDB0000148 | L-Glutamic acid,Glutamate |
| Alanine metabolism | wiki | WP4980 | hmdb:HMDB0000148 | L-Glutamic acid,Glutamate |
| Alanine, aspartate and glutamate metabolism | kegg | map00250 | hmdb:HMDB0000148 | L-Glutamic acid,Glutamate |
| Amino acid metabolism in triple-negative breast cancer cells | wiki | WP5213 | hmdb:HMDB0000148 | L-Glutamic acid,Glutamate |
| Amino acid metabolism pathway excerpt: histidine catabolism extension | wiki | WP4661 | hmdb:HMDB0000148 | L-Glutamic acid,Glutamate |
| Amino acid transport across the plasma membrane | reactome | R-HSA-352230 | hmdb:HMDB0000148 | L-Glutamic acid,Glutamate |
| Amino acid transport defects (IEMs) | wiki | WP5029 | hmdb:HMDB0000148 | L-Glutamic acid,Glutamate |
| Amino Sugar Metabolism | kegg | map00520 | hmdb:HMDB0000148 | L-Glutamic acid,Glutamate |
| Amyotrophic lateral sclerosis (ALS) | wiki | WP2447 | hmdb:HMDB0000148 | L-Glutamic acid,Glutamate |
| Arachidonic acid metabolism | wiki | WP2650 | hmdb:HMDB0000148 | L-Glutamic acid,Glutamate |
| Arachidonic Acid Metabolism | kegg | map00590 | hmdb:HMDB0000148 | L-Glutamic acid,Glutamate |
| Arginine and proline metabolism | kegg | map00330 | hmdb:HMDB0000148 | L-Glutamic acid,Glutamate |
| Aspartate and asparagine metabolism | reactome | R-HSA-8963693 | hmdb:HMDB0000148 | L-Glutamic acid,Glutamate |
| Aspartate and asparagine metabolism | wiki | WP4979 | hmdb:HMDB0000148 | L-Glutamic acid,Glutamate |
| Astrocytic Glutamate-Glutamine Uptake And Metabolism | reactome | R-HSA-210455 | hmdb:HMDB0000148 | L-Glutamic acid,Glutamate |
| Azathioprine ADME | reactome | R-HSA-9748787 | hmdb:HMDB0000148 | L-Glutamic acid,Glutamate |
| beta-Alanine metabolism | kegg | map00410 | hmdb:HMDB0000148 | L-Glutamic acid,Glutamate |
| Biogenic amine synthesis | wiki | WP550 | hmdb:HMDB0000148 | L-Glutamic acid,Glutamate |
| Biomarkers for urea cycle disorders | wiki | WP4583 | hmdb:HMDB0000148 | L-Glutamic acid,Glutamate |
| Biosynthesis of DHA-derived SPMs | reactome | R-HSA-9018677 | hmdb:HMDB0000148 | L-Glutamic acid,Glutamate |
| Biosynthesis of DHA-derived SPMs | wiki | WP4439 | hmdb:HMDB0000148 | L-Glutamic acid,Glutamate |
| Biosynthesis of DHA-derived sulfido conjugates | reactome | R-HSA-9026395 | hmdb:HMDB0000148 | L-Glutamic acid,Glutamate |
| Biosynthesis of maresin conjugates in tissue regeneration (MCTR) | reactome | R-HSA-9026762 | hmdb:HMDB0000148 | L-Glutamic acid,Glutamate |
| Biosynthesis of protectin and resolvin conjugates in tissue regeneration (PCTR and RCTR) | reactome | R-HSA-9026766 | hmdb:HMDB0000148 | L-Glutamic acid,Glutamate |
| Branched-chain amino acid catabolism | reactome | R-HSA-70895 | hmdb:HMDB0000148 | L-Glutamic acid,Glutamate |
| Branched-chain amino acid catabolism | wiki | WP3556 | hmdb:HMDB0000148 | L-Glutamic acid,Glutamate |
| Cannabinoid receptor signaling | wiki | WP3869 | hmdb:HMDB0000148 | L-Glutamic acid,Glutamate |
| Carboxyterminal post-translational modifications of tubulin | reactome | R-HSA-8955332 | hmdb:HMDB0000148 | L-Glutamic acid,Glutamate |
| Carboxyterminal post-translational modifications of tubulin | wiki | WP4075 | hmdb:HMDB0000148 | L-Glutamic acid,Glutamate |
| Class C/3 (Metabotropic glutamate/pheromone receptors) | reactome | R-HSA-420499 | hmdb:HMDB0000148 | L-Glutamic acid,Glutamate |
| Class C/3 (metabotropic glutamate/pheromone receptors) | wiki | WP4418 | hmdb:HMDB0000148 | L-Glutamic acid,Glutamate |
| CREB1 phosphorylation through NMDA receptor-mediated activation of RAS signaling | reactome | R-HSA-442742 | hmdb:HMDB0000148 | L-Glutamic acid,Glutamate |
| Cysteine and methionine catabolism | wiki | WP4504 | hmdb:HMDB0000148 | L-Glutamic acid,Glutamate |
| Cytosolic tRNA aminoacylation | reactome | R-HSA-379716 | hmdb:HMDB0000148 | L-Glutamic acid,Glutamate |
| Defective GCLC causes HAGGSD | reactome | R-HSA-5578999 | hmdb:HMDB0000148 | L-Glutamic acid,Glutamate |
| Defective SLC1A1 is implicated in schizophrenia 18 (SCZD18) and dicarboxylic aminoaciduria (DCBXA) | reactome | R-HSA-5619067 | hmdb:HMDB0000148 | L-Glutamic acid,Glutamate |
| Defective SLC1A3 causes episodic ataxia 6 (EA6) | reactome | R-HSA-5619062 | hmdb:HMDB0000148 | L-Glutamic acid,Glutamate |
| Degradation of cysteine and homocysteine | reactome | R-HSA-1614558 | hmdb:HMDB0000148 | L-Glutamic acid,Glutamate |
| Degradation of GABA | reactome | R-HSA-916853 | hmdb:HMDB0000148 | L-Glutamic acid,Glutamate |
| Disorders of folate metabolism and transport | wiki | WP4259 | hmdb:HMDB0000148 | L-Glutamic acid,Glutamate |

| | | | | |
|---|---|---|---|---|
| EPH-Ephrin signaling | reactome | R-HSA-2682334 | hmdb:HMDB0000148 | L-Glutamic acid,Glutamate |
| EPH-Ephrin signaling | wiki | WP3309 | hmdb:HMDB0000148 | L-Glutamic acid,Glutamate |
| EPHB-mediated forward signaling | reactome | R-HSA-3928662 | hmdb:HMDB0000148 | L-Glutamic acid,Glutamate |
| ERK pathway in Huntington's disease | wiki | WP3853 | hmdb:HMDB0000148 | L-Glutamic acid,Glutamate |
| GABA metabolism (aka GHB) | wiki | WP4157 | hmdb:HMDB0000148 | L-Glutamic acid,Glutamate |
| GABA receptor signaling | wiki | WP4159 | hmdb:HMDB0000148 | L-Glutamic acid,Glutamate |
| GABA synthesis | reactome | R-HSA-888568 | hmdb:HMDB0000148 | L-Glutamic acid,Glutamate |
| GABA synthesis, release, reuptake and degradation | reactome | R-HSA-888590 | hmdb:HMDB0000148 | L-Glutamic acid,Glutamate |
| GABA synthesis, release, reuptake and degradation | wiki | WP2685 | hmdb:HMDB0000148 | L-Glutamic acid,Glutamate |
| Gluconeogenesis | reactome | R-HSA-70263 | hmdb:HMDB0000148 | L-Glutamic acid,Glutamate |
| Glucose metabolism | reactome | R-HSA-70326 | hmdb:HMDB0000148 | L-Glutamic acid,Glutamate |
| Glucose metabolism | wiki | WP4093 | hmdb:HMDB0000148 | L-Glutamic acid,Glutamate |
| Glutamate and glutamine metabolism | reactome | R-HSA-8964539 | hmdb:HMDB0000148 | L-Glutamic acid,Glutamate |
| Glutamate and glutamine metabolism | wiki | WP4992 | hmdb:HMDB0000148 | L-Glutamic acid,Glutamate |
| Glutamate binding, activation of AMPA receptors and synaptic plasticity | reactome | R-HSA-399721 | hmdb:HMDB0000148 | L-Glutamic acid,Glutamate |
| Glutamate binding, activation of AMPA receptors and synaptic plasticity | wiki | WP4459 | hmdb:HMDB0000148 | L-Glutamic acid,Glutamate |
| Glutamate Neurotransmitter Release Cycle | reactome | R-HSA-210500 | hmdb:HMDB0000148 | L-Glutamic acid,Glutamate |
| Glutathione conjugation | reactome | R-HSA-156590 | hmdb:HMDB0000148 | L-Glutamic acid,Glutamate |
| Glutathione metabolism | wiki | WP100 | hmdb:HMDB0000148 | L-Glutamic acid,Glutamate |
| Glutathione metabolism | kegg | map00480 | hmdb:HMDB0000148 | L-Glutamic acid,Glutamate |
| Glutathione synthesis and recycling | reactome | R-HSA-174403 | hmdb:HMDB0000148 | L-Glutamic acid,Glutamate |
| Histidine catabolism | reactome | R-HSA-70921 | hmdb:HMDB0000148 | L-Glutamic acid,Glutamate |
| Histidine catabolism | wiki | WP4988 | hmdb:HMDB0000148 | L-Glutamic acid,Glutamate |
| Histidine metabolism | kegg | map00340 | hmdb:HMDB0000148 | L-Glutamic acid,Glutamate |
| Interconversion of nucleotide di- and triphosphates | reactome | R-HSA-499943 | hmdb:HMDB0000148 | L-Glutamic acid,Glutamate |
| Interconversion of nucleotide di- and triphosphates | wiki | WP4059 | hmdb:HMDB0000148 | L-Glutamic acid,Glutamate |
| Ionotropic activity of kainate receptors | reactome | R-HSA-451306 | hmdb:HMDB0000148 | L-Glutamic acid,Glutamate |
| Long-term potentiation | reactome | R-HSA-9620244 | hmdb:HMDB0000148 | L-Glutamic acid,Glutamate |
| LTC4-CYSLTR mediated IL4 production | reactome | R-HSA-9664535 | hmdb:HMDB0000148 | L-Glutamic acid,Glutamate |
| Lysine catabolism | reactome | R-HSA-71064 | hmdb:HMDB0000148 | L-Glutamic acid,Glutamate |
| Lysine catabolism | wiki | WP5004 | hmdb:HMDB0000148 | L-Glutamic acid,Glutamate |
| Lysine degradation | kegg | map00310 | hmdb:HMDB0000148 | L-Glutamic acid,Glutamate |
| Metabolic disorders of biological oxidation enzymes | reactome | R-HSA-5579029 | hmdb:HMDB0000148 | L-Glutamic acid,Glutamate |
| Metabolic pathways of fibroblasts | wiki | WP5312 | hmdb:HMDB0000148 | L-Glutamic acid,Glutamate |
| Metabolism of folate and pterines | reactome | R-HSA-196757 | hmdb:HMDB0000148 | L-Glutamic acid,Glutamate |
| Metabolism of folate and pterines | wiki | WP3806 | hmdb:HMDB0000148 | L-Glutamic acid,Glutamate |
| Metabolism overview | wiki | WP3602 | hmdb:HMDB0000148 | L-Glutamic acid,Glutamate |
| Methionine de novo and salvage pathway | wiki | WP3580 | hmdb:HMDB0000148 | L-Glutamic acid,Glutamate |
| Methionine Metabolism | kegg | map00270 | hmdb:HMDB0000148 | L-Glutamic acid,Glutamate |
| Mitochondrial tRNA aminoacylation | reactome | R-HSA-379726 | hmdb:HMDB0000148 | L-Glutamic acid,Glutamate |
| NAD biosynthesis II (from tryptophan) | wiki | WP2485 | hmdb:HMDB0000148 | L-Glutamic acid,Glutamate |
| Negative regulation of NMDA receptor-mediated neuronal transmission | reactome | R-HSA-9617324 | hmdb:HMDB0000148 | L-Glutamic acid,Glutamate |
| Neurotransmitter release cycle | reactome | R-HSA-112310 | hmdb:HMDB0000148 | L-Glutamic acid,Glutamate |
| Neurotransmitter release cycle | wiki | WP1871 | hmdb:HMDB0000148 | L-Glutamic acid,Glutamate |
| Neurotransmitter uptake and metabolism In glial cells | reactome | R-HSA-112313 | hmdb:HMDB0000148 | L-Glutamic acid,Glutamate |
| Neurotransmitter uptake and metabolism In glial cells | wiki | WP1872 | hmdb:HMDB0000148 | L-Glutamic acid,Glutamate |
| Nicotinate and nicotinamide metabolism | kegg | map00760 | hmdb:HMDB0000148 | L-Glutamic acid,Glutamate |
| Nicotinate metabolism | reactome | R-HSA-196807 | hmdb:HMDB0000148 | L-Glutamic acid,Glutamate |
| Nicotinate metabolism | wiki | WP4097 | hmdb:HMDB0000148 | L-Glutamic acid,Glutamate |
| Nitrogen metabolism | kegg | map00910 | hmdb:HMDB0000148 | L-Glutamic acid,Glutamate |
| NO/cGMP/PKG mediated neuroprotection | wiki | WP4008 | hmdb:HMDB0000148 | L-Glutamic acid,Glutamate |
| Nucleobase biosynthesis | wiki | WP4084 | hmdb:HMDB0000148 | L-Glutamic acid,Glutamate |
| Nucleotide biosynthesis | reactome | R-HSA-8956320 | hmdb:HMDB0000148 | L-Glutamic acid,Glutamate |
| One carbon pool by folate | kegg | map00670 | hmdb:HMDB0000148 | L-Glutamic acid,Glutamate |
| Orexin receptor pathway | wiki | WP5094 | hmdb:HMDB0000148 | L-Glutamic acid,Glutamate |
| Organic anion transporters | reactome | R-HSA-428643 | hmdb:HMDB0000148 | L-Glutamic acid,Glutamate |
| Paracetamol ADME | reactome | R-HSA-9753281 | hmdb:HMDB0000148 | L-Glutamic acid,Glutamate |
| Phenylalanine and tyrosine metabolism | reactome | R-HSA-8963691 | hmdb:HMDB0000148 | L-Glutamic acid,Glutamate |
| Phenylalanine and tyrosine metabolism | wiki | WP4981 | hmdb:HMDB0000148 | L-Glutamic acid,Glutamate |

| | | | | |
|---|---|---|---|---|
| Phenylalanine and Tyrosine Metabolism | kegg | map00360 | hmdb:HMDB0000148 | L-Glutamic acid,Glutamate |
| Phosphodiesterases in neuronal function | wiki | WP4222 | hmdb:HMDB0000148 | L-Glutamic acid,Glutamate |
| Post NMDA receptor activation events | reactome | R-HSA-438064 | hmdb:HMDB0000148 | L-Glutamic acid,Glutamate |
| Presynaptic function of Kainate receptors | reactome | R-HSA-500657 | hmdb:HMDB0000148 | L-Glutamic acid,Glutamate |
| Proline catabolism | reactome | R-HSA-70688 | hmdb:HMDB0000148 | L-Glutamic acid,Glutamate |
| Proline catabolism | wiki | WP5009 | hmdb:HMDB0000148 | L-Glutamic acid,Glutamate |
| Propanoate metabolism | kegg | map00640 | hmdb:HMDB0000148 | L-Glutamic acid,Glutamate |
| Protein-protein interactions at synapses | reactome | R-HSA-6794362 | hmdb:HMDB0000148 | L-Glutamic acid,Glutamate |
| Purine metabolism and related disorders | wiki | WP4224 | hmdb:HMDB0000148 | L-Glutamic acid,Glutamate |
| Purine ribonucleoside monophosphate biosynthesis | reactome | R-HSA-73817 | hmdb:HMDB0000148 | L-Glutamic acid,Glutamate |
| Pyrimidine biosynthesis | reactome | R-HSA-500753 | hmdb:HMDB0000148 | L-Glutamic acid,Glutamate |
| Pyrimidine metabolism and related diseases | wiki | WP4225 | hmdb:HMDB0000148 | L-Glutamic acid,Glutamate |
| Ras activation upon Ca2+ influx through NMDA receptor | reactome | R-HSA-442982 | hmdb:HMDB0000148 | L-Glutamic acid,Glutamate |
| SALM protein interactions at the synapse | wiki | WP3812 | hmdb:HMDB0000148 | L-Glutamic acid,Glutamate |
| Sensory perception of sweet, bitter, and umami (glutamate) taste | reactome | R-HSA-9717207 | hmdb:HMDB0000148 | L-Glutamic acid,Glutamate |
| Sensory perception of taste | reactome | R-HSA-9717189 | hmdb:HMDB0000148 | L-Glutamic acid,Glutamate |
| Sensory processing of sound | reactome | R-HSA-9659379 | hmdb:HMDB0000148 | L-Glutamic acid,Glutamate |
| Sensory processing of sound by inner hair cells of the cochlea | reactome | R-HSA-9662360 | hmdb:HMDB0000148 | L-Glutamic acid,Glutamate |
| Serine biosynthesis | reactome | R-HSA-977347 | hmdb:HMDB0000148 | L-Glutamic acid,Glutamate |
| Serine biosynthesis | wiki | WP4435 | hmdb:HMDB0000148 | L-Glutamic acid,Glutamate |
| Serine metabolism | wiki | WP4688 | hmdb:HMDB0000148 | L-Glutamic acid,Glutamate |
| Sulfur amino acid metabolism | reactome | R-HSA-1614635 | hmdb:HMDB0000148 | L-Glutamic acid,Glutamate |
| Sulfur amino acid metabolism | wiki | WP3574 | hmdb:HMDB0000148 | L-Glutamic acid,Glutamate |
| Synaptic adhesion-like molecules | reactome | R-HSA-8849932 | hmdb:HMDB0000148 | L-Glutamic acid,Glutamate |
| Synaptic adhesion-like molecules | wiki | WP4054 | hmdb:HMDB0000148 | L-Glutamic acid,Glutamate |
| Synthesis of Leukotrienes (LT) and Eoxins (EX) | reactome | R-HSA-2142691 | hmdb:HMDB0000148 | L-Glutamic acid,Glutamate |
| Synthesis of substrates in N-glycan biosythesis | reactome | R-HSA-446219 | hmdb:HMDB0000148 | L-Glutamic acid,Glutamate |
| Synthesis of UDP-N-acetyl-glucosamine | reactome | R-HSA-446210 | hmdb:HMDB0000148 | L-Glutamic acid,Glutamate |
| TP53 Regulates Metabolic Genes | reactome | R-HSA-5628897 | hmdb:HMDB0000148 | L-Glutamic acid,Glutamate |
| TP53 Regulates Metabolic Genes | wiki | WP3349 | hmdb:HMDB0000148 | L-Glutamic acid,Glutamate |
| Trans-sulfuration pathway | wiki | WP2333 | hmdb:HMDB0000148 | L-Glutamic acid,Glutamate |
| tRNA Aminoacylation | reactome | R-HSA-379724 | hmdb:HMDB0000148 | L-Glutamic acid,Glutamate |
| tRNA aminoacylation | wiki | WP1938 | hmdb:HMDB0000148 | L-Glutamic acid,Glutamate |
| Tryptophan catabolism | reactome | R-HSA-71240 | hmdb:HMDB0000148 | L-Glutamic acid,Glutamate |
| Tryptophan catabolism | wiki | WP4994 | hmdb:HMDB0000148 | L-Glutamic acid,Glutamate |
| Tryptophan metabolism | kegg | map00380 | hmdb:HMDB0000148 | L-Glutamic acid,Glutamate |
| Tyrosine catabolism | reactome | R-HSA-8963684 | hmdb:HMDB0000148 | L-Glutamic acid,Glutamate |
| Tyrosine metabolism | kegg | map00350 | hmdb:HMDB0000148 | L-Glutamic acid,Glutamate |
| Unblocking of NMDA receptors, glutamate binding and activation | reactome | R-HSA-438066 | hmdb:HMDB0000148 | L-Glutamic acid,Glutamate |
| Urea cycle | reactome | R-HSA-70635 | hmdb:HMDB0000148 | L-Glutamic acid,Glutamate |
| Urea cycle | wiki | WP4973 | hmdb:HMDB0000148 | L-Glutamic acid,Glutamate |
| Urea cycle and associated pathways | wiki | WP4595 | hmdb:HMDB0000148 | L-Glutamic acid,Glutamate |
| Urea cycle and metabolism of amino groups | wiki | WP497 | hmdb:HMDB0000148 | L-Glutamic acid,Glutamate |
| Urea cycle and related diseases | wiki | WP4571 | hmdb:HMDB0000148 | L-Glutamic acid,Glutamate |
| Valine, leucine and isoleucine degradation | kegg | map00280 | hmdb:HMDB0000148 | L-Glutamic acid,Glutamate |
| Vitamin B6-dependent and responsive disorders | wiki | WP4228 | hmdb:HMDB0000148 | L-Glutamic acid,Glutamate |
| Alanine, aspartate and glutamate metabolism | kegg | map00250 | hmdb:HMDB0000641 | L-Glutamine,Glutamine |
| Amino Acid conjugation | reactome | R-HSA-156587 | hmdb:HMDB0000641 | L-Glutamine,Glutamine |
| Amino acid conjugation | wiki | WP715 | hmdb:HMDB0000641 | L-Glutamine,Glutamine |
| Amino acid metabolism in triple-negative breast cancer cells | wiki | WP5213 | hmdb:HMDB0000641 | L-Glutamine,Glutamine |
| Amino acid transport across the plasma membrane | reactome | R-HSA-352230 | hmdb:HMDB0000641 | L-Glutamine,Glutamine |
| Amino Sugar Metabolism | kegg | map00520 | hmdb:HMDB0000641 | L-Glutamine,Glutamine |
| Arginine and proline metabolism | kegg | map00330 | hmdb:HMDB0000641 | L-Glutamine,Glutamine |
| Aspartate and asparagine metabolism | reactome | R-HSA-8963693 | hmdb:HMDB0000641 | L-Glutamine,Glutamine |
| Aspartate and asparagine metabolism | wiki | WP4979 | hmdb:HMDB0000641 | L-Glutamine,Glutamine |
| Astrocytic Glutamate-Glutamine Uptake And Metabolism | reactome | R-HSA-210455 | hmdb:HMDB0000641 | L-Glutamine,Glutamine |
| Azathioprine ADME | reactome | R-HSA-9748787 | hmdb:HMDB0000641 | L-Glutamine,Glutamine |
| Biomarkers for urea cycle disorders | wiki | WP4583 | hmdb:HMDB0000641 | L-Glutamine,Glutamine |
| Conjugation of carboxylic acids | reactome | R-HSA-159424 | hmdb:HMDB0000641 | L-Glutamine,Glutamine |

| | | | | |
|---|---|---|---|---|
| Conjugation of phenylacetate with glutamine | reactome | R-HSA-177162 | hmdb:HMDB0000641 | L-Glutamine,Glutamine |
| Cytosolic tRNA aminoacylation | reactome | R-HSA-379716 | hmdb:HMDB0000641 | L-Glutamine,Glutamine |
| Defective SLC6A19 causes Hartnup disorder (HND) | reactome | R-HSA-5619044 | hmdb:HMDB0000641 | L-Glutamine,Glutamine |
| Defective SLC6A19 causes Hartnup disorder (HND) | reactome | R-HSA-5659735 | hmdb:HMDB0000641 | L-Glutamine,Glutamine |
| Diseases associated with glycosylation precursor biosynthesis | reactome | R-HSA-5609975 | hmdb:HMDB0000641 | L-Glutamine,Glutamine |
| Ferroptosis | wiki | WP4313 | hmdb:HMDB0000641 | L-Glutamine,Glutamine |
| Glutamate and glutamine metabolism | reactome | R-HSA-8964539 | hmdb:HMDB0000641 | L-Glutamine,Glutamine |
| Glutamate and glutamine metabolism | wiki | WP4992 | hmdb:HMDB0000641 | L-Glutamine,Glutamine |
| Glutamate Neurotransmitter Release Cycle | reactome | R-HSA-210500 | hmdb:HMDB0000641 | L-Glutamine,Glutamine |
| Interconversion of nucleotide di- and triphosphates | reactome | R-HSA-499943 | hmdb:HMDB0000641 | L-Glutamine,Glutamine |
| Interconversion of nucleotide di- and triphosphates | wiki | WP4059 | hmdb:HMDB0000641 | L-Glutamine,Glutamine |
| Metabolic pathways of fibroblasts | wiki | WP5312 | hmdb:HMDB0000641 | L-Glutamine,Glutamine |
| Metabolism overview | wiki | WP3602 | hmdb:HMDB0000641 | L-Glutamine,Glutamine |
| Mitochondrial tRNA aminoacylation | reactome | R-HSA-379726 | hmdb:HMDB0000641 | L-Glutamine,Glutamine |
| mRNA, protein, and metabolite inducation pathway by cyclosporin A | wiki | WP3953 | hmdb:HMDB0000641 | L-Glutamine,Glutamine |
| Na+/Cl- dependent neurotransmitter transporters | reactome | R-HSA-442660 | hmdb:HMDB0000641 | L-Glutamine,Glutamine |
| NAD biosynthesis II (from tryptophan) | wiki | WP2485 | hmdb:HMDB0000641 | L-Glutamine,Glutamine |
| Neurotransmitter release cycle | reactome | R-HSA-112310 | hmdb:HMDB0000641 | L-Glutamine,Glutamine |
| Neurotransmitter release cycle | wiki | WP1871 | hmdb:HMDB0000641 | L-Glutamine,Glutamine |
| Neurotransmitter uptake and metabolism In glial cells | reactome | R-HSA-112313 | hmdb:HMDB0000641 | L-Glutamine,Glutamine |
| Neurotransmitter uptake and metabolism In glial cells | wiki | WP1872 | hmdb:HMDB0000641 | L-Glutamine,Glutamine |
| Nicotinate and nicotinamide metabolism | kegg | map00760 | hmdb:HMDB0000641 | L-Glutamine,Glutamine |
| Nicotinate metabolism | reactome | R-HSA-196807 | hmdb:HMDB0000641 | L-Glutamine,Glutamine |
| Nicotinate metabolism | wiki | WP4097 | hmdb:HMDB0000641 | L-Glutamine,Glutamine |
| Nitrogen metabolism | kegg | map00910 | hmdb:HMDB0000641 | L-Glutamine,Glutamine |
| Nucleobase biosynthesis | wiki | WP4084 | hmdb:HMDB0000641 | L-Glutamine,Glutamine |
| Nucleotide biosynthesis | reactome | R-HSA-8956320 | hmdb:HMDB0000641 | L-Glutamine,Glutamine |
| Phenylalanine and Tyrosine Metabolism | kegg | map00360 | hmdb:HMDB0000641 | L-Glutamine,Glutamine |
| Purine metabolism and related disorders | wiki | WP4224 | hmdb:HMDB0000641 | L-Glutamine,Glutamine |
| Purine ribonucleoside monophosphate biosynthesis | reactome | R-HSA-73817 | hmdb:HMDB0000641 | L-Glutamine,Glutamine |
| Pyrimidine biosynthesis | reactome | R-HSA-500753 | hmdb:HMDB0000641 | L-Glutamine,Glutamine |
| Pyrimidine metabolism | wiki | WP4022 | hmdb:HMDB0000641 | L-Glutamine,Glutamine |
| Pyrimidine metabolism and related diseases | wiki | WP4225 | hmdb:HMDB0000641 | L-Glutamine,Glutamine |
| Sterol regulatory element-binding proteins (SREBP) signaling | wiki | WP1982 | hmdb:HMDB0000641 | L-Glutamine,Glutamine |
| Synaptic vesicle pathway | wiki | WP2267 | hmdb:HMDB0000641 | L-Glutamine,Glutamine |
| Synthesis of substrates in N-glycan biosythesis | reactome | R-HSA-446219 | hmdb:HMDB0000641 | L-Glutamine,Glutamine |
| Synthesis of UDP-N-acetyl-glucosamine | reactome | R-HSA-446210 | hmdb:HMDB0000641 | L-Glutamine,Glutamine |
| TP53 Regulates Metabolic Genes | reactome | R-HSA-5628897 | hmdb:HMDB0000641 | L-Glutamine,Glutamine |
| TP53 Regulates Metabolic Genes | wiki | WP3349 | hmdb:HMDB0000641 | L-Glutamine,Glutamine |
| tRNA Aminoacylation | reactome | R-HSA-379724 | hmdb:HMDB0000641 | L-Glutamine,Glutamine |
| tRNA aminoacylation | wiki | WP1938 | hmdb:HMDB0000641 | L-Glutamine,Glutamine |
| Urea cycle and associated pathways | wiki | WP4595 | hmdb:HMDB0000641 | L-Glutamine,Glutamine |
| Urea cycle and related diseases | wiki | WP4571 | hmdb:HMDB0000641 | L-Glutamine,Glutamine |
| Amino acid transport across the plasma membrane | reactome | R-HSA-352230 | hmdb:HMDB0000172 | L-Isoleucine,Isoleucine,2S-Amino-3S-methylpentanoic acid |
| Branched-chain amino acid catabolism | reactome | R-HSA-70895 | hmdb:HMDB0000172 | L-Isoleucine,Isoleucine,2S-Amino-3S-methylpentanoic acid |
| Branched-chain amino acid catabolism | wiki | WP3556 | hmdb:HMDB0000172 | L-Isoleucine,Isoleucine,2S-Amino-3S-methylpentanoic acid |
| Cytosolic tRNA aminoacylation | reactome | R-HSA-379716 | hmdb:HMDB0000172 | L-Isoleucine,Isoleucine,2S-Amino-3S-methylpentanoic acid |
| Defective SLC6A19 causes Hartnup disorder (HND) | reactome | R-HSA-5619044 | hmdb:HMDB0000172 | L-Isoleucine,Isoleucine,2S-Amino-3S-methylpentanoic acid |
| Defective SLC6A19 causes Hartnup disorder (HND) | reactome | R-HSA-5659735 | hmdb:HMDB0000172 | L-Isoleucine,Isoleucine,2S-Amino-3S-methylpentanoic acid |
| Glucose homeostasis | wiki | WP661 | hmdb:HMDB0000172 | L-Isoleucine,Isoleucine,2S-Amino-3S-methylpentanoic acid |
| Leucine, isoleucine and valine metabolism | wiki | WP4686 | hmdb:HMDB0000172 | L-Isoleucine,Isoleucine,2S-Amino-3S-methylpentanoic acid |
| Mitochondrial tRNA aminoacylation | reactome | R-HSA-379726 | hmdb:HMDB0000172 | L-Isoleucine,Isoleucine,2S-Amino-3S-methylpentanoic acid |
| mRNA, protein, and metabolite inducation pathway by cyclosporin A | wiki | WP3953 | hmdb:HMDB0000172 | L-Isoleucine,Isoleucine,2S-Amino-3S-methylpentanoic acid |
| Na+/Cl- dependent neurotransmitter transporters | reactome | R-HSA-442660 | hmdb:HMDB0000172 | L-Isoleucine,Isoleucine,2S-Amino-3S-methylpentanoic acid |
| tRNA Aminoacylation | reactome | R-HSA-379724 | hmdb:HMDB0000172 | L-Isoleucine,Isoleucine,2S-Amino-3S-methylpentanoic acid |
| tRNA aminoacylation | wiki | WP1938 | hmdb:HMDB0000172 | L-Isoleucine,Isoleucine,2S-Amino-3S-methylpentanoic acid |
| Tryptophan catabolism | reactome | R-HSA-71240 | hmdb:HMDB0000172 | L-Isoleucine,Isoleucine,2S-Amino-3S-methylpentanoic acid |
| Tryptophan catabolism | wiki | WP4994 | hmdb:HMDB0000172 | L-Isoleucine,Isoleucine,2S-Amino-3S-methylpentanoic acid |
| Valine, leucine and isoleucine degradation | kegg | map00280 | hmdb:HMDB0000172 | L-Isoleucine,Isoleucine,2S-Amino-3S-methylpentanoic acid |

| | | | | |
|---|---|---|---|---|
| Variant SLC6A14 may confer susceptibility towards obesity | reactome | R-HSA-5619094 | hmdb:HMDB0000172 | L-Isoleucine,Isoleucine,2S-Amino-3S-methylpentanoic acid |
| Amino acid transport across the plasma membrane | reactome | R-HSA-352230 | hmdb:HMDB0000687 | L-Leucine |
| Amino acids regulate mTORC1 | reactome | R-HSA-9639288 | hmdb:HMDB0000687 | L-Leucine |
| Amino acids regulate mTORC1 | wiki | WP5015 | hmdb:HMDB0000687 | L-Leucine |
| Branched-chain amino acid catabolism | reactome | R-HSA-70895 | hmdb:HMDB0000687 | L-Leucine |
| Branched-chain amino acid catabolism | wiki | WP3556 | hmdb:HMDB0000687 | L-Leucine |
| Cytosolic tRNA aminoacylation | reactome | R-HSA-379716 | hmdb:HMDB0000687 | L-Leucine |
| Defective SLC3A1 causes cystinuria (CSNU) | reactome | R-HSA-5619113 | hmdb:HMDB0000687 | L-Leucine |
| Defective SLC6A19 causes Hartnup disorder (HND) | reactome | R-HSA-5619044 | hmdb:HMDB0000687 | L-Leucine |
| Defective SLC6A19 causes Hartnup disorder (HND) | reactome | R-HSA-5659735 | hmdb:HMDB0000687 | L-Leucine |
| Defective SLC7A7 causes lysinuric protein intolerance (LPI) | reactome | R-HSA-5660862 | hmdb:HMDB0000687 | L-Leucine |
| Defective SLC7A9 causes cystinuria (CSNU) | reactome | R-HSA-5660883 | hmdb:HMDB0000687 | L-Leucine |
| Disorders in ketone body synthesis | wiki | WP5175 | hmdb:HMDB0000687 | L-Leucine |
| Leucine, isoleucine and valine metabolism | wiki | WP4686 | hmdb:HMDB0000687 | L-Leucine |
| Mitochondrial tRNA aminoacylation | reactome | R-HSA-379726 | hmdb:HMDB0000687 | L-Leucine |
| mRNA, protein, and metabolite inducation pathway by cyclosporin A | wiki | WP3953 | hmdb:HMDB0000687 | L-Leucine |
| Na+/Cl- dependent neurotransmitter transporters | reactome | R-HSA-442660 | hmdb:HMDB0000687 | L-Leucine |
| tRNA Aminoacylation | reactome | R-HSA-379724 | hmdb:HMDB0000687 | L-Leucine |
| tRNA aminoacylation | wiki | WP1938 | hmdb:HMDB0000687 | L-Leucine |
| Tryptophan catabolism | reactome | R-HSA-71240 | hmdb:HMDB0000687 | L-Leucine |
| Tryptophan catabolism | wiki | WP4994 | hmdb:HMDB0000687 | L-Leucine |
| Valine, leucine and isoleucine degradation | kegg | map00280 | hmdb:HMDB0000687 | L-Leucine |
| Variant SLC6A14 may confer susceptibility towards obesity | reactome | R-HSA-5619094 | hmdb:HMDB0000687 | L-Leucine |
| Amino acid transport across the plasma membrane | reactome | R-HSA-352230 | hmdb:HMDB0000159 | L-Phenylalanine,Phenylalanine |
| Biogenic amine synthesis | wiki | WP550 | hmdb:HMDB0000159 | L-Phenylalanine,Phenylalanine |
| Biosynthesis and regeneration of tetrahydrobiopterin and catabolism of phenylalanine | wiki | WP4156 | hmdb:HMDB0000159 | L-Phenylalanine,Phenylalanine |
| Cytosolic tRNA aminoacylation | reactome | R-HSA-379716 | hmdb:HMDB0000159 | L-Phenylalanine,Phenylalanine |
| Defective SLC6A19 causes Hartnup disorder (HND) | reactome | R-HSA-5619044 | hmdb:HMDB0000159 | L-Phenylalanine,Phenylalanine |
| Defective SLC6A19 causes Hartnup disorder (HND) | reactome | R-HSA-5659735 | hmdb:HMDB0000159 | L-Phenylalanine,Phenylalanine |
| Glucose homeostasis | wiki | WP661 | hmdb:HMDB0000159 | L-Phenylalanine,Phenylalanine |
| Lac-Phe pathway | wiki | WP5239 | hmdb:HMDB0000159 | L-Phenylalanine,Phenylalanine |
| Metabolism of cofactors | reactome | R-HSA-8978934 | hmdb:HMDB0000159 | L-Phenylalanine,Phenylalanine |
| Metabolism of cofactors | wiki | WP4990 | hmdb:HMDB0000159 | L-Phenylalanine,Phenylalanine |
| Mitochondrial tRNA aminoacylation | reactome | R-HSA-379726 | hmdb:HMDB0000159 | L-Phenylalanine,Phenylalanine |
| Mitochondrial Uncoupling | reactome | R-HSA-166187 | hmdb:HMDB0000159 | L-Phenylalanine,Phenylalanine |
| Na+/Cl- dependent neurotransmitter transporters | reactome | R-HSA-442660 | hmdb:HMDB0000159 | L-Phenylalanine,Phenylalanine |
| Oleoyl-phe metabolism | reactome | R-HSA-9673163 | hmdb:HMDB0000159 | L-Phenylalanine,Phenylalanine |
| Phenylalanine and tyrosine metabolism | reactome | R-HSA-8963691 | hmdb:HMDB0000159 | L-Phenylalanine,Phenylalanine |
| Phenylalanine and tyrosine metabolism | wiki | WP4981 | hmdb:HMDB0000159 | L-Phenylalanine,Phenylalanine |
| Phenylalanine and Tyrosine Metabolism | kegg | map00360 | hmdb:HMDB0000159 | L-Phenylalanine,Phenylalanine |
| Phenylalanine metabolism | reactome | R-HSA-8964208 | hmdb:HMDB0000159 | L-Phenylalanine,Phenylalanine |
| Phenylketonuria | reactome | R-HSA-2160456 | hmdb:HMDB0000159 | L-Phenylalanine,Phenylalanine |
| Tetrahydrobiopterin (BH4) synthesis, recycling, salvage and regulation | reactome | R-HSA-1474151 | hmdb:HMDB0000159 | L-Phenylalanine,Phenylalanine |
| tRNA Aminoacylation | reactome | R-HSA-379724 | hmdb:HMDB0000159 | L-Phenylalanine,Phenylalanine |
| tRNA aminoacylation | wiki | WP1938 | hmdb:HMDB0000159 | L-Phenylalanine,Phenylalanine |
| Tryptophan catabolism | reactome | R-HSA-71240 | hmdb:HMDB0000159 | L-Phenylalanine,Phenylalanine |
| Tryptophan catabolism | wiki | WP4994 | hmdb:HMDB0000159 | L-Phenylalanine,Phenylalanine |
| Tyrosine metabolism and related disorders | wiki | WP4506 | hmdb:HMDB0000159 | L-Phenylalanine,Phenylalanine |
| Amino acid transport across the plasma membrane | reactome | R-HSA-352230 | hmdb:HMDB0000929 | L-Tryptophan,Tryptophan |
| Biogenic amine synthesis | wiki | WP550 | hmdb:HMDB0000929 | L-Tryptophan,Tryptophan |
| Biosynthesis and regeneration of tetrahydrobiopterin and catabolism of phenylalanine | wiki | WP4156 | hmdb:HMDB0000929 | L-Tryptophan,Tryptophan |
| Cerebral organic acidurias, including diseases | wiki | WP4519 | hmdb:HMDB0000929 | L-Tryptophan,Tryptophan |
| Cytosolic tRNA aminoacylation | reactome | R-HSA-379716 | hmdb:HMDB0000929 | L-Tryptophan,Tryptophan |
| Defective SLC6A19 causes Hartnup disorder (HND) | reactome | R-HSA-5619044 | hmdb:HMDB0000929 | L-Tryptophan,Tryptophan |
| Defective SLC6A19 causes Hartnup disorder (HND) | reactome | R-HSA-5659735 | hmdb:HMDB0000929 | L-Tryptophan,Tryptophan |
| IDO metabolic pathway | wiki | WP5414 | hmdb:HMDB0000929 | L-Tryptophan,Tryptophan |
| Metabolism of amine-derived hormones | reactome | R-HSA-209776 | hmdb:HMDB0000929 | L-Tryptophan,Tryptophan |
| Metabolism of amine-derived hormones | wiki | WP3548 | hmdb:HMDB0000929 | L-Tryptophan,Tryptophan |
| Mitochondrial tRNA aminoacylation | reactome | R-HSA-379726 | hmdb:HMDB0000929 | L-Tryptophan,Tryptophan |

| | | | | |
|---|---|---|---|---|
| Monoamine transport | wiki | WP727 | hmdb:HMDB0000929 | L-Tryptophan,Tryptophan |
| mRNA, protein, and metabolite inducation pathway by cyclosporin A | wiki | WP3953 | hmdb:HMDB0000929 | L-Tryptophan,Tryptophan |
| Na+/Cl- dependent neurotransmitter transporters | reactome | R-HSA-442660 | hmdb:HMDB0000929 | L-Tryptophan,Tryptophan |
| NAD biosynthesis II (from tryptophan) | wiki | WP2485 | hmdb:HMDB0000929 | L-Tryptophan,Tryptophan |
| NAD+ biosynthetic pathways | wiki | WP3645 | hmdb:HMDB0000929 | L-Tryptophan,Tryptophan |
| NAD+ metabolism | wiki | WP3644 | hmdb:HMDB0000929 | L-Tryptophan,Tryptophan |
| Neurotransmitter disorders | wiki | WP4220 | hmdb:HMDB0000929 | L-Tryptophan,Tryptophan |
| Serotonin and melatonin biosynthesis | reactome | R-HSA-209931 | hmdb:HMDB0000929 | L-Tryptophan,Tryptophan |
| Serotonin transporter activity | wiki | WP1455 | hmdb:HMDB0000929 | L-Tryptophan,Tryptophan |
| T cell modulation in pancreatic cancer | wiki | WP5078 | hmdb:HMDB0000929 | L-Tryptophan,Tryptophan |
| tRNA Aminoacylation | reactome | R-HSA-379724 | hmdb:HMDB0000929 | L-Tryptophan,Tryptophan |
| tRNA aminoacylation | wiki | WP1938 | hmdb:HMDB0000929 | L-Tryptophan,Tryptophan |
| Tryptophan catabolism | reactome | R-HSA-71240 | hmdb:HMDB0000929 | L-Tryptophan,Tryptophan |
| Tryptophan catabolism | wiki | WP4994 | hmdb:HMDB0000929 | L-Tryptophan,Tryptophan |
| Tryptophan catabolism leading to NAD+ production | wiki | WP4210 | hmdb:HMDB0000929 | L-Tryptophan,Tryptophan |
| Tryptophan metabolism | wiki | WP465 | hmdb:HMDB0000929 | L-Tryptophan,Tryptophan |
| Tryptophan metabolism | kegg | map00380 | hmdb:HMDB0000929 | L-Tryptophan,Tryptophan |
| Variant SLC6A14 may confer susceptibility towards obesity | reactome | R-HSA-5619094 | hmdb:HMDB0000929 | L-Tryptophan,Tryptophan |
| Amino acid transport across the plasma membrane | reactome | R-HSA-352230 | hmdb:HMDB0000158 | L-Tyrosine |
| Biogenic amine synthesis | wiki | WP550 | hmdb:HMDB0000158 | L-Tyrosine |
| Biomarkers for urea cycle disorders | wiki | WP4583 | hmdb:HMDB0000158 | L-Tyrosine |
| Biosynthesis and regeneration of tetrahydrobiopterin and catabolism of phenylalanine | wiki | WP4156 | hmdb:HMDB0000158 | L-Tyrosine |
| Carboxyterminal post-translational modifications of tubulin | reactome | R-HSA-8955332 | hmdb:HMDB0000158 | L-Tyrosine |
| Carboxyterminal post-translational modifications of tubulin | wiki | WP4075 | hmdb:HMDB0000158 | L-Tyrosine |
| Catecholamine biosynthesis | reactome | R-HSA-209905 | hmdb:HMDB0000158 | L-Tyrosine |
| Cytosolic tRNA aminoacylation | reactome | R-HSA-379716 | hmdb:HMDB0000158 | L-Tyrosine |
| Defective SLC6A19 causes Hartnup disorder (HND) | reactome | R-HSA-5619044 | hmdb:HMDB0000158 | L-Tyrosine |
| Defective SLC6A19 causes Hartnup disorder (HND) | reactome | R-HSA-5659735 | hmdb:HMDB0000158 | L-Tyrosine |
| Dopamine metabolism | wiki | WP2436 | hmdb:HMDB0000158 | L-Tyrosine |
| Glucose homeostasis | wiki | WP661 | hmdb:HMDB0000158 | L-Tyrosine |
| GPR143 in melanocytes and retinal pigment epithelium cells | wiki | WP4941 | hmdb:HMDB0000158 | L-Tyrosine |
| Melanin biosynthesis | reactome | R-HSA-5662702 | hmdb:HMDB0000158 | L-Tyrosine |
| Melanin biosynthesis | wiki | WP3377 | hmdb:HMDB0000158 | L-Tyrosine |
| Metabolism of amine-derived hormones | reactome | R-HSA-209776 | hmdb:HMDB0000158 | L-Tyrosine |
| Metabolism of amine-derived hormones | wiki | WP3548 | hmdb:HMDB0000158 | L-Tyrosine |
| Mitochondrial tRNA aminoacylation | reactome | R-HSA-379726 | hmdb:HMDB0000158 | L-Tyrosine |
| mRNA, protein, and metabolite inducation pathway by cyclosporin A | wiki | WP3953 | hmdb:HMDB0000158 | L-Tyrosine |
| Na+/Cl- dependent neurotransmitter transporters | reactome | R-HSA-442660 | hmdb:HMDB0000158 | L-Tyrosine |
| Neurotransmitter disorders | wiki | WP4220 | hmdb:HMDB0000158 | L-Tyrosine |
| Nicotine effect on dopaminergic neurons | wiki | WP1602 | hmdb:HMDB0000158 | L-Tyrosine |
| Parkinson's disease pathway | wiki | WP2371 | hmdb:HMDB0000158 | L-Tyrosine |
| Phenylalanine and tyrosine metabolism | reactome | R-HSA-8963691 | hmdb:HMDB0000158 | L-Tyrosine |
| Phenylalanine and tyrosine metabolism | wiki | WP4981 | hmdb:HMDB0000158 | L-Tyrosine |
| Phenylalanine and Tyrosine Metabolism | kegg | map00360 | hmdb:HMDB0000158 | L-Tyrosine |
| Phenylalanine metabolism | reactome | R-HSA-8964208 | hmdb:HMDB0000158 | L-Tyrosine |
| Prader-Willi and Angelman syndrome | wiki | WP3998 | hmdb:HMDB0000158 | L-Tyrosine |
| Thyroxine (thyroid hormone) production | wiki | WP1981 | hmdb:HMDB0000158 | L-Tyrosine |
| Thyroxine biosynthesis | reactome | R-HSA-209968 | hmdb:HMDB0000158 | L-Tyrosine |
| tRNA Aminoacylation | reactome | R-HSA-379724 | hmdb:HMDB0000158 | L-Tyrosine |
| tRNA aminoacylation | wiki | WP1938 | hmdb:HMDB0000158 | L-Tyrosine |
| Tryptophan catabolism | reactome | R-HSA-71240 | hmdb:HMDB0000158 | L-Tyrosine |
| Tryptophan catabolism | wiki | WP4994 | hmdb:HMDB0000158 | L-Tyrosine |
| Tyrosine catabolism | reactome | R-HSA-8963684 | hmdb:HMDB0000158 | L-Tyrosine |
| Tyrosine metabolism | kegg | map00350 | hmdb:HMDB0000158 | L-Tyrosine |
| Tyrosine metabolism and related disorders | wiki | WP4506 | hmdb:HMDB0000158 | L-Tyrosine |
| Acrylamide biotransformation and exposure biomarkers | wiki | WP4233 | hmdb:HMDB0000883 | L-Valine,valine |
| Amino acid transport across the plasma membrane | reactome | R-HSA-352230 | hmdb:HMDB0000883 | L-Valine,valine |
| Branched-chain amino acid catabolism | reactome | R-HSA-70895 | hmdb:HMDB0000883 | L-Valine,valine |
| Branched-chain amino acid catabolism | wiki | WP3556 | hmdb:HMDB0000883 | L-Valine,valine |

| | | | | |
|---|---|---|---|---|
| Cytosolic tRNA aminoacylation | reactome | R-HSA-379716 | hmdb:HMDB0000883 | L-Valine,valine |
| Defective SLC6A19 causes Hartnup disorder (HND) | reactome | R-HSA-5619044 | hmdb:HMDB0000883 | L-Valine,valine |
| Defective SLC6A19 causes Hartnup disorder (HND) | reactome | R-HSA-5659735 | hmdb:HMDB0000883 | L-Valine,valine |
| Glucose homeostasis | wiki | WP661 | hmdb:HMDB0000883 | L-Valine,valine |
| Leucine, isoleucine and valine metabolism | wiki | WP4686 | hmdb:HMDB0000883 | L-Valine,valine |
| Mitochondrial tRNA aminoacylation | reactome | R-HSA-379726 | hmdb:HMDB0000883 | L-Valine,valine |
| mRNA, protein, and metabolite inducation pathway by cyclosporin A | wiki | WP3953 | hmdb:HMDB0000883 | L-Valine,valine |
| Na+/Cl- dependent neurotransmitter transporters | reactome | R-HSA-442660 | hmdb:HMDB0000883 | L-Valine,valine |
| Propanoate metabolism | kegg | map00640 | hmdb:HMDB0000883 | L-Valine,valine |
| Pyrimidine metabolism and related diseases | wiki | WP4225 | hmdb:HMDB0000883 | L-Valine,valine |
| tRNA Aminoacylation | reactome | R-HSA-379724 | hmdb:HMDB0000883 | L-Valine,valine |
| tRNA aminoacylation | wiki | WP1938 | hmdb:HMDB0000883 | L-Valine,valine |
| Tryptophan catabolism | reactome | R-HSA-71240 | hmdb:HMDB0000883 | L-Valine,valine |
| Tryptophan catabolism | wiki | WP4994 | hmdb:HMDB0000883 | L-Valine,valine |
| Valine, leucine and isoleucine degradation | kegg | map00280 | hmdb:HMDB0000883 | L-Valine,valine |
| Variant SLC6A14 may confer susceptibility towards obesity | reactome | R-HSA-5619094 | hmdb:HMDB0000883 | L-Valine,valine |
| Network map of SARS-CoV-2 signaling pathway | wiki | WP5115 | uniprot:P02750 | LRG1 |
| 16p11.2 proximal deletion syndrome | wiki | WP4949 | hmdb:HMDB0012108 | LysoPC(17:0/0:0),LPC(14:1) |
| Phosphatidylcholine catabolism | wiki | WP4195 | hmdb:HMDB0012108 | LysoPC(17:0/0:0),LPC(14:1) |
| Phospholipid Biosynthesis | kegg | map00564 | hmdb:HMDB0012108 | LysoPC(17:0/0:0),LPC(14:1) |
| 16p11.2 proximal deletion syndrome | wiki | WP4949 | hmdb:HMDB0010385 | LysoPC(18:1(11Z)/0:0),LPC(14:1) |
| Phosphatidylcholine catabolism | wiki | WP4195 | hmdb:HMDB0010385 | LysoPC(18:1(11Z)/0:0),LPC(14:1) |
| Phospholipid Biosynthesis | kegg | map00564 | hmdb:HMDB0010385 | LysoPC(18:1(11Z)/0:0),LPC(14:1) |
| Linoleic acid metabolism affected by SARS-CoV-2 | wiki | WP4853 | hmdb:HMDB0010386 | LysoPC(18:2(9Z,12Z)/0:0),Omega-6,LPC(18:2) |
| 16p11.2 proximal deletion syndrome | wiki | WP4949 | hmdb:HMDB0010395 | LysoPC(20:4(5Z,8Z,11Z,14Z)/0:0),LPC(14:1) |
| Phosphatidylcholine catabolism | wiki | WP4195 | hmdb:HMDB0010395 | LysoPC(20:4(5Z,8Z,11Z,14Z)/0:0),LPC(14:1) |
| Phospholipid Biosynthesis | kegg | map00564 | hmdb:HMDB0010395 | LysoPC(20:4(5Z,8Z,11Z,14Z)/0:0),LPC(14:1) |
| 16p11.2 proximal deletion syndrome | wiki | WP4949 | hmdb:HMDB0010396 | LysoPC(20:4(8Z,11Z,14Z,17Z)/0:0),LPC(14:1) |
| Phosphatidylcholine catabolism | wiki | WP4195 | hmdb:HMDB0010396 | LysoPC(20:4(8Z,11Z,14Z,17Z)/0:0),LPC(14:1) |
| Phospholipid Biosynthesis | kegg | map00564 | hmdb:HMDB0010396 | LysoPC(20:4(8Z,11Z,14Z,17Z)/0:0),LPC(14:1) |
| 16p11.2 proximal deletion syndrome | wiki | WP4949 | hmdb:HMDB0010405 | LysoPC(24:0/0:0),LPC(14:1) |
| Phosphatidylcholine catabolism | wiki | WP4195 | hmdb:HMDB0010405 | LysoPC(24:0/0:0),LPC(14:1) |
| Phospholipid Biosynthesis | kegg | map00564 | hmdb:HMDB0010405 | LysoPC(24:0/0:0),LPC(14:1) |
| Acetylcholine synthesis | wiki | WP528 | hmdb:HMDB0007882 | PC(14:0/20:3(8Z,11Z,14Z)),PC(14:0_20:0) |
| Phospholipid Biosynthesis | kegg | map00564 | hmdb:HMDB0007882 | PC(14:0/20:3(8Z,11Z,14Z)),PC(14:0_20:0) |
| Acetylcholine synthesis | wiki | WP528 | hmdb:HMDB0007940 | PC(15:0/18:2(9Z,12Z)),PC(14:0_20:0) |
| Phospholipid Biosynthesis | kegg | map00564 | hmdb:HMDB0007940 | PC(15:0/18:2(9Z,12Z)),PC(14:0_20:0) |
| Acetylcholine synthesis | wiki | WP528 | hmdb:HMDB0007960 | PC(15:0/24:1(15Z)) |
| Phospholipid Biosynthesis | kegg | map00564 | hmdb:HMDB0007960 | PC(15:0/24:1(15Z)) |
| Acetylcholine synthesis | wiki | WP528 | hmdb:HMDB0008091 | PC(18:1(11Z)/24:0),PC(14:0_20:0) |
| Phospholipid Biosynthesis | kegg | map00564 | hmdb:HMDB0008091 | PC(18:1(11Z)/24:0),PC(14:0_20:0) |
| Acetylcholine synthesis | wiki | WP528 | hmdb:HMDB0008092 | PC(18:1(11Z)/24:1(15Z)),PC(14:0_20:0) |
| Phospholipid Biosynthesis | kegg | map00564 | hmdb:HMDB0008092 | PC(18:1(11Z)/24:1(15Z)),PC(14:0_20:0) |
| Acetylcholine synthesis | wiki | WP528 | hmdb:HMDB0008308 | PC(20:1(11Z)/20:1(11Z)),PC(14:0_20:0) |
| Phospholipid Biosynthesis | kegg | map00564 | hmdb:HMDB0008308 | PC(20:1(11Z)/20:1(11Z)),PC(14:0_20:0) |
| Acetylcholine synthesis | wiki | WP528 | hmdb:HMDB0008416 | PC(20:3(8Z,11Z,14Z)/22:4(7Z,10Z,13Z,16Z)),PC(14:0_20:0) |
| Phospholipid Biosynthesis | kegg | map00564 | hmdb:HMDB0008416 | PC(20:3(8Z,11Z,14Z)/22:4(7Z,10Z,13Z,16Z)),PC(14:0_20:0) |
| Acetylcholine synthesis | wiki | WP528 | hmdb:HMDB0008444 | PC(20:4(5Z,8Z,11Z,14Z)/20:4(8Z,11Z,14Z,17Z)) |
| Phospholipid Biosynthesis | kegg | map00564 | hmdb:HMDB0008444 | PC(20:4(5Z,8Z,11Z,14Z)/20:4(8Z,11Z,14Z,17Z)) |
| Acetylcholine synthesis | wiki | WP528 | hmdb:HMDB0008507 | PC(20:5(5Z,8Z,11Z,14Z,17Z)/20:3(5Z,8Z,11Z)) |
| Phospholipid Biosynthesis | kegg | map00564 | hmdb:HMDB0008507 | PC(20:5(5Z,8Z,11Z,14Z,17Z)/20:3(5Z,8Z,11Z)) |
| Acetylcholine synthesis | wiki | WP528 | hmdb:HMDB0008508 | PC(20:5(5Z,8Z,11Z,14Z,17Z)/20:3(8Z,11Z,14Z)),PC(14:0_20:0) |
| Phospholipid Biosynthesis | kegg | map00564 | hmdb:HMDB0008508 | PC(20:5(5Z,8Z,11Z,14Z,17Z)/20:3(8Z,11Z,14Z)),PC(14:0_20:0) |
| Acetylcholine synthesis | wiki | WP528 | hmdb:HMDB0008509 | PC(20:5(5Z,8Z,11Z,14Z,17Z)/20:4(5Z,8Z,11Z,14Z)),PC(14:0_20:0) |
| Phospholipid Biosynthesis | kegg | map00564 | hmdb:HMDB0008509 | PC(20:5(5Z,8Z,11Z,14Z,17Z)/20:4(5Z,8Z,11Z,14Z)),PC(14:0_20:0) |
| Acetylcholine synthesis | wiki | WP528 | hmdb:HMDB0008510 | PC(20:5(5Z,8Z,11Z,14Z,17Z)/20:4(8Z,11Z,14Z,17Z)) |
| Phospholipid Biosynthesis | kegg | map00564 | hmdb:HMDB0008510 | PC(20:5(5Z,8Z,11Z,14Z,17Z)/20:4(8Z,11Z,14Z,17Z)) |
| Acetylcholine synthesis | wiki | WP528 | hmdb:HMDB0008515 | PC(20:5(5Z,8Z,11Z,14Z,17Z)/22:4(7Z,10Z,13Z,16Z)),PC(14:0_20:0) |
| Phospholipid Biosynthesis | kegg | map00564 | hmdb:HMDB0008515 | PC(20:5(5Z,8Z,11Z,14Z,17Z)/22:4(7Z,10Z,13Z,16Z)),PC(14:0_20:0) |

| | | | | |
|---|---|---|---|---|
| Acetylcholine synthesis | wiki | WP528 | hmdb:HMDB0008537 | PC(22:0/20:0),PC(14:0_20:0) |
| Phospholipid Biosynthesis | kegg | map00564 | hmdb:HMDB0008537 | PC(22:0/20:0),PC(14:0_20:0) |
| Acetylcholine synthesis | wiki | WP528 | hmdb:HMDB0008609 | PC(22:2(13Z,16Z)/20:5(5Z,8Z,11Z,14Z,17Z)),PC(14:0_20:0) |
| Phospholipid Biosynthesis | kegg | map00564 | hmdb:HMDB0008609 | PC(22:2(13Z,16Z)/20:5(5Z,8Z,11Z,14Z,17Z)),PC(14:0_20:0) |
| Acetylcholine synthesis | wiki | WP528 | hmdb:HMDB0008658 | PC(22:5(4Z,7Z,10Z,13Z,16Z)/15:0) |
| Phospholipid Biosynthesis | kegg | map00564 | hmdb:HMDB0008658 | PC(22:5(4Z,7Z,10Z,13Z,16Z)/15:0) |
| Acetylcholine synthesis | wiki | WP528 | hmdb:HMDB0008670 | PC(22:5(4Z,7Z,10Z,13Z,16Z)/20:2(11Z,14Z)) |
| Phospholipid Biosynthesis | kegg | map00564 | hmdb:HMDB0008670 | PC(22:5(4Z,7Z,10Z,13Z,16Z)/20:2(11Z,14Z)) |
| Acetylcholine synthesis | wiki | WP528 | hmdb:HMDB0008674 | PC(22:5(4Z,7Z,10Z,13Z,16Z)/20:4(8Z,11Z,14Z,17Z)) |
| Phospholipid Biosynthesis | kegg | map00564 | hmdb:HMDB0008674 | PC(22:5(4Z,7Z,10Z,13Z,16Z)/20:4(8Z,11Z,14Z,17Z)) |
| Acetylcholine synthesis | wiki | WP528 | hmdb:HMDB0008691 | PC(22:5(7Z,10Z,13Z,16Z,19Z)/15:0) |
| Phospholipid Biosynthesis | kegg | map00564 | hmdb:HMDB0008691 | PC(22:5(7Z,10Z,13Z,16Z,19Z)/15:0) |
| Acetylcholine synthesis | wiki | WP528 | hmdb:HMDB0008698 | PC(22:5(7Z,10Z,13Z,16Z,19Z)/18:3(6Z,9Z,12Z)) |
| Phospholipid Biosynthesis | kegg | map00564 | hmdb:HMDB0008698 | PC(22:5(7Z,10Z,13Z,16Z,19Z)/18:3(6Z,9Z,12Z)) |
| Acetylcholine synthesis | wiki | WP528 | hmdb:HMDB0008699 | PC(22:5(7Z,10Z,13Z,16Z,19Z)/18:3(9Z,12Z,15Z)) |
| Phospholipid Biosynthesis | kegg | map00564 | hmdb:HMDB0008699 | PC(22:5(7Z,10Z,13Z,16Z,19Z)/18:3(9Z,12Z,15Z)) |
| Acetylcholine synthesis | wiki | WP528 | hmdb:HMDB0008700 | PC(22:5(7Z,10Z,13Z,16Z,19Z)/18:4(6Z,9Z,12Z,15Z)) |
| Phospholipid Biosynthesis | kegg | map00564 | hmdb:HMDB0008700 | PC(22:5(7Z,10Z,13Z,16Z,19Z)/18:4(6Z,9Z,12Z,15Z)) |
| Acetylcholine synthesis | wiki | WP528 | hmdb:HMDB0008703 | PC(22:5(7Z,10Z,13Z,16Z,19Z)/20:2(11Z,14Z)) |
| Phospholipid Biosynthesis | kegg | map00564 | hmdb:HMDB0008703 | PC(22:5(7Z,10Z,13Z,16Z,19Z)/20:2(11Z,14Z)) |
| Acetylcholine synthesis | wiki | WP528 | hmdb:HMDB0008707 | PC(22:5(7Z,10Z,13Z,16Z,19Z)/20:4(8Z,11Z,14Z,17Z)) |
| Phospholipid Biosynthesis | kegg | map00564 | hmdb:HMDB0008707 | PC(22:5(7Z,10Z,13Z,16Z,19Z)/20:4(8Z,11Z,14Z,17Z)) |
| Acetylcholine synthesis | wiki | WP528 | hmdb:HMDB0008730 | PC(22:6(4Z,7Z,10Z,13Z,16Z,19Z)/18:2(9Z,12Z)),PC(14:0_20:0) |
| Phospholipid Biosynthesis | kegg | map00564 | hmdb:HMDB0008730 | PC(22:6(4Z,7Z,10Z,13Z,16Z,19Z)/18:2(9Z,12Z)),PC(14:0_20:0) |
| Acetylcholine synthesis | wiki | WP528 | hmdb:HMDB0008732 | PC(22:6(4Z,7Z,10Z,13Z,16Z,19Z)/18:3(9Z,12Z,15Z)),PC(14:0_20:0) |
| Phospholipid Biosynthesis | kegg | map00564 | hmdb:HMDB0008732 | PC(22:6(4Z,7Z,10Z,13Z,16Z,19Z)/18:3(9Z,12Z,15Z)),PC(14:0_20:0) |
| Acetylcholine synthesis | wiki | WP528 | hmdb:HMDB0008737 | PC(22:6(4Z,7Z,10Z,13Z,16Z,19Z)/20:3(5Z,8Z,11Z)) |
| Phospholipid Biosynthesis | kegg | map00564 | hmdb:HMDB0008737 | PC(22:6(4Z,7Z,10Z,13Z,16Z,19Z)/20:3(5Z,8Z,11Z)) |
| Acetylcholine synthesis | wiki | WP528 | hmdb:HMDB0008738 | PC(22:6(4Z,7Z,10Z,13Z,16Z,19Z)/20:3(8Z,11Z,14Z)),PC(14:0_20:0) |
| Phospholipid Biosynthesis | kegg | map00564 | hmdb:HMDB0008738 | PC(22:6(4Z,7Z,10Z,13Z,16Z,19Z)/20:3(8Z,11Z,14Z)),PC(14:0_20:0) |
| Acetylcholine synthesis | wiki | WP528 | hmdb:HMDB0008748 | PC(22:6(4Z,7Z,10Z,13Z,16Z,19Z)/22:6(4Z,7Z,10Z,13Z,16Z,19Z)),PC(14:0_20:0) |
| Phospholipid Biosynthesis | kegg | map00564 | hmdb:HMDB0008748 | PC(22:6(4Z,7Z,10Z,13Z,16Z,19Z)/22:6(4Z,7Z,10Z,13Z,16Z,19Z)),PC(14:0_20:0) |
| Acetylcholine synthesis | wiki | WP528 | hmdb:HMDB0008761 | PC(24:0/18:1(11Z)),PC(14:0_20:0) |
| Phospholipid Biosynthesis | kegg | map00564 | hmdb:HMDB0008761 | PC(24:0/18:1(11Z)),PC(14:0_20:0) |
| Acetylcholine synthesis | wiki | WP528 | hmdb:HMDB0008762 | PC(24:0/18:1(9Z)),PC(14:0_20:0) |
| Phospholipid Biosynthesis | kegg | map00564 | hmdb:HMDB0008762 | PC(24:0/18:1(9Z)),PC(14:0_20:0) |
| Retinol Metabolism | kegg | map00830 | hmdb:HMDB0008790 | PC(24:1(15Z)/15:0) |
| Beta-oxidation of pristanoyl-CoA | reactome | R-HSA-389887 | hmdb:HMDB0000824 | Propionylcarnitine,PROPIONYLCARNITINE (C3),O-Propanoylcarnitine |
| Krebs cycle disorders | wiki | WP4236 | hmdb:HMDB0000824 | Propionylcarnitine,PROPIONYLCARNITINE (C3),O-Propanoylcarnitine |
| Leucine, isoleucine and valine metabolism | wiki | WP4686 | hmdb:HMDB0000824 | Propionylcarnitine,PROPIONYLCARNITINE (C3),O-Propanoylcarnitine |
| Peroxisomal lipid metabolism | reactome | R-HSA-390918 | hmdb:HMDB0000824 | Propionylcarnitine,PROPIONYLCARNITINE (C3),O-Propanoylcarnitine |
| Peroxisomal lipid metabolism | wiki | WP1878 | hmdb:HMDB0000824 | Propionylcarnitine,PROPIONYLCARNITINE (C3),O-Propanoylcarnitine |
| Folate metabolism | wiki | WP176 | uniprot:P01011 | SERPINA3 |
| Platelet degranulation | reactome | R-HSA-114608 | uniprot:P01011 | SERPINA3 |
| Response to elevated platelet cytosolic Ca ions | wiki | WP1903 | uniprot:P01011 | SERPINA3 |
| Response to elevated platelet cytosolic Ca2+ | reactome | R-HSA-76005 | uniprot:P01011 | SERPINA3 |
| Vitamin B12 metabolism | wiki | WP1533 | uniprot:P01011 | SERPINA3 |
| Glucocorticoid biosynthesis | reactome | R-HSA-194002 | uniprot:P08185 | SERPINA6 |
| Metabolism of steroid hormones | reactome | R-HSA-196071 | uniprot:P08185 | SERPINA6 |
| Metabolism of steroid hormones | wiki | WP2749 | uniprot:P08185 | SERPINA6 |
| Prednisone ADME | reactome | R-HSA-9757110 | uniprot:P08185 | SERPINA6 |
| Common Pathway of Fibrin Clot Formation | reactome | R-HSA-140875 | uniprot:P01008 | SERPINC1 |
| Complement and coagulation cascades | wiki | WP558 | uniprot:P01008 | SERPINC1 |
| Formation of Fibrin Clot (Clotting Cascade) | reactome | R-HSA-140877 | uniprot:P01008 | SERPINC1 |
| Formation of Fibrin Clot and the Clotting Cascade | wiki | WP1818 | uniprot:P01008 | SERPINC1 |
| Intrinsic Pathway of Fibrin Clot Formation | reactome | R-HSA-140837 | uniprot:P01008 | SERPINC1 |
| Post-translational protein phosphorylation | reactome | R-HSA-8957275 | uniprot:P01008 | SERPINC1 |
| Post-translational protein phosphorylation | wiki | WP4110 | uniprot:P01008 | SERPINC1 |
| Regulation of IGF transport and uptake by IGF binding proteins | wiki | WP2799 | uniprot:P01008 | SERPINC1 |

| | | | | |
|---|---|---|---|---|
| Regulation of Insulin-like Growth Factor (IGF) transport and uptake by Insulin-like Growth Factor Binding Proteins (IGFBPs) | reactome | R-HSA-381426 | uniprot:P01008 | SERPINC1 |
| Common Pathway of Fibrin Clot Formation | reactome | R-HSA-140875 | uniprot:P05546 | SERPIND1 |
| Complement and coagulation cascades | wiki | WP558 | uniprot:P05546 | SERPIND1 |
| Formation of Fibrin Clot (Clotting Cascade) | reactome | R-HSA-140877 | uniprot:P05546 | SERPIND1 |
| Formation of Fibrin Clot and the Clotting Cascade | wiki | WP1818 | uniprot:P05546 | SERPIND1 |
| Intrinsic Pathway of Fibrin Clot Formation | reactome | R-HSA-140837 | uniprot:P05546 | SERPIND1 |
| Post-translational protein phosphorylation | reactome | R-HSA-8957275 | uniprot:P05546 | SERPIND1 |
| Post-translational protein phosphorylation | wiki | WP4110 | uniprot:P05546 | SERPIND1 |
| Regulation of IGF transport and uptake by IGF binding proteins | wiki | WP2799 | uniprot:P05546 | SERPIND1 |
| Regulation of Insulin-like Growth Factor (IGF) transport and uptake by Insulin-like Growth Factor Binding Proteins (IGFBPs) | reactome | R-HSA-381426 | uniprot:P05546 | SERPIND1 |
| ABC transporters in lipid homeostasis | reactome | R-HSA-1369062 | hmdb:HMDB0013464 | SM(d18:0/16:1(9Z)) |
| ABC-family proteins mediated transport | reactome | R-HSA-382556 | hmdb:HMDB0013464 | SM(d18:0/16:1(9Z)) |
| ABC-family proteins mediated transport | wiki | WP1780 | hmdb:HMDB0013464 | SM(d18:0/16:1(9Z)) |
| Ceramide signalling | reactome | R-HSA-193681 | hmdb:HMDB0013464 | SM(d18:0/16:1(9Z)) |
| Glycosphingolipid catabolism | reactome | R-HSA-9840310 | hmdb:HMDB0013464 | SM(d18:0/16:1(9Z)) |
| Glycosphingolipid metabolism | reactome | R-HSA-1660662 | hmdb:HMDB0013464 | SM(d18:0/16:1(9Z)) |
| Neurodegeneration with brain iron accumulation (NBIA) subtypes pathway | wiki | WP4577 | hmdb:HMDB0013464 | SM(d18:0/16:1(9Z)) |
| Oligodendrocyte specification and differentiation, leading to myelin components for CNS | wiki | WP4304 | hmdb:HMDB0013464 | SM(d18:0/16:1(9Z)) |
| p75 NTR receptor-mediated signaling | wiki | WP4443 | hmdb:HMDB0013464 | SM(d18:0/16:1(9Z)) |
| p75 NTR receptor-mediated signalling | reactome | R-HSA-193704 | hmdb:HMDB0013464 | SM(d18:0/16:1(9Z)) |
| Phosphatidylcholine catabolism | wiki | WP4195 | hmdb:HMDB0013464 | SM(d18:0/16:1(9Z)) |
| Sphingolipid metabolism: integrated pathway | wiki | WP4726 | hmdb:HMDB0013464 | SM(d18:0/16:1(9Z)) |
| ABC transporters in lipid homeostasis | reactome | R-HSA-1369062 | hmdb:HMDB0013467 | SM(d18:0/22:2(13Z,16Z)(OH)) |
| ABC-family proteins mediated transport | reactome | R-HSA-382556 | hmdb:HMDB0013467 | SM(d18:0/22:2(13Z,16Z)(OH)) |
| ABC-family proteins mediated transport | wiki | WP1780 | hmdb:HMDB0013467 | SM(d18:0/22:2(13Z,16Z)(OH)) |
| Ceramide signalling | reactome | R-HSA-193681 | hmdb:HMDB0013467 | SM(d18:0/22:2(13Z,16Z)(OH)) |
| Glycosphingolipid catabolism | reactome | R-HSA-9840310 | hmdb:HMDB0013467 | SM(d18:0/22:2(13Z,16Z)(OH)) |
| Glycosphingolipid metabolism | reactome | R-HSA-1660662 | hmdb:HMDB0013467 | SM(d18:0/22:2(13Z,16Z)(OH)) |
| Neurodegeneration with brain iron accumulation (NBIA) subtypes pathway | wiki | WP4577 | hmdb:HMDB0013467 | SM(d18:0/22:2(13Z,16Z)(OH)) |
| Oligodendrocyte specification and differentiation, leading to myelin components for CNS | wiki | WP4304 | hmdb:HMDB0013467 | SM(d18:0/22:2(13Z,16Z)(OH)) |
| p75 NTR receptor-mediated signaling | wiki | WP4443 | hmdb:HMDB0013467 | SM(d18:0/22:2(13Z,16Z)(OH)) |
| p75 NTR receptor-mediated signalling | reactome | R-HSA-193704 | hmdb:HMDB0013467 | SM(d18:0/22:2(13Z,16Z)(OH)) |
| Phosphatidylcholine catabolism | wiki | WP4195 | hmdb:HMDB0013467 | SM(d18:0/22:2(13Z,16Z)(OH)) |
| Sphingolipid metabolism: integrated pathway | wiki | WP4726 | hmdb:HMDB0013467 | SM(d18:0/22:2(13Z,16Z)(OH)) |
| Sphingolipid metabolism: integrated pathway | wiki | WP4726 | hmdb:HMDB0010169 | SM(d18:1/16:0) |
| ABC transporters in lipid homeostasis | reactome | R-HSA-1369062 | hmdb:HMDB0012107 | SM(d18:1/24:1(15Z)),SPHM,N-lauroyl-D-erythro-Sphingosylphosphoryl choline |
| ABC-family proteins mediated transport | reactome | R-HSA-382556 | hmdb:HMDB0012107 | SM(d18:1/24:1(15Z)),SPHM,N-lauroyl-D-erythro-Sphingosylphosphoryl choline |
| ABC-family proteins mediated transport | wiki | WP1780 | hmdb:HMDB0012107 | SM(d18:1/24:1(15Z)),SPHM,N-lauroyl-D-erythro-Sphingosylphosphoryl choline |
| Ceramide signalling | reactome | R-HSA-193681 | hmdb:HMDB0012107 | SM(d18:1/24:1(15Z)),SPHM,N-lauroyl-D-erythro-Sphingosylphosphoryl choline |
| Glycosphingolipid catabolism | reactome | R-HSA-9840310 | hmdb:HMDB0012107 | SM(d18:1/24:1(15Z)),SPHM,N-lauroyl-D-erythro-Sphingosylphosphoryl choline |
| Glycosphingolipid metabolism | reactome | R-HSA-1660662 | hmdb:HMDB0012107 | SM(d18:1/24:1(15Z)),SPHM,N-lauroyl-D-erythro-Sphingosylphosphoryl choline |
| Neurodegeneration with brain iron accumulation (NBIA) subtypes pathway | wiki | WP4577 | hmdb:HMDB0012107 | SM(d18:1/24:1(15Z)),SPHM,N-lauroyl-D-erythro-Sphingosylphosphoryl choline |
| Oligodendrocyte specification and differentiation, leading to myelin components for CNS | wiki | WP4304 | hmdb:HMDB0012107 | SM(d18:1/24:1(15Z)),SPHM,N-lauroyl-D-erythro-Sphingosylphosphoryl choline |
| p75 NTR receptor-mediated signaling | wiki | WP4443 | hmdb:HMDB0012107 | SM(d18:1/24:1(15Z)),SPHM,N-lauroyl-D-erythro-Sphingosylphosphoryl choline |
| p75 NTR receptor-mediated signalling | reactome | R-HSA-193704 | hmdb:HMDB0012107 | SM(d18:1/24:1(15Z)),SPHM,N-lauroyl-D-erythro-Sphingosylphosphoryl choline |
| Phosphatidylcholine catabolism | wiki | WP4195 | hmdb:HMDB0012107 | SM(d18:1/24:1(15Z)),SPHM,N-lauroyl-D-erythro-Sphingosylphosphoryl choline |
| Sphingolipid metabolism: integrated pathway | wiki | WP4726 | hmdb:HMDB0012107 | SM(d18:1/24:1(15Z)),SPHM,N-lauroyl-D-erythro-Sphingosylphosphoryl choline |
| Extra-nuclear estrogen signaling | wiki | WP4977 | hmdb:HMDB0003334 | Symmetric dimethylarginine,ADMA |
| Metabolism of nitric oxide: NOS3 activation and regulation | wiki | WP1850 | hmdb:HMDB0003334 | Symmetric dimethylarginine,ADMA |
| Nitric oxide metabolism in cystic fibrosis | wiki | WP4947 | hmdb:HMDB0003334 | Symmetric dimethylarginine,ADMA |
| Signaling by VEGF | wiki | WP1919 | hmdb:HMDB0003334 | Symmetric dimethylarginine,ADMA |
| 1p36 copy number variation syndrome | wiki | WP5345 | uniprot:P04004 | VTN |
| Complement system | wiki | WP2806 | uniprot:P04004 | VTN |
| Complement system in neuronal development and plasticity | wiki | WP5090 | uniprot:P04004 | VTN |
| ECM proteoglycans | reactome | R-HSA-3000178 | uniprot:P04004 | VTN |
| Elastic fibre formation | reactome | R-HSA-1566948 | uniprot:P04004 | VTN |
| Elastic fibre formation | wiki | WP2666 | uniprot:P04004 | VTN |
| Extracellular matrix organization | wiki | WP2703 | uniprot:P04004 | VTN |
| Inflammatory response pathway | wiki | WP453 | uniprot:P04004 | VTN |

| | | | | |
|---|---|---|---|---|
| Integrin cell surface interactions | reactome | R-HSA-216083 | uniprot:P04004 | VTN |
| Molecules associated with elastic fibres | reactome | R-HSA-2129379 | uniprot:P04004 | VTN |
| Non-integrin membrane-ECM interactions | reactome | R-HSA-3000171 | uniprot:P04004 | VTN |
| Primary focal segmental glomerulosclerosis (FSGS) | wiki | WP2572 | uniprot:P04004 | VTN |
| Regulation of Complement cascade | reactome | R-HSA-977606 | uniprot:P04004 | VTN |
| Syndecan interactions | reactome | R-HSA-3000170 | uniprot:P04004 | VTN |
| Syndecan interactions | wiki | WP2787 | uniprot:P04004 | VTN |
