## supplementary_information1 for "Molecular characterization of liver steatosis through circulating proteins, metabolites and lipids" (from another version of this preprint: version 1, 2025-12-02)

**Title**

**Short Title**

Blood-based profiling of liver steatosis

**Authors**

Marilyn De Graeve[1], Essi Hantikainen[1], Philippine Louail[1], Vinicius Verri Hernandes[1,2], Cristian Pattaro[1], Francisco S. Domingues[1], Peter P. Pramstaller[1], Herbert Tilg[3], Markus Ralser[4,5,6], Christoph Grander[3,*], Johannes Rainer[1,*]

**Affiliations**

[1]Institute for Biomedicine, Eurac Research, 39100 Bolzano, Italy

[2]Faculty of Chemistry, Department of Food Chemistry and Toxicology, University of Vienna, Währinger Straße 38, 1090 Vienna, Austria

[3]Department of Internal Medicine I, Gastroenterology, Hepatology, Endocrinology & Metabolism, Medical University Innsbruck, 6020 Innsbruck, Austria.

[4]Institute of Biochemistry, Charité Universitätsmedizin Berlin, 10117 Berlin, Germany

[5]Max Planck Institute for Molecular Genetics, 14195 Berlin, Germany

[6]The Wellcome Centre for Human Genetics, Nuffield Department of Medicine, University of Oxford, Oxford OX3 7BN, UK

* Contributed equally

**Corresponding author**

### SI information 1: Investigation of steatosis related liver fibrosis and MASH

#### Classification of steatosis related liver parameters

Hepatic fibrosis was assessed using liver stiffness measurement (FibroScan®, Echosens, France): scores below 7 kPa were considered normal, those between ≥7 and ≤12.5 kPa indicated mild fibrosis, and those above 12.5 kPa indicated severe fibrosis.[1] In addition, we used FIB4 scores to estimate liver fibrosis by using participant's age and blood markers[2] (ALT, AST and Platelet count, see Table 1).

To investigate the presence of metabolic dysfunction-associated steatohepatitis (MASH), the non-invasive FibroScan-AST (FAST) score was calculated using the above-mentioned CAP and liver stiffness values, together with aspartate aminotransferase[3,4] (AST, in U/L). Participants were classified at-risk of MASH if they presented a FAST score above 0.67 and not at-risk of MASH when they had a FAST score below 0.35. FAST scores between ≥0.35 and ≤0.67 were defined as inconclusive.

Other relevant measurements available for CHRIS NAFLD sub-study participants used to investigate initial correlations with steatosis were visual fat, MASLD and MetS. Visual fat was derived from sonographic evaluation of the liver fat by hepatologist (categorical from 1 to 3, with 2-3 depicting steatosis).

MASLD was defined by (i) presence of steatotic liver disease using visual fat score of 2-3, (ii) positive for at least one cardiometabolic criteria (BMI≥25 or waist circumference (WC>94 (Males) WC>80 (Females) cm); fasting glucose≥100 mg/dl or HbA1c≥5.7% or T2D or following treatment for type 2 diabetes; high blood pressure (BP≥130/85 mmHg) or following specific antihypertensive drug treatment; triglycerides≥150 mg/dl or following lipid lowering treatment; and low high-density lipoprotein (HDL≤40 (M) HDL≤50 (F) mg/dl) or following lipid lowering treatment); but (iii) having a low alcohol consumption (below 40g/day).

MetS was defined by individuals with a high waist circumference (WC≥94 (M), WC≥80 (F) cm), and at least two of the following traits: triglycerides≥150 mg/dl or following specific treatment for this lipid abnormality; low high-density lipoprotein (HDL<40 (M) HDL<50 (F) mg/dl) or following specific treatment for this lipid abnormality; blood pressure (BP≥ 130/85 mmHg) or following treatment for hypertension; and fasting glucose≥100 mg/dl or diagnosed T2D.

#### Multiple linear regression analysis

Similarly to the liver steatosis analysis (Section: Statistical data analysis), an additional hypothesis regarding liver steatosis and MASH liver was tested. For this, multiple linear regression models were fitted to identify associations between proteins and metabolites and lipids in healthy controls, i.e. negative for fibrosis (<7 kPa, n=297) and severe fibrosis individuals (>12.5 kPa, n=4).

#### Circulating proteins, metabolites and lipids associated with liver fibrosis or MASH

As higher levels of steatosis are associated with increased risk for MASH and development of fibrosis[5], participants were evaluated for presence of MASH and fibrosis, respectively, and multiple linear regression analyses were performed for these phenotypes to reveal circulating proteins, metabolites and lipids associated with liver fibrosis or MASH.

Fibroscan was used for non-invasive measurement of liver stiffness and quantification of liver fibrosis. Participants with liver stiffness values above 12.5 kPa were classified as severe fibrosis (cirrhosis)[6]. The participants were mostly categorized in the group having no fibrosis, the gender was balanced, and no relationship between age and fibrosis classifications was observed. Participants from the highest fibrosis group (n=4, >12.5 kPa), displayed signs of liver injury depicted by a decreased platelet count, and increased AST and ALT concentration levels (Table S7). Notably, the FIB4 scores, calculated from the participant's age and the blood clinical markers ALT, AST and Platelet count, did not correlate well with the participant's fibrosis classifications (liver stiffness measurements using Fibroscan) (Figure S5). This can most likely be due to a small number of participants with a high liver stiffness value over 12.5 kPa (n=4), thus afflicted with cirrhosis.

Table S7. Demographics of fibrosis classification based upon the liver stiffness measurements.

| **Relevant Metadata** | **Unit** | **No Fibrosis** | **Mild fibrosis** | **Severe fibrosis** |
|---|---|---|---|---|
| Sex | Nr. male | 146 | 11 | 2 |
| | Nr. female | 153 | 10 | 4 |
| Age | years | 67.82 ± 10.39 | 63.33 ± 11.75 | 66.67 ± 11.79 |
| AST | U/L | 20.22 ± 5.31 | 25.19 ± 10.21 | 29.00 ± 13.58 |
| ALT | U/L | 20.45 ± 10.73 | 29.57 ± 17.18 | 35.83 ± 27.32 |
| Platelet Count | 10E9/L | 240.77 ± 61.60 | 217.62 ± 48.35 | 210.33 ± 59.41 |

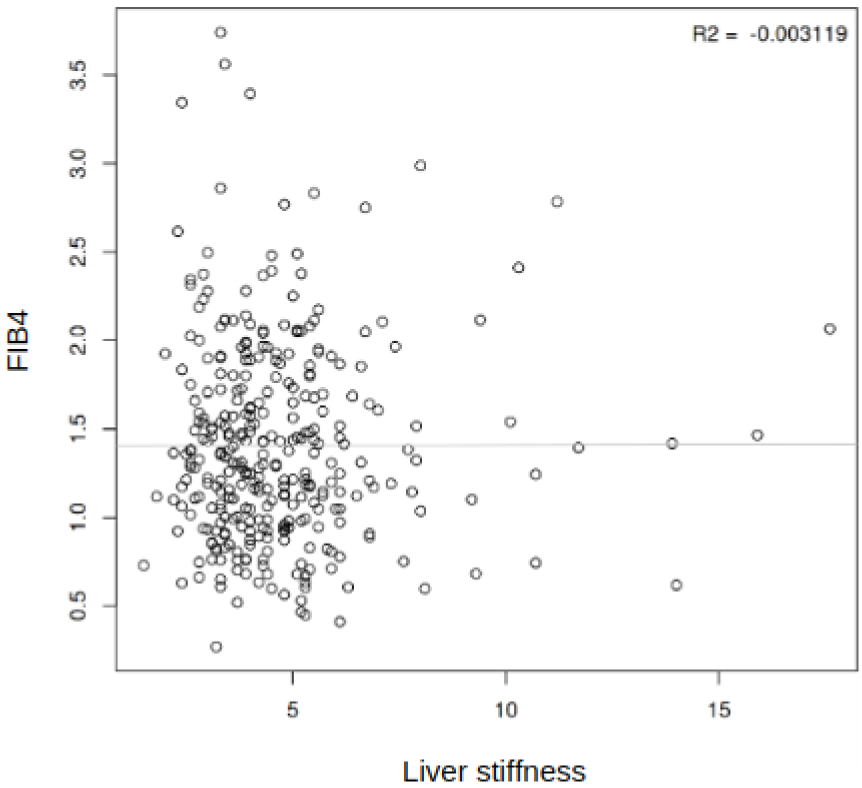


Figure S5. Participant's fibrosis score (liver stiffness measurements) versus their calculated FIB4 score depicts weak correlation.

Comparing control individuals (n=297, liver stiffness<7 kPa) versus individuals with severe fibrosis (n=4, liver stiffness >12.5 kPa), 2 significant proteins were retrieved (Table S8). In our cohort, we found CLEC3B to be significantly decreased in individuals with advanced fibrosis. Plasminogen-binding tetranectin (CLEC3B) was described to be a biomarker for hepatocellular[7] and for cholangiocarcinoma[8]. Vitamin D-binding protein (GC) was also found to be decreased in participants with increased liver fibrosis compared to controls. The very low number of cases precludes, however, a generalizable interpretation of this result. Also, no metabolites or lipids were found significantly associated with fibrosis.

Table S8. Significant proteins when comparing control (n=297, liver stiffness<7 kPa) versus severe fibrosis (n=4, liver stiffness >12.5 kPa) using linear regression analysis with age and sex as covariates. $CV_{QC}$, coefficient of variance of pooled quality control samples of CHRIS study. Alternative nomenclature can be consulted on UniProt (https://www.uniprot.org/). Formula: protein ~ sex + age/10 + steatosis grade.

| **Protein** | **Gene** | **$CV_{QC}$** | **Coefficient** | **Adjusted p-value** | **Effect Size** |
|---|---|---|---|---|---|
| P02774;P02774-3 | GC | 0.0613 | -0. 3534 | 0.00201 | -2.066 |

| | | | | | |
|---|---|---|---|---|---|
| P05452 | CLEC3B | 0.1391 | -0.4796 | 0.03965 | -1.688 |

Using the liver fat, liver stiffness and AST values, the at-risk for MASH was calculated using the FAST score. The participants were mostly categorized in the group being not at-risk MASH (n=300). The remaining participants were inconclusive, contained mostly missing values (n=13) precluding calculation of the FAST. Only a single individual was classified as at-risk MASH and hence no association analyses were performed.
