## supplementary_information2 for "Molecular characterization of liver steatosis through circulating proteins, metabolites and lipids" (from another version of this preprint: version 1, 2025-12-02)

**Title**

**Short Title**

Blood-based profiling of liver steatosis

**Authors**

Marilyn De Graeve[1], Essi Hantikainen[1], Philippine Louail[1], Vinicius Verri Hernandes[1,2], Cristian Pattaro[1], Francisco S. Domingues[1], Peter P. Pramstaller[1], Herbert Tilg[3], Markus Ralser[4,5,6], Christoph Grander[3,*], Johannes Rainer[1,*]

**Affiliations**

[1]Institute for Biomedicine, Eurac Research, 39100 Bolzano, Italy

[2]Faculty of Chemistry, Department of Food Chemistry and Toxicology, University of Vienna, Währinger Straße 38, 1090 Vienna, Austria

[3]Department of Internal Medicine I, Gastroenterology, Hepatology, Endocrinology & Metabolism, Medical University Innsbruck, 6020 Innsbruck, Austria.

[4]Institute of Biochemistry, Charité Universitätsmedizin Berlin, 10117 Berlin, Germany

[5]Max Planck Institute for Molecular Genetics, 14195 Berlin, Germany

[6]The Wellcome Centre for Human Genetics, Nuffield Department of Medicine, University of Oxford, Oxford OX3 7BN, UK

* Contributed equally

**Corresponding author**

### SI information 2: Untargeted metabolomics analysis of grade 3 steatosis participants

#### Sample preparation for untargeted metabolomics

For untargeted metabolomics, 50 µL of blood serum samples as well as pooled QC samples were defrosted in ice and dispensed onto an Ostro plate (Waters) for protein precipitation and phospholipid removal, followed by the addition of 200 µL of cold acetonitrile + 1% formic acid solution containing lab eled internal standards. The plate was shaken for 5 min at 400 rpm and incubated for 30 min at 4°C for enhanced protein precipitation. Next, a positive pressure of 12 psi was applied for 7 min followed by an increase to 25 psi, kept for an additional 5 min for ensuring complete sample filtering. Samples were dried in a concentrator for 2h at 38°C and additional 30 min at 40°C. After 5 min cooling at room temperature, 200 µL of acetonitrile/water 50:50 v/v solution was added for reconstitution. The plate was then shaken for 5 min at 400 rpm and incubated for 25 min at room temperature. Finally, an additional protein precipitation step via filtration using Sirocco filters (Waters) was performed using the same pressure and time previously described for the Ostro plate. The final extract was split into one additional plate for LC-MS data acquisition in both positive and negative ionization modes.

#### Untargeted metabolomics data acquisition

Untargeted metabolomics data was obtained using an Agilent 1290 Infinity LC system coupled with a Sciex 5600 QTOF mass spectrometer. An ACQUITY UPLC BEH Amide Column (130Å, 1.7 µm, 2.1 mm X 100 mm, Waters Corporation) was used for compound separation, with a total injection volume of 10 µL. Acetonitrile (A) and water (B) were used as mobile phase, both with 0.1% formic acid as additive and a flow rate of 0.6 mL/min. Chromatographic gradient was set as follows: 0 – 1 min (95% A), 1 – 4 min (95 to 30% A), 4 – 5 min (30% A), 5 – 5.1 min (30 to 95% A), 5.10 – 8 min (95% A). Compound ionization was performed using an electrospray source set at 5120V (positive mode) or -3800V (negative mode). Further source parameters included source temperature (700°C for positive mode, 650°C for negative mode), ion source gas1 (60psi for positive mode and 30psi for negative mode) and ion source gas 2 (60psi for positive mode and 30psi for negative mode). Curtain gas (25psi) was set the same for both polarities. Spectra were acquired in profile mode under the 50-1000 m/z range.

#### Untargeted metabolomics data processing

For untargeted metabolomics analysis, raw data were preprocessed using the publicly available xcms software package.[1] A detailed description of the preprocessing workflow is available at: https://github.com/EuracBiomedicalResearch/nafld_proteo_metabolomics. Samples were analyzed in seven batches, and pooled quality control (QC) samples were included in each batch. Initial inspection of the raw data revealed substantial retention time shifts across certain regions between batches. Retention time correction was therefore performed using QC samples, with spiked internal standards serving as anchor peaks. Features were defined as valid if they were detected in at least 30% of the experimental samples. Signals observed in blank samples were removed as potential contaminants. Subsequently, batch effects were corrected using a combined linear model that accounted for both injection order and batch.

### Grade 3 steatosis associated blood serum untargeted metabolome

Multiple linear modeling (Figure S10, Table S9) was calculated to identify associations between metabolic features in healthy controls, i.e. negative for steatosis (none, n=140) and individuals with grade 3 steatosis (n=81), as described for the targeted metabolomics (Section: Multiple linear regression analysis). Prior statistical analysis, the data was median scaled, as the exploratory unsupervised analyses revealed considerable batch effects, and log2 transformed.

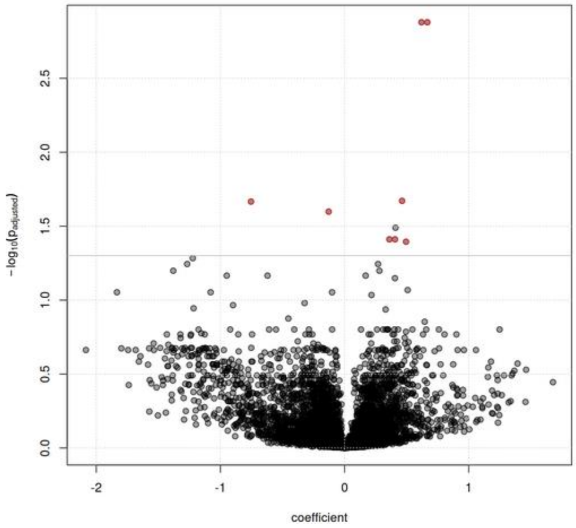


Figure S10. Volcano plot for normal versus grade 3 steatosis, after linear modeling of the untargeted acquired but unidentified metabolic features, from the untargeted metabolomics. Red colored points indicate significance (FDR adjusted p-value<0.05).

Table S9. Significant metabolic features, acquired using untargeted metabolomics, when comparing control (n=141, CAP<238 dB/m) versus grade 3 steatosis (n=79, naab13>290 dB/m) using linear regression analysis with age and sex as covariates. $CV_{QC}$, coefficient of variance of pooled quality control samples of CHRIS study. *m/z,* mass to charge ratio (Da). RT, retention time (sec). 80 kit). Formula: metabolic feature ~ sex + age/10 + steatosis grade.

| | ***m/z*** | **RT** | **$CV_{QC}$** | **Coefficient** | **Adjusted p-value** | **Effect Size** |
|---|---|---|---|---|---|---|
| FT00689 | 131.1 | 169.4 | 0.2572 | 0.6664 | 0.001321 | 0.711 |
| FT00940 | 147.6 | 169.3 | 0.2453 | 0.6192 | 0.001321 | 0.7051 |
| FT03857 | 329.1 | 148.7 | 0.3063 | 0.4632 | 0.02132 | 0.6063 |
| FT01935 | 214.1 | 207 | 0.7897 | -0.7542 | 0.02157 | -0.6002 |
| FT00688 | 131.1 | 175.8 | 0.1374 | -0.1285 | 0.02521 | -0.6075 |
| FT00949 | 148.1 | 169.3 | 0.2163 | 0.361 | 0.03882 | 0.5832 |
| FT00968 | 149.1 | 169.3 | 0.4662 | 0.4062 | 0.03882 | 0.5869 |

| | | | | | | |
|---|---|---|---|---|---|---|
| FT00181 | 84.05 | 169.4 | 0.3353 | 0.4948 | 0.04028 | 0.5797 |

### Altered pathways involving steatosis grade 3 associated circulating metabolites, derived from the untargeted metabolomics

The results (molecular features together with their associated p-values) from the linear modeling were used to enable a richer pathway enrichment analysis, utilizing the Mummichog algorithm (http://mummichog.org, v2.7.0) using metabolic model MFN_1.10.4.[2] All metabolic features from the untargeted metabolomics were included, even though they were not significant or annotated. Using the power of mummichog, pathways were revealed by finding patterns of up- or downregulated small molecules, hence identifying their mutual response in a certain biochemical pathway.

The analysis revealed 14 enriched pathways containing the putative annotations of 3 significantly discriminating metabolites: N-Acetylmethionine, 1-Pyrroline-4-hydroxy-2-carboxylate and L-4-Hydroxyglutamate semialdehyde (Table S10). As these molecules are not covered by the Biocrates AbsoluteIDQ® p180 kit, they are related to amino acid metabolism, either via methionine or proline, and were added to Figure S11 its putative results. N-Acetylmethionine, a derivative from the essential amino acid methionine, is either the result of direct synthesis by specific N-acetyltransferases or via the proteolytic degradation of N-acetylated proteins by hydrolases and is less abundantly present in the grade 3 steatosis. 1-Pyrroline-4-hydroxy-2-carboxylate is most likely the breakdown product from hydroxy-L-proline and is more abundant in grade 3 steatosis. L-4-Hydroxyglutamate semialdehyde is also an intermediate breakdown product of proline, but via 1-Pyrroline-3-hydroxy-5-carboxylate and entering the citrate cycle via pyruvate and is more abundantly present in the grade 3 steatosis.

Table S10. Top pathways revealed upon mummichog of untargeted metabolomics. The compounds retrieved in the pathways are putative IDs, not experimentally confirmed, and contain retrieved metabolites which were not significantly discriminating between control and grade 3 steatosis as well. The significantly discriminating metabolites are highlighted in bold.

| **Pathways** | **Overlap size** | **Pathway size** | **p-value** | **Retrieved Compounds in pathway** |
|---|---|---|---|---|
| Purine metabolism | 4 | 28 | 0.00143 | Inosine, 5-Hydroxyisourate, D-Glutamine, **L-4-Hydroxyglutamate semialdehyde** |
| Nitrogen metabolism | 2 | 6 | 0.00302 | D-Glutamine, **L-4-Hydroxyglutamate semialdehyde** |
| Glutathione Metabolism | 2 | 9 | 0.00672 | **1-Pyrroline-4-hydroxy-2-carboxylate**, **L-4-Hydroxyglutamate semialdehyde** |
| Arginine and Proline Metabolism | 3 | 26 | 0.00723 | **1-Pyrroline-4-hydroxy-2-carboxylate**, D-Glutamine, **L-4-Hydroxyglutamate semialdehyde** |
| Vitamin B3 (nicotinate and nicotinamide) metabolism | 2 | 10 | 0.00781 | D-Glutamine, **L-4-Hydroxyglutamate semialdehyde** |
| Glutamate metabolism | 2 | 10 | 0.00781 | D-Glutamine, **L-4-Hydroxyglutamate semialdehyde** |

| | | | | |
|---|---|---|---|---|
| Alanine and Aspartate Metabolism | 2 | 11 | 0.01042 | D-Glutamine, **L-4-Hydroxyglutamate semialdehyde** |
| Aminosugars metabolism | 2 | 16 | 0.01983 | D-Glutamine, **L-4-Hydroxyglutamate semialdehyde** |
| Valine, leucine and isoleucine degradation | 2 | 19 | 0.02412 | L-Isoleucine, **L-4-Hydroxyglutamate semialdehyde** |
| Carbon fixation | 1 | 3 | 0.02571 | D-Erythrose 4-phosphate |
| Lysine metabolism | 2 | 22 | 0.03042 | N6,N6,N6-Trimethyl-L-lysine, **L-4-Hydroxyglutamate semialdehyde** |
| Aspartate and asparagine metabolism | 3 | 49 | 0.03176 | **1-Pyrroline-4-hydroxy-2-carboxylate**, D-Glutamine, **L-4-Hydroxyglutamate semialdehyde** |
| Methionine and cysteine metabolism | 2 | 26 | 0.03832 | **L-4-Hydroxyglutamate semialdehyde**, **N-Acetylmethionine** |
| Pyrimidine metabolism | 2 | 29 | 0.04571 | D-Glutamine, **L-4-Hydroxyglutamate semialdehyde** |

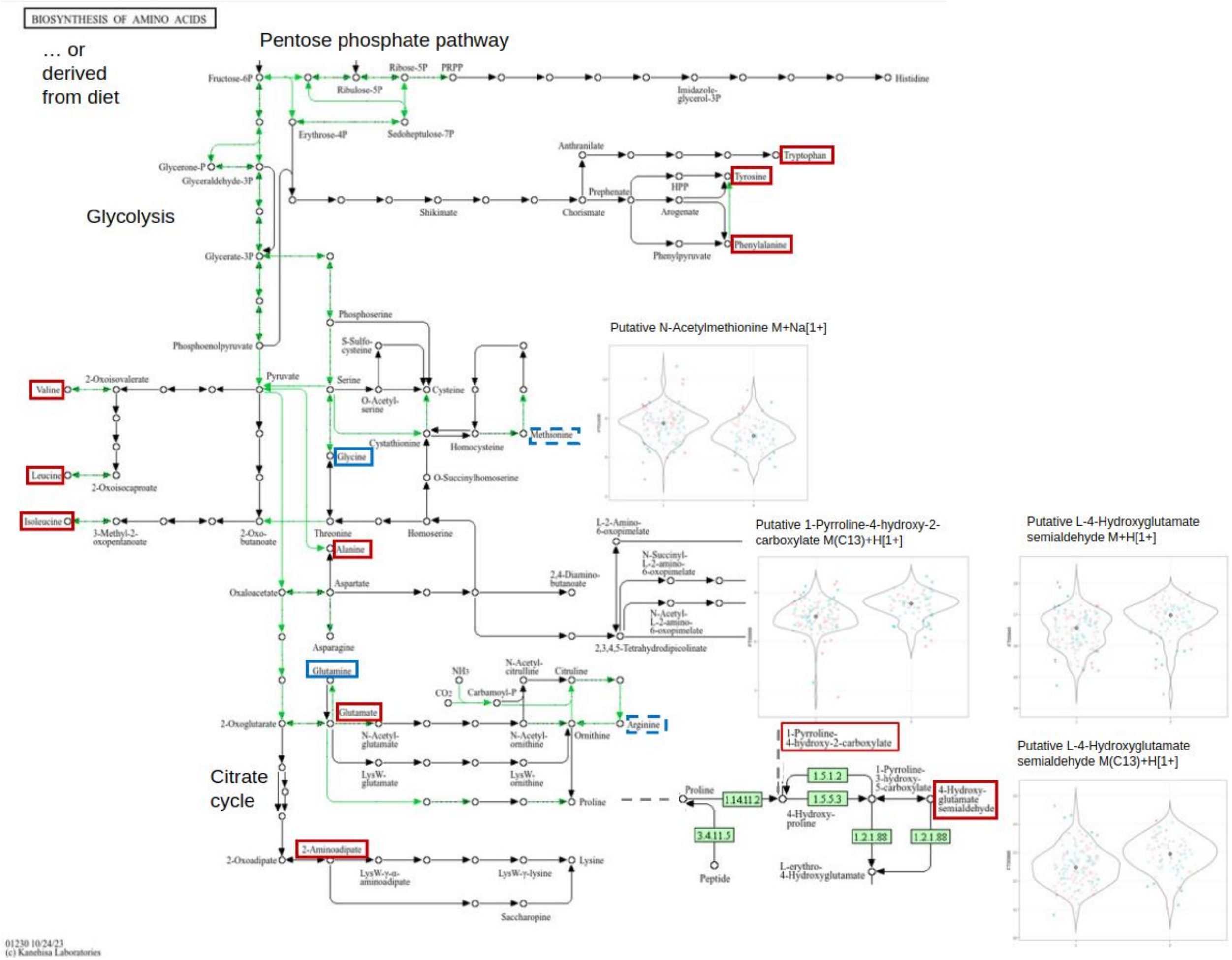


Figure S11. Altered biochemical amino acid pathways of differentiating putatively identified metabolic features (adducts) between control and grade 3 steatosis. Boxplots left, control; right, grade 3 steatosis. Red and blue dots are female and male participants, respectively. Figure based on KEGG, enzyme reaction catalyzed in humans in green lines, closely related amino acid in dashed box (Kanehisa Laboratories 01230 10/24/23). Red boxes indicate higher abundance in steatosis grade 3, blue boxes higher abundance in control individuals. Violin plots left, control; right, grade 3 steatosis.
