## supplementary_information3 for "Molecular characterization of liver steatosis through circulating proteins, metabolites and lipids" (from another version of this preprint: version 1, 2025-12-02)

**Title**

**Short Title**

Blood-based profiling of liver steatosis

**Authors**

Marilyn De Graeve[1], Essi Hantikainen[1], Philippine Louail[1], Vinicius Verri Hernandes[1,2], Cristian Pattaro[1], Francisco S. Domingues[1], Peter P. Pramstaller[1], Herbert Tilg[3], Markus Ralser[4,5,6], Christoph Grander[3,*], Johannes Rainer[1,*]

**Affiliations**

[1]Institute for Biomedicine, Eurac Research, 39100 Bolzano, Italy

[2]Faculty of Chemistry, Department of Food Chemistry and Toxicology, University of Vienna, Währinger Straße 38, 1090 Vienna, Austria

[3]Department of Internal Medicine I, Gastroenterology, Hepatology, Endocrinology & Metabolism, Medical University Innsbruck, 6020 Innsbruck, Austria.

[4]Institute of Biochemistry, Charité Universitätsmedizin Berlin, 10117 Berlin, Germany

[5]Max Planck Institute for Molecular Genetics, 14195 Berlin, Germany

[6]The Wellcome Centre for Human Genetics, Nuffield Department of Medicine, University of Oxford, Oxford OX3 7BN, UK

* Contributed equally

**Corresponding author**

### SI information 3: Altered pathways involving circulating proteins (cont.)

Another second biological process of interest is the increased platelet degranulation due to elevated cytosolic $Ca^{2+}$ levels, upon which SERPINA3, ITIH3 and ITIH4 and other proteins are secreted. While SERPINA3 undergoes exocytosis from platelet alpha granule lumen, the ITIHs undergo exocytosis from platelet dense granule lumen[1] (Figure S6A). It is a hypothesis, since plasma circulating SERPINA3, ITIH3 and ITIH4 are all down, less such elevated cytosolic $Ca^{2+}$ levels occur under grade 3 steatosis conditions.

While the third enriched pathway, the network map of SARS-CoV-2 signaling pathway (wiki pathways WP5115) revealed the collectively controlled positive regulation of protein expression of ITIH3, ITIH4, LRG1, CFI and CFH in the peripherical blood mononuclear cells (PBMC) and of GSN, APOD and APOM in serum of severe/critical patients upon SARS-CoV-2 infection[2], it was considered off topic because of the different inflammation grade it instigates. Viral infection gives a high inflammatory response, while steatosis is considered the result of a low-grade long-term pro-inflammatory state, similar to obesity[3]. In addition, there is currently an overreporting bias on COVID compared to steatosis, and for grade 3 steatosis in the CHRIS-NAFLD cohort, an upregulation of CFI and CFH and downregulation of LGR1, ITIH3 and ITIH4 were noted instead of a collective upregulation (Figure S6B). However, the overlap in involved circulating proteins confirms our finding that the question to look at differences between control and grade 3 steatosis is not exclusively related to steatosis but also finds some overlap with non- versus MASLD afflicted participants in literature. This leads us to a hypothesis that the altered protein levels represent more of a metabolically healthy versus metabolically unhealthy phenotype, than a signature biomarker to define steatosis.

A

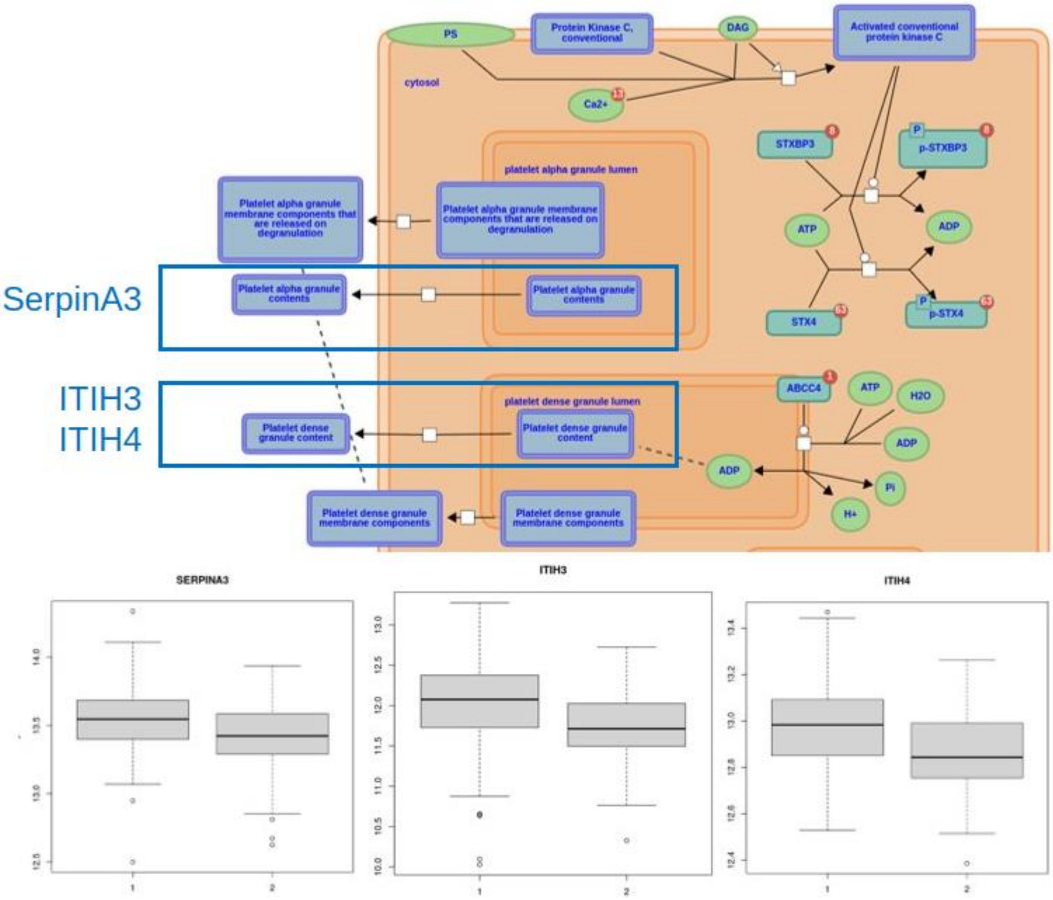
PS
Protein Kinase C, conventional
DAG
Activated conventional protein kinase C
cytosol
Ca2+
STXBP3
p-STXBP3
platelet alpha granule lumen
Platelet alpha granule membrane components that are released on degranulation
Platelet alpha granule membrane components that are released on degranulation
ATP
ADP
SerpinA3
Platelet alpha granule contents
Platelet alpha granule contents
STX4
p-STX4
ITIH3
ITIH4
platelet dense granule lumen
ABCC4
ATP
H2O
Platelet dense granule content
Platelet dense granule content
ADP
ADP
Pi
H+
Platelet dense granule membrane components
Platelet dense granule membrane components
SERPINA3
ITIH3
ITIH4

B

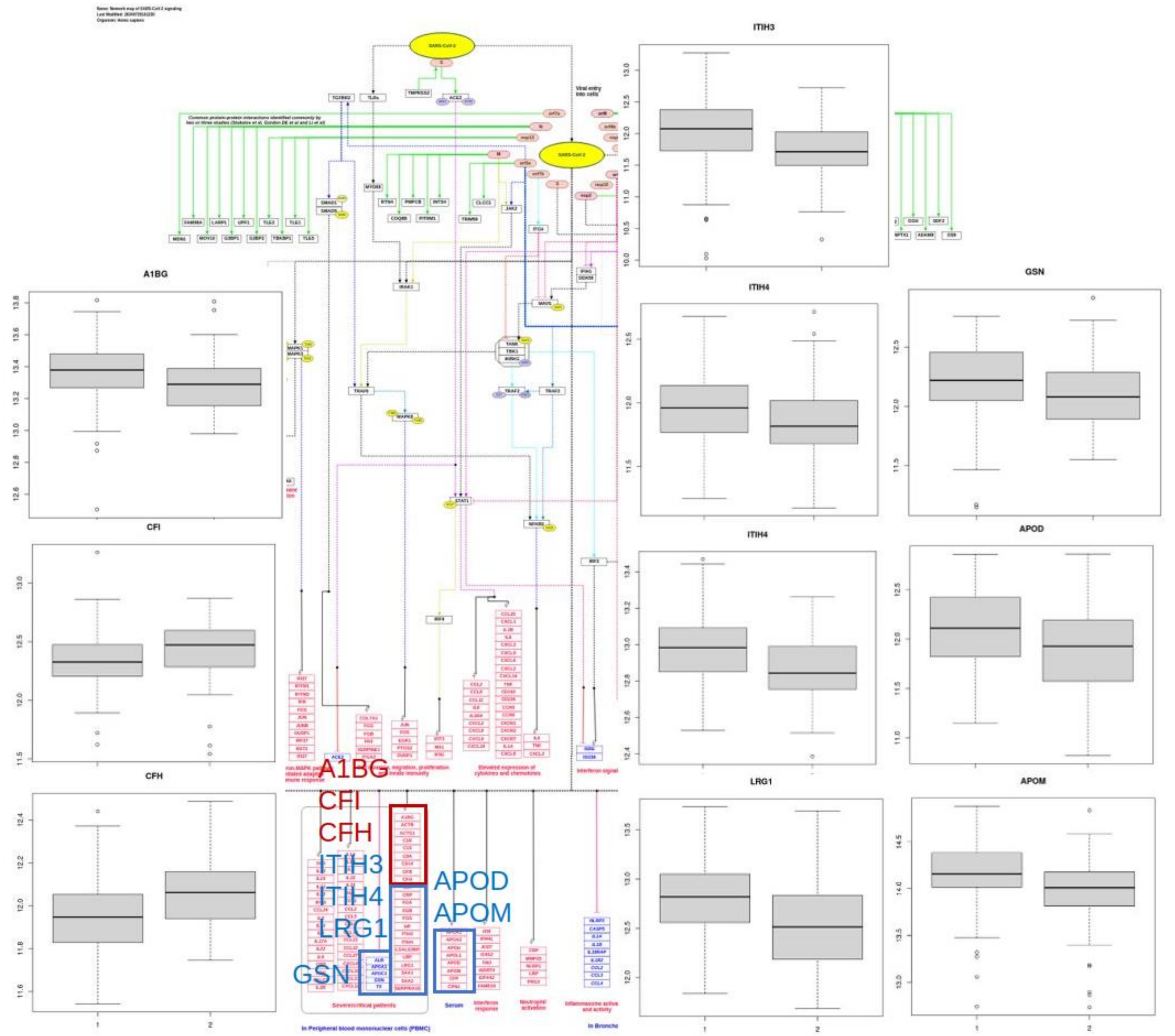


Figure S. Altered biochemical pathways of differentiating proteins between control and grade 3 steatosis. Response to elevated platelet cytosolic Ca2+ (A). Altered biochemical pathways of differentiating proteins between control and grade 3 steatosis. Network map of SARS-CoV-2 signaling pathway (B). Red boxes indicate higher abundance in steatosis grade 3, blue boxes higher abundance in control individuals. Boxplots left, control; right, grade 3 steatosis.
