## supplementary_information4 for "Molecular characterization of liver steatosis through circulating proteins, metabolites and lipids" (from another version of this preprint: version 1, 2025-12-02)

**Title**

**Short Title**

Blood-based profiling of liver steatosis

**Authors**

Marilyn De Graeve[1], Essi Hantikainen[1], Philippine Louail[1], Vinicius Verri Hernandes[1,2], Cristian Pattaro[1], Francisco S. Domingues[1], Peter P. Pramstaller[1], Herbert Tilg[3], Markus Ralser[4,5,6], Christoph Grander[3,*], Johannes Rainer[1,*]

**Affiliations**

[1]Institute for Biomedicine, Eurac Research, 39100 Bolzano, Italy

[2]Faculty of Chemistry, Department of Food Chemistry and Toxicology, University of Vienna, Währinger Straße 38, 1090 Vienna, Austria

[3]Department of Internal Medicine I, Gastroenterology, Hepatology, Endocrinology & Metabolism, Medical University Innsbruck, 6020 Innsbruck, Austria.

[4]Institute of Biochemistry, Charité Universitätsmedizin Berlin, 10117 Berlin, Germany

[5]Max Planck Institute for Molecular Genetics, 14195 Berlin, Germany

[6]The Wellcome Centre for Human Genetics, Nuffield Department of Medicine, University of Oxford, Oxford OX3 7BN, UK

* Contributed equally

**Corresponding author**

### SI information 4: Altered pathways involving circulating metabolites and lipids (cont.)

The carnitines are involved in two crucial central metabolism pathways, segregated in two distinct locations. Carnitine and C3 are involved in the lipid metabolism, i.e. the b-oxidation, in the peroxisomes (Figure S7A); while the other significantly upgraded acylcarnitines (C3, C4, C5) are products of the catabolism of the branched-chain amino acids (BCAAs) leu, ile and val in the cytosol and subsequently in the mitochondria. C3 is not transported across cellular membranes, so in both locations, C3 is more abundant in grade 3 steatosis. As the metabolites were circulating, i.e. measured in blood serum, their measurements are a direct result of BCAA catabolism. Carnitine is also linked with glycine, as the biosynthesis of C0 includes the release of glycine in the conversion step from 3-hydroxy-N(6),N(6),N(6)-trimethyl-L-lysine to 4-trimethylammoniobutanal by a serine hydroxymethyltransferase tetramer (SHMT1, Reactome R-HSA-71243, Figure S7B).

A

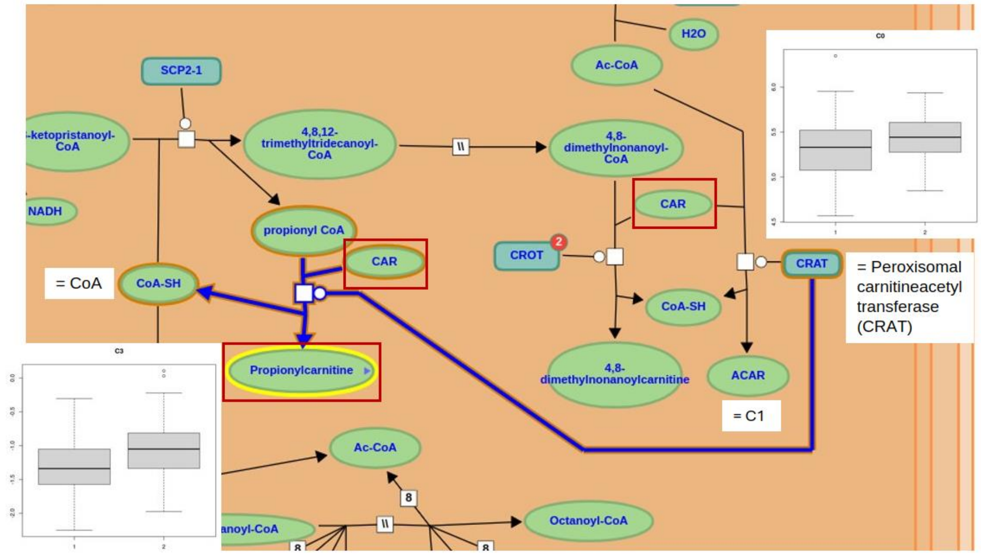


B

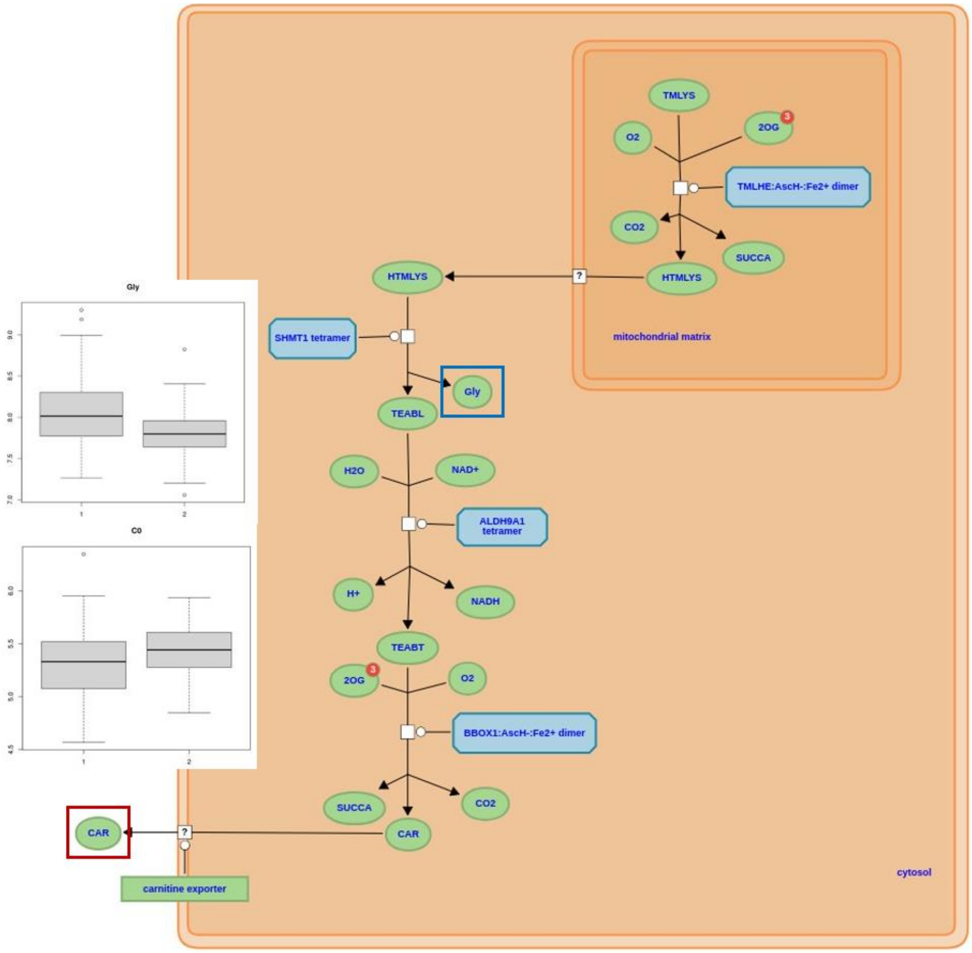


Figure S7. Altered biochemical pathways of differentiating metabolites and lipids between control and grade 3 steatosis. Peroxisomal lipid metabolism (A). Carnitine synthesis (B). Red boxes indicate higher abundance in steatosis grade 3, blue boxes higher abundance in control individuals. Boxplots left, control;

right, grade 3 steatosis.

For the lyso- and PCs are collected under the biochemical pathway of the glycerophospholipid metabolism (KEGG map00564, Figure S8A) and the phosphatidylcholine catabolism (wiki pathways WP4195, Figure S8B). SMs are also connected to (lyso)PCs in the latter pathway via choline as a shared intermediate molecule. With the 2 main routes of biosynthesis either from activated choline and 1,2-diacyl-sn-glycerol, or from phosphatidylethanolamine (PE) to PC. As half of the discriminating PCs have more than two fatty acyl unsaturations (so more than 1 unsaturation per acyl chain), most of them are PCs with polyunsaturated fatty acyl chains (PUFA, n=12). These are exclusively derived from the diet and can be swapped with an unsaturated or monosaturated acyl chain in the PC moiety after biosynthesis. Diets sources are vegetable, fish and animal oils and fats, e.g. linoleic acid (C18:2) from sunflower oil and arachidonic acid (C20:4) from animal fat. These two omega 6 PUFAs are also incorporated in 2 of the lysoPCs as acyl moiety. The decrease of these important omega 6 PUFA-bound lysoPCs, and possibly also omega 3 fatty acids in PCs, does not reflect the abundance of free PUFAs. Derived from the diet, but no evidence of this could not be extracted from the FFQ. And increased hydrolysis of PUFA containing (lyso)PCs, can result in more ARA and LA as free fatty acid, which might be diverted into the eicosanoid synthesis and result in a pro-inflammatory state. Therefore, these 2 specific lysoPCs could be biomarkers of metabolically liver lipoprotein dysregulation, though probably not exclusive to steatosis.

A

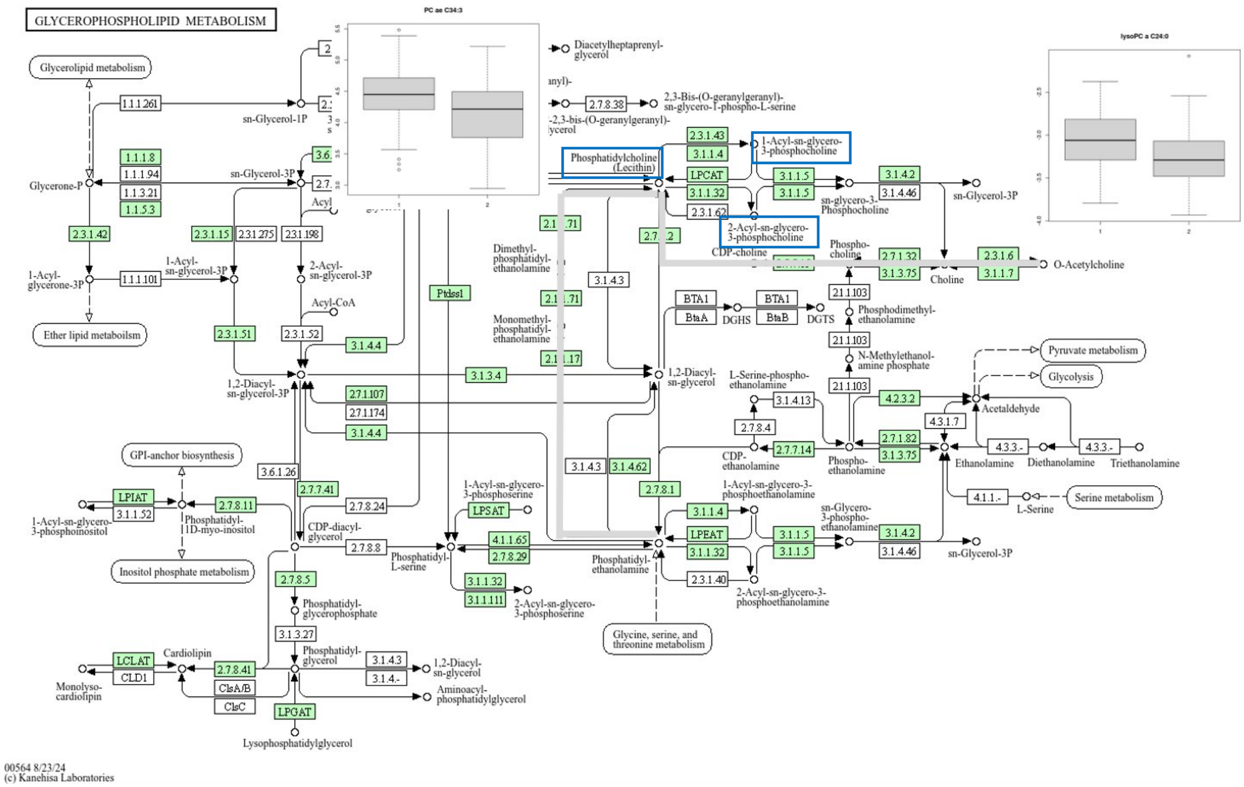


B

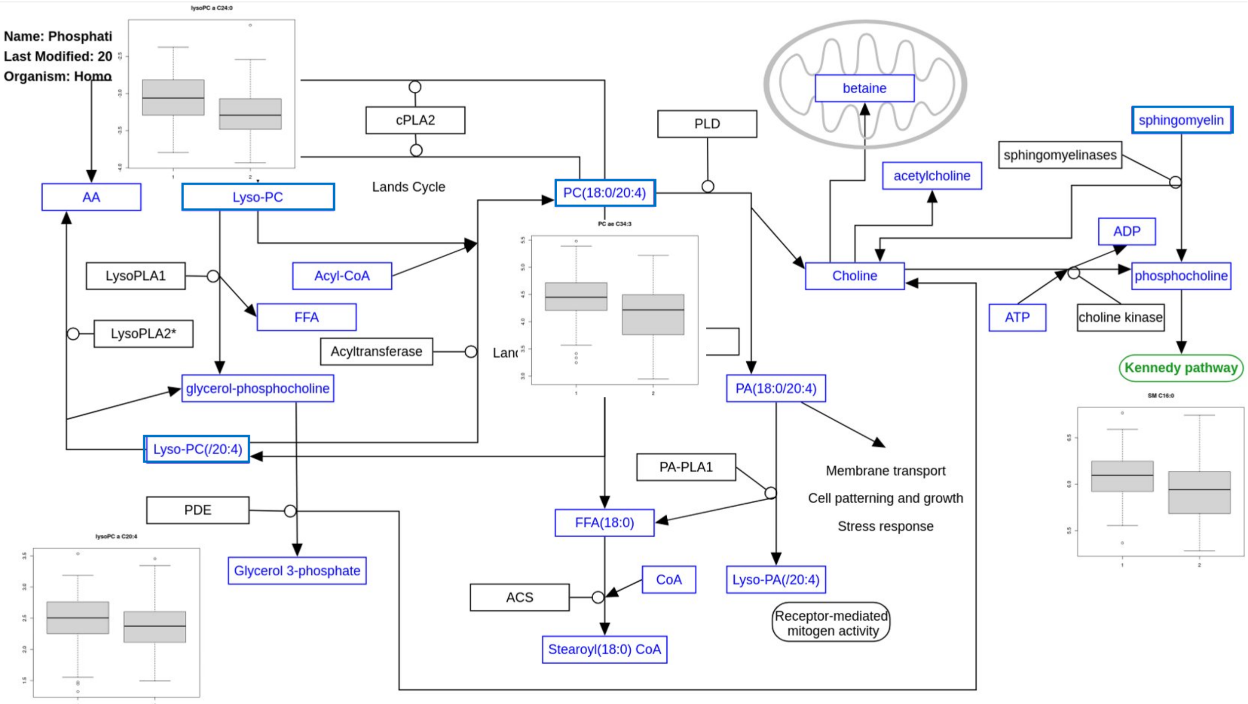


Figure S8. Altered biochemical pathways of differentiating metabolites and lipids between control and grade 3 steatosis. Glycerophospholipid metabolism (A). Phosphatidylcholine catabolism (B). Red boxes indicate higher abundance in steatosis grade 3, blue boxes higher abundance in control individuals.

Boxplots left, control; right, grade 3 steatosis.

The significantly downregulated apolipoproteins, APOD and APOM were not represented in enriched pathways, therefore no direct link to the listed significant metabolites and lipids was detected. Though, indirectly APOs are linked to the transport of small hydrophobic molecules such as lipids, retinoids, steroids and bilins. A decrease of APO proteins is possibly linked to a decreased level of (lyso)PCs and SMs in serum of grade 3 steatosis afflicted participants.

In addition to its connection to PCs, significant SMs are also represented in the sphingolipid metabolism: integrated pathway (Figure S9), but don't interact with other metabolites of interest. Therefore, it is not further discussed.

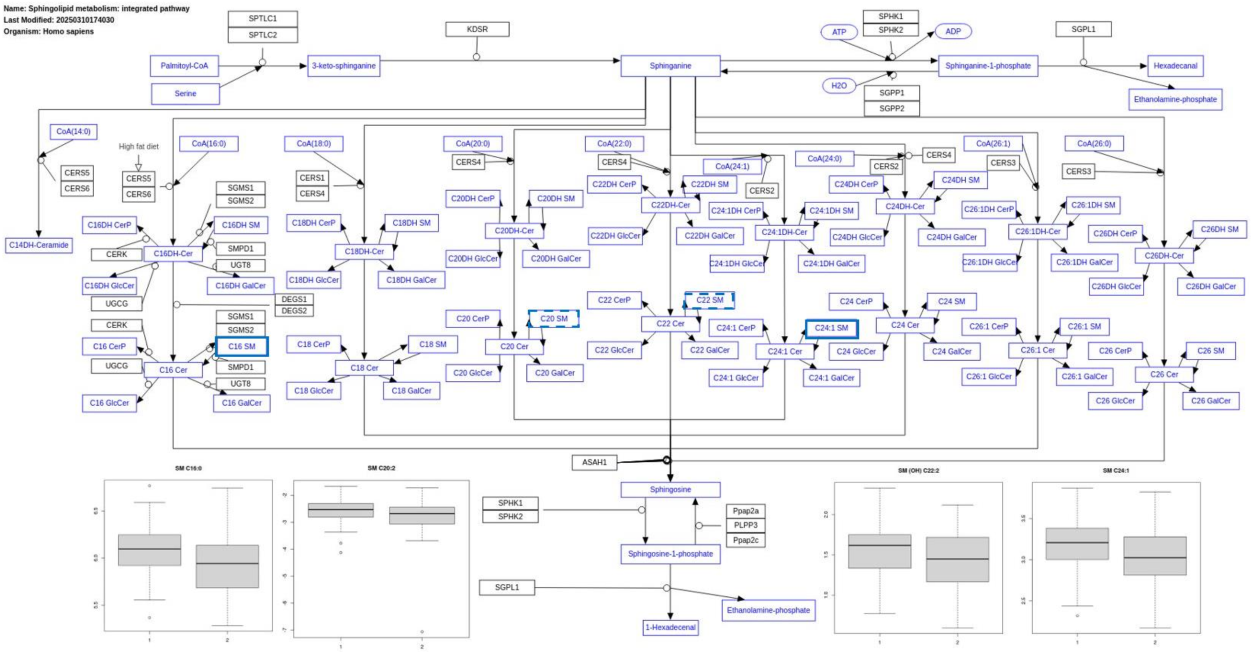


Figure S9. Altered biochemical pathways of differentiating metabolites and lipids between control and grade 3 steatosis. Sphingolipid metabolism: integrated pathway. SM (OH) C22:2 and SM C20:2 are related to SM(d18:1/22:0) and SM(d18:1/20:0) respectively and therefore depicted by a dotted box. Red boxes indicate higher abundance in steatosis grade 3, blue boxes higher abundance in control individuals. Boxplots left, control; right, grade 3 steatosis.
